# A germline KDM3C polymorphism impairs DNA repair and sensitizes to chemoradiotherapy

**DOI:** 10.64898/2026.08.26.26360896

**Authors:** Adria Hasan, Elena V. Demidova, Pragya Priyadarshini, Philip Czyzewicz, Leonny Gathuka, Takahiko Murayama, Yan Zhou, Zachary AC Kiss, Raksha KR Shastry, Mark Andrake, Gavin Hearne, Karthik Devarajan, Chao Wu, Anshul Shah, Bryant M. Schultz, Denise C. Connolly, Gail L. Rosen, Israel Cañadas, Jeffrey C. Liu, Barbara A. Burtness, J Joshua Smith, Roland L. Dunbrack, Erica A. Golemis, Johnathan R. Whetstine, Joshua E. Meyer, Sanjeevani Arora

**Affiliations:** Cancer Prevention and Control Program, Fox Chase Cancer Center, Philadelphia, PA; Cancer Signaling and Microenvironment, Fox Chase Cancer Center, Philadelphia, PA; Nuclear Dynamics and Cancer, Fox Chase Cancer Center, Philadelphia, PA; Department of Biostatistics and Bioinformatics, Fox Chase Cancer Center, Philadelphia, PA; Department of Radiation Oncology, Fox Chase Cancer Center, Philadelphia, PA; Ecological and Evolutionary Signal-Processing and Informatics Laboratory, Department of Electrical and Computer Engineering, College of Engineering, Drexel University, Philadelphia, PA; Department of Colon and Rectal Surgery, Division of Surgery, The University of Texas MD Anderson Cancer Center, Houston, TX; Biosample Repository Facility, Fox Chase Cancer Center, Philadelphia, PA; Cancer Epigenetics Institute, Fox Chase Cancer Center, Philadelphia, PA; Department of Internal Medicine, Yale Cancer Center, Yale School of Medicine, New Haven, CT; Department of Cancer and Cellular Biology, Lewis Katz School of Medicine, Temple University, Philadelphia, PA

## Abstract

Chemoradiotherapy (CRT) is the standard-of-care therapy for many solid malignancies, yet predictive biomarkers of treatment response remain limited. We identified a germline single nucleotide polymorphism (SNP) in an intrinsically disordered region of the lysine demethylase KDM3C/JMJD1C (p.S464T) that is associated with CRT outcomes in locally advanced rectal cancers (LARC) and head and neck squamous cell carcinoma (LA-HNSCC). *In silico* modeling with AlphaFold predicted S464T substitution influenced interaction between phosphorylated KDM3C and RNF8 FHA domain. In cellular models, conversion of S464 to T464 increased sensitivity to DNA-damaging agents. S464T substitution impaired damage-induced MDC1-RAP80 signaling and downstream RAP80-BRCA1 colocalization. SNP carrying cells impaired DNA repair causing genotoxic stress that is associated with increased cGAS-cGAMP innate immune signaling and increased apoptosis. Population analyses with the SNP highlighted an increase incidence of UV-induced skin and other cancers, linking inherited variation in the chromatin regulatory gene *KDM3C* to genome instability, cancer risk, and therapeutic vulnerability.

**Statement of Significance:** We identify a germline polymorphism in *KDM3C*/*JMJD1C* that disrupts DNA damage repair signaling and enhances sensitivity to DNA-damaging therapy. These findings reveal a mechanistic link between inherited variation in chromatin regulation and therapeutic response, providing a framework for biomarker-driven stratification of patients receiving chemoradiotherapy.

## INTRODUCTION

Chemoradiotherapy (CRT) is a cornerstone of multimodal therapy for multiple solid tumors including locally advanced rectal cancer (LARC) (1,2) and head and neck squamous cell carcinoma (LA-HNSCC) (3,4), yet patient responses vary widely (5–7). In LARC, neoadjuvant CRT (nCRT) is routinely utilized to reduce tumor burden, improve surgical outcomes or allow for organ preservation through surgical omission (8), yet responses range from complete pathological regression to minimal or no therapeutic benefit (9–11). Similarly, CRT is a primary curative modality for many LA-HNSCC patients, but treatment outcomes are heterogeneous with substantial differences in locoregional control and long-term survival (12,13). Despite the central role of CRT in the management of these malignancies, the biological factors that determine therapeutic response remain incompletely understood (7,14,15).

In LARC, response to neoadjuvant chemoradiotherapy has been associated with several clinical (16,17) and tumor-intrinsic features, including tumor stage (16), *KRAS* mutation status (18,19), tumor hypoxia (20,21), and immune infiltration (22); however, these factors have not consistently explained the variability in therapeutic outcomes observed across patients (19,23). Similarly, in LA-HNSCC, in addition to HPV status (24,25), predictors such as tumor burden (26), hypoxia (27), and somatic genomic alterations (28–30) have been associated with CRT sensitivity, yet these parameters incompletely account for the wide heterogeneity in treatment response.

Because CRT exerts its cytotoxic effects primarily through the induction of DNA damage, particularly DNA double-strand breaks (DSBs) (31,32), individual differences in DNA repair capacity represent a plausible and underexplored source of variability in treatment response. It is well established that defects in DSB repair influence responses to genotoxic therapies (33). For example, germline and somatic alterations central mediators of homologous recombination (HR), such as *BRCA1* and *BRCA2*, profoundly influence responses to DNA-damaging treatments (34). However, the extent to which broader germline variation across DNA repair networks contribute to variability in CRT response is an open question. This knowledge gap provides a strong rationale for systematic investigation of determinants of DNA repair capacity across both core DNA repair pathways and the broader cellular networks that regulate repair as potential drivers of variability in CRT response.

KDM3C is a Jumonji domain-containing chromatin regulator that was initially characterized as a histone demethylase capable of removing methyl groups from H3K9me1/2 chromatin marks, modifications generally associated with transcriptional control (35–43). KDM3C functions as a H3K9 demethylase, however, the function appears to go beyond histone demethylation alone (43). Recent studies also suggest that KDM3C may also have non-enzymatic mechanisms as a transcriptional co-activator for transcription factors [*e.g.*, HOXA9] (35,44).

Prior work has also demonstrated that *KDM3C* directly participates in double-strand break (DSB) repair by modulating the MDC1–RNF8 signaling axis, promoting downstream recruitment of RAP80–BRCA1 complexes to sites of DNA damage events in a process essential for BRCA1-mediated DSB repair and cellular responses to genotoxic therapy (45). These observations support KDM3C as a potential regulator of therapeutic response to DNA-damaging treatments.

Here, we performed a discovery analysis of DNA repair pathways and regulatory networks that impact DNA repair to identify germline determinants of response to CRT. Our analysis encompassed canonical DNA repair pathways alongside complementary processes that influence repair capacity, including cell-cycle regulation, chromatin programs, immune signaling, and post-translational protein modification networks. Through this framework, we identified *KDM3C* (also known as *JMJD1C*) as a candidate regulator of response to CRT. A germline coding missense single nucleotide polymorphism (SNP) in *KDM3C* (p.S464T; T/T) associated with improved response to CRT in patients with LARC and LA-HNSCC. Mechanistically, this variant disrupts assembly of the MDC1–RNF8–RAP80–BRCA1 repair complex, leading to impaired DNA damage and enhanced sensitivity to DNA-damaging therapies. Our findings establish germline variation in *KDM3C* as a previously unrecognized determinant of therapeutic vulnerability to DNA damage. This discovery highlights inherited variation in epigenetic proteins and DNA damage response pathways as an important contributor to interindividual differences in responses to genotoxic cancer therapies.

## RESULTS

### Germline missense polymorphism in *KDM3C* associates significantly with favorable outcomes

To identify candidate germline determinants associated with response to CRT, we performed whole-exome sequencing (WES; 100× coverage) on peripheral blood-derived DNA from an initial cohort of 30 LARC patients. We curated a 161-gene panel encompassing core DNA repair pathways and processes that influence DNA repair, reasoning that both common and rare variation could be relevant to therapy response. (**Figure 1A**, **Supplementary Table 1**, see **Methods** for analysis pipeline). This approach ensured that the panel encompassed not only canonical DNA repair genes but also supporting pathways critical for maintaining genomic integrity. To functionally organize the gene set, we assigned genes to pathway categories using WebGestalt (WEB-based Gene Set Analysis Toolkit) 2019 (46), which allowed systematic evaluation of their distribution across biologically relevant pathways (**Supplementary Table 1-5**). Analysis of WES data identified 77 genes harboring candidate variants (**Supplementary Table 3, 6**).

**Figure 1.**
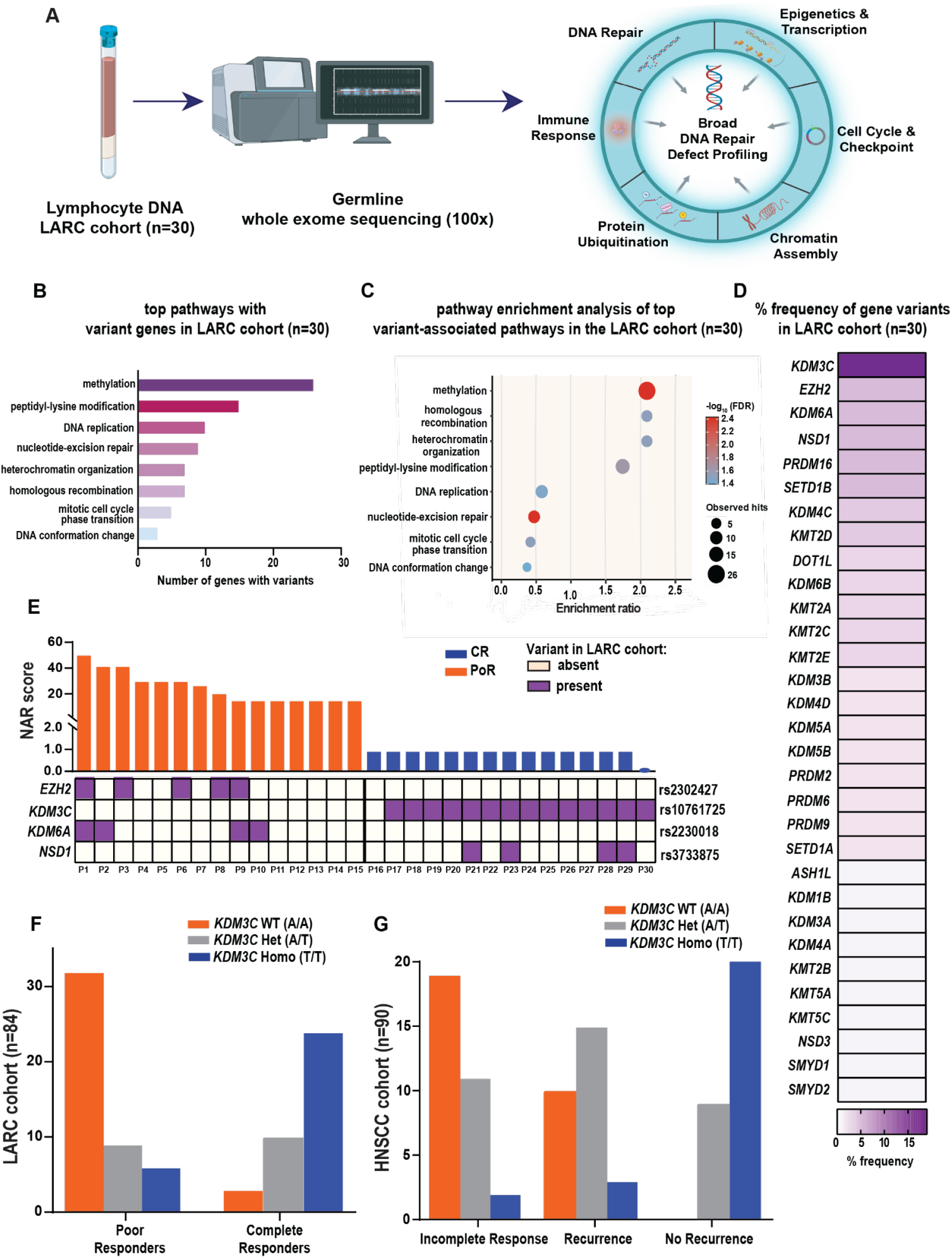
KDM3C p.S464T germline SNP associates with CRT outcomes in LARC and HNSCC. **(A)** Schematic of the study workflow. Pretreatment lymphocyte DNA was isolated from patients with LARC (n=30) and subjected to germline whole-exome sequencing (100×) to enable broad profiling of inherited variants across pathways related to DNA repair, epigenetics and transcription, cell cycle and checkpoint control, protein ubiquitination, chromatin assembly, and immune response. Created in BioRender. Adria Hasan, F. (2026) https://BioRender.com/bnshk69. **(B)** Top pathways containing variant genes identified across the LARC cohort (n=30) **(C)** Pathway enrichment analysis of top variant-associated pathways in the LARC cohort. Dot size indicates the number of observed hits, and dot color represents significance as –log10 (FDR). **(D)** Frequency of variants in methylation-associated genes across the LARC cohort. **(E) Top:** NAR scores associated with response to nCRT in LARC cohort (n=30). **Bottom:** SNPs associated with response to nCRT in 30 LARC patients**. (F)** Genotyping in LARC patients (n=84) reveals that KDM3C SNP-S464T (T/T) is significantly enriched in the CR group A/A - PoR vs. CR, ***P<0.0001, A/T - PoR vs. CR, NS, T/T - PoR vs. CR, **p<0.001. **(G)** LA-HNSCC patients (n=90) were segregated into 3 groups and SNP genotyping was performed. No-recurrence is less likely in the WT-S464(A/A) than in the non-WT counterparts (p=0.0003, OR=0), Association between outcome & genotype: p<0.000001, Fisher’s Exact Test.

We tested whether the observed distribution of variants across functional categories was non-random, reasoning that certain pathways might harbor a disproportionate number of variants. Among the significantly enriched pathways, methylation genes showed the strongest enrichment (26 genes with variants; q= 0.0037). Additional enriched pathways included peptidyl-lysine modification (15 genes; q= 0.0234), HR (7 genes; q= 0.0302), heterochromatin organization (7 genes; q= 0.0302), DNA conformation change (3 genes; q= 0.0399), mitotic cell-cycle phase transition (5 genes; q= 0.0302), nucleotide-excision repair (9 genes; q= 0.0037), and DNA replication (10 genes; q= 0.0361) (**Figure 1B–C**). Notably, SNPs identified in multiple patients were found in methylation genes, including *KDM3C, EZH2, KDM6A*, and *NSD1* (**Figure 1D**).

Next, we stratified the LARC patients by clinical outcomes based on Neoadjuvant Rectal (NAR) score, a validated surrogate endpoint in LARC that integrates tumor downstaging and nodal response following nCRT that captures the cumulative response to therapy (47,48). Based on NAR score, the cohort was segregated into complete responders (CR; n=15), defined as patients with most favorable post-treatment response and poor responders (PoR; n=15) (**Figure 1E, top**), defined as patients with the least favorable post-treatment response. Across the cohort, common single nucleotide polymorphisms (SNPs) were most frequently detected in *KDM3C* (T/T, p.S464T, rs10761725; 14/30 patients), *EZH2* (C/C, p.D185H, rs2302427), *KDM6A* (A/A; p.T778K, rs2230018; 4/30), and *NSD1* (T/T p.V614L, rs3733875; 4/30). Strikingly, KDM3C p.S464T (14/15; p<0.0001) and NSD1 p.V614L (4/15) were found exclusively in CRs, whereas EZH2 p.D185H (5/15, p<0.05) and KDM6A p.T778K (4/15) were found exclusively in PoRs (**Figure 1E, bottom right**). At the pathway level, CRs were significantly enriched in methylation (16 genes; q=0.0019), peptidyl-lysine modification (13 genes; q= 0.0019), and nucleotide-excision repair (4 genes; q= 0.0494), while no pathways were significantly enriched in the PoR group after FDR correction (**Supplementary Figure 1, Supplementary Table 7 and 8**). These collective candidates support the role of chromatin regulatory pathways in contributing to therapeutic response outcomes following DNA-damaging therapy.

Given the striking association of the *KDM3C* T/T with treatment response outcomes in the initial cohort, we performed expanded genotyping in an additional set of 54 LARC patients (bringing the total in LARC cohort to 84) (**Supplementary Table 9**). Here, the CRs predominantly carried the *KDM3C* T/T genotype, whereas PoRs carried the *KDM3C* A/A genotype (**Figure 1F**; ***p<0.0001, Fisher’s Exact test). The heterozygous *KDM3C* A/T genotype was not associated with response status. Herein, the reference A/A allele is designated as WT-S464, and the alternative T/T allele, corresponding to the p.S464T substitution, is designated as SNP-S464T.

We next evaluated the *KDM3C* SNP-S464T in an independent retrospective cohort of LA-HNSCC patients who had received CRT (n=90) (**Supplementary Table 10**). Recurrence outcome was used as a clinically relevant endpoint in HNSCC as it reflects failure of durable response to CRT and more directly captures tumor-intrinsic treatment resistance than overall survival (49,50). We observed a significant association between recurrence status, and the *KDM3C* genotype in the HNSCC cohort (p<0.000001). Here, recurrence was observed more among patients with the A/A genotype (WT-S464) compared with patients carrying the non-WT genotypes, indicating enrichment of the T-allele–containing genotypes among patients who remained recurrence-free after treatment (**Figure 1G**; p=0.0003, OR=0; Fisher’s Exact Test). Collectively, these findings identify the *KDM3C* homozygous T/T genotype (SNP-S464T) as a promising candidate for predicting favorable CRT outcome across two cancer types. Conversely, enrichment of the WT-S464 among less favorable outcome groups may have future relevance for identifying patients who could benefit from alternative or intensified treatment strategies.

Population-level annotation in gnomAD v4.1.1 demonstrated that *KDM3C* rs10761725 is a common polymorphism, with allele frequencies approaching ∼0.7-0.8 across several gnomAD populations (**Supplementary Table 11**). Among the gnomAD ancestry groups examined, the expected T/T genotype frequency for *KDM3C* rs10761725 was highest in the Ashkenazi Jewish population (0.67453), followed by Amish (0.64112), European (Finnish) (0.62806), and European (non-Finnish) (0.61325), and lowest in the African/African American population (0.14273) (**Supplementary Table 12**). The high population prevalence is consistent with a potential role as a population-level modifier of DNA repair and therapeutic sensitivity.

We leveraged TCGA dataset to evaluate the relationship between germline *KDM3C* status, and patient demographics or underlying somatic driver landscapes in rectal cancer (n = 145) and HNSCC (n = 493) cohorts (see **Supplementary Results**). We first assessed whether *KDM3C* genotype distribution was associated with patient demographic characteristics. In the rectal cancer cohort, *KDM3C* genotype distribution was largely independent of patient age (p= 0.535) or sex (p= 0.548), displaying only a borderline association with race (p= 0.056). Conversely, within the larger LA-HNSCC cohort, the distribution of the *KDM3C* genotype showed a highly significant association with race (p= 0.00024), driven by an enrichment of the A/A allele (WT-S464) in African American patients, directly mirroring the population-level gnomAD analysis. Additionally, *KDM3C* genotype correlated significantly with patient age groups in HNSCC (p= 0.049), while remaining independent of sex (p= 0.734). Collectively, these findings indicate that the *KDM3C* SNP-S464T distribution exhibits distinct population-level and age-related stratification depending on the cancer context, although differences in TCGA cohort size between the rectal cancer and HNSCC groups may influence these associations.

To analyze the somatic driver landscapes according to the germline *KDM3C* genotype status, we stratified TCGA cohorts accordingly (**Supplementary Results** and **Supplementary Table 13)**. Across TCGA rectal tumors and HNSCCs, *KDM3C* genotype stratification did not reveal major genotype-specific differences in the distribution of common somatic driver alterations. Rectal tumors showed recurrent alterations in *TP53, KRAS, APC, SMAD4*, and *BRAF*, consistent with established rectal cancer genomic landscapes in which these genes are among commonly altered driver genes (51). HNSCCs demonstrated recurrent mutations in *TP53, PIK3CA, NOTCH1, CDKN2A, FAT1*, *KMT2D*, and *NSD1* across all *KDM3C* genotypes consistent with established HNSCC genomic landscape in which these are among commonly altered driver genes (52).

Beyond these conserved profiles, we identified a significant association between *KDM3C* status and *BRAF* alterations in rectal cancer (p= 0.0318), where the canonical V600E variant were restricted to the A/A WT-S464 tumors. Across *KDM3C* genotype groups in LARC, *KRAS* mutations were consistently dominated by codon G12 variants, particularly G12V, G12D, and G12C. Within this overall pattern, T/T SNP-S464T carriers showed a modest relative enrichment of G12D and a greater diversity of rare *KRAS* variants. *TP53* mutations were highly recurrent and consistently centered on canonical hotspots (R175, R248, R273, R282, G245) across all genotypes. Although descriptive differences in raw mutation counts were noted across *KDM3C* genotype groups—including a higher number of *SMAD4* truncating events in T/T SNP-S464T tumors, exclusive *CDKN2A* W110* mutations in carriers of the T allele tumors (both A/T, T/T), and modest increases in *NSD1*, *FAT1*, and *KMT2D* truncating alterations or *PIK3CA* hotspots among T/T SNP-S464T tumors—these patterns were observed in small, imbalanced genotype groups. Therefore, these somatic alteration patterns are considered hypothesis-generating and do not fully account for the observed association between *KDM3C* genotype.

### KDM3C SNP-S464T enhances cytotoxic response to DNA-damaging therapies across patient-derived cellular models

To determine whether the *KDM3C* SNP-S464T alters response to DNA damaging agents, we tested its impact across patient-derived and isogenic cellular systems. We first evaluated lymphoblastoid cell lines generated from the peripheral blood monocytes of a subset of LARC patients analyzed above (n=12) and exposed these cell lines to irradiation (IR), 5-fluorouracil (5-FU), or oxaliplatin. Across all agents tested, KDM3C SNP-S464T carrying lymphoblastoid lines exhibited a significantly higher percent of cell killing compared with KDM3C WT-S464 lymphoblastoid lines (**Figure 2A-C**). Patient-derived rectal tumor organoids were genotyped and assessed for response to LARC therapies, such as 5-FU, FOLFOX (leucovorin, 5-FU, oxaliplatin) or FOLFIRI (leucovorin, 5-FU, irinotecan). Rectal tumoroids harboring the SNP-S464T consistently exhibited lower IC_50_ values for 5-FU, FOLFOX or FOLFIRI compared with organoids with WT-S464 (A/A) or heterozygous A/T genotypes (**Figure 2D**), mirroring the observations in rectal cancer patient-derived lymphoblastoid cell lines.

**Figure 2.**
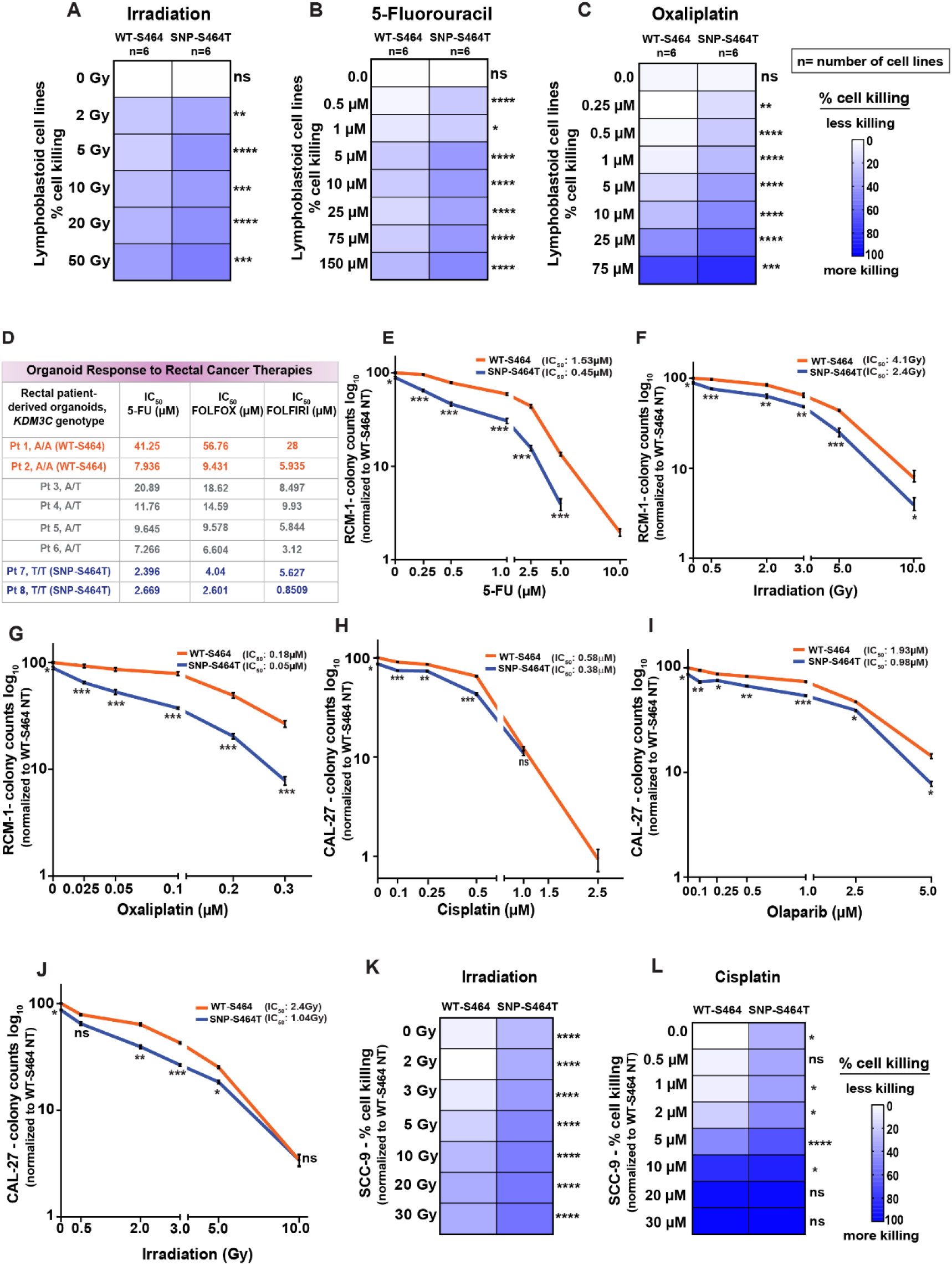
KDM3C SNP-S464T enhances cellular sensitivity to DNA-damaging therapies across patient-derived models. Percent cell killing in lymphoblastoid lines from LARC patients with KDM3C WT-S464 (n=6) or SNP-S464T (n=6) genotypes. Cells were left untreated or treated with **(A)** irradiation, **(B)** 5-fluorouracil, or **(C)** oxaliplatin. Viability was measured 72 h after treatment using CellTiter-Glo assay and plotted as percent cell killing. n=3, *p<0.05, **p<0.01, ***p<0.001, ****p<0.0001, Mann-Whitney. **(D)** IC_50_ values for 5-FU, FOLFOX, and FOLFIRI in rectal cancer patient-derived organoids, stratified by *KDM3C* genotype. Clonogenic survival of RCM-1 isogenic cells (SNP-S464T vs. WT-S464) following **(E)** 5-fluorouracil, **(F)** irradiation, or **(G)** oxaliplatin. Clonogenic survival of CAL-27 isogenic cells (SNP-S464T vs. WT-S464) following **(H)** cisplatin, **(I)** olaparib, or **(J)** irradiation. For all clonogenic assays, isogenic lines were plated and then treated with the indicated DNA-damaging agents. Colonies were stained with crystal violet, and colony counts were normalized to untreated WT-S464 controls and plotted on a log_10_ scale. Error bars=SEM; n=3 biological replicates, ns= non-significant; *p<0.05, **p<0.01, ***p<0.001; unpaired two-tailed t-test with Welch’s correction. Percent cell killing in SCC-9 (SNP-S464T vs. WT-S464) isogenic cells following **(K)** irradiation or **(L)** cisplatin. Cell viability was assessed using CellTiter-Glo assay 72h post-treatment and plotted as percent cell killing. n=3 biological replicates, ns= non-significant; *p<0.05, **p<0.01, ***p<0.001; unpaired two-tailed t-test with Welch’s correction.

We next used CRISPR/Cas9-editing to generate several isogenic SNP-S464T or WT-S464 cancer cell lines. Using these models, we performed clonogenic survival assays following treatment with DNA-damaging agents commonly used in the treatment of LARC and HNSCC. RCM-1 rectal cancer SNP-S464T cells demonstrated markedly reduced colony survival following treatment with 5-FU, IR, or oxaliplatin compared with WT-S464 cells, consistent with increased sensitivity to agents used in LARC treatment (**Figure 2E-G**). CAL-27 HNSCC SNP-S464T cells showed significantly reduced colony survival compared to the WT-S464 cells upon treatment with cisplatin, olaparib, or IR (**Figures 2H-J**) indicating increased sensitivity to agents used in HNSCC treatment. Although SCC-9 HNSCC WT-S464 and SNP-S464T cells did not form quantifiable colonies, SNP-S464T cells showed significantly elevated cell killing following IR or cisplatin treatment compared to WT-S464 cells (**Figure 2K-L**).

### KDM3C SNP-S464T disrupts MDC1–RNF8–RAP80-BRCA1 pathway assembly

The scaffolding protein MDC1 is a central mediator of DNA repair, binding γH2AX at the site of DSBs, and recruiting downstream repair factors. Previous studies have suggested that KDM3C regulates MDC1-dependent DDR signaling and downstream RNF8-RAP80-BRCA1-mediated repair complex assembly (45). Given this role, we tested whether the SNP-S464T influenced this DNA repair axis. Notably, S464 amino acid site lies outside the JmjC catalytic domain of KDM3C. Therefore, we reasoned that p.S464T could influence protein–protein interactions or the recruitment of KDM3C within DNA damage signaling complexes. To test this possibility, we first examined engagement between KDM3C and MDC1 following DNA damage.

Co-immunoprecipitation (Co-IP) of endogenous proteins from the CAL-27 HNSCC isogenic model revealed significantly attenuated KDM3C-MDC1 interaction in KDM3C SNP-S464T cells compared with WT-S464 cells following IR (**Figure 3A, top**). Minimal KDM3C-MDC1 interactions were observed in the absence of IR. Minimal KDM3C-MDC1 interactions were observed in the absence of IR. No significant changes in total KDM3C or MDC1 protein levels were observed following IR (**Figure 3A, bottom**), indicating no effect on KDM3C or MDC1 protein abundance.

**Figure 3.**
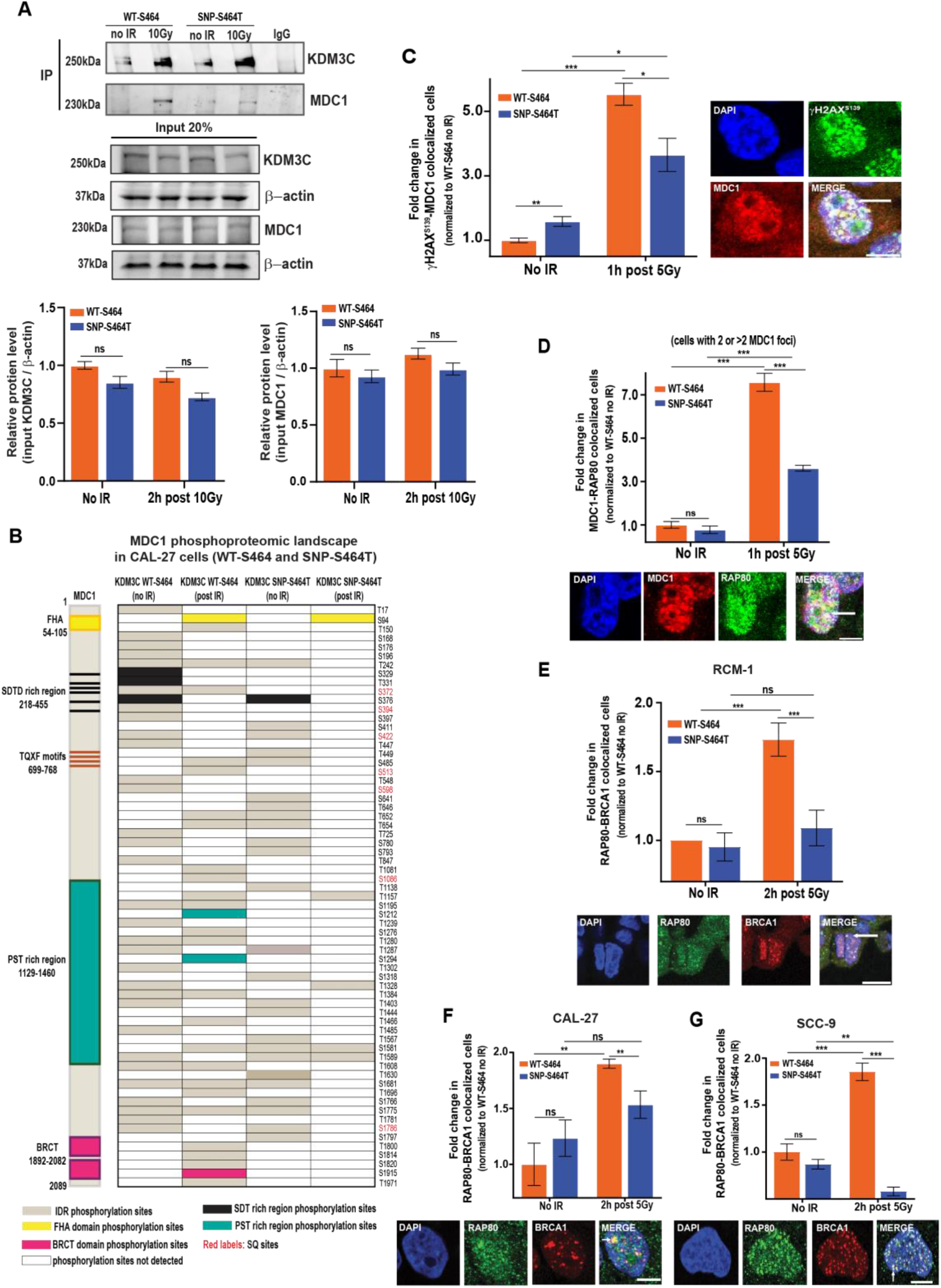
KDM3C SNP-S464T disrupts assembly of the MDC1-RNF8-RAP80-BRCA1 repair pathway. **(A) Top:** Isogenic *KDM3C* SNP/WT cells were not treated or irradiated (10 Gy), and lysates were collected 2h post IR. Co-IP was performed using KDM3C as bait, followed by immunoblotting for KDM3C and MDC1. Input controls and IgG pulldowns confirm specificity **Bottom:** Immunoblot shows no changes in MDC1 and KDM3C protein expression in Cal-27 cells bearing SNP-S464T vs. WT-S464. Densitometry analysis of MDC1 and KDM3C normalized to β-actin and shown as relative protein levels. **(B)** Schematic of MDC1 domain architecture and phosphorylation mapping in CAL-27 bearing KDM3C WT-S464 and SNP-S464T cells. Linear stick diagram of MDC1 highlighting the FHA domain (yellow), SDTD or Ser-Asp-Thr-Asp repeats motifs (black), TQXF motifs (brown), PST or Pro-Ser-Thr rich region (teal), and the tandem BRCT domain (pink). Heatmap showing MDC1 phosphorylation sites identified by nanoLC/MS in CAL-27 cells expressing KDM3C WT-S464 or SNP-S464T at baseline (or no IR) and post IR. Each column represents an individual phosphorylation site. Colored boxes denote detected phosphorylation events, with light brown indicating phosphorylation within IDR, yellow indicating phosphorylation within the FHA region, teal indicating phosphorylation within the PST region, and pink indicating phosphorylation within the BRCT region; white indicates sites not detected. Sites labeled in red are SQ motifs. **(C) Top:** Reduced γH2AX^S139^-MDC1 colocalization was observed post IR in CAL-27 isogenic cancer models bearing KDM3C SNP-S464T vs. WT-S464. **Bottom**: representative γH2AX^S139^-MDC1 colocalization image are shown. **(D) Top:** Reduced MDC1-RAP80 colocalization was observed post IR in CAL-27 isogenic cancer models bearing KDM3C SNP-S464T vs. WT-S464. **Bottom**: representative MDC1-RAP80 colocalization images are shown. Reduced RAP80-BRCA1 colocalization was observed post IR in **Top: (E)** RCM-1, **(F)** CAL-27, and **(G)** SCC-9 isogenic cancer models bearing KDM3C SNP-S464T vs. WT-S464. **Bottom**: representative RAP80-BRCA1 colocalization image are shown. **Quantification:** Colocalization values were normalized to the corresponding WT-S464 no IR condition within each isogenic cell-line pair. Scale bar: 5μm, Error bars=SEM; n=3 biological replicates, ns= non-significant; *p<0.05, **p<0.01, ***p<0.001; unpaired two-tailed t-test with Welch’s correction. All images were captured using Leica TCS Advanced SP8 Confocal microscope (Leica Microsystems, Wetzlar, Germany).

MDC1 is extensively regulated by phosphorylation upon DNA damage (53), with modifications occurring across distinct functional regions of the protein, including ATM/ATR-responsive SQ motifs (54). Since phosphorylation of MDC1 is required for the downstream signaling that promotes BRCA1 pathway assembly (55), we tested whether the attenuated KDM3C-MDC1 engagement observed in KDM3C SNP-S464T cells impacts DNA damage–responsive phospho-signaling of MDC1. To address this, we performed Co-IP/mass spectrometry of KDM3C–MDC1 complexes from the isogenic cell lines (WT-S464 and SNP-S464T, CAL-27) at baseline and following IR to study impact on MDC1 phosphorylation (**Figure 3B; Supplementary Figure 2; Supplementary Table 14**).

In KDM3C WT-S464 cells, multiple phosphorylation sites were detected across functional regions of MDC1 at baseline, including residues within the N-terminal intrinsically disordered region (IDR) and SDTD repeat motifs involved in interactions with DNA damage signaling factors. Following IR, WT-S464 cells exhibited increased phosphorylation at several established DNA damage–responsive sites, including S94 within the FHA (Forkhead-associated) domain, ATR-associated phosphosite such as S513 and the SQ motif site S1086, indicating robust DNA damage–responsive MDC1 phosphorylation. Additional phosphosites were identified in the SDTD-rich region, including T331 and S376, which are annotated as CK2A1-associated sites, and T847, which is annotated as a PLK1-associated site.

In contrast, KDM3C SNP-S464T cells showed a restricted MDC1 phosphorylation profile at baseline and a substantially attenuated phosphorylation profile following IR. Phosphorylation events were largely confined to residues within MDC1 IDR regions, with limited detection of phosphorylation at SDTD motifs and SQ motifs associated with downstream signaling (**Figure 3B; Supplementary Figure 2; Supplementary Table 14**). Together, these results suggest that weakened KDM3C-MDC1 interaction SNP-S464T cells may compromise the efficiency or extent of downstream MDC1 phospho-signaling events.

We also examined the functional consequence of the SNP-S464T on the assembly of proteins required for DNA damage response. Since MDC1 recruitment to sites of DNA damage depends on phosphorylation-dependent binding of the MDC1 BRCT domain to γH2AX (56), we quantified γH2AX-MDC1 colocalization. At baseline, SNP-S464T cells exhibited increased γH2AX-MDC1 colocalization relative to WT-S464 cells, suggesting elevated endogenous DNA damage and persistent retention of MDC1 at pre-existing lesions in SNP-S464T cells. Following IR, γH2AX-MDC1 colocalization increased markedly in WT-S464 cells, reflecting robust recruitment of MDC1 to newly induced DNA lesions. In contrast, SNP-S464T cells showed a significant decrease in γH2AX-MDC1 colocalization despite increased γH2AX induction at baseline (**Figure 3C**), indicating impaired damage-induced MDC1 recruitment or stabilization at sites of DNA damage. These findings are consistent with impaired activation of MDC1-dependent DNA damage response (DDR) signaling in SNP-S464T cells.

In published models of DSB repair, MDC1 recruitment precedes RAP80 accumulation, and perturbations disrupting upstream interactions affect downstream assembly of repair complexes (57). Because the KDM3C– MDC1 interaction is attenuated, we assessed whether this impacts RAP80 recruitment by quantifying MDC1-RAP80 colocalization. MDC1-RAP80 colocalization was comparable between CAL-27 WT-S464 and SNP-S464T cells at baseline. Following IR, WT-S464 cells showed a marked increase in MDC1-RAP80 colocalization, whereas this damage-induced colocalization was significantly attenuated in SNP-S464T cells (**Figure 3D**). These data are consistent with impaired MDC1 interactions after IR.

Since RAP80-mediated recruitment of BRCA1 represents a critical downstream output of MDC1–RNF8 ubiquitin signaling (58), we then quantified RAP80-BRCA1 colocalization following IR across three isogenic models (RCM-1, CAL-27, and SCC-9). In all models, *KDM3C* SNP-S464T cells displayed significantly reduced RAP80-BRCA1 colocalized foci after DNA damage compared with WT-S464 cells (**Figure 3E-G**), suggesting defective BRCA1 pathway assembly downstream of MDC1-RAP80.

Consistent with these findings, analysis of TCGA rectal tumors revealed a significant enrichment of the homologous recombination (HR) deficiency–associated SBS3 mutational signature (59) in T/T homozygous *KDM3C* SNP-S464T carriers compared with A/T heterozygotes and A/A wildtype (WT-S464) tumors (**Supplementary Figure 3A**). In HNSCC, although the SBS3 signature was detectable across all genotypes, it was significantly enriched in tumors harboring the *KDM3C* T/T genotype (**Supplementary Figure 3B**).

### The highly conserved S464 residue localizes to an intrinsically disordered region associated with BRCA1 repair

To gain structural insight into how the SNP-S464T might impair pathway assembly, we modeled the KDM3C protein architecture using AlphaFold2 (AF2) (**Figure 4A**); a more detailed analysis of the structure features is provided in the **Supplementary Results**. AF2 predicted three structured modules in KDM3C: a previously unannotated complex of intertwined SH3-like β-barrel domains at the N-terminus (residues 1-254); a central zinc-binding regulatory region (residues 1694–1938), including a C6-type zinc-finger module conserved across KDM3 family members (60); and a C-terminal catalytic JmjC domain (residues 2150-2540). During this work, a novel arrangement of three SH3-like domains, labeled as ‘SH3-like domain’, was found upstream of the SNP containing region. These predicted SH3-like domains may facilitate protein–protein interactions (61) and complex assembly, consistent with a potential scaffolding role in DDR signaling.

**Figure 4.**
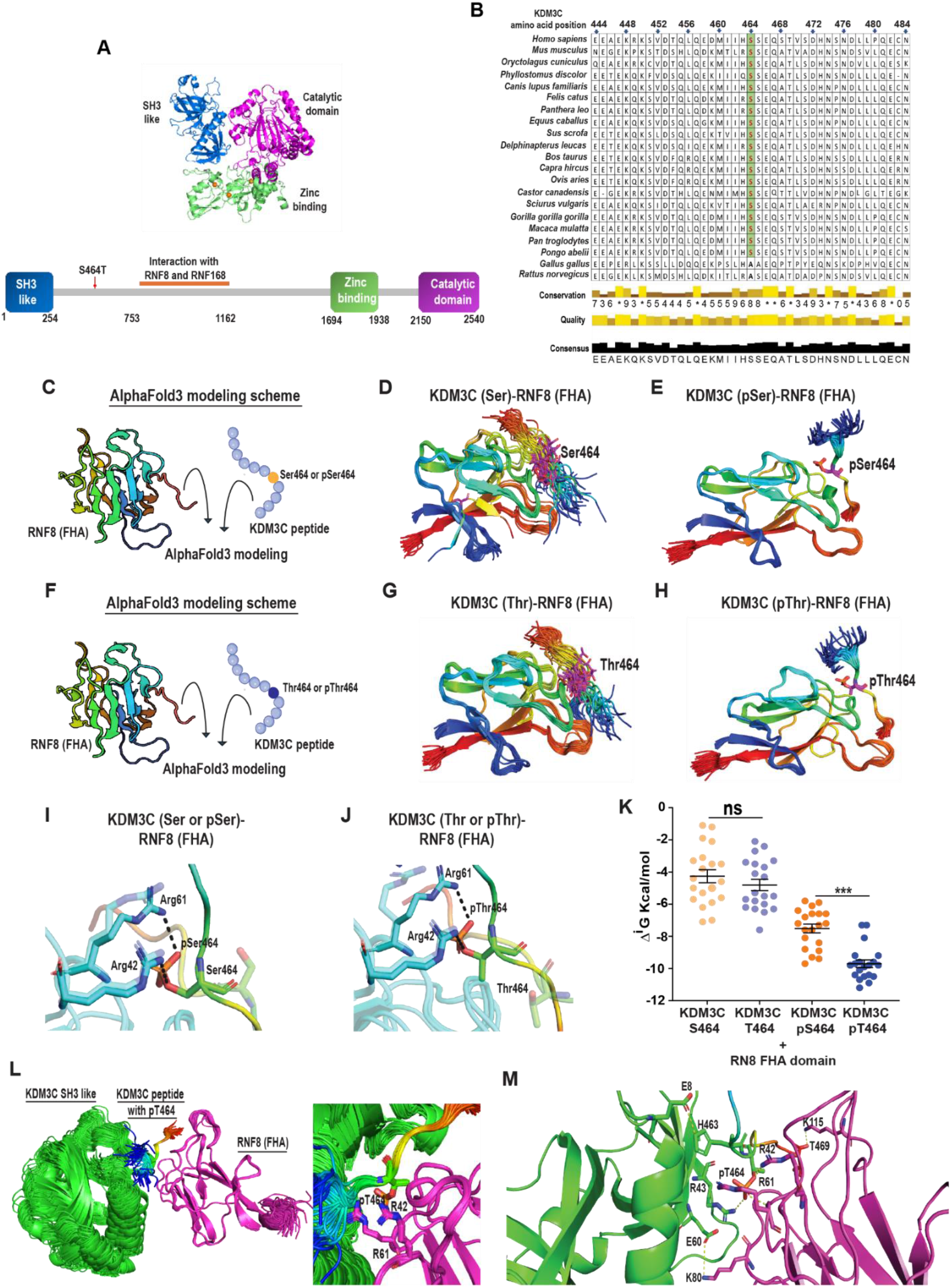
Phospho-threonine promotes significantly more stable interaction between KDM3C and RNF8 FHA complex in AlphaFold3 models. **(A) Top:** AF2 structural model of full-length KDM3C showing three predicted folded modules: an N-terminal SH3-like β-barrel domain (blue), a central zinc-binding regulatory module (green), and a C-terminal JmjC catalytic domain (purple). **Bottom:** folded modules of KDM3C (blue, green, purple), *KDM3C* SNP and putative region of interactions with ubiquitin ligases (RNF8, RNF168) are shown. **(B)** The sequence alignment surrounding S464 is shown for multiple species, showing high conservation of the S464 residue. Sequences for KDM3C protein of 21 different organisms were selected and retrieved from UniProt database. Alignment was done using the Clustal Omega server and visualized using Jalview software. Jalview automatically calculates quantitative alignment annotations such as conservation, quality, and consensus scores. These annotations are displayed as histograms below the multiple sequence alignment. Conservation annotation shown in numerical value (scale of 1 to 9 with 9 being most conserved) as well as histogram portrayal. The columns which have identical residues are indicated by *. The quality annotation shows the chances of observing mutations whereas consensus annotation depicts the percentage of residues in a specific column of alignment. **(C)** Schematic representation of AlphaFold3 modeling of KDM3C peptides (S or pS) in complex with the RNF8 FHA domain. Created in BioRender. Adria Hasan, F. (2026) https://BioRender.com/hxkm487. Superposition of 50 models of RNF8-FHA domain in complex with KDM3C peptide respectively bearing **(D)** serine, **(E)** phosphoserine. **(F)** Schematic representation of AlphaFold3 modeling of KDM3C peptides (T or pT in complex with the RNF8 FHA domain. Created in BioRender. Adria Hasan, F. (2026) https://BioRender.com/hxkm487. Superposition of 50 models of RNF8-FHA domain in complex with KDM3C peptide respectively bearing **(G)** threonine, and **(H)** phospho-threonine variant. Note that in panels D, E, G, and H, the rainbow coloring of peptides from N-terminal blue to C-terminal red. **(I, J)** Superimposition of the top-ranking complex of RNF8 FHA domain (Cyan) with KDM3C peptide bearing pS/pT (side chains rendered darker) shows formation of H-bonds with interacting amino acids while no H-bond is formed when S/T is not phosphorylated (side chains rendered lighter). **(K)** Average solvation free energy gain (ΔiG) of top 20 models of RNF8-FHA in complex with WT (S464) and SNP (T464) variant of KDM3C peptide with and without phosphorylation obtained using PISA server. **(L)** Superimposition of 50 models of RNF8-FHA domain with KDM3C pT peptide and KDM3C SH3-like domain. Note that in panel L the rainbow coloring of peptides from N-terminal blue to C-terminal red. **(M)** Enlarged view showing key residues at the RNF8-FHA interface with KDM3C pT464 and the SH3-like domain.

The remaining regions of KDM3C, including the SNP-containing segment, were predicted to be intrinsically disordered. The broader N-terminal region of KDM3C spanning amino acids 1-753, which includes the SNP-S464T-containing segment, contains multiple SQ motifs that may represent potential ATM/ATR phosphorylation sites during DDR signaling. Sequence analysis of the KDM3C protein sequence (UniProtKB entry <u>Q15652</u>) identified SQ motifs at residues 96-97, 291-292, 430-431, 665-666, and 734-735 (45,62). Prior work has shown that the region of KDM3C spanning amino acids 753–1162 interacts with the E3 ubiquitin ligases RNF8 and RNF168 (45). Although this region is distal to the S464 residue, these findings support a broader role for KDM3C in DNA repair signaling and suggest that additional regions, including the SNP-containing region, may modulate repair-complex assembly. Despite its IDR features, evolutionary analysis demonstrated that both the SNP-containing region surrounding S464, and the S464 residue itself are highly conserved KDM3C across mammals (**Figure 4B**). Comparative analysis of the KDM3 family further indicated that KDM3C contains a larger IDR region (**Supplementary Figure 4**) that could contain functional roles not shared by KDM3A or KDM3B IDRs. Since there are no extended regions of significant homology/similarity when comparing the three IDRs of KDM3A, KDM3B, and KDM3C, this larger KDM3C IDR may support functional roles that are not shared by the KDM3A/B paralogs.

### AlphaFold3 modeling predicts potential phospho-dependent interaction between KDM3C and the RNF8 FHA domain

To explore a potential structural basis for the impact of the KDM3C SNP-S464T on DDR signaling, we first performed AF2 modeling of 79 KDM3C-RNF8/MDC1 domain pairs using information from the ECOD database (63). We utilized both AlphaFold’s ipTM score as well as a recently developed score, ipSAE (interaction predicted Score from Aligned Errors) (64) that fixes known artifacts in the ipTM score. Here, 14 domain pairs yielded models with an iPTM (interface predicted template modeling score) ≥ 0.6 (**Supplementary Table 15)**. The strongest predicted interaction was between the KDM3C SH3-like domain (aa 6-180) and the RNF8 SMAD/FHA domain (aa 16-135), with 41 of 50 models meeting both the ipTM ≥ 0.6 and ipSAE score ≥ 0.4 thresholds. Additional RNF8-associated models included predicted interactions between the RNF8 SMAD/FHA domain (aa 16-135) and the KDM3C catalytic/double-stranded β-helix domain (2156–2495), as well as KDM3C inter-domain regions spanning aa 256–1695 and aa 1926–2155.

Therefore, we performed a focused, hypothesis-driven AlphaFold3 (AF3) analysis using KDM3C-derived peptides containing either S464 (WT) or T464 (SNP) in complex with the RNF8 FHA domain (**Figure 4C-H**). Because FHA domains are phospho-peptide-binding domains with a known preference for phospho-threonine over phospho-serine containing motifs (65), we modeled both non-phosphorylated and hypothetical phosphorylated states of the residue 464-centered peptide to assess whether residue identity and phosphorylation state could influence peptide positioning within the FHA binding interface. AF3 generated 50 models for each peptide state. The peptide comprised 11 residues around the S464 or T464 site at the center and was modeled in both phosphorylated and non-phosphorylated forms.

Superimposition of the RNF8-FHA domain with S464- (**Figure 4C-E**) and T464- (**Figure 4F-H**) containing peptides revealed that phosphorylation significantly enhanced peptide alignment and structural consistency. Phosphorylated peptides (pS464 and pT464) showed tight superimposition with consistent side-chain orientations, while non-phosphorylated forms exhibited marked variability. Notably, phosphorylation induced a distinct flip in peptide orientation (rainbow coloring of peptides from N-terminal blue to C-terminal red) and resulted in high uniformity of peptide position among the 50 models, predicting a conformational switch that may impact FHA domain binding. Structural analysis showed that pS464 and pT464 formed stable hydrogen bonds with key RNF8-FHA residues R42 and R61 (**Figure 4I-J**). These interactions were absent in the non-phosphorylated peptides, although variable hydrophobic contacts persisted. Consistent with this, binding free energy (ΔG) calculations using the PISA server (66) predicted more favorable interactions for the modeled phosphorylated peptides, with the pT464 peptide showing the strongest predicted binding affinity among the states examined, followed by pS464, T464, and S464 (**Figure 4K**). Non-phosphorylated peptides also showed broader ΔG distributions, suggesting higher conformational flexibility and lower structural stability. Furthermore, computational experiments showed evidence that both the pT464 region and the SH3-like region of KDM3C could cooperate to bind the RNF8-FHA domain. All 50 models of RNF8-FHA and pT464-KDM3C peptide pair in complex with the SH3 module had ipSAE score above 0.6. For non-phosphorylated complexes, no interacting interface between KDM3C peptide and RNF8 FHA was observed as reflected by very low iPTM scores (∼0.2) and ipSAE scores of zero. High confidence complexes were formed only when KDM3C pT464 was in the interface (**Figure 4L-M**).

These modeling results suggest that if the 464 amino acid site in KDM3C undergoes phosphorylation, the p.S464T substitution could enhance RNF8-FHA domain engagement and alter the conformation or stability of KDM3C–RNF8 interactions. Specifically, an abnormally tight or prolonged engagement between the KDM3C pT464 and the RNF8-FHA domain may kinetically trap the complex, preventing the dynamic release or hand-off of repair factors required to properly assemble downstream components like RAP80 and BRCA1.

Next, *in silico* kinase motif modeling using NetPhos 3.1 (67,68) of the WT-S464 sequence (MIIHSSEQS) yielded no positive predictions for active kinase recognition at this position. Conversely, introducing the SNP-S464T (MIIHTSEQS) significantly enhanced the local phosphorylation potential, generating a positive prediction score for Protein Kinase C (PKC; score= 0.560) (**Supplementary Table 16)**. These data suggest that the SNP-S464T substitution creates a distinct neo-phosphorylation motif, potentially altering downstream protein assembly. Additional experimental studies will be needed to determine whether residue 464 is phosphorylated *in vivo* in response to DNA damage and to definitively validate the relevant upstream kinase(s).

### KDM3C SNP-S464T induces S-phase stalling and γH2AX accumulation

We evaluated transcriptome-wide gene expression profiles to determine whether isogenic cell lineages harboring the KDM3C SNP-S464T versus WT-S464 converged on shared transcriptional programs. Given that KDM3C is a chromatin-remodeling histone demethylase implicated in transcriptional regulation (69,70) baseline comparisons identified hundreds of differentially expressed genes in RCM-1, CAL-27, and SCC-9 models (**Supplementary Table 17**). This heterogeneity indicates that the shared impact on response to DNA-damaging therapy is likely not driven by a uniform downstream transcriptional signature.

Individual gene-level overlap was modest; however, *DAPK1* which encodes death associated protein kinase 1 (71), a positive mediator of apoptosis and autophagy with known tumor suppressor function (71,72) was consistently downregulated in all KDM3C SNP-S464T cell lines. On the other hand, *CXCL8*, which encodes pro-inflammatory chemokine IL-8, and promotes neutrophil recruitment and activation, was consistently upregulated in all KMD3C SNP-S464T cells (73,74) (**Supplementary Figure 5)**.

Building on the impaired BRCA1 pathway assembly downstream of MDC1-RAP80 observed above, we assessed cell-cycle progression across the isogenic models. We observed prolonged S-phase accumulation across all KDM3C SNP-S464T models compared to WT-S464 models. In RCM-1 cells, the SNP-S464T cells exhibited a significantly elevated S-phase fraction at early time points following release from synchronization compared with WT-S464 cells (**Figure 5A**). Similarly, SCC-9 SNP-S464T cells showed sustained S-phase accumulation from 0-48 h after release compared with WT-S464 cells (**Figure 5B**). Consistent with these findings, the CAL-27 SNP-S464T cells displayed delayed exit from S phase under basal conditions and persistent S-phase accumulation following cisplatin exposure (**Supplementary Figure 6A-B**), indicating impaired S-phase progression that may reflect altered DDR signaling.

**Figure 5.**
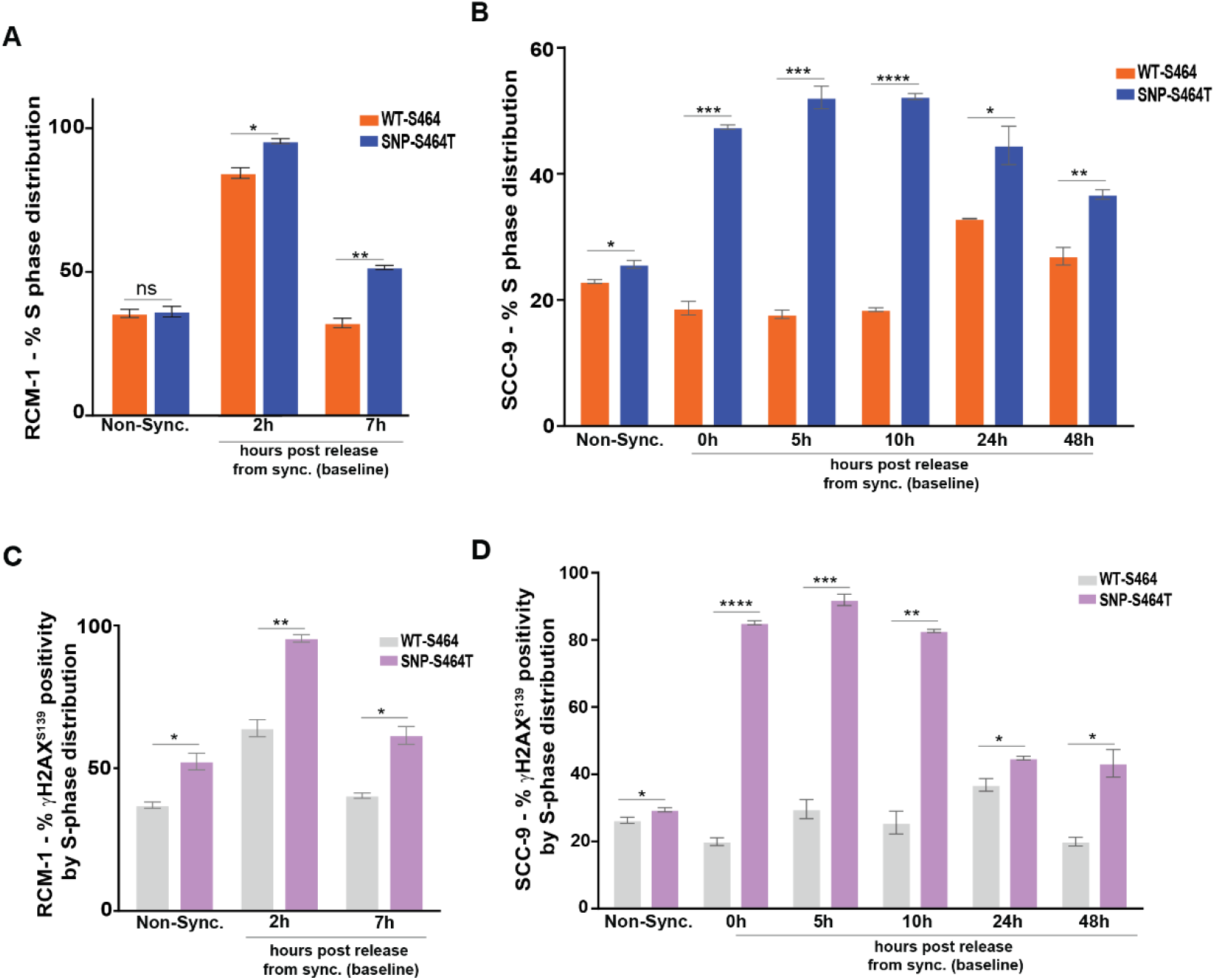
KDM3C SNP-S464T induces S-phase stalling and γH2AX accumulation. Cell-cycle distribution following synchronization and release under baseline, non-irradiated conditions in **(A)** RCM-1 and **(B)** SCC-9 isogenic cells. SNP-S464T cells showed delayed S-phase exit compared with WT-S464 cells. γH2AX^S139^ positivity within the S-phase population following synchronization and release in **(C)** RCM-1 and **(D)** SCC-9 cells show increased γH2AX^S139^ positivity within S phase. Error bars=SEM; n=3 biological replicates, ns= non-significant; *p<0.05, **p<0.01, ***p<0.001; unpaired two-tailed t-test with Welch’s correction.

To determine whether S-phase stalling was accompanied by increased endogenous DNA damage, we quantified γH2AX^S139^ under baseline, non-IR synchronization-release conditions. Although both genotypes accumulated γH2AX ^S139^ during S phase, WT-S464 cells efficiently resolved γH2AX ^S139^ over time, whereas SNP-S464T cells exhibited persistent γH2AX ^S139^ signal, indicating sustained replication-associated DNA damage (**Figure 5C-D**).

### KDM3C SNP-S464T enhances cGAS-cGAMP-STING signaling and increases apoptotic cell death after DNA damage

Given that pro-inflammatory *CXCL8* was consistently upregulated alongside other inflammatory mediators across the KDM3C SNP-S464T isogenic models, we next examined whether this inflammatory-response signature persisted following genotoxic stress. RNA-seq performed 24 hours after IR showed robust enrichment of interferon and inflammatory-response pathways in SNP-S464T HNSCC cells relative to WT-S464 control cells, including humoral immune-response pathways in CAL-27 and cytokine-activity signatures in SCC-9 cells (**Supplementary Figure 7A-B; Supplementary Table 18**).

Since cGAS–STING signaling is a well-established DNA damage–responsive pathway that can link cytosolic DNA to interferon and inflammatory gene induction (75), we tested whether this pathway was activated after IR. Because unresolved DNA damage can promote micronuclei formation and cytosolic DNA accumulation (75), we quantified cGAS-dsDNA colocalized micronuclei as an upstream readout of cGAS engagement with damage-derived DNA. In this pathway, cytoplasmic dsDNA binding activates cGAS and promotes production of cGAMP, which activates STING and downstream TBK1-dependent transcriptional programs that mediate interferon responses (76,77). Here, cGAS–dsDNA colocalization at baseline was similar between the WT and SNP groups; however, at 72 h after IR, SNP-S464T cells exhibited a significantly higher frequency of cGAS-positive micronuclei colocalized with cytosolic dsDNA compared with WT-S464 cells (**Figure 6A**). Consistent with this, cGAMP levels were significantly elevated in SCC-9 SNP-S464T cells at 72 h and 96 h after IR, while baseline levels were comparable between these groups (**Figure 6B**). In CAL-27 cells, although significant differences were not observed in cGAS–dsDNA colocalization following IR treatment (**Supplementary Figure 7C**), cGAMP remained significantly higher in SNP-S464T cells at later time points (72h and 96h) compared to WT-S464 cells post IR (**Figure 6C**).

**Figure 6.**
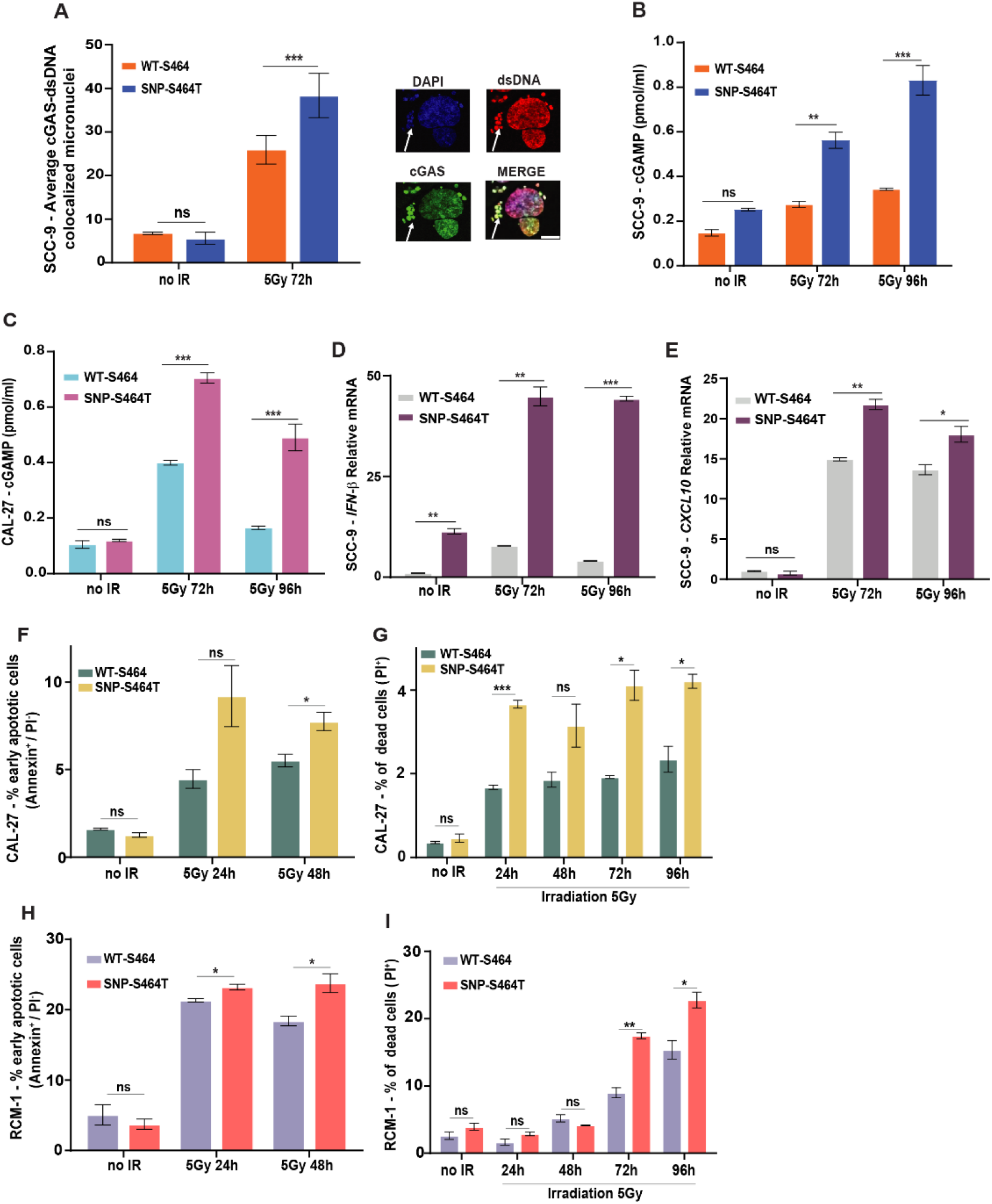
KDM3C SNP-S464T enhances cGAS-cGAMP activation and increases apoptosis following genotoxic stress. **(A)** Activation of the cGAS pathway in SCC-9 cancer cells carrying the KDM3C SNP-S464T post IR. **Top:** Increased colocalization in cGAS-dsDNA micronuclei in SCC-9 SNP-S464T cells at 72h, was observed post IR. **Bottom:** representative colocalization image using the Leica TCS Advanced SP8 Confocal microscope (Leica Microsystems, Wetzlar, Germany). **Quantification:** cells with cGAS-dsDNA colocalized foci. Scale bar: 5μm. **(B)** Increased cGAMP levels in SCC-9 SNP-S464T cells at 72h, and 96h post IR was observed. Protein was isolated from untreated or irradiated SCC-9 WT-S464 and SNP-S464T cells, and cGAMP activity was measured using ELISA. Error bars=SEM; n=3 biological replicates, ns= non-significant; *p<0.05, **p<0.01, ***p<0.001; unpaired two-tailed t-test with Welch’s correction. **(C)** Increased cGAMP levels in CAL-27 SNP-S464T cells at 72h, and 96h post IR was observed. Error bars=SEM; n=2 biological replicates, ns= non-significant; ***p<0.001; unpaired two-tailed t-test with Welch’s correction. Increase in relative mRNA expression of **(D)** *IFN-β*, and **(E)** *CXCL10* in SCC-9 SNP-S464T cells vs. WT-S464 after DNA damage. Total RNA was isolated after IR and mRNA expression of the target genes was measured by qRT-PCR. Error bars=SEM; n=3 biological replicates, ns= non-significant; *p<0.05, **p<0.01, ***p<0.001; unpaired two-tailed t-test with Welch’s correction. Increase in IR induced **(F)** % positive early apoptotic cells and **(G)** % PI^+^ dead cells were observed in CAL-27 cells bearing SNP-S464T vs. WT-S464. Similarly, increase in IR induced **(H)** % positive early apoptotic cells and **(I)** % PI^+^ dead cells were observed in RCM-1 cells bearing SNP-S464T vs. WT-S464. The isogenic cell lines were exposed to 5Gy IR and analyzed by Annexin V/PI staining. Error bars=SEM; n=3 biological replicates, ns= non-significant; *p<0.05, **p<0.01, ***p<0.001; unpaired two-tailed t-test with Welch’s correction.

Downstream SCC-9 SNP-S464T cells maintained significantly elevated levels of active, S172-phosphorylated TBK1 at 72–96 h after IR, relative to WT-S464 cells (**Supplementary Figure 7D**). Correspondingly, *IFN-β* and *CXCL10* mRNA levels were significantly increased in SCC-9 SNP-S464T cells at these time points (**Figure 6D-E**), consistent with sustained activation of the cGAS–STING–type I interferon axis.

We next assessed apoptotic outcomes following DNA damage. In CAL-27 cells exposed to 5 Gy IR, SNP-S464T cells exhibited a significantly higher proportion of early apoptotic cells (Annexin⁺/PI⁻) at 24–48 h compared with WT-S464T cells (**Figure 6F**), along with increased accumulation of PI⁺ dead cells from 24–96 h (**Figure 6G**). Similarly, in RCM-1 cells exposed to 5 Gy IR, SNP-S464T cells exhibited a significantly higher proportion of early apoptotic cells (Annexin⁺/PI⁻) at 24–48 h compared with WT-S464 cells (**Figure 6H**), along with increased accumulation of PI⁺ dead cells from 24–96 h (**Figure 6I**). These results suggest that while the WT-S464 cells initiated apoptosis upon IR treatment, they progressed slowly into terminal stages of cell death, whereas SNP-S464T cells significantly continued to accumulate PI⁺ cells over time, consistent with impaired repair and persistent damage signaling. Similar patterns were observed following cisplatin treatment in CAL-27 WT S464 and SNP-S464T cells (**Supplementary Figure 7E-F**).

Together, these results suggest that defective resolution of DNA damage in KDM3C SNP-S464T cells leads to accumulation of cytosolic DNA and persistent activation of the cGAS–STING–interferon signaling axis, thereby enhancing inflammatory signaling and apoptotic sensitivity following genotoxic stress.

### KDM3C SNP-S464T is associated with increased cancer incidence in population cohorts

Given that defective repair can promote genome instability and cancer development, we assessed whether the SNP-S464T was associated with cancer incidence. In the UK Biobank cohort (n= 501,946), SNP-S464T carriers (T/T; n= 281,763) showed a modest but significant increase in overall cancer incidence relative to WT-S464 carriers (A/A; n= 59,108), with the strongest enrichments observed for skin cancer and cervical carcinoma *in situ*. Relative-risk analysis showed increased overall cancer risk among individuals younger than 45 years (p= 0.029), increased skin cancer risk in both younger and older age groups (p= 0.017 and p< 0.001, respectively) and increased cervical carcinoma *in situ* risk among individuals younger than 45 years (p= 0.031) (**Figure 7A**).

**Figure 7.**
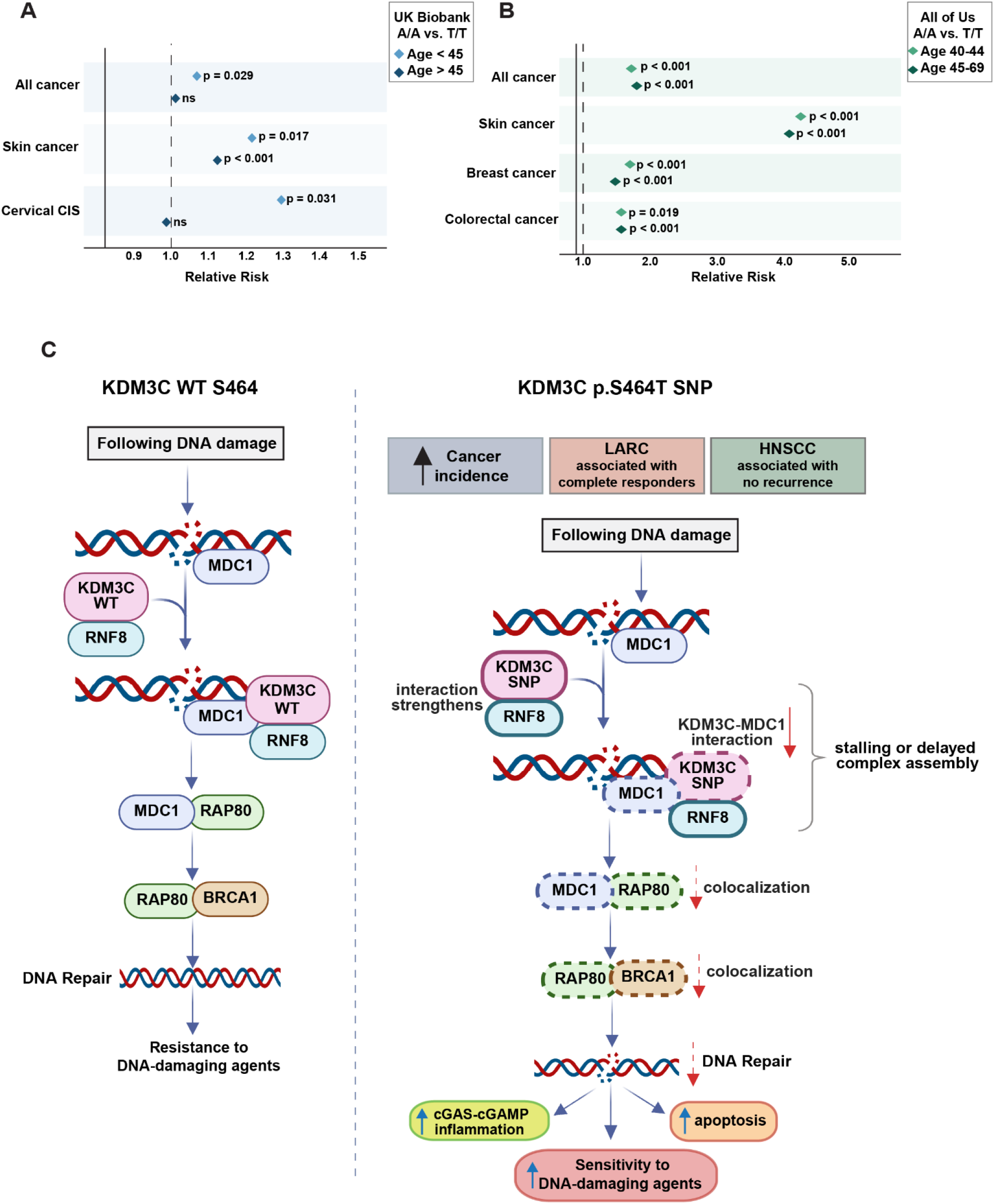
Relative risk of cancer by subtype in homozygous KDM3C SNP carriers (T/T; reference group: WT:A/A), stratified by age. **(A)** In the UK Biobank cohort, homozygous T/T carriers (SNP-S464T) showed significant increase in overall cancer incidence, with the strongest enrichment observed for skin cancer and cervical carcinoma *in situ* (CIS). Horizontal bars represent the RR of cancer diagnosis in homozygous T/T carriers across cancer types and age strata. The dashed vertical line indicates the baseline risk ratio (RR = 1.0). **(B)** In the All of Us cohort, homozygous *T/T* carriers (SNP-S464T) showed significant associations across multiple cancer subtypes and age groups, with skin cancer showing the strongest effect in both age groups. Significant associations were also observed for breast cancer and colorectal cancer. Horizontal bars represent the RR of cancer diagnosis in homozygous T/T carriers (SNP-S464T) across cancer types and age strata. The dashed vertical line indicates the baseline relative risk (RR = 1.0). Statistical analysis was performed using crude/unadjusted association tests (OR/RR) based on 2 × 2 contingency tables. **(C)** Schematic illustrating the proposed model for KDM3C SNP-S464T mediated disruption of DNA damage-response complex assembly. Created in BioRender. Adria Hasan, F. (2026) https://BioRender.com/rbapl4q.

We then evaluated the All of Us cohort (n= 414,814 with genotype data), in which SNP-S464T carriers (T/T; n= 180,159) similarly showed increased cancer incidence relative to WT-S464 carriers (A/A; n= 69,774). Relative-risk analysis in the All of Us cohort showed increased skin cancer risk in both age groups (40-44 years and 45-69 years; both p< 0.001). Increased overall cancer risk and breast cancer risk were also observed in both age groups (all p< 0.001). Colorectal cancer risk was similarly increased among individuals aged 40-44 years (p= 0.019) and 45-69 years (p< 0.001) (**Figure 7B**). Together, these independent cohort analyses indicate that the SNP-S464T is associated with increased cancer incidence, with the most consistent association observed for skin cancer across two population cohorts.

These findings support a model in which the KDM3C SNP-S464T disrupts complex assembly required for DNA repair, contributing to cancer risk and therapeutic vulnerability (**Figure 7C**). Our results highlight this mechanism, though additional downstream or parallel effects remain to be explored.

## DISCUSSION

In this study, we identify a conserved germline coding missense SNP-S464T in *KDM3C* that is associated with altered BRCA1-associated repair-complex assembly, and improved CRT outcomes in patients with LARC and LA-HNSCC. Across independent clinical cohorts, the SNP-S464T was enriched in individuals with better outcomes, with expanded genotyping supporting a robust association and population data indicating that this common polymorphism is associated with ancestry-dependent frequency. Mechanistically, SNP-S464T impaired damage-induced KDM3C-MDC1 interaction and attenuated damage-induced colocalization of MDC1, RAP80, and BRCA1 repair factors and reduced repair complex assembly. These molecular defects were accompanied by persistent γH2AX accumulation, delayed S-phase exit, increased sensitivity to IR and other DNA-damaging agents, and enhanced cell death following genotoxic stress. In HNSCC models, unresolved DNA damage was further associated with activation of the cGAS-cGAMP-STING pathway and induction of inflammatory-response programs, features consistent with chronic DNA damage (77–81).

Tumors harboring the KDM3C SNP-S464T showed distinct patient demographic stratification depending on the cancer context. Further, TCGA analyses in rectal and HNSCC cohorts revealed a broadly conserved somatic driver landscape across *KDM3C* genotypes. The modest differences observed in specific mutation frequencies, such as alterations in *BRAF*, *KRAS*, or *SMAD4*, were restricted to small genotype groups and are likely hypothesis-generating. Rectal and head and neck tumors with the SNP allele were enriched for mutational signature, SBS3, which is typically observed in tumors with alterations in *BRCA1*, *BRCA2*, or *PALB2* (82–84). In large population cohorts, the SNP-S464T is associated with increased cancer incidence, particularly skin cancer. As skin cancers are strongly linked to UV-induced DNA damage and defective DNA repair (85), this association is consistent with a potential role for KDM3C in genome maintenance (45,86), although a direct role for KDM3C in skin cancer remains to be established.

Our findings suggest that the KDM3C SNP-S464T may modulate DNA damage through effects on damage-induced repair-complex assembly. This interpretation is also consistent with prior genome-wide screening data that identified KDM3C as a candidate regulator of HR, although its role in HR was not directly validated in that study (87).The SNP-S464T lies within a conserved region in the IDR of KDM3C. Our domain analysis identified a putative SH3-like domain near this region, suggesting potential protein–protein interaction capacity. Prior work has also mapped RNF8/RNF168 binding to of KDM3C (45), distal to S464. Together, these observations support a broader role for KDM3C in DNA repair signaling, although the specific contribution of the S464-containing region remains to be defined. IDRs are increasingly recognized as critical hubs for protein–protein interactions and are frequently subject to regulatory post-translational modifications (88,89). Consistent with this architecture, many core DDR proteins, including MDC1 (56), 53BP1 (90), BRCA1 (91,92), and RNF168 (93) contain extensive IDRs that facilitate flexible docking, phospho-dependent interactions, and ubiquitin-mediated signaling at sites of DNA damage. In isogenic models, we show that the SNP-S464T reduces KDM3C interaction with MDC1 and attenuates DNA damage-induced MDC1 phosphorylation signaling, including ATM/ATR-associated SQ/TQ responses and CK2-dependent SDT motifs critical for downstream signaling. This results in reduced MDC1–γH2AX and MDC1–RAP80 colocalization, ultimately impairing BRCA1-RAP80 (BRCA1-A) complex recruitment, potentially shifting repair pathway balance away from coordinated HR. Here, structural modeling suggests that substitution of serine with threonine at this position alters phospho-dependent interactions with the RNF8-FHA domain, providing a potential mechanistic basis for disrupted KDM3C-MDC1 complex formation and impaired downstream signaling. Additional experimental studies are needed to determine whether residue 464 is phosphorylated and to define the relevant upstream kinase(s).

In large population-based cohorts, the SNP-S464T is associated with increased cancer incidence, particularly skin cancer, consistent with UV-driven DNA damage and known links between HR/DSB repair capacity and skin cancer susceptibility (94–96). While not causal, these findings align with our mechanistic data showing impaired MDC1–RNF8–RAP80–BRCA1 pathway assembly, suggesting that altered KDM3C-dependent repair may promote genome instability while creating a vulnerability to DNA-damaging therapies. Clinically, the SNP-S464T is strongly associated with improved CRT outcomes in both LARC and HNSCC, extending prior observations that germline variation in canonical DDR genes (e.g., *BRCA1, BRCA2*, ATM) modulates treatment sensitivity (97). Our findings suggest that germline variation in chromatin regulators may similarly influence CRT response. Notably, a germline coding SNP in *KDM4A* was previously shown to alter protein turnover, enhance sensitivity to mTOR inhibitors and was associated with differential outcomes in lung cancer (98), establishing a precedent for lysine demethylase germline variation influencing both protein function and therapeutic response.

Together, these data identify KDM3C SNP-S464T as a germline modifier of DSB repair complex assembly, linking genome instability, cancer susceptibility, and treatment response. Larger prospective studies are needed to validate predictive utility and address potential confounding factors. Overall, our findings support consideration of chromatin-regulatory germline variants as response modifiers in precision oncology and reveal a previously unrecognized link between inherited chromatin-regulator variation and therapeutic DNA damage responses across tumor types.

## METHODS

### Study approval

The work in this study was performed under FCCC (Fox Chase Cancer Center) Institutional Review Board approved protocols, #18-4005, #19-4001 and #21-9921.

### Human specimens, cell lines and cell culture

De-identified cryopreserved peripheral blood mononuclear cell specimens from LARC and HNSCC patients used for this study were obtained from the FCCC Biosample Repository Facility (BRF). The FCCC BRF obtains informed consent and HIPAA authorization from all BRF participants for the use of de-identified specimens and associated clinical data prior to specimen collection (IRB# 11-866). Human rectal cancer cell line RCM-1 and HNSCC cancer cell lines (CAL-27 and SCC-9) were obtained from American Type Culture Collection (ATCC) and used for all experiments. All cell lines were authenticated for short tandem repeat (STR) genotyping and were routinely tested for *Mycoplasma*. RCM-1 cells were cultured in 1:1 RPMI-1640 and Ham’s F-12 media supplemented with 10% fetal bovine serum (FBS, HyClone, Logan, Utah), 2mM L-glutamine (Corning, Corning, NY), and 1× penicillin/streptomycin (10,000 units/ml and 10,000 ug/mL; Thermo Fisher Scientific, Waltham, MA). CAL-27 cells were cultured in Dulbecco’s Modified Eagles Medium (DMEM) media supplemented with 10% FBS, 4.5 g/L glucose; 4mM L-glutamine, 1mM sodium pyruvate, and 1x penicillin/streptomycin. SCC-9 cells were maintained in DMEM/Ham’s F-12 media supplemented with 10% FBS, 2.5mM L-glutamine; 15mM HEPES, 0.5Mm sodium pyruvate, 0.4µg/ml hydrocortisone, and 1× penicillin/streptomycin. All the cells were maintained in a humidified incubator with 5% CO_2_ at 37°C.

For Epstein-Barr virus (EBV) transformed lymphoblastoid cell lines (B-lines), PBMCs from LARC patients were collected and immortalized with EBV as previously described (99). EBV-transformed lymphoblastoid cell lines from LARC patients were genotyped and designated as KDM3C WT-S464 (n=6) and KDM3C SNP-S464T (n=6).

### CRISPR-Cas9 gene-editing

CRISPR-Cas9 Gene-Editing was performed by Synthego Corporation (Redwood City, CA, USA) for the generation of isogenic pairs. RCM-1 originally carries the homozygous WT allele (A/A) and was edited to generate isogenic pairs of homozygous SNP (T/T) and homozygous WT allele (A/A). SCC-9 originally carries the homozygous *KDM3C* SNP of interest, while Cal-27 carries the heterozygous A/T and these were edited to generate isogenic pairs of homozygous SNP (T/T) and homozygous WT allele (A/A). The single-guide RNA (sgRNA) sequence was AGAAUUAUGAUCAGAAACUG and the sgRNA cut location is chr10:63,214,759. The PCR primers used were forward primer 5’ACATCACTGACCCAGCTCTG3’ and reverse primer 5’AAGGCAGGAGAAGAGACCCT3’.

### Colony formation assay

Colony survival was assessed by clonogenic survival assay. Isogenic pairs of RCM-1 (1000 per well) or CAL-27 (100 cells per well) cells were seeded in a 6-well plate and incubated overnight at 37°C with 5% CO_2_. After overnight incubation, cells were treated with different doses of either irradiation (IR; 0.5 Gy to 10 Gy), cisplatin (0.1µM to 5µM, cat no. 25021-233-20, Sagent Pharmaceuticals, IL), olaparib (0.1µM to 5µM, cat no. SML3705, Sigma, St. Louis, MO), 5-flurouracil (0.25µM to 10µM, cat no. 16729-276-38, Accord Healthcare, Durham, NC), oxaliplatin (0.025µM to 1µM, cat no. 25021-233-20, Sagent Pharmaceuticals, IL), irinotecan (0.25µM to 10µM, cat no. 25021-230-05, Sagent Pharmaceuticals), combination of cisplatin (0.1µM to 5µM) and olaparib (0.25µM, cat no. SML3705, Sigma, St. Louis, MO) or combination of cisplatin (0.1µM to 5µM) and IR (2 Gy) and further incubated at 37°C with 5% CO_2_ for 14 days. The colonies were then washed with 1x PBS and stained with 0.2% crystal violet solution for 15 minutes. Following another wash with 1x PBS, the plates were left to dry, and the colonies were manually counted. Percentage survival was calculated, transformed to log_10_, and data is presented as colonies normalized to WT-S464, formed at baseline, from 3 independent experiments.

### Cellular viability assay

EBV-transformed lymphoblastoid cell lines cell line from LARC patients, both WT-S464 (n=6) and SNP-S464T (n=6) were plated in 96-well plates and treated with IR (2 Gy to 50 Gy), 5-fluorouracil (0.5µM to 150µM; cat no. 16729-276-38, Accord Healthcare, Durham, NC), or oxaliplatin (0.25µM to 75µM; cat no. 25021-233-20, Sagent Pharmaceuticals, IL). Similarly, SCC-9 (WT-S464 and SNP-S464T) cells were seeded in 96-well plates at a density of 7000 cells per well. Next day, cells were treated with IR (2 Gy to 30 Gy), or cisplatin (0.5µM to 30µM, cat no. 25021-233-20, Sagent Pharmaceuticals, IL). Cellular viability was measured by short-term CellTiter-Glo® assay (cat no. G7570, Promega Corporation, Madison, WI) 72h after treatment. Data is presented as percent cell killing from 3 independent experiments.

### Immunofluorescence assay

RCM-1, Cal-27 or SCC-9 (WT-S464 and SNP-S464T) isogenic pair cell lines were seeded in chamber slides (cat no. NST-230104, DiagnoCine, NJ, USA) at a density of 10,000 cells per well. Cells were irradiated at 5Gy and fixed with 4% PFA, 1h after IR for γH2AX-MDC1 or MDC1-RAP80 colocalization, 2h after IR for RAP80-BRCA1 colocalization and 24h-96h after IR for cGAS-dsDNA colocalization. Following fixation, cells were permeabilized with 0.2% Triton X-100 and then incubated in MAXblock™ Blocking Medium (cat no. 15252, Active Motif, Carlsbad, CA) for 2hr at room temperature (RT). This was followed by overnight incubation with primary antibodies against MDC1 (1:500, cat no. M2444, Millipore Sigma, Burlington, MA (used for RAP80 colocalization) and cat no. ab11169, Abcam, Cambridge, UK (used for γH2AX colocalization)), BRCA1 (1:500, cat no. sc-6954 (D-9), Santa Cruz Biotechnology Dallas, TX), RAP80 (1:10000, cat no. A300-763A-1, Thermo Fisher Scientific, Waltham, MA), γH2AX (1:500, cat no. 16-193, Millipore Sigma, Burlington, MA), cGAS (1:500, cat no. 79978, Cell Signaling Technology, Danvers, MA), and dsDNA (1:1000, cat no. ab27156, Abcam, Cambridge, UK) at 4°C.

Following overnight incubation, the cells were washed with 0.1% PBS-T and then incubated with either secondary anti-mouse (1:1000, cat no. ab150115, Abcam, Cambridge, UK), anti-mouse (1:500, cat no. A11029, Thermo Fisher Scientific, Waltham, MA), anti-rabbit (1:1000, cat no. ab150079, Abcam, Cambridge, UK) or anti-rabbit (1:500, cat no. A11034, Thermo Fisher Scientific, Waltham, MA) antibodies in the dark for 1h at RT. The cells were further washed with 0.1% PBS-T, then counter-stained with 300nM DAPI (cat no. D1306, Thermo Fisher Scientific, Waltham, MA), and incubated in the dark for 15 minutes at RT. The cells were again washed with 0.1% PBS-T. After a final wash with 1x PBS, coverslips were mounted on the slides with ProLongTM Diamond antifade (cat no. P36965, Thermo Fisher Scientific, Waltham, MA) and kept in dark to cure overnight.

All the images were obtained with Leica TCS Advanced SP8 Confocal microscope (Leica Microsystems, Wetzlar, Germany) using 40x or 63x oil immersion objective. For colocalization analysis between MDC1-γH2AX, MDC1-RAP80, RAP80-BRCA1 or cGAS-dsDNA, data was normalized to the WT-S464 at baseline. The data are presented as mean ±SEM of 3 independent experiments.

### Molecular modeling studies

AlphaFold2 was used to generate the model of KDM3C. Further, evolutionary conservation of the *KDM3C* SNP region was assessed by retrieving KDM3C sequences from 21 different species from UniProt (100) and performing multiple sequence alignment using Clustal Omega (101,102). The multiple sequence alignment was visualized in Jalview (103), which provided annotations for conservation, quality, and consensus scores. The KDM3C peptide consists of a five-amino acid sequence flanking the SNP site at Ser464. The KDM3C peptide model preparation included four variants to study structural effects at the SNP site Ser464: the wild-type peptide with non-phosphorylated Ser464, a phosphorylated Ser464 variant (pSer), a mutant variant with Ser464 replaced by Thr464, and a phosphorylated Thr464 variant (pThr).

#### Structural Modeling of Complexes using AlphaFold3

Each variant of the KDM3C peptide was modeled in complex with the respective protein domains (RNF8-FHA, MDC1-FHA). Specifically, 50 models were generated for each combination of peptide and protein domains. These models were designed to assess the impact of phosphorylation and mutation on peptide structure as well as KDM3C peptide affinity towards the two different protein domains. Structure models were subsequently analyzed for stability, structural rearrangements, and interaction patterns at the peptide-protein domain interface. AlphaFold 3 scoring matrices such as protein-protein interface (iPTM) score and Predicted Aligned Error (PAE) score were used to evaluate the best models. Models having higher iPTM suggest the accuracy of predicted interface in protein-protein/peptide complexes. In contrast, low PAE for residue pairs from different domains indicates that AlphaFold3 predicts their relative positions and orientations with confidence.

#### PISA server Analysis of Complexes

We used PISA server (66) which takes coordinate files as an input and gives structural information such as solvation energy, buried surface area, H-bonds and salt bridges. Average solvation free energy gain (ΔiG) for top 20 models from each variant of KDM3C-peptide and with the RNF8-FHA and, MDC1-FHA domain complexes were obtained using the server. A negative ΔiG indicates favorable binding and strong protein-protein interaction upon interface formation.

#### Interaction Analysis of Complexes

To investigate the interactions between variants of the KDM3C peptide and specific protein do-mains, molecular visualization was performed using PyMOL software. The top-ranking complexes of KDM3C peptide with and without phosphorylation at position 464, with the FHA domains of RNF8 and MDC1 were superimposed to evaluate the presence or absence of interaction in complexes. Superimposition of all 50 models enabled a comparative assessment of the structural impact of phosphorylation on the orientation of the KDM3C peptide when bound to the respective protein domains. Additionally, we also investigated the involvement of SH3 domain of KDM3C in stabilization of complex of RNF8-FHA domain and phosphorylated-SNP variant of KDM3C peptide, as well as examined its potential interactions with the peptide and FHA domain of complex. We generated 50 models of the ternary complex comprising SH3 domains of KDM3C, FHA domain of RNF8 with all four variants of peptide as mentioned above.

### Statistical analysis

All graphs depict mean ± SEM. Tests for differences between two groups were performed using two-tailed unpaired Student *t* test with Welch’s correction or Mann-Whitney test. Values of p < 0.05 (*), p < 0.01 (**), p < 0.001 (***), or p < 0.0001 (****) were considered significant. GraphPad Prism9 was used for statistical analysis of experiments. Statistical analysis for LARC and HNSCC genotyping data was tested for association between response group and genotype using the Fisher’s Exact Test as follows: A/A-PoR vs. CR; A/T-PoR vs. CR and T/T-PoR vs. CR. The statistical analysis for the population level analysis of the cohorts is described in the **supplementary document**.

## Supporting information

All supplementary files

## Data availability statement

The data generated in this study is available on reasonable request from the corresponding author. The patients in this study did not consent to sharing their raw germline sequencing data.

## Conflict of interest

The authors declare there is no conflict associated with this study.

## Ethics Statement

The FCCC Biosample Repository Facility obtains informed consent and HIPAA authorization from all participants for the use of de-identified specimens and associated clinical data prior to specimen collection (IRB #11-866). The work in this study was performed under FCCC (Fox Chase Cancer Center) Institutional Review Board approved protocols, #18-4005, #19-4001 and #21-9921.

## Acknowledgments

The work in this grant was supported by the resources and expertise provided by several FCCC core facilities: FCCC Cell Culture Facility, Biosample Repository Facility, Biostatistics and Bioinformatics Facility, Biological Imaging Facility, Radiation Safety Facility, and Molecular Modeling and Screening Facility (MMSF). All work with patient biospecimens and corresponding clinical and demographic data were approved by the FCCC IRB (IRB# 18-4005 and #19-4001). We acknowledge Shreya Shah (summer research assistant at FCCC), for assistance with reviewing TCGA files, and Margret B. Einarson, PhD, (Manager, MMSF), for guidance on these studies. We acknowledge George Shenoda and Samhita Ganti (High school volunteers, Arora Lab, FCCC) for assistance with sample preparation for integrated γH2AX and cell cycle studies. We also acknowledge Paulina Bleu (MSKCC), Johanna D. James (FCCC), and Dr. Thomas J. Galloway (FCCC) for their assistance during this study. During this work, A.H. was awarded the C. David Allis Travel Award at the 2025 CEI Annual Symposium and received two AACR Scholar-in-Training Awards in 2025 related to this study. Part of this research has been conducted using the UK Biobank Resource under application number 533532. The datasets used for the analyses described in this manuscript were obtained from dbGaP under approval number (Project-18630; Title-Mutagenic impact of DNA replication-repair variants).

## Author contributions

**A.H.,** performed most studies, all data analysis, writing, editing; **P.P., M.A., R.L.D.,** structural modeling studies; **E.V.D., P.C.**, performed clinical data studies and analysis; **L.G.,** performed integrated cell cycle and γH2AX experiments; **T.M., I.S.**, performed cGAMP, mRNA expression experiments and data analysis; **C.W., J.J.S.,** rectal tumoroids study; **Z.A.C.K., R.K.R.S.,** UK Biobank and All of Us cohort analysis; **G.H., G.L.R.,** mutational signature analysis; **Y.Z.,** UK Biobank and All of Us cohort analysis review and TCGA studies; **K.D.,** statistical analysis; **A.S.,** Bulk RNA-seq gene expression data analysis; **B.M.S., D.C.C.,** assistance with biospecimen selection and provision of specimens and clinical data; **J.L.,** intellectual contribution; **B.B.,** intellectual contribution; **E.A.G.,** editing manuscript and intellectual contribution; **J.W.,** editing manuscript and intellectual contribution**; J.E.M.,** editing manuscript and intellectual contribution; and **S.A.,** designing and planning all studies, writing, editing and funding acquisition.

## Funding Statement

All Fox Chase Cancer Center (FCCC) Core Facilities and authors are in part supported by the NCI Comprehensive Cancer Center Support Grant, P30CA006927, to FCCC. This work was supported by the Career Enhancement Proposal (CEP) Award from the Yale Head and Neck Cancer NIH SPORE (P50 DE030707) to S.A., DOD Career Development Award to S.A. (W81XWH-18-1-0148), DOD Idea Award to S.A. (HT9425-23-1-0840), and NIH UH2/UH3 (1UH2CA271230-01/5UH3CA271230-04) to S.A. and J.E.M. A.H. was supported by the DOD Idea Award to S.A. (HT9425-23-1-0840), NIH UH2/UH3 (1UH2CA271230-01/5UH3CA271230-04) to S.A. and by FCCC Board of Directors Fellowship (to A.H.). R.D. was supported by NIH grant R35 GM122517. E.A.G., was supported by NIH grant 1R03CA292552 and the William Wikoff Smith Charitable Trust. E.A.G., along with B.B., was also supported by the DOD grant CA201045/W81XWH2110487. B.B. and D.C.C. were additionally supported by NIH grant P50 DE030707. I.C., was supported by NIH grant R37 Award CA283552-01A1. G.L.R., was supported by the National Science Foundation under grant numbers #1936791, #1919691, and #2107108. J.R.W. was supported by NIH grant R35GM144131. J.J.S. is supported by NIH/NCI R37 Award CA248289. Z.K. was awarded the *In Vino Vita* Young Investigator Award for the UK Biobank cohort studies performed here. T.M. was additionally supported by the Grant-in-Aid, Japan Society for Promotion of Science fellowship (202160224).

## Notes

### Competing Interest Statement

Author Disclosure Information: J.J.S received travel support for fellow education and personal education from Intuitive Surgical (August 2015 and April 2026); served as a clinical advisor for Guardant Health (March 2019); served as a clinical advisor for Foundation Medicine (April 2022); served as a consultant and speaker for Johnson and Johnson (May 2022); served as a clinical advisor for Urogen (January 2025); served as a clinical advisor for Regeneron (January 2025); serves as a clinical advisor and consultant for GSK (2023-26); served as a clinical consultant for Gerson Lehrman Group (March 2025); served as a clinical consultant for Agenus (May 2025, May 2026); served as a clinical consultant for BioNTech (May 2025); served as a clinical consultant for Clario (2025-26). J.R.W. at Fox Chase Cancer Center is an inventor on U.S. Provisional Patent Application No.: 63/938,301. J.R.W. has served or is serving as a consultant or advisor for Qsonica, Daiichi Sankyo, Inc., and Vyne Therapeutics in past year. J.R.W. received an honorarium from Arima Genomics and is a co-investigator on a sponsored project. J.R.W also received funding from Oryzon Genomics. B.B. has served as a consultant for Astra Zeneca, Bicara, Genmab, Glaxo Smith Kline, Johnson and Johnson, Merck, Merus, One Carbon, and Takeda. J.E.M. has patents issued related to colorectal cancer diagnostics, receives teaching honoraria from Varian Medical Systems, has a strategic advisor relationship with Quantigic Genomics. I.C. has received non-financial support from Accent Therapeutics and consulting/advisory honoraria from Storm Therapeutics. S.A. patent(s) issued related to colorectal cancer diagnostics and has received funding from Merck & Co. during the conduct of the study. M.A. has a patent issued related to methods for diagnosing a predisposition to develop colon cancer. No disclosures were reported by the other authors.

### Author Declarations

The FCCC Biosample Repository Facility (BRF) obtains informed consent and HIPAA authorization from all BRF participants for the use of de-identified specimens and associated clinical data prior to specimen collection (IRB# 11-866). Deidentified biospecimens and associated clinical data from LARC and HNSCC patients were obtained from the FCCC BRF under FCCC Institutional Review Board (IRB)-approved protocols, #18-4005, #19-4001 and #21-9921.

