## Supplementary material for "A germline KDM3C polymorphism impairs DNA repair and sensitizes to chemoradiotherapy": All supplementary files

### Supplementary Results.

#### Exploratory analysis of somatic driver mutations in rectal and head and neck cancer by *KDM3C* genotype.

To characterize the genomic landscape of tumors by the specific *KDM3C* genotypes (A/A, A/T, and T/T), we stratified rectal and HNSCC tumors by the germline status of *KDM3C* in TCGA. Here, the genotype groups were not evenly represented, with enrichment of T-allele carriers, reflecting cohort composition, thereby limiting formal statistical analyses and interpretation to descriptive patterns.

A total of 145 rectal tumors were examined, comprising A/A (WT, n= 11), heterozygous A/T (n= 49), and homozygous T/T (n= 85) genotypes. For downstream analyses, samples were grouped into WT A/A (n= 11) and combining the heterozygous (het) and homozygous (homo) T-allele carriers, A/T + T/T, (n= 134). We first assessed whether the *KDM3C* genotype distribution was associated with demographic variables in this rectal cancer cohort using Fisher's exact test. Genotype distribution showed a borderline association with race (p= 0.056), with A/A WT and SNP T-allele counts of 2 and 3, respectively, among self-reported African American patients, and 5 and 72, respectively, among European American patients. No significant association was observed between genotype and sex (p= 0.548), with A/A WT and SNP T-allele counts of 6 and 63 among female patients, and 5 and 78 among male patients. Similarly, genotype distribution was not significantly associated with age group (p= 0.535), with A/A WT and SNP T-allele counts of 4 and 70 among older patients (>61 years), and 7 and 71 among younger patients (≤61 years).

Next, we assessed whether KDM3C genotype groups were associated with mutational profiles of recurrently altered driver genes in rectal cancer, using Fisher's exact test. Among the genes tested, *BRAF* showed a significant association with genotype group (p= 0.0318). *EGFR* demonstrated a non-significant trend toward association (p = 0.0964). No significant associations were observed for *TP53*, *APC*, *KRAS*, *NRAS*, *PIK3CA*, *SMAD4*, *PTEN*, *FBXW7*, *POLE*, or *POLD1*. Finally, we examined the type of mutations present in a subset of recurrent rectal driver genes, including *APC*, *KRAS*, *BRAF*, *SMAD4*, *TP53* (**Supplementary Table 13**).

*APC* mutations were observed across *KDM3C* genotypes and were predominantly truncating, consisting mainly of nonsense and frameshift mutations. In raw counts, nonsense mutations were identified in 7 WT, 29 HET SNP, and 49 Homo SNP tumors, while frameshift mutations were observed in 6 WT, 12 HET SNP, and 30 Homo SNP tumors. In contrast, *APC* splice-site and missense mutations were infrequent across all groups, consistent with the canonical tumor-suppressor inactivation pattern of *APC* in colorectal tumorigenesis (1,2)

*KRAS* mutations, primarily at codon G12, were also detected across all genotypes. G12V represented the most frequent alteration (1 WT, 6 HET SNP, and 9 Homo SNP tumors), followed by G12D, which was restricted to HET SNP and Homo SNP carriers, and G12C, which was present across all groups. Additional less common G12 substitutions were observed were also observed in the Homo SNP group.

*BRAF* mutations were extremely rare, with the canonical V600E alteration observed only in the WT, while non-V600 missense mutations were detected in SNP carrier groups. *SMAD4* alterations were distributed across the *KDM3C* genotypes and were predominantly missense (2 WT, 4 HET SNP, and 11 Homo SNP tumors). However, truncating events, including frameshift and in-frame deletions, were observed exclusively in the Homo SNP group.

*TP53* mutations were highly prevalent across the *KDM3C* genotypes and followed the expected spectrum of missense mutations (7 WT, 24 HET SNP, and 38 Homo SNP tumors). These were accompanied by lower frequencies of nonsense, splice-site, frameshift, and in-frame indel alterations. Recurrent hotspot mutations-including R175H, R248Q/W, R273H/C, R282W, and G245S.

Next, a total of 493 HNSCCs were examined, including A/A (n=48), A/T (n=178), and T/T (n=267) status. For downstream analyses, samples were grouped into WT A/A (n= 48) and combining the het and homo T-allele carriers (A/T + T/T; n= 445). We first assessed whether *KDM3C* genotype distribution was associated with demographic variables in this HNSCC cancer cohort using Fisher's exact test. Genotype distribution showed significant association by race (p= 0.0002405) with WT and SNP counts of 12 and 33, among self-reported African American patients, respectively, and 32 and 104 among European American patients. Further, genotype distribution showed significant association with age group (p= 0.04878) with WT A/A and SNP T-allele counts of 17 and 233 among older patients (>61 years), and 31 and 226 among younger patients (≤61 years). However, no significant association was observed between genotype and sex (p= 0.7336) with WT A/A and SNP T-allele counts of 14 and 123 among female patients and 34 and 337 among male patients.

We next assessed whether *KDM3C* genotype groups were associated with mutational profiles of recurrently altered driver genes in HNSCC genes, using Fisher's exact test. No significant genotype-associated enrichment was observed for any of the genes tested, including *TP53*, *PTEN*, *MDM2*, *EGFR*, *CDKN2A*, *FAT1*, *NOTCH1*, *PIK3CA*, *KMT2D*, *FGF3*, *CCND1*, *HRAS*, *BRAF*, or *NSD1*. Finally, we examined the type of mutations present in a subset of recurrent HNSCC genes, including *NOTCH1*, *NSD1*, *PIK3CA*, *TP53*, *CDKN2A*, *FAT1* and *KMT2D* (**Supplementary Table 13**).

*NOTCH1* mutations were observed across the *KDM3C* genotypes and were predominantly missense, representing the most frequent type of alteration (4 WT, 20 HET SNP, and 31 Homo SNP tumors). Nonsense and frameshift mutations were also detected across all groups, while splice-site and in-frame deletion events were comparatively infrequent.

*NSD1* mutations exhibited a broad spectrum of alteration types, including missense, nonsense, frameshift, splice-site, and in-frame deletion events. While nonsense and frameshift mutations were more prominent in the *KDM3C* Homo SNP group, the HET SNP group showed relatively higher raw counts of missense alterations. Overall, *NSD1* variants were distributed across all genotype groups, with truncating or disruptive alterations observed in each group.

*PIK3CA* mutations were predominantly missense and included recurrent hotspot mutations at E542, E545, Q546, and H1047. E542K was observed in 3 WT, 12 HET SNP, and 4 Homo SNP tumors, whereas E545K was observed in 1 WT, 9 HET SNP, and 14 Homo SNP tumors. H1047R was observed in 4 HET SNP and 8 Homo SNP tumors, while Q546R, H1047L, and H1047Q were observed only in the Homo SNP group.

*TP53* mutations were highly prevalent across all *KDM3C* genotypes and were dominated by missense alterations (26 WT, 77 HET SNP, and 134 Homo SNP tumors). Additional mutation classes, including nonsense, frameshift, splice/splice-region, and in-frame indels were observed at lower frequencies. Recurrent hotspot mutations, including Y220C, R175H, R248Q/W, R273H/C, R282W, and G245 variants, were consistently present across genotype groups, with no hotspot uniquely enriched in any specific *KDM3C* genotype.

*CDKN2A* mutations were observed across all *KDM3C* genotypes and were primarily truncating (5 WT, 19 HET SNP, and 33 Homo SNP tumors). Frameshift, splice-site, missense, and in-frame deletion events occurred less frequently. Recurrent alterations included R80*, R58*, W110*, E88*/E88fs, and X153_splice, with R80* and R58* representing the most frequently observed events. W110* was detected only in HET SNP and Homo SNP groups in this dataset.

*FAT1* mutations were observed across all *KDM3C* genotypes and were dominated by truncating alterations. Here, nonsense mutations were observed in 7 WT, 26 HET SNP, and 31 Homo SNP tumors, while frameshift mutations were observed in 3 WT, 12 HET SNP, and 24 Homo SNP tumors. Missense mutations were less frequent, and splice-site and in-frame deletion events were rare. Recurrent *FAT1* alterations were limited, with R937* observed in the HET SNP group and S3373*, R885*, R1627*, and S2826Kfs*2 recurrent in the Homo SNP group.

*KMT2D* mutations were observed across all *KDM3C* genotypes and included missense, nonsense, frameshift, splice/splice-region, and in-frame deletion events. Missense mutations were observed in 4 WT, 5 HET SNP, and 18 Homo SNP tumors, while nonsense mutations were observed in 3 WT, 10 HET SNP, and 15 Homo SNP tumors. Frameshift alterations were also frequent, occurring in 2 WT, 7 HET SNP, and 15 Homo SNP tumors.

Together, these analyses provide an exploratory overview of somatic mutation patterns across *KDM3C* genotypes. Future work would be to study a dataset specifically designed with balanced genotype representation will be required to rigorously assess genotype–somatic mutation relationships.

**KDM3C contains three folded modules with subdomain organization.**

While the catalytic domain of KDM3C is the only part with an experimentally determined structure (PDB: 5FZO), we have used AlphaFold2/AlphaFold3 to model the full-length structure of KDM3C. These programs are capable of high-resolution predictions of protein domain folds. To further analyze the potential relationships between KDM3C and other proteins, we employed the Foldseek (3) server, which searches for the AlphaFold Database (AFDB) containing models for over 200 million proteins from UniProt (4).

The results indicate that KDM3C contains three folded modules, each with distinct sub-domains: **Module 1** (residues 1-254) forms a complex of three intertwined SH3-like small beta barrels, a structure shared with KDM3A and KDM3B across different species. **Module 2** (residues 1694-1938) appears to be a regulatory domain. The region spanning residues 1694-1775 forms a novel zinc-binding domain, unique to KDM3 proteins. This module also contains a ZZ (zinc-zinc) domain (residues 1830-1906) that binds two zinc ions, shared with KDM6 proteins and EP300. **Module 3** (residues 2150–2450) includes the catalytic JmjC domain, which forms a beta helix and two sub-domains—one unique to KDM3s and another shared with KDM families 2, 3, and 7. Modules 2 and 3 appear to form a single complex, while Module 1 is typically detached in most models.

**KDM3C SNP-S464T is associated with cell-line-specific gene-expression changes.**

Using bulk mRNA sequencing, we assessed *KDM3C* genotype-dependent gene-expression differences across three isogenic cell-line models. RNA-seq did not identify a uniform transcriptional signature shared across RCM-1, CAL-27, and SCC-9 cells (**Supplementary Table 18**). Instead, genotype-associated gene-expression changes were largely context dependent, likely reflecting the distinct parental backgrounds and tissue origins of the rectal and HNSCC models.

In RCM-1, SNP-S464T cells showed downregulation of pathways related to DNA methylation, inflammatory response, nucleosome organization, and HDAC-associated histone regulation, with upregulation of oxidative phosphorylation, mitochondrial respiratory chain, ROS metabolic processes, interferon α/β signaling, and cytokine activity (**Supplementary Figure 5C**). In CAL-27 SNP-S464T cells, gene-expression changes were characterized by downregulation of genome-maintenance pathways, including DNA replication, cell-cycle checkpoint, chromosome segregation, double-strand break repair, nucleotide-excision repair, and mismatch repair, while upregulated pathways included apoptotic signaling, chemokine production, adaptive immune response, and protein kinase B signaling (**Supplementary Figure 5D**). In SCC-9 SNP-S464T cells, downregulated pathways included extracellular matrix and extracellular structure organization, whereas upregulated pathways included cytokine–cytokine receptor interaction, receptor ligand activity, growth factor activity, mesenchymal-to-epithelial transition, and extrinsic apoptotic pathway (**Supplementary Figure 5E**).

**Supplementary Methods.**

**Exome sequencing, variant annotation, filtering, and prioritization.** We performed WES on lymphocyte DNA from 30 LARC cases in the study. Whole-exome sequencing of DNA was performed by BGI Americas Corporation (Cambridge, MA) at 100X average coverage. Agilent SureSelect XT All Exon V6 kit was used for exon capture (Agilent Technologies, Wilmington, DE). Library preparation was performed using standard Illumina protocols. Each captured library was indexed and loaded onto Hiseq2000 platform (Illumina, Hayward, CA) for 100 bp paired-end high-throughput sequencing. Sequence reads were mapped to human reference genome (hg19) using the Burrows-Wheeler Aligner (BWA) (5). Single Nucleotide Polymorphisms (SNPs) and small Insertion/Deletions (InDels) were detected using Genome Analysis Toolkit (GATK) (6). Variant annotations were computed with ANNOVAR (7) version 2019-10-24 and included: population frequency from (a) ANNOVAR-provided versions of the gnomAD genome and exome call sets (8), version 2019-11-04 release; (b) predicted protein impact, using ANNOVAR-provided versions of RefSeq (9), and (c) predicted deleteriousness, using the ANNOVAR file dbnsfp30a (10). Annotations from InterVar (11), a bioinformatics software tool for clinical interpretation of genetic variants by the ACMG/AMP 2015 guidelines (12) were also included in the report.

After variant calling and annotation, selected filters were applied to prioritize variants for downstream analysis and manual review. Although not directly associated to the study hypothesis, the sequencing data were reviewed for any pathogenic or likely pathogenic variants (PV/LPV) in cancer risk genes that are frequently tested on multi-gene cancer panels (13-15). Based on the study hypothesis, we focused our attention on genes involved in core DNA repair, as well as pathways supporting genome maintenance, including chromatin regulation, transcription, DNA replication, cell-cycle control, protein ubiquitination, chromatin assembly, and immune-associated processes. A 161-gene analysis set was compiled from candidate gene lists previously published by our group and others, together with additional chromatin-associated genes relevant to genome stability (**Supplementary Table 1** (16,17)).

Variants identified by WES were prioritized using a multi-step filtering strategy. Only missense variants were prioritized. Rare variants were required to meet quality-control thresholds of genotype quality ≥10, read depth ≥10, and maximum population allele frequency <0.01. Common variants were evaluated separately, with prioritization based on differences in allele-frequency across gnomAD subpopulations, including ancestry- and sex-stratified groups, and zygosity was also considered. Rare variants were further prioritized if they were predicted to alter the protein sequence in any RefSeq transcript or were annotated as pathogenic or likely pathogenic by at least two variant effect prediction tools, including SIFT, PolyPhen-2, MutationAssessor, and CADD. Candidate variants taken forward for analysis were confirmed by Sanger sequencing (GENEWIZ, South Plainfield, NJ). Additional DNA specimens from an independent LARC set were also assessed by Sanger sequencing for selected candidate SNPs of interest.

To functionally organize the 161-gene background set, genes were assigned to pathway categories using WebGestalt 2019 (WEB-based GEne SeT AnaLysis Toolkit), enabling evaluation of how the screened genes segregated across biologically relevant pathways (**Supplementary Table 2**). Following application of the prespecified filtering strategy, we identified 77 genes harboring germline variants (**Supplementary Table 3, 6**). Enrichment was assessed by comparing the number of genes observed in the candidate germline variants list (n=77) with full gene set (n=161). Pathway enrichment was assessed using Fisher's exact test on 2 × 2 contingency tables comparing genes in the candidate list versus genes not in the candidate list, stratified by pathways identified via WebGestalt 2019 (**Supplementary Table 4**). To account for multiple testing across pathways, p values were adjusted using the Benjamini–Hochberg false discovery rate (FDR) method (18). Pathways with q<0.05 were considered enriched (**Supplementary Table 5)**.

Finally, we calculated the expected genotype frequencies for *KDM3C* rs10761725 from the gnomAD (v4.1.1) T-allele frequency under Hardy–Weinberg equilibrium (19,20), where p represents the A-allele frequency and q represents the T-allele frequency. Expected genotype frequencies were calculated as $p^2$ for A/A, 2pq for A/T, and $q^2$ for T/T.

**SNP genotyping.** SNP genotyping was performed by Azenta Life Sciences (South Plainfield, NJ). Genomic DNA was isolated from PBMCs used for SNP genotyping of the KDM3C SNP-S464T variant from the LARC (n=84) and HNSCC (N=90) cohort. Details of the primers used are mentioned in **Supplementary Table 19**.

**Drug treatment in rectal organoids**. Drug sensitivity assays in patient-derived rectal cancer organoids were performed by the Smith group as previously described (21).

**Bulk RNA sequencing.** Total RNA from RCM-1, CAL-27 or SCC-9 (WT-S464 and SNP-S464T) cells was isolated at baseline, and 24h after 2Gy IR using the Qiagen RNA isolation kit (74104, Qiagen, Hilden, Germany). RNA sequencing was performed by Novogene Inc. (Sacramento, CA).

Messenger RNA was purified from total RNA using poly-T oligo-attached magnetic beads. After fragmentation, the first strand cDNA was synthesized using random hexamer primers, followed by the second strand cDNA synthesis using either dUTP for directional library or dTTP for non-directional library. For the non-directional library, it was ready after end repair, A-tailing, adapter ligation, size selection, amplification, and purification. For the directional library, it was ready after end repair, A tailing, adapter ligation, size selection, USER enzyme digestion, amplification. The library was checked with Qubit and real time PCR for quantification and bioanalyzer for size distribution detection. Quantified libraries were pooled and sequenced on Illumina platforms, according to effective library concentration and data amount. Further, clustering of the index coded samples was performed according to the manufacturer's instructions. After cluster generation, the library preparations were sequenced on an Illumina platform, and paired end reads were generated.

Raw data (raw reads) of fastq format were first processed through fastp software. In this step, clean data (clean reads) were obtained by removing reads containing adapter, reads containing ploy-N and low-quality reads from raw data. At the same time, Q20, Q30 and GC content the clean data were calculated. All the downstream analyses were based on clean data with high quality. Reference genome and gene model annotation files were downloaded from genome website directly. Index of the reference genome was built using Hisat2 v2.0.5 and paired-end clean reads were aligned to the reference genome using Hisat2 v2.0.5. Hisat2 was selected as the mapping tool for that Hisat2 can generate a database of splice junctions based on the gene model annotation file and thus a better mapping result than other non-splice mapping tools.

Differential expression analysis of two conditions/groups (two biological replicates per condition) was performed using the DESeq2 R package (1.20.0). DESeq2 provides statistical routines for determining differential expression in digital gene expression data using a model based on the negative binomial distribution. The resulting P-values were adjusted using Benjamini and Hochberg's approach for controlling the false discovery rate. Genes with an adjusted Q-value<=0.05 found by DESeq2 were assigned as differentially expressed.

Further the clusterProfiler R package (22) software was used for enrichment analysis. GO terms with Q-value less than 0.05 were considered significantly enriched by differential expressed genes. KEGG is a database resource for understanding high-level functions and utilities of the biological system, such as the cell, the organism, and the ecosystem, from molecular-level information, especially large-scale molecular datasets generated by genome sequencing and other high-through put experimental technologies (http://www.genome.jp/kegg/). The cluster Profiler R package was used to test the statistical enrichment of differential expression genes in KEGG pathways. The Reactome database brings together the various reactions and biological pathways of human model species. Reactome pathway pathways with corrected P value less than 0.05 were considered significantly enriched by differential expressed genes.

For Gene Set Enrichment Analysis (GSEA), genes were ranked according to the degree of differential expression in the two samples, and then the predefined Gene Set were tested to see if they were enriched at the top or bottom of the list. Gene set enrichment analysis can include subtle expression changes. The local version of the GSEA analysis tool (http://www.broadinstitute.org/gsea/index.jsp; (23)), GO, KEGG, Reactome, and WebGestalt (WEB-based GEne SeT AnaLysis Toolkit) (24-26) were used for GSEA independently.

**Integrated $\gamma$H2AX$^{S139}$ quantification and cell-cycle analysis.** RCM-1, CAL-27 or SCC-9 (WT-S464 and SNP-S464T) cells were at a density of 2 x $10^6$ cells/plate. RCM-1 and CAL-27 cells were synchronized at the G1/S boundary using a double thymidine block, whereas SCC-9 cells were synchronized in G0/G1 by serum starvation. For double thymidine synchronization, CAL-27 and RCM-1 cells at ~60% confluence were treated with 2mM thymidine for ~17h (Block 1), washed twice with PBS, and released into thymidine-free medium for ~8h. Cells were then treated with 2mM thymidine for an additional ~17 h (Block 2) and released into complete medium, after which samples were collected at the different time points with or without treatment. SCC-9 cells were synchronized by culturing in medium containing 1% FBS for 48h to induce G0/G1 arrest, followed by release into complete medium containing 10% FBS; samples were collected at defined time points after serum re-addition.

For all conditions, cells were harvested by trypsinization, washed twice with 1x PBS, and ~1.5 x $10^6$ cells were collected per sample. Cell pellets were fixed for 24h at −20°C in ice-cold 70% ethanol, which was added dropwise with gentle vortexing to prevent clumping. For $\gamma$H2AX staining, fixed samples were centrifuged to remove ethanol, permeabilized with 0.2% Triton X-100, and incubated in MAXblock™ Blocking Medium (cat no. 15252, Active Motif, Carlsbad, CA) for 2h at RT. Cells were then incubated overnight at 4°C with $\gamma$H2AX$^{S139}$ primary antibody (1:500, 16-193, Millipore Sigma, Burlington, MA). Following primary incubation, cells were washed with 1x PBS and incubated with Alexa Fluor-conjugated anti-mouse secondary antibody (1:1000, cat no. A11029, Thermo Fisher Scientific, Waltham, MA) for 1h at RT in the dark. After washing, cells were resuspended in PI/RNase Staining Buffer (cat no. 550825, BD Biosciences, Franklin Lakes, NJ) for 30 min at RT and analyzed by flow cytometry to obtain integrated $\gamma$H2AX levels within defined cell-cycle phases

**Protein extraction, co-immunoprecipitation, and immunoblotting.** CAL-27 or SCC-9 (WT-S464 and SNP-S464T) were seeded at a density of 1 x $10^6$ cells per 10-cm$^2$ culture plate. CAL-27 cells were harvested 2h after 10Gy IR, washed with cold PBS, and lysed in RIPA lysis buffer (cat no. 89900, Thermo Fisher Scientific, Waltham, MA) supplemented with protease inhibitor cocktail Set III (cat no. 535140, EMD Millipore Sigma, Burlington, MA). For SCC-9 cells (WT-S464 and SNP-S464T), whole cell lysates were prepared 24-96 h after 5 Gy IR. Lysates were incubated at 4°C for 20 min under constant agitation, and the supernatants were collected by centrifugation at 15,000 rpm for 15 min. Protein content was quantified using Qubit™ Protein BR assay kit (cat no. A50668, Thermo Fisher Scientific, Waltham, MA).

For Co-IP using CAL-27 lysates, 250µg total cellular protein was incubated with KDM3C primary antibody (1:100, cat no. sc-101073 (BA-09), Santa Cruz Biotechnology, Dallas, TX) for 5h at 4°C. After binding, 25µl of the resuspended Protein A/G PLUS-agarose beads (cat no. sc-2003, Santa Cruz Biotechnology, Dallas, TX) was added and incubated overnight at 4°C on a rotating device. Immune complexes were pelleted by centrifugation at 2,500 rpm for 5 min at 4°C, washed four times with cold 1× PBS, and centrifuged after each wash. Pellets were resuspended in 50µL of 1× electrophoresis sample buffer, boiled for 5 min, and centrifuged to remove the beads. Immunoblotting was performed using 25µL of the immunoprecipitated proteins.

For immunoblotting, either 25µL of CAL-27 immunoprecipitated protein, 50µg of CAL-27 whole cell lysate or 20µg of SCC-9 whole-cell lysate was resolved on Novex™ 8% Tris–Glycine gels (cat no. XP00082BOX, Thermo Fisher Scientific, Waltham, MA) gel and transferred to 0.2µm nitrocellulose membrane (cat no. 1620112, Bio-Rad, Hercules, CA). Membranes were blocked using 2% BSA and incubated overnight at 4°C with the following primary antibodies: KDM3C (1:1000, cat no. sc-101073 (BA-09), Santa Cruz Biotechnology, Dallas, TX), MDC1 (1:1000, cat no. M2444, Millipore Sigma, Burlington, MA), TBK1 (1:1000, cat no. 3504, Cell Signaling Technology, Danvers, MA), pTBK1 (Ser172) (1:1000, cat no. 5483, Cell Signaling Technology, Danvers, MA), and β-actin (1:1000, cat no. sc-47778, Santa Cruz Biotechnology Dallas, TX). After incubation, membranes were washed with 0.05% PBS-T and incubated with HRP-conjugated anti-mouse (1:2000, cat no. 7076, Cell Signaling Technology, Danvers, MA) or anti-rabbit secondary antibody (1:2000, cat no. 7074, Cell Signaling Technology, Danvers, MA) for 1h at RT. Blots were developed using SuperSignal™ West Pico PLUS chemiluminescent substrate (cat no. 34580, Thermo Fisher Scientific) and imaged using the ChemiDoc™ MP system (Bio-Rad). β-actin served as a loading control for normalization. A protein ladder (10–250 kDa, cat no. 161-0375, Bio-Rad, Hercules, CA) was used to verify molecular weights.

**Quantitative PCR with reverse transcription.** SCC-9 (WT-S464 and SNP-S464T) cells were seeded in a T-75cm$^2$ flask at a density of 1 x 10$^6$ cells/flask and irradiated at 5Gy. Total RNAs were extracted at baseline, 72h and 96h post 5G IR using the RNeasy Mini Kit (cat no. 74104, Qiagen, Hilden, Germany) according to the manufacturer's instructions. A total of 800ng extracted RNA was used to generate cDNA with the ProtoScript® II First Strand cDNA Synthesis Kit (cat no. E6560L, New England Biolabs, Ipswich, MA), for RT-qPCR kit (cat no. 18080-044, Thermo Fisher Scientific, Waltham, MA). qRT-PCR of the indicated genes (**Supplementary Table 20**) was performed using Power SYBR Green PCR Master Mix (cat no. 4367659, Applied Biosystems, Waltham, MA) using Applied Biosystems QuantStudio 6 Pro Real-Time PCR System and software. The relative expression was normalized with the expression of the housekeeping genes *36B4* and analyzed with the –ΔΔCt relative quantification method.

**ELISA for 2′,3′-Cyclic GAMP (cGAMP).** CAL-27 or SCC-9 (WT-S464 and SNP-S464T) cells were seeded in a T-75cm$^2$ flask at a density of 1 x 10$^6$ cells/flask. Cell lysates were collected at baseline, 72h and 96h post 5Gy

IR. The lysates were analyzed for 2′,3′-Cyclic GAMP (cGAMP) using ELISA (cat no. K067-H1, Arbor Assays, Washtenaw, MI) kit according to the manufacturer's instructions.

**Apoptosis assessment.** CAL-27 or RCM-1(WT-S464 and SNP-S464T) cells were seeded in $10cm^2$ plates at a density of 1 x $10^6$ cells/plate. Cell suspension was collected at baseline and at 24-96h after treatment with different DNA damaging agents. Suspended cells were diluted in Annexin V-binding buffer and stained using the FITC Annexin V/Dead Cell (cat no. 556547, BD Biosciences, Franklin Lakes, NJ) according to manufacturer's instructions.

**Mass spectrometry analysis.** Phospho-proteomic nanoLC-MS/MS mass spectrometry on immunoprecipitated KDM3C from isogenic CAL-27 WT-S464 and SNP-S464T cell lines under basal conditions (no IR) and following ionizing radiation (post IR) was performed by Creative Proteomics (Shirley, NY) using an Ultimate 3000 nano-UHPLC system (ThermoFisher Scientific, Waltham, MA). KDM3C was immunoprecipitated from each cell lysate, separated by gel electrophoresis, excised using 3 enzymes (Glu-C, chymotrypsin, and trypsin), and subjected to LC-MS/MS to assess post-translational modifications. The full scan was performed between 300-1,650 m/z at the resolution 60,000 at 200 m/z, the automatic gain control target for the full scan was set to 3e6. The MS/MS scan was operated in Top 20 mode using the following settings: resolution 15,000 at 200 m/z; automatic gain control target 1e5; maximum injection time 19 milliseconds; normalized collision energy at 28%; isolation window of 1.4 Th; charge sate exclusion: unassigned, 1, > 6; dynamic exclusion 30 s. 18 raw MS files were analyzed and searched against MDC1 reference sequence (UniProt: Q14676), using PEAKS STUDIO 8.5, with phosphorylation (STY) as variable modification. The enzyme specificity was set to trypsin, Glu-C, or chymotrypsin; the maximum missed cleavages were set to 2; the precursor ion mass tolerance was set to 10 ppm, and MS/MS tolerance was 0.02 Da.

**Phosphorylation prediction analysis.** Potential kinase recognition at KDM3C S464 site were evaluated using NetPhos-3.1 (27,28). Short peptide sequences surrounding the WT S464 residue and the p.S464T variant residue were analyzed separately. The WT sequence was entered as MIIHSSEQS, and the variant sequence was entered as MIIHTSEQS. NetPhos generates kinase-specific phosphorylation prediction scores ranging from 0 to 1, with scores above the standard 0.5 threshold considered positive predictions. Predictions were compared between the WT-S464 and SNP-S464T sequences to determine whether the S464T substitution altered local phosphorylation potential or predicted kinase specificity.

**UK Biobank and All of Us analysis.** The population level analysis of the cohorts as well as statistical tests are described below.

UK Biobank (discovery/validation). UK Biobank is a prospective, population-based cohort of 501,946 participants aged 40–69 years recruited across the United Kingdom between 2006–2010. The resource includes genomic and deep phenotypic data with longitudinal follow-up through linkage to national cancer registries, hospital episode statistics, and death registries. UK Biobank received ethical approval from the North West Multi-Centre Research Ethics Committee, and all participants provided written informed consent.

All of Us Research Program (cross-validation). To evaluate reproducibility in a cohort with greater ancestral and sociodemographic diversity, analyses were replicated in the All of Us Research Program, a nationwide longitudinal cohort designed to advance precision medicine through inclusion of historically underrepresented populations (approximately 50% self-identify as racial/ethnic minorities). The program integrates multiple data modalities, including electronic health records, genomic data (genotyping and whole-genome sequencing), participant-provided survey data, and physical measurements, with ongoing longitudinal follow-up. All of Us has institutional review board approval and has obtained informed consent from all participants. Analyses were performed within the All of Us Researcher Workbench, a secure cloud-based platform that provides controlled access to de-identified data and supports analysis using integrated tools (e.g., R, Python, and SQL-based workflows) without direct data download.

Genotyping and genotype groups (UK Biobank). Genotype data were obtained from UK Biobank genotyping array and/or whole-genome sequencing resources. Participants were categorized as homozygous T/T variant (SNP/SNP), heterozygous A/T (SNP/Ref), or homozygous A/A reference (Ref/Ref). Primary analyses focused on homozygous T/T variant carriers versus homozygous A/A reference individuals (recessive model). **All of Us.** Genotypes were obtained from the whole-genome sequencing dataset within the Researcher Workbench using the variant identifier from the wgs_variant_calls_ld_pruned table. Genomic analyses were conducted within the Controlled Tier of the All of Us Research Program Researcher Workbench, which provides access to individual-level whole-genome sequencing data under controlled data use policies. Allele calls were coded as 1 (variant) or 0 (reference) for allele1 and allele2, and participants were classified as homozygous T/T variant (1/1), heterozygous A/T (1/0), or homozygous A/A reference (0/0). Primary analyses compared homozygous T/T variant carriers to homozygous A/A reference individuals.

Cancer outcome ascertainment (UK Biobank). Cancer outcomes were ascertained through linkage to national cancer registries, hospital episode statistics, and death registries, and mapped using ICD-10 codes. Age at diagnosis was computed from the year of diagnosis and the year of birth. Participants were stratified by age at diagnosis into <45 years (early-onset) and ≥45 years. Cancer endpoints included: all cancers combined; skin cancer (C43–C44); breast cancer (C50); cervical carcinoma *in situ* (D06); and colorectal cancer (C18–C20). **All of Us.** Cancer outcomes were identified from EHR data using ICD-10 codes recorded in the condition_occurrence table. To align with UK Biobank age stratification while accounting for the All of Us age distribution, participants were stratified by age at diagnosis into 40–45 years and 45–65 years. Age at diagnosis was calculated from the year of condition_start_date and year_of_birth. Cancer endpoints included: all cancers combined; skin cancer (C43–C44); breast cancer (C50); and colorectal cancer (C18–C20).

Control definition. Controls were defined as participants without a documented cancer diagnosis during the observation period, operationalized as absence of ICD-10 codes beginning with “C” (malignant neoplasms) or “D0” (in situ neoplasms). For sex-specific cancer analyses, controls were restricted to the relevant biological sex.

Statistical analysis (primary comparison). For each cohort, cancer endpoint, and age stratum, 2×2 contingency tables were constructed comparing cases versus controls by KDM3C genotype. The primary analysis tested a recessive model by comparing homozygous variant carriers to homozygous reference individuals. **Effect size.** Relative risk (RR) was the primary effect measure:

$$RR = \frac{a/(a+b)}{c/(c+d)}$$

where *a* = cases with homozygous variant genotype, *b* = controls with homozygous variant genotype, *c* = cases with homozygous reference genotype, and *d* = controls with homozygous reference genotype. Odds ratios (OR) were computed as a secondary measure:

$$OR = \frac{a \times d}{b \times c}.$$

Significance testing. Two-sided P values were calculated using Pearson's $\chi^2$ test; Fisher's exact test was used when any expected cell count was <5. P values were not adjusted for multiple comparisons, given the exploratory cross-cohort replication framework; results were interpreted based on consistency of direction and magnitude across cohorts. Cross-cohort comparison. Results were compared qualitatively across cohorts to assess concordance in effect direction, magnitude, and statistical support in populations with differing ancestral composition and ascertainment structures. Analyses were not adjusted for genetic ancestry; results were interpreted in the context of consistency in effect direction and magnitude across cohorts with differing population structures. Computational implementation. UK Biobank analyses were performed in Python 3.10 using pandas (v1.5.x), numpy (v1.24.x), and scipy.stats (v1.10.x), with cohort assembly and extraction performed using SQL queries executed within the UK Biobank analysis environment. All of Us analyses were conducted within the All of Us Researcher Workbench using Python 3.10 notebooks. Data were extracted from controlled-tier datasets using SQL in BigQuery (Google Cloud Platform). Statistical analyses used the same Python libraries to maintain consistency across cohorts.

**Mutational signatures analysis.** All data used for mutational signature analysis were obtained from the National Cancer Institute Genomic Data Commons (GDC) data portal. High-throughput sequencing data in variant call format (VCF), which details somatic mutations identified in tumor samples, along with associated clinical annotations, were retrieved for two TCGA cancer cohorts: TCGA Rectum Adenocarcinoma (TCGA-READ) (29) and TCGA Head and Neck Squamous Cell Carcinoma (TCGA-HNSC) (30). Following data acquisition, the study cohort consisted of 157 READ and 510 HNSC patients. GDC provides pre-processed VCF files generated from four somatic variant-calling pipelines (MuSE, MuTect2, VarScan2, and Pindel). Consistent with prior benchmarking studies demonstrating improved mutation detection with MuTect2 (31,32) and based on its substantially higher mutation counts in our dataset, all downstream analyses were performed on MuTect2-derived calls. Details of the GDC preprocessing pipelines are available in the GDC documentation (NCI GDC Documentation, 2024, (33)).

Signature Assignment. Mutational signatures represent characteristic patterns of somatic alterations generated by distinct mutational processes and are cataloged in resources such as COSMIC (34). To determine the contribution of known SBS signatures to each tumor sample, we used SigProfilerAssignment (35), which decomposes observed mutational profiles into non-negative contributions of reference signatures using a non-negative least squares (NNLS) optimization framework. For each sample, MuTect2-derived mutations were mapped to the GRCh38 reference genome to annotate trinucleotide sequence context, after which single base substitution (SBS) mutational profiles were constructed and assigned to COSMIC signatures.

Signature Analysis. Signature contributions were examined using two complementary measures: frequency, defined as the proportion of patients in whom a signature was detected, and weight, representing the quantitative contribution of the signature within positive samples. For example, SBS1 was detected in all READ tumors (frequency = 1.0) but contributed modestly on average (mean weight= 0.071), whereas SBS3 appeared in only 4.2% of cases but exhibited a substantially higher mean weight (0.26), indicating a stronger effect in those tumors. To identify signatures associated with clinical or molecular features, patients were stratified by variables such as age, BMI, geographic origin, or driver gene mutations (e.g., *APC, TP53, KRAS*), and two-sample t-tests were performed to compare signature weights between groups. Signatures designated as potential sequencing artifacts in COSMIC (34) were excluded from all downstream interpretation (see full list at (https://cancer.sanger.ac.uk/signatures/)). For clarity, additional visualizations were generated focusing on signatures that demonstrated significance in statistical t-tests or high variation among signature frequency with respect to selected relevant clinical data.

**TCGA data analysis.** Germline variant calling was performed for the TCGA Head and Neck Squamous Cell Carcinoma (TCGA-HNSC) and TCGA Rectum Adenocarcinoma (TCGA-READ) cohorts using the Seven Bridges Cancer Genomics Cloud (CGC) platform (www.cancergenomicscloud.org). Briefly, aligned whole-exome sequencing (WES) BAM files were retrieved from the Genomic Data Commons (GDC) and processed through a standardized germline variant calling pipeline. Variants were called using GATK HaplotypeCaller in GVCF mode, followed by joint genotyping and variant quality score recalibration (VQSR) according to GATK best practices. The resulting variant call format (VCF) files were filtered and annotated to extract genotype information at the *KDM3C* locus, enabling classification of samples as WT-S464 (A/A), heterozygous SNP (A/T), or homozygous SNP-S464T (T/T).

Somatic mutation data for rectal adenocarcinoma and head and neck squamous cell carcinoma were obtained from the Genomic Data Commons (GDC). To ensure accurate integration of somatic mutation data with germline genotype data, cBioPortal sample identifiers were systematically cross-referenced with corresponding TCGA sample identifiers retrieved from the GDC project pages for TCGA-READ and TCGA-HNSC, respectively.

Access to controlled-access TCGA data, including potentially identifying germline allele information, was obtained through the TCGA Data Access Committee via dbGaP. In addition, authorization to access somatic single-nucleotide variant data derived from TCGA donors was obtained through dbGaP under approval number

(Project-18630; Title- Mutagenic impact of DNA replication-repair variants). These approved datasets were used to match somatic variant profiles with available genotype information for downstream analysis.

### Supplemental References.

### Supplementary Table Legends

**Supplementary table 1.** Full list of genes (n=161).

**Supplemental table 2.** Genes grouped into pathways based on WebGestalt 2019 classification.

**Supplementary table 3.** Genes with candidate variants identified by WES (n= 77).

**Supplementary table 4.** 2 × 2 contingency tables generated for each pathway

**Supplementary table 5.** Pathway-level enrichment summary: total genes, variant-containing genes, Fisher's exact test p-value, and FDR-adjusted q value.

**Supplementary table 6.** Germline variants found in LARC cohort (n=30) after WES analysis.

**Supplementary Table 7.** Pathway-level enrichment summary in Complete Responders (CRs): total genes, variant-containing genes, Fisher's exact test p-value, and FDR-adjusted q value.

**Supplementary Table 8.** Pathway-level enrichment summary in Poor Responders (PoRs): total genes, variant-containing genes, Fisher's exact test p-value, and FDR-adjusted q value.

**Supplementary table 9.** Clinical and demographic characteristics of LARC cohort (n=84).

**Supplementary table 10.** Clinical and demographic characteristics of HNSCC cohort (n=90).

**Supplementary table 11.** Allele frequency of SNPs in LARC patients (n=30). GnomAD version used is gnomAD v4.1.1.

**Supplementary table 12.** Expected genotype frequencies of KDM3C rs10761725 across gnomAD ancestry groups.

**Supplementary Table 13.** Somatic mutations in Rectal and HNSC separated by *KDM3C* genotype.

**Supplementary table 14.** Phosphorylation sites detected in MDC1 in CAL-27 cells bearing KMD3C WT-S464 or SNP-S464T at baseline and 1h post 10Gy IR.

**Supplementary table 15.** Protein-pair models with ipTM ≥ 0.6 and ipSAE ≥0.4 out of 50 models.

**Supplementary table 16.** NetPhos prediction results.

**Supplementary table 17.** Analysis of baseline differentially expressed genes across KDM3C SNP-S464T vs. WT-S464 isogenic models.

**Supplementary table 18.** Bulk RNA-sequencing results.

**Supplementary table 19.** Primer sequence used for SNP genotyping.

**Supplementary table 20.** Primer sequences for target genes used in the study.

### Supplementary Figure Legends.

**Supplementary Figure 1. (A)** Pathway enrichment of variant-containing genes in LARC complete responders (CRs).

**Supplementary Figure 2. MDC1 phosphoproteomic landscape in CAL-27 cells bearing KDM3C WT-S464 and SNP-S464T cells.** Schematic of MDC1 domain architecture and phosphorylation mapping in CAL-27 cells bearing KDM3C WT-S464 and SNP-S464T cells. Linear stick diagram of MDC1 highlighting the FHA domain (yellow), SDTD or Ser-Asp-Thr-Asp repeats motifs (black), TQXF motifs (brown), PST or Pro-Ser-Thr rich region (teal), and the tandem BRCT domain (pink). Heatmap showing MDC1 phosphorylation sites identified by nanoLC/MS in CAL-27 cells expressing KDM3C WT-S464 or SNP-S464T at baseline (or no IR) and post IR. Each column represents an individual phosphorylation site. Colored boxes denote detected phosphorylation events, with light brown indicating phosphorylation within IDR, yellow indicating phosphorylation within the FHA region, teal indicating phosphorylation within the PST region, and pink indicating phosphorylation within the BRCT region; white indicates sites not detected.

**Supplementary Figure 3. SBS3 in READ and HNSC tumors bearing *KDM3C* T/T (SNP-S464T) and A/A (WT-S464) alleles.** VCF files, from the MuTect2 pipeline, containing the protected germline data from TCGA HNSC patients (n=510) were used. SigProfilerAssignment decomposed the mutational spectra of individual tumors, presented by *KDM3C* genotypes (A/A WT-S464, A/T heterozygous, T/T homozygous SNP-S464T).

**Supplementary Figure 4. Comparative KDM3 family domain organization.** Schematic representation of domain organization in KDM3A, KDM3B, and KDM3C. All three KDM3 paralogs contain an N-terminal SH3-like domain, a zinc-binding region, an LXXLL motif, and a C-terminal catalytic domain, but differ substantially in overall protein length. KDM3C contains an expanded IDR region separating the N-terminal SH3-like domain from the zinc-binding and catalytic domains, supporting potential paralog-specific regulatory or protein-interaction functions. Domain boundaries are indicated by amino acid positions.

**Supplementary Figure 5. Baseline gene-expression and pathway differences in KDM3C SNP-S464T isogenic models.** Bulk RNA-sequencing was performed in isogenic RCM-1, CAL-27, and SCC-9 cells comparing *KDM3C* SNP-S464T cells to WT-S464 cells at baseline. Venn diagrams showing overlap of baseline differentially expressed genes in RCM-1, CAL-27, and SCC-9 cells. **(A)** Genes downregulated in SNP-S464T cells compared with WT-S464 cells, with *DAPK1* identified as the only gene commonly downregulated across all three isogenic models. **(B)** Genes upregulated in SNP-S464T cells compared with WT-S464 cells, with *CXCL8* identified as the only gene commonly upregulated across all three models. Bar plots show significantly enriched upregulated and downregulated pathways in SNP-S464T cells compared to WT-S464 cells in **(C)** RCM-1, **(D)** CAL-27, and **(E)** SCC-9 cells. Bulk RNA-sequencing and bioinformatic analysis were performed by Novogene Corporation. Enriched pathways are reported using FDR-corrected q-values. *$p < 0.05$; n = 2 technical replicates.

**Supplementary Figure 6. Cell-cycle distribution and γH2AX positivity during S-phase progression in CAL-27 isogenic cells.** S-phase distribution of CAL-27 SNP-S464T and WT-S464 isogenic cells at **(A)** baseline and **(B)** treated with 5μM cisplatin. Cells were synchronized by double-thymidine block and released; S-phase percentages were measured at the indicated timepoints. Error bars=SEM; n=3 biological replicates, ns= non-significant, *$p<0.05$, **$p<0.01$; unpaired two-tailed t-test with Welch's correction.

**Supplementary Figure 7. KDM3C SNP-S464T is associated with genotoxic stress-induced inflammatory signaling and cell death in HNSCC models.** Bulk RNA-sequencing shows that immune-related pathways were enriched in **(A)** CAL-27 and **(B)** SCC-9 cells bearing KDM3C SNP-S464T vs. WT-S464 24h after 2Gy IR. Bulk

RNA-sequencing and bioinformatic analysis was performed by Novogene Corporation. Enriched pathways reported here are by corrected False Discovery Rate (FDR) q-values; *p<0.05 for technical replicates, n=2. **(C)** Quantification of cGAS-dsDNA colocalization in CAL-27 cells at baseline and 72h after 5Gy IR along with their representative images. Scale bar: 50μm. Error bars=SEM; n=3 biological replicates **(D)** Immunoblot shows increase in pTBK1 levels in SCC-9 cells bearing *KDM3C* SNP-S464T vs. WT-S464. **Left:** Protein lysates were prepared from untreated or irradiated SCC-9 WT-S464 and SNP-S464T cells and immunoblot analysis of $pTBK1^{Ser172}$ and total TBK1 in SCC-9 cells was performed at indicated timepoints after IR. **Right:** Densitometry analysis of pTBK1/total TBK1 shows increase in $pTBK1^{S172}$ after DNA damage. Increase in cisplatin induced **(E)** % positive early apoptotic cells and **(F)** % $PI^+$ dead cells were observed in CAL-27 cells bearing SNP-S464T vs. WT-S464. The isogenic cell lines were treated with 10μM cisplatin and analyzed by Annexin V/PI staining. Error bars=SEM; n=3 biological replicates, ns= non-significant; *p<0.05, **p<0.01, ***p<0.001; unpaired two-tailed t-test with Welch's correction.

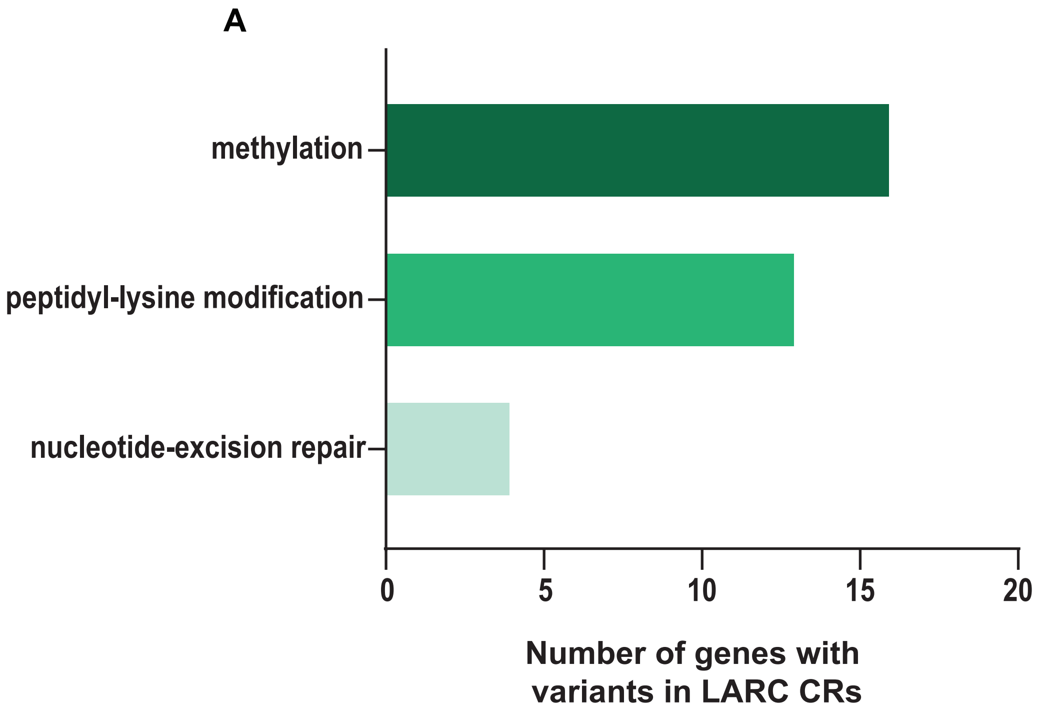

Supplementary Figure 1. Hasan et al.

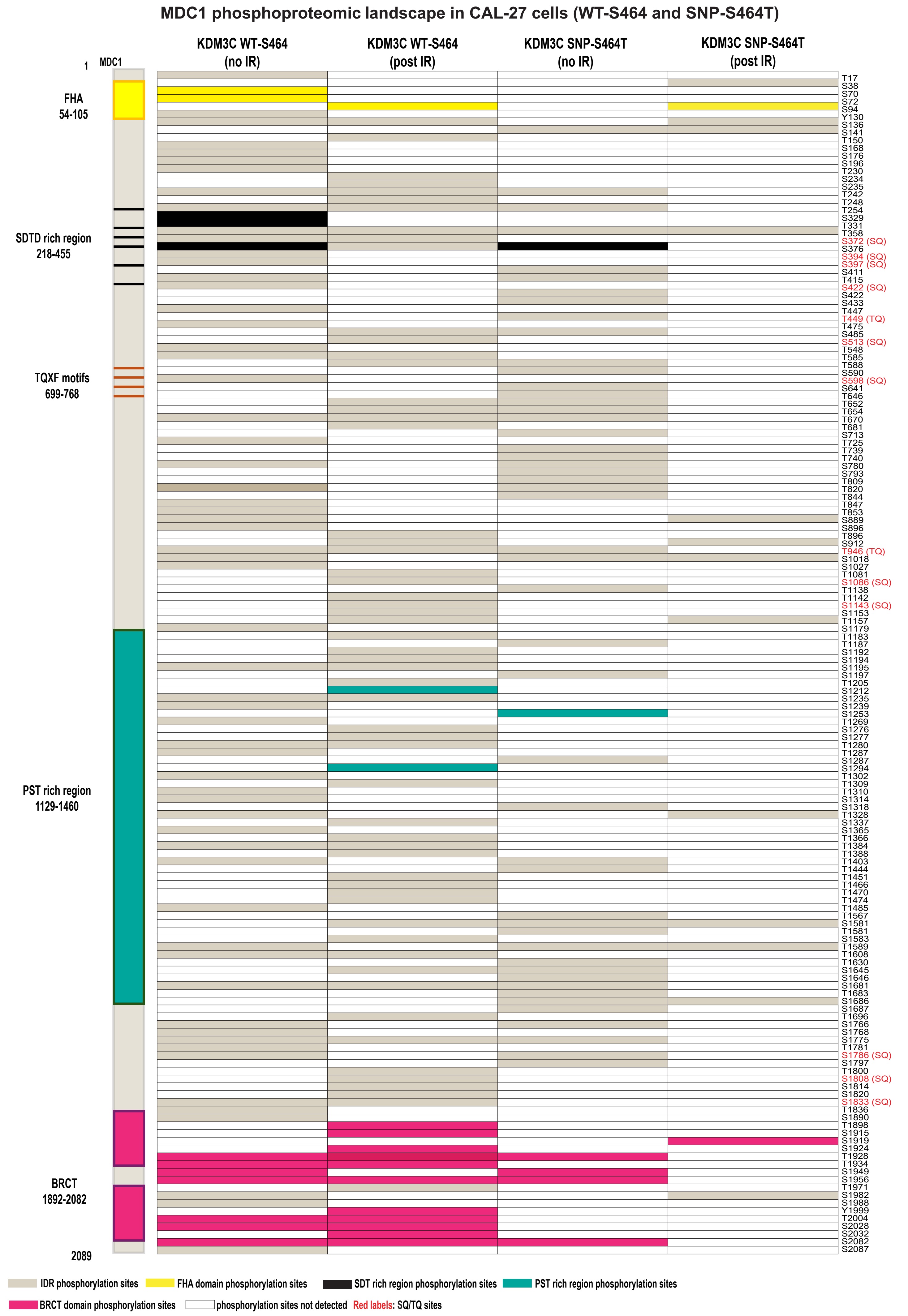

Supplementary Figure 2. Hasan et al.

A

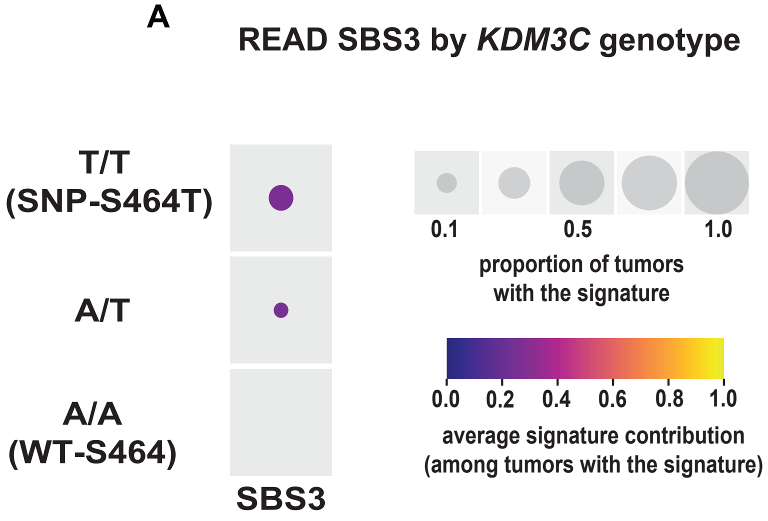

B

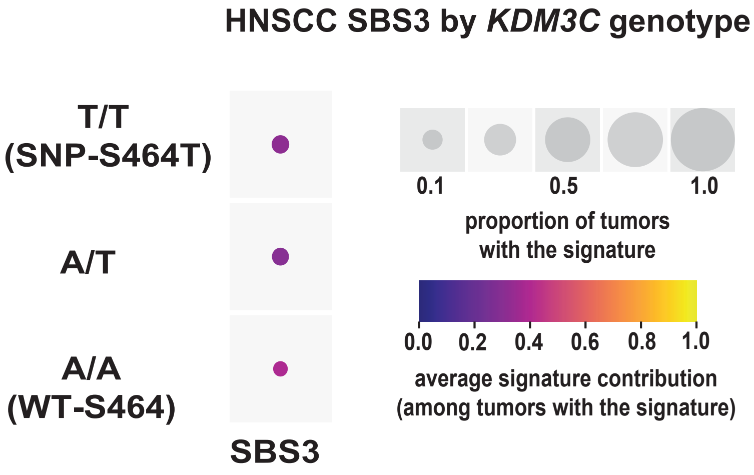

Supplementary Figure 3. Hasan et al.

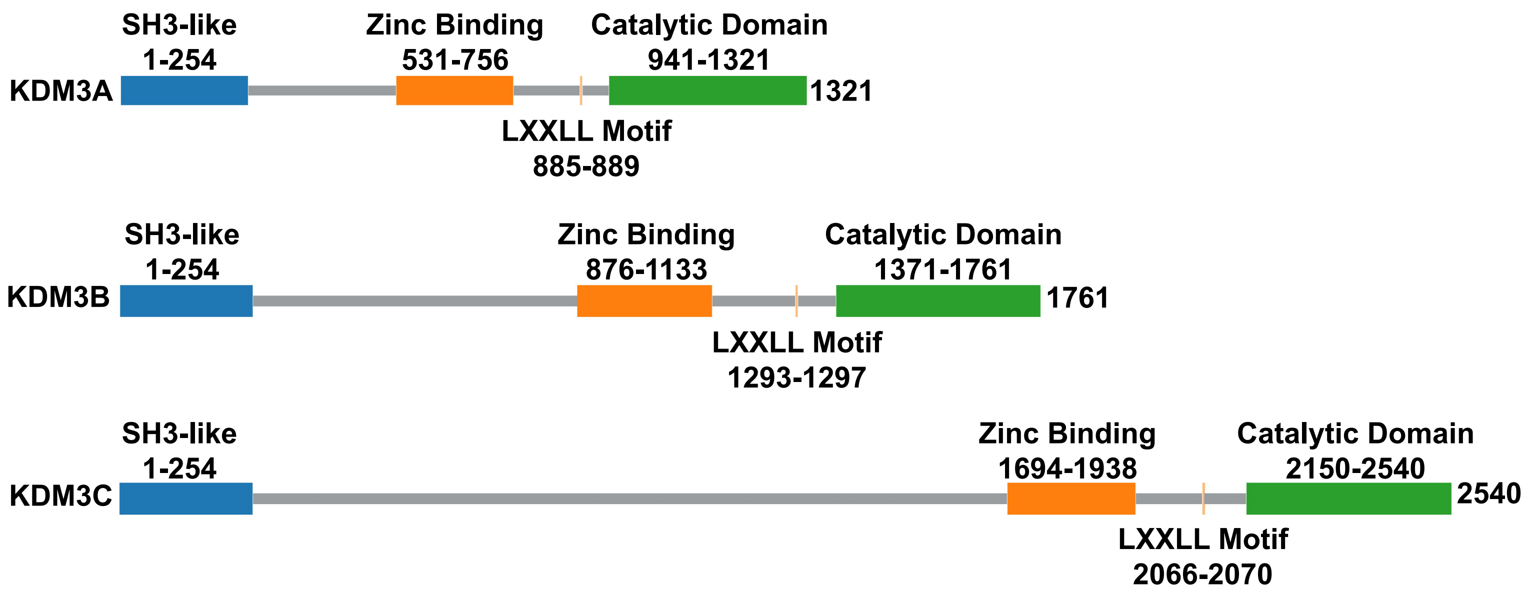

Supplementary Figure 4. Hasan et al.

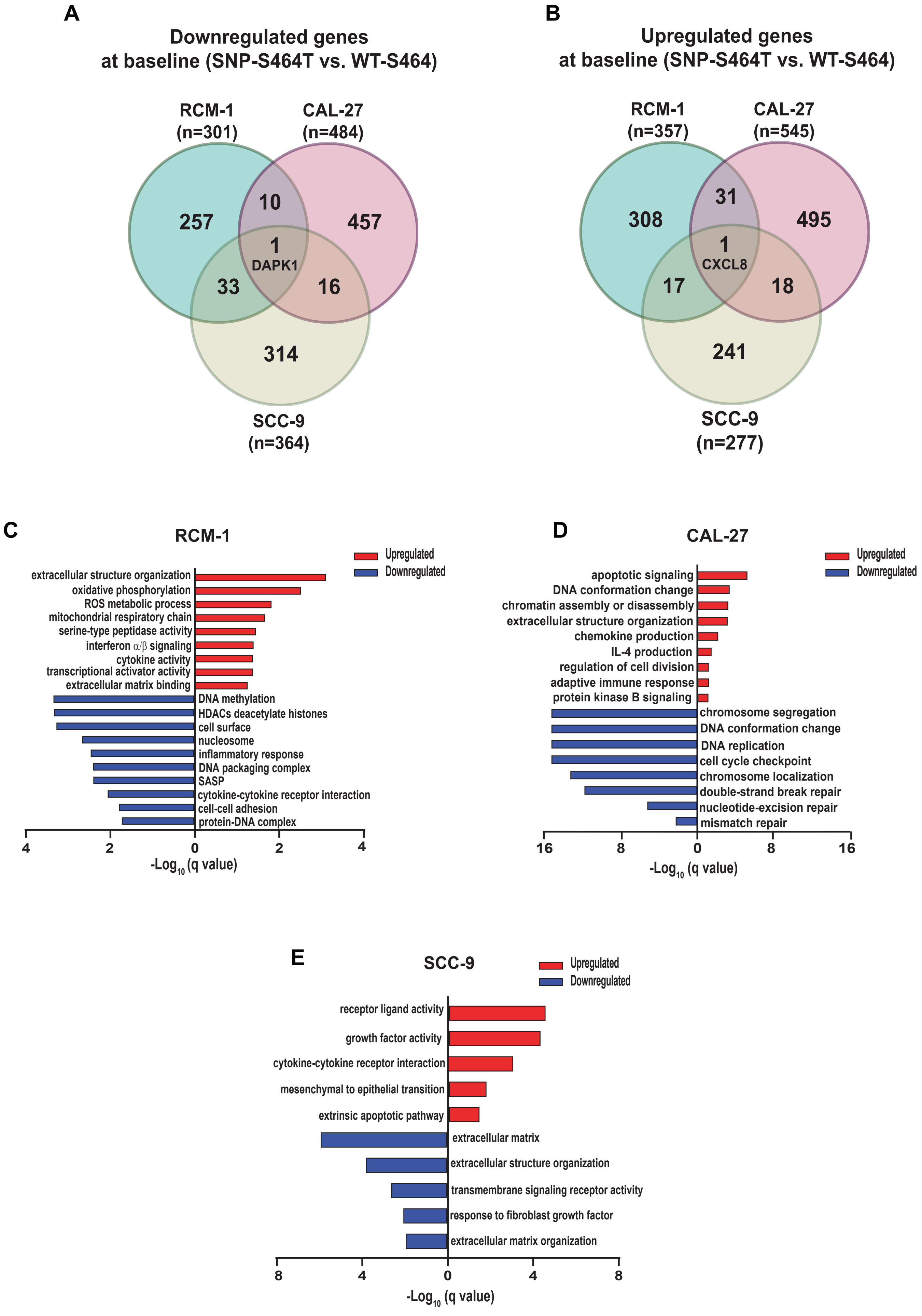

Supplementary Figure 5. Hasan et al.

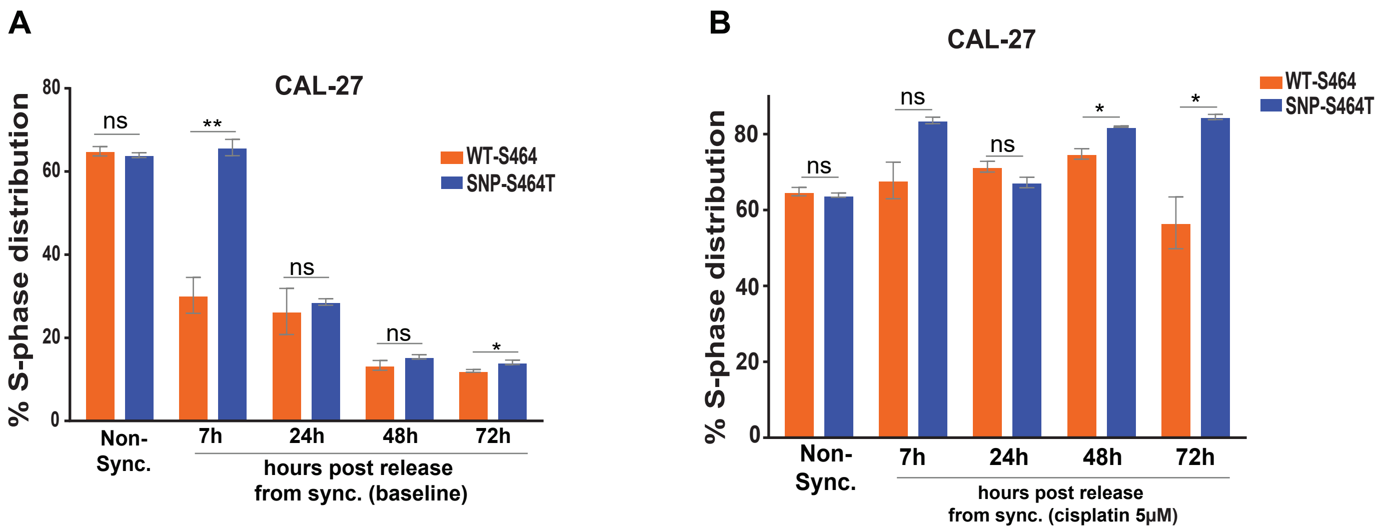

Supplementary Figure 6. Hasan et al.

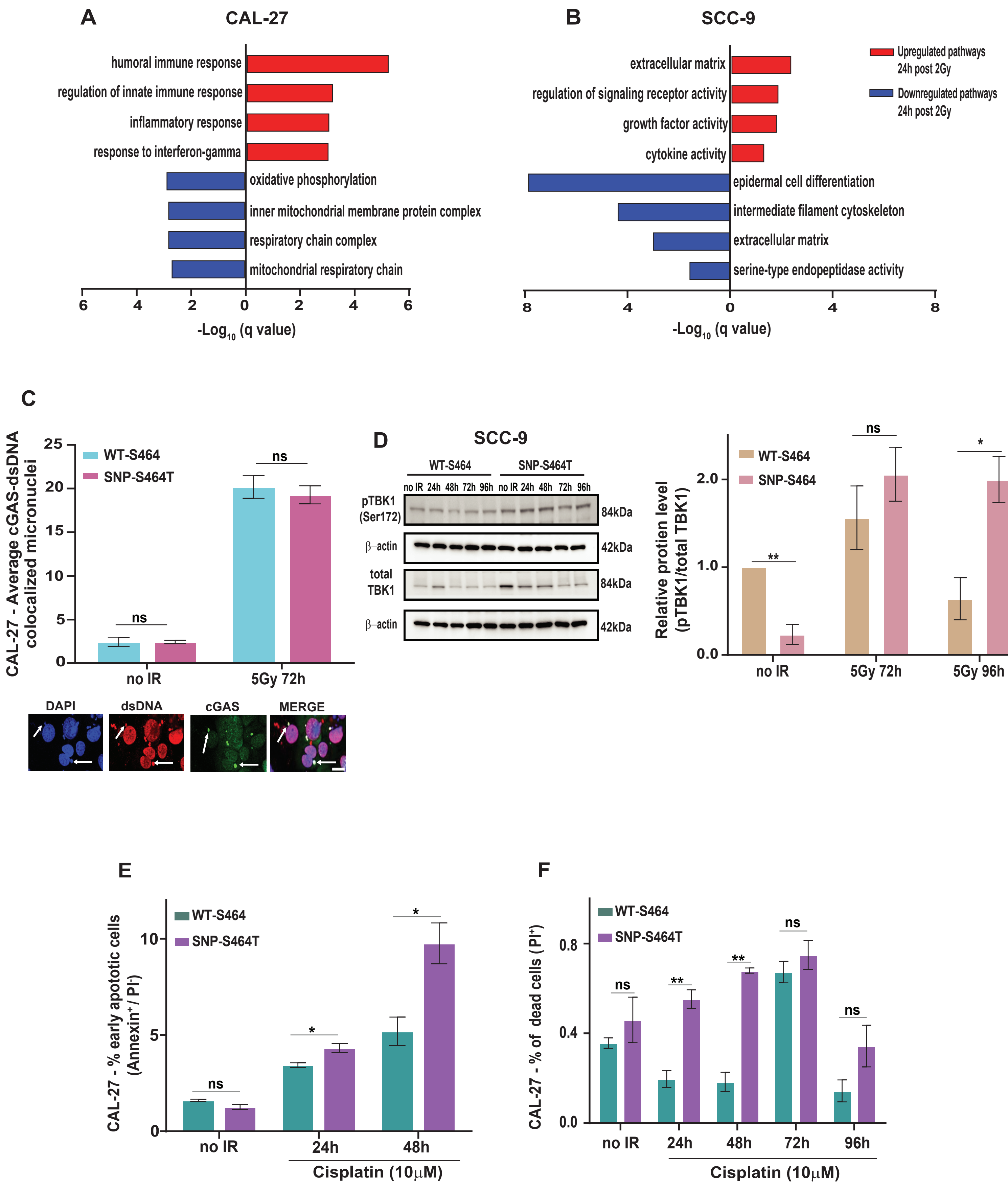

Supplementary Figure 7. Hasan et al.

**Supplementary Table 1. Full list of genes (n=161)**

*APEX2*
*ASH1L*
*ATM*
*ATR*
*ATRIP*
*BLM*
*BRCA1*
*BRCA2*
*BRIP1*
*CCNH*
*CDK7*
*CETN2*
*CHAF1A*
*CHEK1*
*CHEK2*
*DDB1*
*DMC1*
*DOT1L*
*DUT*
*EME1*
*EME2*
*ERCC1*
*ERCC4*
*EZH2*
*FAAP24*
*FAN1*
*FANCA*
*FANCB*
*FANCC*
*FANCD2*
*FANCE*
*FANCF*
*FANCI*
*FANCL*
*FANCM*
*GEN1*
*GTF2H1*
*GTF2H2*
*GTF2H3*
*GTF2H4*
*HELQ*
*HUS1*
*KDM1A*
*KDM1B*
*KDM2A*
*KDM2B*
*KDM3A*
*KDM3B*
*KDM3C*

*KDM4A*
*KDM4B*
*KDM4C*
*KDM4D*
*KDM5A*
*KDM5B*
*KDM5C*
*KDM5D*
*KDM6A*
*KDM6B*
*KDM7A*
*KDM8*
*KMT2A*
*KMT2B*
*KMT2C*
*KMT2D*
*KMT2E*
*KMT5A*
*KMT5B*
*KMT5C*
*LIG1*
*LIG3*
*LIG4*
*MBD4*
*MDC1*
*MGMT*
*MLH1*
*MLH3*
*MMS19*
*MNAT1*
*MPG*
*MSH2*
*MSH3*
*MSH4*
*MSH5*
*MSH6*
*MUS81*
*MUTYH*
*NEIL1*
*NEIL2*
*NEIL3*
*NSD1*
*NSD3*
*OGG1*
*PALB2*
*PARP2*
*PCNA*
*PER1*
*PMS1*
*PMS2*

*PNKP*
*POLB*
*POLD1*
*POLE*
*POLG*
*POLH*
*POLI*
*POLL*
*POLM*
*POLQ*
*PRDM16*
*PRDM2*
*PRDM6*
*PRDM9*
*PRKDC*
*PRPF19*
*RAD1*
*RAD18*
*RAD23A*
*RAD23B*
*RAD50*
*RAD51*
*RAD51B*
*RAD51D*
*RAD52*
*RAD54B*
*RAD54L*
*RAD9A*
*RECQL*
*RECQL4*
*RECQL5*
*RIF1*
*RNF168*
*RNF4*
*RNF8*
*RPA1*
*RPA2*
*RPA3*
*RPA4*
*SETD1A*
*SETD1B*
*SETDB1*
*SHPRH*
*SMUG1*
*SMYD1*
*SMYD2*
*SPO11*
*TDG*
*TDP1*
*TOPBP1*

*TP53*
*TP53BP1*
*UNG*
*UVSSA*
*WRN*
*XAB2*
*XPA*
*XPC*
*XRCC1*
*XRCC2*
*XRCC3*
*XRCC4*

**Supplementary Table 2. Genes grouped into pathways based on WebGestalt 2019 classificatio**

| base-excision repair | mismatch repair | interstrand cross-link repair | nucleotide-excision repair | demethylation | non-recombinational repair |
|---|---|---|---|---|---|
| *APEX2* | *ERCC1* | *ATR* | *CCNH* | *KDM1A* | *ATM* |
| *LIG1* | *LIG1* | *ATRIP* | *CDK7* | *KDM1B* | *BLM* |
| *LIG3* | *MLH1* | *ERCC1* | *CETN2* | *KDM2A* | *BRCA1* |
| *MBD4* | *MLH3* | *FANCA* | *DDB1* | *KDM2B* | *ERCC1* |
| *MPG* | *MSH2* | *FANCB* | *ERCC1* | *KDM3A* | *KDM2A* |
| *MUTYH* | *MSH3* | *FANCC* | *ERCC4* | *KDM3B* | *KDM4D* |
| *NEIL1* | *MSH4* | *FANCD2* | *FANCC* | *KDM3C* | *LIG3* |
| *NEIL2* | *MSH5* | *FANCE* | *GTF2H1* | *KDM4A* | *LIG4* |
| *NEIL3* | *MSH6* | *FANCF* | *GTF2H2* | *KDM4B* | *MDC1* |
| *OGG1* | *PCNA* | *FANCL* | *GTF2H3* | *KDM4C* | *MLH1* |
| *PARP2* | *PMS1* | *FANCM* | *GTF2H4* | *KDM4D* | *OGG1* |
| *POLB* | *PMS2* | *MSH6* | *HUS1* | *KDM5A* | *POLL* |
| *POLD1* | *RPA1* | *MUS81* | *LIG1* | *KDM5B* | *POLM* |
| *POLE* | *RPA2* | *RAD51* | *LIG3* | *KDM5C* | *POLQ* |
| *POLG* | *RPA3* | *RAD51D* | *LIG4* | *KDM5D* | *PRKDC* |
| *POLL* | *RPA4* | *RNF168* | *MMS19* | *KDM6A* | *RAD50* |
| *POLQ* | *TDG* | *RNF8* | *MNAT1* | *KDM6B* | *RAD52* |
| *RECQL4* | *XPC* | *RPA1* | *NEIL1* | *KDM7A* | *RIF1* |
| *RPA1* | | *RPA2* | *NEIL2* | *KDM8* | *RNF168* |
| *RPA2* | | *RPA3* | *NEIL3* | *TDG* | *RNF8* |
| *RPA3* | | *XPA* | *OGG1* | | *XRCC1* |
| *RPA4* | | *XRCC3* | *PCNA* | | *XRCC4* |
| *SMUG1* | | | *PNKP* | | |
| *TDG* | | | *POLB* | | |
| *TP53* | | | *POLD1* | | |
| *UNG* | | | *POLE* | | |
| *WRN* | | | *POLL* | | |
| *XPA* | | | *RAD23A* | | |
| *XRCC1* | | | *RAD23B* | | |
| | | | *RAD52* | | |
| | | | *RPA1* | | |
| | | | *RPA2* | | |
| | | | *RPA3* | | |
| | | | *RPA4* | | |
| | | | *TP53* | | |
| | | | *UVSSA* | | |
| | | | *XAB2* | | |
| | | | *XPA* | | |
| | | | *XPC* | | |
| | | | *XRCC1* | | |

n

| somatic diversification of immune receptors | double-strand break repair | DNA strand elongation | DNA damage response, detection of DNA damage | post replication repair | telomere organization | DNA replication |
|---|---|---|---|---|---|---|
| *ERCC1* | *ATM* | *LIG1* | *DDB1* | *BRCA1* | *ATM* | *ATM* |
| *LIG1* | *BLM* | *LIG3* | *PCNA* | *MSH2* | *ATR* | *ATR* |
| *LIG3* | *BRCA1* | *PCNA* | *PNKP* | *PCNA* | *BLM* | *ATRIP* |
| *LIG4* | *CHEK1* | *POLE* | *POLD1* | *POLD1* | *ERCC1* | *BLM* |
| *MLH1* | *CHEK2* | *RAD50* | *RAD18* | *POLH* | *HUS1* | *BRCA1* |
| *MSH2* | *DMC1* | | *RPA1* | *RAD18* | *PCNA* | *CHEK1* |
| *MSH6* | *EME1* | | *RPA2* | *RPA1* | *PNKP* | *CHEK2* |
| *PMS2* | *EME2* | | *RPA3* | *RPA2* | *POLD1* | *DUT* |
| *POLB* | *ERCC1* | | | *RPA3* | *PRKDC* | *EME2* |
| *POLL* | *FANCB* | | | | *RAD50* | *FANCM* |
| *POLM* | *GEN1* | | | | *RAD51* | *GEN1* |
| *POLQ* | *HUS1* | | | | *RAD51D* | *HUS1* |
| *PRKDC* | *KDM1A* | | | | *RECQL4* | *LIG1* |
| *RIF1* | *KDM2A* | | | | *RIF1* | *LIG3* |
| *RNF168* | *KDM4D* | | | | *RPA1* | *LIG4* |
| *RNF8* | *LIG3* | | | | *RPA2* | *MSH3* |
| *UNG* | *LIG4* | | | | *RPA3* | *MSH6* |
| | *MDC1* | | | | *RPA4* | *PCNA* |
| | *MGMT* | | | | *WRN* | *PNKP* |
| | *MLH1* | | | | *XRCC1* | *POLB* |
| | *MMS19* | | | | *XRCC3* | *POLD1* |
| | *MSH2* | | | | | *POLE* |
| | *MUS81* | | | | | *POLG* |
| | *OGG1* | | | | | *POLH* |
| | *POLL* | | | | | *POLI* |
| | *POLM* | | | | | *POLL* |
| | *POLQ* | | | | | *POLQ* |
| | *PRKDC* | | | | | *RAD50* |
| | *PRPF19* | | | | | *RAD51* |
| | *RAD50* | | | | | *RECQL4* |
| | *RAD51* | | | | | *RECQL5* |
| | *RAD51B* | | | | | *RPA1* |
| | *RAD51D* | | | | | *RPA2* |
| | *RAD52* | | | | | *RPA3* |
| | *RAD54B* | | | | | *RPA4* |
| | *RAD54L* | | | | | *WRN* |
| | *RECQL* | | | | | |
| | *RECQL4* | | | | | |
| | *RECQL5* | | | | | |
| | *RIF1* | | | | | |
| | *RNF168* | | | | | |
| | *RNF8* | | | | | |
| | *RPA1* | | | | | |
| | *RPA2* | | | | | |
| | *RPA3* | | | | | |
| | *RPA4* | | | | | |
| | *SPO11* | | | | | |

*TDP1*
*TP53BP1*
*WRN*
*XRCC1*
*XRCC2*
*XRCC3*
*XRCC4*

| DNA modification | cell cycle checkpoint | DNA conformation change | protein auto ubiquitination | cell cycle G2/M phase transition | chromosome segregation | B cell activation |
|---|---|---|---|---|---|---|
| *BRCA1* | *ATM* | *BLM* | *BRCA1* | *ATM* | *ATM* | *ATM* |
| *KDM1B* | *ATR* | *FANCM* | *RAD18* | *BLM* | *BLM* | *ERCC1* |
| *KMT2A* | *ATRIP* | *GTF2H2* | *RNF4* | *BRCA1* | *BRCA1* | *LIG4* |
| *KMT2E* | *BLM* | *MNAT1* | *RNF8* | *CETN2* | *DMC1* | *MLH1* |
| *MBD4* | *BRCA1* | *RAD50* | | *CHEK1* | *EME2* | *MSH2* |
| *MGMT* | *CHEK1* | *RAD51* | | *CHEK2* | *FANCD2* | *MSH6* |
| *MPG* | *CHEK2* | *RAD54B* | | *HUS1* | *FANCM* | *POLM* |
| *NEIL1* | *EME2* | *RAD54L* | | *KDM8* | *GEN1* | *PRKDC* |
| *NEIL2* | *GEN1* | *RECQL4* | | *MSH6* | *MLH1* | *RIF1* |
| *OGG1* | *HUS1* | *RECQL5* | | *RAD51B* | *MLH3* | *RNF168* |
| *SMUG1* | *MDC1* | *RNF8* | | *RECQL4* | *MMS19* | *RNF8* |
| *TDG* | *MSH2* | *RPA1* | | *TOPBP1* | *MSH4* | *UNG* |
| *UNG* | *MSH6* | *RPA2* | | *TP53* | *MSH5* | |
| | *MUS81* | *RPA4* | | | *MUS81* | |
| | *PCNA* | *SHPRH* | | | *RAD18* | |
| | *PRKDC* | *TP53* | | | *RECQL5* | |
| | *RAD1* | *WRN* | | | *SPO11* | |
| | *RAD9A* | | | | *XRCC3* | |
| | *RPA2* | | | | | |
| | *RPA4* | | | | | |
| | *TOPBP1* | | | | | |
| | *TP53* | | | | | |
| | *XPC* | | | | | |
| | *XRCC3* | | | | | |

| **mitotic cell cycle phase transition** | **regulation of chromosome organization** | **peptidyl-lysine modification** | **cell cycle arrest** | **cyclin-dependent protein kinase activity** | **adaptive immune response** | **response to oxidative stress** |
|---|---|---|---|---|---|---|
| *ATM* | *ATM* | *BRCA1* | *ATM* | *BLM* | *ERCC1* | *ERCC1* |
| *BLM* | *ATR* | *CHEK1* | *BRCA1* | *CCNH* | *KDM5D* | *FANCC* |
| *BRCA1* | *BRCA1* | *KDM3A* | *CDK7* | *CDK7* | *LIG4* | *FANCD2* |
| *CETN2* | *CHEK1* | *KDM3C* | *CHEK2* | *GTF2H1* | *MLH1* | *KDM6B* |
| *CHEK2* | *ERCC1* | *KDM4A* | *KMT2E* | *MNAT1* | *MSH2* | *MGMT* |
| *DDB1* | *GEN1* | *KDM4C* | *MSH2* | | *MSH6* | *NEIL1* |
| *GEN1* | *GTF2H2* | *KDM4D* | *PCNA* | | *RIF1* | *OGG1* |
| *HUS1* | *KDM1A* | *KDM6A* | *TP53* | | *RNF168* | *PCNA* |
| *KDM8* | *KDM3A* | *KMT2A* | | | *RNF8* | *PNKP* |
| *KMT2E* | *KDM3C* | *KMT2B* | | | *UNG* | *RAD52* |
| *MNAT1* | *KDM4A* | *KMT2C* | | | | *TP53* |
| *MSH6* | *KDM4C* | *KMT2D* | | | | *WRN* |
| *PCNA* | *KDM4D* | *KMT2E* | | | | *XPA* |
| *POLE* | *KDM5A* | *KMT5A* | | | | *XRCC1* |
| *PRKDC* | *KMT2A* | *KMT5B* | | | | |
| *RAD51B* | *KMT2B* | *KMT5C* | | | | |
| *RECQL4* | *KMT2E* | *PER1* | | | | |
| *RPA1* | *LIG4* | *RIF1* | | | | |
| *RPA2* | *MNAT1* | | | | | |
| *RPA3* | *PNKP* | | | | | |
| *RPA4* | *RAD50* | | | | | |
| *TOPBP1* | *RIF1* | | | | | |
| *TP53* | *RNF4* | | | | | |
| *XPC* | *XRCC1* | | | | | |
| *XRCC3* | *XRCC3* | | | | | |

| **lymphocyte mediated immunity** | **methylation** | **cell cycle G1/S phase transition** | **epigenetic regulation of gene expression** | **intrinsic apoptotic signaling pathway** | **regulation of cell cycle phase transition** |
|---|---|---|---|---|---|
| *ERCC1* | *BRCA1* | *ATM* | *BRCA1* | *ATM* | *ATM* |
| *KDM5D* | *ASH1L* | *CHEK2* | *CHEK1* | *BRCA1* | *BLM* |
| *LIG4* | *DOT1L* | *KMT2E* | *DOT1L* | *CHEK2* | *BRCA1* |
| *MLH1* | *EZH2* | *MNAT1* | *EZH2* | *KDM1A* | *CETN2* |
| *MSH2* | *KDM1B* | *PCNA* | *KDM1B* | *MLH1* | *CHEK1* |
| *MSH6* | *KDM3A* | *POLE* | *KDM5A* | *MSH2* | *CHEK2* |
| *RIF1* | *KDM4A* | *PRKDC* | *KMT2A* | *MSH6* | *DDB1* |
| *RNF168* | *KDM4C* | *RPA1* | *KMT2B* | *POLB* | *GEN1* |
| *RNF8* | *KDM4D* | *RPA2* | *KMT2D* | *PRKDC* | *HUS1* |
| *UNG* | *KDM6A* | *RPA3* | *RIF1* | *RAD9A* | *KMT2E* |
| | *KMT2A* | *RPA4* | *SETDB1* | *TP53* | *MSH6* |
| | *KMT2B* | *TP53* | *TDG* | *XPA* | *PCNA* |
| | *KMT2C* | | *TP53* | | *PRKDC* |
| | *KMT2E* | | | | *RAD51B* |
| | *KMT5A* | | | | *RECQL4* |
| | *KMT5C* | | | | *RPA2* |
| | *NSD1* | | | | *TOPBP1* |
| | *NSD3* | | | | *TP53* |
| | *PRDM16* | | | | *XPC* |
| | *PRDM2* | | | | *XRCC3* |
| | *PRDM6* | | | | |
| | *PRDM9* | | | | |
| | *SETD1A* | | | | |
| | *SETDB1* | | | | |
| | *SMYD1* | | | | |
| | *SMYD2* | | | | |

| RNA capping | homologous recombination | heterochromatin organization | regulation of chromatin organization | protein monoubiquitination |
|---|---|---|---|---|
| *CCNH* | *BRCA2* | *DOT1L* | *KMT2A* | *DDB1* |
| *CDK7* | *FAAP24* | *EZH2* | *SETD1A* | *FANCL* |
| *GTF2H1* | *FAN1* | *KDM5A* | *SETDB1* | *FANCM* |
| *GTF2H2* | *PALB2* | *KMT2A* | | *KDM2B* |
| *GTF2H3* | *BRIP1* | *KMT2D* | | *RAD18* |
| *GTF2H4* | *FANCI* | *PRDM16* | | *RNF168* |
| *MNAT1* | *HELQ* | *SETDB1* | | |

**Supplementary Table 3. Genes with candidate variants identified by WES (n = 77)**

*ASH1L*
*ATM*
*ATR*
*BRCA1*
*BRCA2*
*BRIP1*
*CDK7*
*CHAF1A*
*DOT1L*
*EME1*
*ERCC4*
*EZH2*
*FAAP24*
*FAN1*
*FANCC*
*FANCI*
*FANCM*
*HELQ*
*KDM1B*
*KDM3A*
*KDM3B*
*KDM3C*
*KDM4A*
*KDM4C*
*KDM4D*
*KDM5A*
*KDM5B*
*KDM6A*
*KDM6B*
*KMT2A*
*KMT2B*
*KMT2C*
*KMT2D*
*KMT2E*
*KMT5A*
*KMT5C*
*LIG1*
*MBD4*
*MLH3*
*MMS19*
*MPG*
*MSH2*
*MSH6*
*MUS81*
*MUTYH*
*NEIL3*
*NSD1*
*NSD3*
*PALB2*
*PARP2*
*PER1*

*POLG*
*POLI*
*POLM*
*POLQ*
*PRDM16*
*PRDM2*
*PRDM6*
*PRDM9*
*PRPF19*
*RAD1*
*RAD50*
*RAD51B*
*RAD51D*
*RECQL*
*SETD1A*
*SETD1B*
*SETDB1*
*SHPRH*
*SMUG1*
*SMYD1*
*SMYD2*
*TDG*
*TP53BP1*
*UVSSA*
*XAB2*
*XRCC1*

**Supplementary Table 4. 2 × 2 contingency tables generated for each pathway**

| DNA damage response, detection of DNA damage | Genes in candidate variant list | Genes not in candidate variant list | | protein auto-ubiquitination |
|---|---|---|---|---|
| Genes in pathway | 0 | 8 | | Genes in pathway |
| Genes outside pathway | 77 | 76 | | Genes outside pathway |

| DNA strand elongation | Genes in candidate variant list | Genes not in candidate variant list | | post replication repair |
|---|---|---|---|---|
| Genes in pathway | 2 | 3 | | Genes in pathway |
| Genes outside pathway | 75 | 81 | | Genes outside pathway |

| cell cycle G1/S phase transition | Genes in candidate variant list | Genes not in candidate variant list | | DNA conformation change |
|---|---|---|---|---|
| Genes in pathway | 2 | 10 | | Genes in pathway |
| Genes outside pathway | 75 | 74 | | Genes outside pathway |

| cell cycle G2/M phase transition | Genes in candidate variant list | Genes not in candidate variant list | | B cell activation |
|---|---|---|---|---|
| Genes in pathway | 4 | 9 | | Genes in pathway |
| Genes outside pathway | 73 | 75 | | Genes outside pathway |

| mitotic cell cycle phase transition | Genes in candidate variant list | Genes not in candidate variant list | | cell cycle arrest |
|---|---|---|---|---|
| Genes in pathway | 5 | 20 | | Genes in pathway |
| Genes outside pathway | 72 | 64 | | Genes outside pathway |

| interstrand cross-link repair | Genes in candidate variant list | Genes not in candidate variant list | | chromosome segregation |
|---|---|---|---|---|
| Genes in pathway | 6 | 16 | | Genes in pathway |
| Genes outside pathway | 71 | 68 | | Genes outside pathway |

| Homologous recombination | Genes in candidate variant list | Genes not in candidate variant list | | heterochromatin organization |
|---|---|---|---|---|
| Genes in pathway | 7 | 0 | | Genes in pathway |
| Genes outside pathway | 70 | 84 | | Genes outside pathway |

| DNA replication | Genes in candidate variant list | Genes not in candidate variant list |
|---|---|---|
| Genes in pathway | 10 | 26 |
| Genes outside pathway | 67 | 58 |

| epigenetic regulation of gene expression |
|---|
| Genes in pathway |
| Genes outside pathway |

| regulation of chromosome organization | Genes in candidate variant list | Genes not in candidate variant list |
|---|---|---|
| Genes in pathway | 14 | 11 |
| Genes outside pathway | 63 | 73 |

| peptidyl-lysine modification |
|---|
| Genes in pathway |
| Genes outside pathway |

| somatic diversification of immune receptors | Genes in candidate variant list | Genes not in candidate variant list |
|---|---|---|
| Genes in pathway | 5 | 12 |
| Genes outside pathway | 72 | 72 |

| protein monoubiquitination |
|---|
| Genes in pathway |
| Genes outside pathway |

| | Genes in candidate variant list | Genes not in candidate variant list |
|---|---|---|
| | 1 | 3 |
| | 76 | 81 |

| cyclin-dependent protein kinase activity | Genes in candidate variant list | Genes not in candidate variant list |
|---|---|---|
| Genes in pathway | 1 | 4 |
| Genes outside pathway | 76 | 80 |

| | Genes in candidate variant list | Genes not in candidate variant list |
|---|---|---|
| | 2 | 7 |
| | 75 | 77 |

| adaptive immune response | Genes in candidate variant list | Genes not in candidate variant list |
|---|---|---|
| Genes in pathway | 2 | 8 |
| Genes outside pathway | 75 | 76 |

| | Genes in candidate variant list | Genes not in candidate variant list |
|---|---|---|
| | 3 | 14 |
| | 74 | 70 |

| response to oxidative stress | Genes in candidate variant list | Genes not in candidate variant list |
|---|---|---|
| Genes in pathway | 3 | 11 |
| Genes outside pathway | 74 | 73 |

| | Genes in candidate variant list | Genes not in candidate variant list |
|---|---|---|
| | 4 | 8 |
| | 73 | 76 |

| mismatch repair | Genes in candidate variant list | Genes not in candidate variant list |
|---|---|---|
| Genes in pathway | 5 | 13 |
| Genes outside pathway | 72 | 71 |

| | Genes in candidate variant list | Genes not in candidate variant list |
|---|---|---|
| | 5 | 3 |
| | 72 | 81 |

| intrinsic apoptotic signaling pathway | Genes in candidate variant list | Genes not in candidate variant list |
|---|---|---|
| Genes in pathway | 5 | 7 |
| Genes outside pathway | 72 | 77 |

| | Genes in candidate variant list | Genes not in candidate variant list |
|---|---|---|
| | 6 | 12 |
| | 71 | 72 |

| non-recombinational repair | Genes in candidate variant list | Genes not in candidate variant list |
|---|---|---|
| Genes in pathway | 7 | 15 |
| Genes outside pathway | 70 | 69 |

| | Genes in candidate variant list | Genes not in candidate variant list |
|---|---|---|
| | 7 | 0 |
| | 70 | 84 |

| DNA modification | Genes in candidate variant list | Genes not in candidate variant list |
|---|---|---|
| Genes in pathway | 8 | 5 |
| Genes outside pathway | 69 | 79 |

| Genes in candidate variant list | Genes not in candidate variant list |
|---|---|
| 10 | 3 |
| 67 | 81 |

| base-excision repair | Genes in candidate variant list | Genes not in candidate variant list |
|---|---|---|
| Genes in pathway | 11 | 18 |
| Genes outside pathway | 66 | 66 |

| Genes in candidate variant list | Genes not in candidate variant list |
|---|---|
| 15 | 3 |
| 62 | 81 |

| double-strand break repair | Genes in candidate variant list | Genes not in candidate variant list |
|---|---|---|
| Genes in pathway | 16 | 31 |
| Genes outside pathway | 61 | 46 |

| Genes in candidate variant list | Genes not in candidate variant list |
|---|---|
| 1 | 5 |
| 76 | 79 |

| RNA capping | Genes in candidate variant list | Genes not in candidate variant list |
|---|---|---|
| Genes in pathway | 1 | 6 |
| Genes outside pathway | 76 | 78 |

| lymphocyte mediated immunity | Genes in candidate variant list | Genes not in candidate variant list |
|---|---|---|
| Genes in pathway | 2 | 8 |
| Genes outside pathway | 75 | 76 |

| regulation of chromatin organization | Genes in candidate variant list | Genes not in candidate variant list |
|---|---|---|
| Genes in pathway | 3 | 0 |
| Genes outside pathway | 74 | 84 |

| telomere organization | Genes in candidate variant list | Genes not in candidate variant list |
|---|---|---|
| Genes in pathway | 5 | 16 |
| Genes outside pathway | 72 | 68 |

| regulation of cell cycle phase transition | Genes in candidate variant list | Genes not in candidate variant list |
|---|---|---|
| Genes in pathway | 5 | 15 |
| Genes outside pathway | 72 | 69 |

| cell cycle checkpoint | Genes in candidate variant list | Genes not in candidate variant list |
|---|---|---|
| Genes in pathway | 7 | 17 |
| Genes outside pathway | 70 | 67 |

| nucleotide-excision repair | Genes in candidate variant list | Genes not in candidate variant list |
|---|---|---|
| Genes in pathway | 9 | 31 |
| Genes outside pathway | 68 | 53 |

| demethylation | Genes in candidate variant list | Genes not in candidate variant list |
|---|---|---|
| **Genes in pathway** | 12 | 8 |
| **Genes outside pathway** | 65 | 76 |

| methylation | Genes in candidate variant list | Genes not in candidate variant list |
|---|---|---|
| **Genes in pathway** | 26 | 0 |
| **Genes outside pathway** | 51 | 84 |

**Supplementary Table 5. Pathway-level enrichment summary: total genes, variant-containing genes**

| Pathways | DNA damage response, detection of DNA damage | protein auto ubiquitination | cyclin-dependent protein kinase activity | RNA capping |
|---|---|---|---|---|
| **total genes out of list of genes (n=161)** | 8 | 4 | 5 | 7 |
| **total genes out of candidate list (n=77)** | 0 | 1 | 1 | 1 |
| **p value from Fisher Exact Test** | 0.0069 | 0.6217 | 0.3695 | 0.1193 |
| **FDR corrected p value (q value)** | 0.0361 | 0.6572 | 0.4404 | 0.1919 |

**pathways with q value < 0.05 were considered to be enriched**

**s, Fisher’s exact test p-value, and FDR-adjusted q value**

| DNA strand elongation | post replication repair | adaptive immune response | lymphocyte mediated immunity | cell cycle G1/S phase transition | DNA conformation change |
|---|---|---|---|---|---|
| 5 | 9 | 10 | 10 | 12 | 17 |
| 2 | 2 | 2 | 2 | 2 | 3 |
| >0.9999 | 0.1711 | 0.1017 | 0.1017 | 0.0338 | 0.0097 |
| 0.9999 | 0.2532 | 0.1881 | 0.1881 | 0.0962 | 0.0399 |

| response to oxidative stress | regulation of chromatin organization | cell cycle G2/M phase transition | B cell activation | mismatch repair | somatic diversification of immune receptors |
|---|---|---|---|---|---|
| 14 | 3 | 13 | 12 | 18 | 17 |
| 3 | 3 | 4 | 4 | 5 | 5 |
| 0.05 | 0.1072 | 0.2529 | 0.3748 | 0.0833 | 0.1285 |
| 0.1156 | 0.1889 | 0.3466 | 0.4404 | 0.1712 | 0.1981 |

| telomere organization | mitotic cell cycle phase transition | cell cycle arrest | intrinsic apoptotic signaling pathway | regulation of cell cycle phase transition | interstrand cross-link repair | chromosome segregation |
|---|---|---|---|---|---|---|
| 21 | 25 | 8 | 12 | 20 | 22 | 18 |
| 5 | 5 | 5 | 5 | 5 | 6 | 6 |
| 0.0201 | 0.0039 | 0.4811 | 0.7684 | 0.0329 | 0.0416 | 0.219 |
| 0.0676 | 0.0302 | 0.5236 | 0.7897 | 0.0962 | 0.1026 | 0.3117 |

| non-recombinational repair | cell cycle checkpoint | homologous recombination | heterochromatin organization | DNA modification | nucleotide-excision repair |
|---|---|---|---|---|---|
| 22 | 24 | 7 | 7 | 13 | 40 |
| 7 | 7 | 7 | 7 | 8 | 9 |
| 0.115 | 0.0748 | 0.0049 | 0.0049 | 0.3892 | 0.0002 |
| 0.1919 | 0.1628 | 0.0302 | 0.0302 | 0.4404 | 0.0037 |

| DNA replication | epigenetic regulation of gene expression | base-excision repair | demethylation | regulation of chromosome organization | peptidyl-lysine modification |
|---|---|---|---|---|---|
| 36 | 13 | 29 | 20 | 25 | 18 |
| 10 | 10 | 11 | 12 | 14 | 15 |
| 0.0078 | 0.0408 | 0.3055 | 0.3391 | 0.3928 | 0.0019 |
| 0.0361 | 0.1026 | 0.4037 | 0.4326 | 0.4404 | 0.0234 |

| double-strand break repair | methylation | protein monoubiquitination |
|---|---|---|
| 54 | 26 | 6 |
| 16 | 26 | 1 |
| 0.0138 | <0.0001 | 0.2126 |
| 0.0511 | 0.0037 | 0.3082 |

**Supplementary Table 6. Germline variants found in LARC cohort (n=30) after WES analysis**

| Patient | Gene | Position | Ref | Alt | AA Change |
|---|---|---|---|---|---|
| **Complete responders** | | | | | |
| p23 | *ATM* | rs150757822 | A | C | K1992T |
| p17, p18, p26, p30 | *BRCA1* | rs1799950 | T | C | Q356R |
| p30 | *BRCA2* | rs28897745 | A | G | D2665G |
| p26 | *ERCC4* | rs150244523 | T | C | L313P |
| p18 | *ERCC4* | rs1800124 | A | G | E875G |
| p17 | *FAAP24* | rs36017455 | C | T | S126F |
| p29 | *FAAP24* | rs148106526 | C | T | T212M |
| p29 | *FAN1* | rs771137282 | G | A | V687I |
| p30 | *FANCM* | rs61746943 | C | T | T1600I |
| p24 | *FANCM* | rs147021911 | C | T | Q1701X |
| p28 | *MBD4* | rs2307293 | C | G | D568H |
| p16 | *MMS19* | rs36023427 | C | T | G1029D |
| p17 | *MMS19* | rs29001280 | G | A | R98W |
| p25 | *MPG* | rs145824088 | G | A | V179I |
| p26 | *MSH2* | rs63750875 | G | C | A636P |
| p20 | *MUS81* | rs34891773 | C | T | R351W |
| p24 | *NEIL3* | rs34007209 | C | T | R38C |
| p18 | *PALB2* | rs45624036 | C | T | V932M |
| p18 | *PARP2* | rs3093921 | A | G | D235G |
| p26 | *PER1* | rs72845601 | G | A | P37S |
| p26 | *POLI* | rs3218786 | T | C | F532S |
| p24, p30 | *POLQ* | rs3218634 | G | C | L2538V |
| p24, p30 | *POLQ* | rs532411 | G | A | A2304V |
| p27 | *RAD50* | rs28903086 | G | A | V127I |
| p24 | *RAD51D* | rs28363284 | T | C | E233G |
| p22 | *SHPRH* | . | C | T | R1479H |
| p28 | *TP53BP1* | rs45482998 | A | G | V1031A |
| p17 | *TP53BP1* | rs148009445 | T | C | Q308R |
| p30 | *XRCC1* | rs2307186 | C | A | R7L |
| p18 | *DOT1L* | rs144165419 | G | A | A1053T |
| p18, p21 | *DOT1L* | rs113164803 | C | A | L1379M |
| p21 | *KDM3C* | rs200797706 | A | G | L1458S |

| | | | | | |
|---|---|---|---|---|---|
| p22 | *KDM3C* | rs200160728 | G | A | P1089S |
| p17, p18, p19, p20, p21, p22, p23, p24, p25, p26, p27, p28, p29, p30 | *KDM3C* | rs10761725 | A | T | S464T |
| p19 | *KDM3A* | rs143405154 | G | T | C417F |
| p29, p30 | *KDM3B* | rs200772506 | G | A | A5T |
| p25 | *KDM4A* | rs138721228 | C | T | A493V |
| p30 | *KDM4C* | rs34369202 | T | C | I603T |
| p25 | *KDM4D* | rs144086807 | C | G | P457A |
| p23 | *KDM5A* | rs781474100 | C | T | R1282H |
| p22, p26 | *KDM5B* | rs112284833 | C | G | E939Q |
| p20 | *KDM6B* | rs148641957 | G | T | V209L |
| p25 | *KMT2A* | rs9332745 | C | G | A30G |
| p24 | *KMT2A* | rs778675942 | C | T | R3228C |
| p21 | *KMT2A* | rs199656572 | C | G | P3531A |
| p23 | *KMT2B* | . | G | A | E117K |
| p22 | *KMT2C* | rs142997680 | T | A | T3575S |
| p19 | *KMT2C* | rs755062207 | C | T | G2986D |
| p25 | *KMT2D* | rs75937132 | T | C | M3398V |
| p26 | *KMT2E* | rs200289007 | A | T | M316L |
| p18 | *KMT2E* | rs145923995 | A | G | N1195S |
| p22 | *KMT2E* | rs74959149 | G | A | S1532N |
| p26 | *KMT5C* | rs200852152 | C | A | P328H |
| p17 | *NSD1* | rs768074196 | C | T | R2195C |
| p21, p23, p28, p29 | *NSD1* | rs3733875 | G | T | V345L |
| p20 | *NSD3* | rs185811622 | G | A | S754L |
| p24 | *PRDM16* | rs201904226 | G | A | G818S |
| p20 | *SETD1A* | . | C | T | R694W |
| p20 | *SETD1A* | rs374966225 | C | T | T1122M |
| p21 | *SETD1B* | rs58801491 | T | C | L1355P |
| p20 | *SMYD1* | rs565724752 | G | A | V258I |
| **Poor responders** | | | | | |
| p4 | *ASH1L* | rs78904392 | T | C | N1941D |
| p1, p3, p6, p8, p9 | *EZH2* | rs2302427 | C | G | D185H |
| p2, p9 | *KDM3C* | rs34491125 | G | C | D2400E |
| p1 | *KDM3C* | rs368852030 | T | C | E1577G |
| p14 | *KDM1B* | rs149734475 | A | C | Q162P |
| p7 | *KDM4C* | rs111451391 | A | C | S534R |
| p13 | *KDM4C* | rs191848178 | G | T | A630S |
| p4 | *KDM4C* | rs35389625 | A | G | N697S |
| p14 | *KDM4D* | rs372591204 | C | T | R451C |
| p3 | *KDM5A* | rs200969654 | G | C | S103C |
| p3 | *KDM6A* | rs138723332 | T | C | C520R |
| p1, p2, p9, p10 | *KDM6A* | rs2230018 | C | A | T778K |
| p1 | *KDM6B* | rs79548905 | G | C | E221D |
| p7 | *KDM6B* | rs749981813 | G | T | R988L |
| p11 | *KMT2C* | rs138119145 | C | G | D1319H |
| p3 | *KMT2D* | rs201481646 | C | T | R5266H |
| p9 | *KMT2D* | rs112921115 | G | A | T1246M |
| p9 | *KMT2D* | rs201994402 | G | C | R228G |
| p6 | *KMT5A* | rs376869233 | G | A | A108T |

| | | | | | |
|---|---|---|---|---|---|
| p1 | *PRDM16* | rs187194973 | G | A | A34T |
| p8 | *PRDM16* | rs554385722 | G | A | A643T |
| p13 | *PRDM16* | rs149333409 | G | A | V764M |
| p7 | *PRDM16* | rs201814961 | C | T | P889L |
| p11 | *PRDM2* | rs41269803 | G | A | C334Y |
| p2 | *PRDM2* | rs116238585 | C | G | P949A |
| p3, p15 | *PRDM6* | rs139705595 | A | G | I262V |
| p8 | *PRDM9* | rs202118545 | C | G | T769R |
| p1 | *PRDM9* | rs77287813 | A | C | N790H |
| p12 | *SETD1B* | rs773794483 | A | C | H8P |
| p7 | *SETD1B* | rs113279920 | C | T | P367L |
| p12 | *SETD1B* | rs144871159 | G | A | A1488T |
| p7 | *SETDB1* | rs150892641 | C | T | A1021V |
| p7 | *SMYD2* | rs34259050 | A | G | M384V |
| p2, p12 | *ATR* | rs77208665 | T | C | K764E |
| p10 | *BRCA2* | rs80358431 | C | G | S476C |
| p8 | *BRIP1* | rs587781325 | A | G | Y369H |
| p1, p3, p12 | *CDK7* | rs34584424 | C | T | T285M |
| p6 | *CHAF1A* | rs369272426 | C | T | T904M |
| p7 | *EME1* | rs148490439 | C | A | P245Q |
| p4, p15 | *ERCC4* | rs1800067 | G | A | R415Q |
| p6 | *FANCC* | rs1800361 | G | A | S26F |
| p9 | *FANCI* | rs149223439 | G | C | G53A |
| p9 | *FANCI* | rs151038616 | T | A | L619Q |
| p9, p10, p13 | *FANCM* | rs45547534 | A | G | I208M |
| p2 | *HELQ* | rs113520876 | T | A | Y876F |
| p9 | *LIG1* | rs3730947 | C | T | V349M |
| p4 | *MLH3* | rs138006166 | C | T | A1394T |
| p7 | *MMS19* | rs29001311 | T | G | Q409P |
| p8 | *MSH6* | rs63749999 | C | T | R1035X |
| p1 | *MUS81* | rs61754785 | G | A | R432H |
| p12 | *MUTYH* | rs150792276 | G | A | R423C |
| p13 | *MUTYH* | rs200872702 | T | C | I220V |
| p3 | *POLG* | rs113994096 | G | A | P587L |
| p2 | *POLI* | rs3218784 | A | G | I261M |
| p10 | *POLI* | rs200852409 | C | G | P390R |
| p13 | *POLM* | rs28382644 | C | G | G220A |
| p9 | *PRPF19* | rs188856023 | G | T | T91N |
| p4, p15 | *RAD1* | rs2308957 | C | T | G114D |

| | | | | | |
|---|---|---|---|---|---|
| p10 | *RAD51B* | rs34094401 | T | G | L172W |
| p14 | *RAD51B* | rs34594234 | A | G | K243R |
| p6 | *RECQL* | . | G | A | P126L |
| p13 | *SMUG1* | rs3136389 | G | A | R105W |
| p3 | *TDG* | rs4135113 | G | A | G199S |
| p15 | *TP53BP1* | rs28903077 | C | T | R1454Q |
| p8 | *UVSSA* | rs116741007 | G | A | G355R |
| p15 | *UVSSA* | rs147609449 | C | T | R445W |
| p15 | *XAB2* | rs61761630 | C | T | E303K |

| Location |
| --- |
| outside of the known functional domains |
| outside of the known functional domains |
| outside of the known functional domains |
| outside of the known functional domains |
| outside of the known functional domains |
| outside of the known functional domains |
| outside of the known functional domains |
| outside of the known functional domains |
| outside of the known functional domains |
| outside of the known functional domains |
| outside of the known functional domains |
| outside of the known functional domains |
| inal Dos2-interacting transcription regulator of RNA-Pol-II (50 |
| Methylpurine-DNA glycosylase (MPG) (89 - 283) |
| MutS domain V (619 - 853) |
| ERCC4 nuclease domain (273 - 413) |
| outside of the known functional domains |
| outside of the known functional domains |
| outside of the known functional domains |
| outside of the known functional domains |
| outside of the known functional domains |
| DNA polymerase family A domain (2313 - 2584) |
| outside of the known functional domains |
| outside of the known functional domains |
| Rad51 (74 - 258) |
| Zinc finger, C3HC4 type (RING finger) (1432 - 1478) |
| outside of the known functional domains |
| outside of the known functional domains |
| N-terminal domain (1 - 151) |
| outside of the known functional domains |
| outside of the known functional domains |
| outside of the known functional domains |

outside of the known functional domains
outside of the known functional domains

outside of the known functional domains
outside of the known functional domains
outside of the known functional domains
outside of the known functional domains
outside of the known functional domains
outside of the known functional domains
PLU-1-like domain (756 - 1088)
outside of the known functional domains
outside of the known functional domains
outside of the known functional domains
outside of the known functional domains
outside of the known functional domains
outside of the known functional domains
outside of the known functional domains
outside of the known functional domains
outside of the known functional domains
outside of the known functional domains
outside of the known functional domains
outside of the known functional domains
outside of the known functional domains
Pro-Trp-Trp-Pro (PWWP) domain (321 - 414)
outside of the known functional domains
outside of the known functional domains
outside of the known functional domains
outside of the known functional domains
outside of the known functional domains
outside of the known functional domains

outside of the known functional domains
outside of the known functional domains
JmjC hydroxylase domain (2382 - 2481)
outside of the known functional domains
CW-type Zinc Finger (137 - 192)
outside of the known functional domains
outside of the known functional domains
outside of the known functional domains
outside of the known functional domains
ARID/BRIGHT DNA binding domain (82 - 170)
outside of the known functional domains
outside of the known functional domains
outside of the known functional domains
outside of the known functional domains
outside of the known functional domains
FYR-C domain (5236-5321)
outside of the known functional domains
outside of the known functional domains
outside of the known functional domains

outside of the known functional domains
outside of the known functional domains
outside of the known functional domains
outside of the known functional domains

outside of the known functional domains
outside of the known functional domains
SET domain (260 - 365)
Zinc-finger double domain (762 - 785)
Zinc-finger double domain (790 - 815)
outside of the known functional domains
outside of the known functional domains
outside of the known functional domains
outside of the known functional domains
outside of the known functional domains
outside of the known functional domains
outside of the known functional domains
DEAD_2 helicase domain (248 - 415)

Protein kinase domain (12 - 295)
CAF1 complex subunit p150, region binding to CAF1-p60 at C-term (665 - 956)
outside of the known functional domains
outside of the known functional domains

Fanconi anaemia group C protein (1 - 558)

FANCI solenoid 1 cap domain (1 - 53)
FANCI helical domain 2 (555 - 787)
DEAD/DEAH box helicase domain (101 - 254)
outside of the known functional domains
N-terminal domain (287 - 465)
outside of the known functional domains

outside of the known functional domains
MutS domain III (738 - 1064)

outside of the known functional domains
NUDIX domain (382 - 491)
PD superfamily base excision DNA repair protein domain (12[illegible]

outside of the known functional domains

outside of the known functional domains
impB/mucB/samB family C-terminal domain (316 - 439)
outside of the known functional domains
Prp19/Pso4-like domain (67 - 134)
Repair protein Rad1/Rec1/Rad17 (17 - 257)

Rad51 (82 - 342)

Rad51 (82 - 342)

DEAD/DEAH box helicase domain (94 - 259)
Uracil DNA glycosylase superfamily domain (71 - 262)
Uracil DNA glycosylase superfamily domnain (131 - 278)

outside of the known functional domains
outside of the known functional domains
outside of the known functional domains
outside of the known functional domains

| ClinVar | SIFT prediction |
|---|---|
| Conflicting interpretations, familial breast cancer, ataxia-telangiectasia syndrome | T |
| Benign, hereditary breast and ovarian cancer predisposing | D |
| Benign, hereditary breast and ovarian cancer predisposing | D |
| not provided, highly penetrant | D |
| not provided, highly penetrant, Fanconi anemia complementation group Q, Xeroderma pigmentosum group F,Cockayne syndrome | D |
| Benign | . |
| | . |
| | D |
| Benign, Falconi anemia | D |
| Likely pathogenic/x2c risk factor, Falconi anemia, familial breast cancer, spermatogenic failure, premature ovarian failure, ovary neoplasm | . |
| Uncertain significance | D |
| | D |
| [illegible] - 311) | D |
| | T |
| Pathogenic, Lynch syndrome, colorectal cancer | T |
| | D |
| | D |
| Benign, familial breast cancer, PALB2-related cancer susceptibility | T |
| | D |
| | D |
| | D |
| | D |
| | D |
| Conflicting interpretations, Nijmegen breakage syndrome-like disorder, hereditary cancer-predisposing | T |
| Benign, hereditary breast and ovarian cancer-predisposing | T |
| | D |
| | D |
| | D |
| | D |
| | T |
| | T |
| Likely benign | T |

| | |
|---|---|
| Uncertain significance, early myoclonic encephalopathy | D |
| | D |
| | D |
| | D |
| | T |
| | T |
| | T |
| | D |
| | D |
| Conflicting interpretations, inborn genetic diseases | D |
| Benign | T |
| | D |
| | T |
| | D |
| | T |
| | D |
| Benign, Kabuki syndrome | D |
| | T |
| | T |
| | T |
| | T |
| | D |
| Benign, history of neurodevelopmental disorder | T |
| Benign | T |
| Benign, left ventricular noncompaction | T |
| | D |
| | D |
| | T |
| | T |
| | T |
| Benign, highly penetrant | D |
| Benign, early myoclonic encephalopathy | T |
| | D |
| | D |
| | T |
| | D |
| | T |
| | T |
| | T |
| Benign, Kabuki syndrome | T |
| Benign, highly penetrant | T |
| | D |
| | D |
| Benign, highly penetrant | D |
| Conflicting interpretations, highly penetrant, Kabuki syndrome | D |
| Likely benign, Kabuki syndrome | D |
| Likely benign | T |
| | T |

| | |
|---|---|
| Benign, left ventricular noncompaction | D |
| | T |
| Benign, left ventricular noncompaction | T |
| Uncertain significance, left ventricular noncompaction, Wolff-Parkinson-White pattern | T |
| | D |
| | T |
| | T |
| | D |
| Likely benign | T |
| | D |
| | D |
| | T |
| Benign | T |
| | T |
| Conflicting interpretations, Seckel syndrome 1 | D |
| Conflicting interpretations, hereditary breast and ovarian cancers | D |
| Uncertain significance, hereditary breast cancer, Fanconi anemia complementation group J | D |
| | D |
| | D |
| | D |
| Benign, Xeroderma pigmentosum group F | D |
| Benign, Falconi anemia | D |
| Likely benign, Falconi anemia | T |
| Conflicting interpretations, Falconi anemia | D |
| Benign, Falconi anemia | D |
| | T |
| | D |
| Uncertain significance, Lynch syndrome, colorectal cancer | D |
| Benign | T |
| Pathogenic, Lynch syndrome, Turcot syndrome, endometrial cancer, hereditary colorectal cancer | . |
| | T |
| Conflicting interpretations, MYH-associated polyposis, colon cancer | D |
| Uncertain significance, MYH-associated polyposis, hereditary cancer-predisposing | T |
| Conflicting interpretations, global developmental delay, mitochondrial DNA depletion syndrome, progressive sclerosing poliodystrophy, POLG-related disorders, seizures | D |
| | . |
| | D |
| | D |
| | T |
| | D |

| | |
|---|---|
| | D |
| | T |
| | D |
| | D |
| | T |
| | D |
| Likely benign | D |
| | D |
| | D |

| Polyphen2 prediction | References | Previously reported |
| --- | --- | --- |
| D | ;45, PMID: 28767289, PN | Found in pancreatic cancer cohort, and metastatic CRC (case report). In tumors of patients with chronic lymphocytic leukemia and 11q deletion. |
| D | 10.1093/hmg/ddm050, PI | Significantly associated with ovarian cancer risk. Enriched in breast cancer, may be associated with it (no access article) |
| D | ): 20104584, PMID: 1855 | Reported in breast and ovarian cancers. VUS. In conservative region. |
| D | | |
| P | PMID: 24465539 | Reported in breast cancer. Relation to cancer risk is not reported. |
| P | | |
| D | | |
| D | | |
| P | 351780, doi:10.1093/carc | Found in breast and ovarian cancer cohorts. Relation to cancer risk is not reported. |
| . | l0.1073/pnas.1407909111, | High frequency in breast cancer, especially in TNBC. Associated with poor 10-year breast cancer-specific survival. Immunohistochemical analyses show that variant carriers have reduced PAR-activity. |
| D | | |
| D | | |
| D | | |
| D | | |
| P | 515(02)01808-2, DOI: 1C | High risk in CRC or other HNPCC-associated cancers in Ashkenazi Jewish (founder mutation), endometrial cancer, Lynch syndrome. Increased frequency in CRC Jewish patients at age of 40 or younger. |
| D | | |
| P | | |
| D | ɔringer.com/article/10.10C | Reported in Australiam cohort of TNBC, and Italian cohort of breast cancer. Case report: found in proband with pancreatic, breast cancer, and melanoma |
| P | | |
| D | | |
| D | | |
| D | | |
| P | | |
| D | ɜr.biomedcentral.com/arti | Reported in family with familial breast cancer. No association with cancer risk, including breast, childhood leukemia, laryngeal, and head and neck cancers. |
| D | 10.1002/ijc.20169, https:/. | Highly represented in high-risk, site-specific, familial breast [illegible] |
| D | | |
| D | | |
| D | | |
| D | | |
| B | | |
| B | | |
| B | | |

| | | |
|---|---|---|
| D | | |
| B | | |
| P | | |
| P | | |
| B | | |
| B | | |
| P | | |
| D | | |
| D | | |
| B | | |
| P | | |
| D | | |
| B | | |
| P | | |
| B | | |
| B | | |
| B | | |
| B | | |
| B | | |
| B | | |
| B | | |
| D | | |
| B | | |
| D | | |
| D | | |
| D | | |
| P | | |
| B | | |
| B | | |
| B | | |
| P | ): 30608448, PMID: 2669 | rome. Case report: D185H persisted at transformation + EZH2 |
| P | | |
| D | | |
| B | | |
| P | | |
| D | | |
| D | | |
| B | | |
| D | | |
| D | | |
| B | | |
| D | | |
| D | | |
| D | | |
| D | PMID: 26032282 | Probably damaging in pheochromocytoma |
| D | | |
| B | | |
| B | | |

| | | |
|---|---|---|
| D | | |
| B | | |
| P | | |
| D | | |
| B | | |
| P | | |
| D | | |
| D | | |
| B | | |
| B | | |
| P | | |
| B | | |
| B | | |
| B | | |
| P | | |
| D | | |
| D | | |
| D | | |
| D | | |
| D | | |
| D | PMID: 17018596 | Found in breast cancer cohort. No association with breast cancer risk. |
| D | PMID: 12670332 | Presented at a fourfold greater frequency in children with acute myeloid leukaemia than in the cord blood samples. Also been reported in FA patients. |
| D | | |
| D | | |
| D | | |
| D | | |
| D | | |
| D | [illegible]50, PMID: 17656264, PM | Present in familial CRC in Korean cohort. Tumors with the variant are MSI-H. Functional studies in T293 cells showed no difference in MMR vs. wt. In conservative region. |
| D | | |
| . | doi.org/10.1053/j.gastro.2 | Identified in CRC cohort. No further studies. |
| D | | |
| D | | |
| D | | |
| D | | |
| P | | |
| D | | |
| D | | |
| P | | |
| D | PMID: 27782108 | No significant association with breast/ovarian cancer. More common in cancer cohorts, may be low-to-moderate risk, no validation. |

| | | |
|---|---|---|
| D | PMID: 26261251 | Common germline variant in (MAF>1%) in ovarian cancer cohort. Relation to cancer risk is not reported. |
| D | PMID: 26261251 | Common germline variant in (MAF>1%) in ovarian cancer cohort. Relation to cancer risk is not reported. |
| D | | |
| D | | |
| P | ): 25375110, PMID: 1522 | Significantly associated with CRC risk in Saudi cohort (AA genotype increased the risk by >3.6-fold, the minor allele A |
| D | | |
| D | | |
| D | | |
| D | | |

**Supplementary Table 7. Pathway-level enrichment summary in Complete Responders (CRs): total genes,**

| Pathways | base-excision repair | mismatch repair | interstrand cross-link repair | nucleotide-excision repair | demethylation |
|---|---|---|---|---|---|
| **total genes out of list of genes (n=161)** | 8 | 4 | 5 | 7 | 5 |
| **total variant genes in CRs (n=45)** | 6 | 1 | 3 | 4 | 9 |
| **p value from Fisher Exact Test** | 0.3726 | 0.0252 | 0.1298 | 0.0039 | 0.1069 |
| **FDR corrected p value (q value)** | 0.5446 | 0.1596 | 0.411 | 0.0494 | 0.37 |

**pathways with q value < 0.05 were considered to be enriched**

, variant-containing genes, Fisher’s exact test p-value, and FDR-adjusted q value

| non-recombinational repair | somatic diversification of immune receptors | double-strand break repair | DNA strand elongation | DNA damage response, detection of DNA damage |
|---|---|---|---|---|
| 9 | 10 | 10 | 12 | 17 |
| 6 | 2 | 11 | 1 | 0 |
| 0.9999 | 0.156 | 0.141 | 0.9999 | 0.1071 |
| 0.9999 | 0.4121 | 0.4121 | 0.9999 | 0.37 |

| post replication repair | telomere organization | DNA replication | DNA modification | cell cycle checkpoint | DNA conformation change | protein auto ubiquitination |
|---|---|---|---|---|---|---|
| 14 | 3 | 13 | 12 | 18 | 17 | 21 |
| 2 | 4 | 6 | 5 | 4 | 3 | 1 |
| 0.9999 | 0.438 | 0.0961 | 0.3559 | 0.2237 | 0.4017 | 0.9999 |
| 0.9999 | 0.5944 | 0.37 | 0.541 | 0.425 | 0.5654 | 0.9999 |

| cell cycle G2/M phase transition | chromosome segregation | B cell activation | mitotic cell cycle phase transition | regulation of chromosome organization |
|---|---|---|---|---|
| 25 | 8 | 12 | 20 | 22 |
| 2 | 5 | 2 | 3 | 13 |
| 0.5191 | 0.9999 | 0.5125 | 0.0563 | 0.0066 |
| 0.6363 | 0.9999 | 0.6363 | 0.3056 | 0.0627 |

| peptidyl-lysine modification | cell cycle arrest | cyclin-dependent protein kinase activity | adaptive immune response | response to oxidative stress | lymphocyte mediated immunity |
|---|---|---|---|---|---|
| 18 | 22 | 24 | 7 | 7 | 13 |
| 13 | 4 | 0 | 1 | 2 | 1 |
| 0.0001 | 0.2208 | 0.3231 | 0.2854 | 0.3529 | 0.2854 |
| 0.0019 | 0.425 | 0.5338 | 0.493 | 0.541 | 0.493 |

| methylation | cell cycle G1/S phase transition | epigenetic regulation of gene expression | intrinsic apoptotic signaling pathway | regulation of cell cycle phase transition | RNA capping |
|---|---|---|---|---|---|
| 40 | 36 | 13 | 29 | 20 | 25 |
| 16 | 2 | 6 | 3 | 3 | 0 |
| 0.0001 | 0.5125 | 0.1931 | 0.9999 | 0.1952 | 0.1919 |
| 0.0019 | 0.6363 | 0.4121 | 0.9999 | 0.4121 | 0.4121 |

| homologous recombination | heterochromatin organization | regulation of chromatin organization | protein monoubiquitination |
|---|---|---|---|
| 18 | 54 | 26 | 6 |
| 4 | 5 | 2 | 1 |
| 0.0962 | 0.0187 | 0.189 | 0.9999 |
| 0.37 | 0.1421 | 0.4121 | 0.9999 |

**Supplementary Table 8. Pathway-level enrichment summary in Poor Responders (PoRs): total genes, va**

| Pathways | base-excision repair | mismatch repair | interstrand cross-link repair | nucleotide-excision repair | demethylation |
|---|---|---|---|---|---|
| **total genes out of list of genes (n=161)** | 8 | 4 | 5 | 7 | 5 |
| **total variant genes in PoRs (n=48)** | 5 | 4 | 5 | 7 | 8 |
| **p value from Fisher Exact Test** | 0.1202 | 0.5891 | 0.6165 | 0.0716 | 0.3035 |
| **FDR corrected p value (q value)** | 0.3045 | 0.7221 | 0.7321 | 0.2267 | 0.5242 |

**pathways with q value < 0.05 were considered to be enriched**

**riant-containing genes, Fisher's exact test p-value, and FDR-adjusted q value**

| non-recombinational repair | somatic diversification of immune receptors | double-strand break repair | DNA strand elongation | DNA damage response, detection of DNA damage |
|---|---|---|---|---|
| 9 | 10 | 10 | 12 | 17 |
| 2 | 3 | 9 | 1 | 0 |
| 0.0236 | 0.4001 | 0.0106 | 0.9999 | 0.1065 |
| 0.1197 | 0.5848 | 0.1007 | 0.9999 | 0.3005 |

| post replication repair | telomere organization | DNA replication | DNA modification | cell cycle checkpoint | DNA conformation change | protein auto ubiquitination |
|---|---|---|---|---|---|---|
| 14 | 3 | 13 | 12 | 18 | 17 | 21 |
| 0 | 1 | 6 | 3 | 4 | 1 | 0 |
| 0.0586 | 0.0049 | 0.0627 | 0.7564 | 0.1519 | 0.0238 | 0.3185 |
| 0.2166 | 0.0969 | 0.2166 | 0.8212 | 0.3608 | 0.1197 | 0.5262 |

| cell cycle G2/M phase transition | chromosome segregation | B cell activation | mitotic cell cycle phase transition | regulation of chromosome organization |
|---|---|---|---|---|
| 25 | 8 | 12 | 20 | 22 |
| 2 | 4 | 2 | 2 | 5 |
| 0.3474 | 0.5891 | 0.5125 | 0.0085 | 0.342 |
| 0.528 | 0.7221 | 0.6716 | 0.1007 | 0.528 |

| peptidyl-lysine modification | cell cycle arrest | cyclin-dependent protein kinase activity | adaptive immune response | response to oxidative stress | lymphocyte mediated immunity |
|---|---|---|---|---|---|
| 18 | 22 | 24 | 7 | 7 | 13 |
| 7 | 1 | 1 | 1 | 2 | 1 |
| 0.4157 | 0.4375 | 0.9999 | 0.2837 | 0.2335 | 0.2837 |
| 0.5851 | 0.5938 | 0.9999 | 0.5134 | 0.467 | 0.5134 |

| methylation | cell cycle G1/S phase transition | epigenetic regulation of gene expression | intrinsic apoptotic signaling pathway | regulation of cell cycle phase transition | RNA capping |
|---|---|---|---|---|---|
| 40 | 36 | 13 | 29 | 20 | 25 |
| 14 | 0 | 6 | 1 | 2 | 1 |
| 0.0051 | 0.0188 | 0.2099 | 0.1107 | 0.0395 | 0.6752 |
| 0.0969 | 0.1197 | 0.4431 | 0.3005 | 0.1668 | 0.7546 |

| homologous recombination | heterochromatin organization | regulation of chromatin organization | protein monoubiquitination |
|---|---|---|---|
| 18 | 54 | 26 | 6 |
| 4 | 5 | 1 | 1 |
| 0.1979 | 0.0252 | 0.9999 | 0.6701 |
| 0.4424 | 0.1197 | 0.9999 | 0.7546 |

**Supplementary Table 9. Clinical and demographic characteristics of LARC co**

| Coded ID | Response | NAR Score | Gender | Race | Ethnicity | Clinical T Stage | Clinical N Stage |
|---|---|---|---|---|---|---|---|
| 1 | CR | 0.94 | Male | White | Non-Spanish, Non-His | c3 | c2A |
| 2 | CR | 0.94 | Female | White | Non-Spanish, Non-His | c3 | c0 |
| 3 | CR | 0.00 | Female | White | Non-Spanish, Non-His | c4 | cX |
| 4 | CR | 0.94 | Male | White | Non-Spanish, Non-His | c3 | c0 |
| 5 | CR | 0.94 | Male | White | Non-Spanish, Non-His | c3 | c0 |
| 6 | CR | 0.94 | Male | White | Non-Spanish, Non-His | c3 | c0 |
| 7 | CR | 0.94 | Female | White | Non-Spanish, Non-His | c3 | c0 |
| 8 | CR | 0.94 | Male | White | Non-Spanish, Non-His | c3 | c0 |
| 9 | CR | 0.94 | Male | White | Non-Spanish, Non-His | c3 | c1A |
| 10 | CR | 0.94 | Female | White | Non-Spanish, Non-His | c3 | c0 |
| 11 | CR | 0.94 | Male | White | Non-Spanish, Non-His | c3 | c0 |
| 12 | CR | 0.9365245 | Female | White | Non-Spanish, Non-His | c3 | c1 |
| 13 | CR | 0.9365245 | Male | White | Non-Spanish, Non-His | c3 | c1 |
| 14 | CR | 0.9365245 | Male | White | Non-Spanish, Non-His | c3 | c0 |
| 15 | CR | 0.9365245 | Female | White | Non-Spanish, Non-His | c3 | cX |
| 16 | CR | 0.9365245 | Male | White | Non-Spanish, Non-His | c3 | c1 |
| 17 | CR | 0.9365245 | Female | White | Non-Spanish, Non-His | c3 | c1 |
| 18 | CR | 0.9365245 | Female | White | Non-Spanish, Non-His | c3 | c0 |
| 19 | CR | 0.9365245 | Female | White | Non-Spanish, Non-His | c3 | c0 |
| 20 | CR | 0.9365245 | Male | White | Non-Spanish, Non-His | c3 | c1B |
| 21 | CR | 0.94 | Male | White | Non-Spanish, Non-His | c3 | c1B |
| 22 | CR | 0.94 | Male | White | Non-Spanish, Non-His | c3 | c1B |
| 23 | CR | 0.94 | Male | White | Non-Spanish, Non-His | c3 | c1 |

| | | | | | | | |
|---|---|---|---|---|---|---|---|
| 24 | CR | 0.9365245 | Female | White | Non-Spanish, Non-His | c3 | c1 |
| 25 | CR | 0 | Female | White | Non-Spanish, Non-His | c4B | c1 |
| 26 | CR | 0.9365245 | Female | White | Non-Spanish, Non-His | c3 | c0 |
| 27 | CR | 0.9365245 | Male | White | Non-Spanish, Non-His | c3 | c1 |
| 28 | CR | 0.94 | Male | White | Non-Spanish, Non-His | c3 | c0 |
| 29 | CR | 0.9365245 | Female | White | Non-Spanish, Non-His | c4 | c1 |
| 30 | CR | 0.9365245 | Female | Black | Non-Spanish, Non-His | c3 | c1 |
| 31 | CR | 0.9365245 | Female | White | Non-Spanish, Non-His | c3 | c0 |
| 32 | CR | 0.94 | Male | White | Non-Spanish, Non-His | c3 | c0 |
| 33 | CR | 0.94 | Female | White | Non-Spanish, Non-His | c3 | c0 |
| 34 | CR | 0.94 | Male | White | Non-Spanish, Non-His | c3 | c0 |
| 35 | CR | 0.94 | Male | White | Non-Spanish, Non-His | c3 | c0 |
| 36 | CR | 0.94 | Female | White | Non-Spanish, Non-His | c3 | c0 |
| 37 | CR | 0.94 | Male | White | Non-Spanish, Non-His | c3 | c0 |
| 38 | PoR | 14.98 | Male | White | Non-Spanish, Non-His | c3 | c0 |
| 39 | PoR | 14.98 | Male | White | Non-Spanish, Non-His | c3 | c0 |
| 40 | PoR | 14.98 | Female | White | Non-Spanish, Non-His | c3 | c0 |
| 41 | PoR | 50.36 | Female | White | Non-Spanish, Non-His | c3 | c0 |
| 42 | PoR | 30.07 | Male | White | Non-Spanish, Non-His | c3 | c0 |
| 43 | PoR | 41.62 | Female | White | Non-Spanish, Non-His | c3 | c0 |
| 44 | PoR | 14.98 | Female | White | Non-Spanish, Non-His | c3 | c0 |
| 45 | PoR | 20.40 | Male | White | Non-Spanish, Non-His | c3 | c0 |
| 46 | PoR | 14.98 | Female | White | Non-Spanish, Non-His | c3 | c0 |
| 47 | PoR | 30.07 | Male | White | Non-Spanish, Non-His | c3 | c1 |
| 48 | PoR | 14.984391 | Male | White | Non-Spanish, Non-His | c3 | c1 |
| 49 | PoR | 20.395421 | Male | White | Non-Spanish, Non-His | c3 | c1 |

| | | | | | | | |
|---|---|---|---|---|---|---|---|
| 50 | PoR | 14.984391 | Male | White | Non-Spanish, Non-His | c3 | c1 |
| 51 | PoR | 14.984391 | Female | White | Non-Spanish, Non-His | c3 | c1 |
| 52 | PoR | 30.072841 | Male | Black | Non-Spanish, Non-His | c3 | c1 |
| 53 | PoR | 30.072841 | Male | White | Non-Spanish, Non-His | c3 | c1 |
| 54 | PoR | 14.984391 | Female | White | Non-Spanish, Non-His | c3 | c0 |
| 55 | PoR | 14.984391 | Male | White | Non-Spanish, Non-His | c3 | c1 |
| 56 | PoR | 50.364204 | Male | White | Non-Spanish, Non-His | c3 | c2 |
| 57 | PoR | 30.072841 | Male | White | Non-Spanish, Non-His | c3 | c1 |
| 58 | PoR | 20.395421 | Male | White | Non-Spanish, Non-His | c3 | cX |
| 59 | PoR | 14.984391 | Male | Black | Non-Spanish, Non-His | c3 | c1 |
| 60 | PoR | 50.364204 | Female | White | Non-Spanish, Non-His | c3 | c1 |
| 61 | PoR | 50.364204 | Male | White | Non-Spanish, Non-His | c3 | c1 |
| 62 | PoR | 30.072841 | Female | White | Non-Spanish, Non-His | c4 | c2 |
| 63 | PoR | 30.07 | Male | White | Non-Spanish, Non-His | c3 | c0 |
| 64 | PoR | 14.984391 | Male | White | Non-Spanish, Non-His | c3 | c1 |
| 65 | PoR | 30.072841 | Male | White | Non-Spanish, Non-His | c3 | c1 |
| 66 | PoR | 14.984391 | Male | White | Non-Spanish, Non-His | c3 | c1 |
| 67 | PoR | 50.364204 | Male | Black | Non-Spanish, Non-His | c3 | c1 |
| 68 | PoR | 14.984391 | Female | White | Non-Spanish, Non-His | c3 | c1 |
| 69 | PoR | 30.072841 | Female | Black | Non-Spanish, Non-His | c3 | c1 |
| 70 | PoR | 30.07 | Male | White | Non-Spanish, Non-His | c3 | c1 |
| 71 | PoR | 14.984391 | Female | White | Non-Spanish, Non-His | c2 | c1 |
| 72 | PoR | 14.984391 | Male | White | Non-Spanish, Non-His | c3 | c0 |
| 73 | PoR | 30.072841 | Male | White | Non-Spanish, Non-His | c3 | c1 |
| 74 | PoR | 30.072841 | Male | White | Non-Spanish, Non-His | c4B | c0 |

| | | | | | | | |
|---|---|---|---|---|---|---|---|
| 75 | PoR | 20.395421 | Male | White | Non-Spanish, Non-His | c4 | c2 |
| 76 | PoR | 50.364204 | Male | White | Non-Spanish, Non-His | c3 | c1 |
| 77 | PoR | 30.07 | Male | White | Non-Spanish, Non-His | c3 | c0 |
| 78 | PoR | 14.98 | Male | White | Non-Spanish, Non-His | c3 | c0 |
| 79 | PoR | 14.98 | Male | White | Non-Spanish, Non-His | c3 | c0 |
| 80 | PoR | 41.62 | Male | White | Non-Spanish, Non-His | c3 | c0 |
| 81 | PoR | 26.64 | Female | White | Non-Spanish, Non-His | c3 | c1B |
| 82 | PoR | 14.98 | Male | Black | Non-Spanish, Non-His | c3 | c1 |
| 83 | PoR | 14.98 | Male | White | Non-Spanish, Non-His | c3 | c0 |
| 84 | PoR | 14.98 | Male | White | Non-Spanish, Non-His | c2 | c0 |

| Clinical M Stage | Pathologic T Stage | Pathologic N Stage | Pathologic Stage Group | Treatment Name I | Treatment Name II |
|---|---|---|---|---|---|
| c0 | p2 | p0 | 1 | Chemotherapy | Radiation |
| c0 | p0 | p0 | 99 | Chemotherapy | Radiation |
| c0 | pX | pX | 99 | Chemotherapy | Radiation |
| c0 | p0 | p0 | N/A | Chemotherapy | Radiation |
| c0 | p0 | p0 | 99 | Chemotherapy | Radiation |
| c0 | p0 | p0 | | Chemotherapy | Radiation |
| c0 | p0 | p0 | 99 | Chemotherapy | Radiation |
| c0 | p0 | p0 | 99 | Radiation | Surgery |
| c0 | p0 | p0 | 99 | Chemotherapy | Radiation |
| c0 | p0 | p0 | 99 | | Radiation |
| c0 | p0 | p0 | | | Radiation |
| c0 | p0 | p0 | 99 | Radiation | Chemotherapy |
| c0 | p0 | p0 | 99 | Chemotherapy | Radiation |
| c0 | pIS | p0 | 0 | Chemotherapy | Radiation |
| c0 | p0 | p0 | 99 | Chemotherapy | Radiation |
| c0 | p0 | p0 | 99 | Chemotherapy | Radiation |
| c0 | p0 | p0 | 99 | Chemotherapy | Radiation |
| c0 | pIS | p0 | 0 | Chemotherapy | Radiation |
| c0 | pIS | p0 | 0 | Radiation | Chemotherapy |
| c0 | p0 | p0 | 99 | Chemotherapy | Radiation |
| c0 | p0 | p0 | 99 | Chemotherapy | Radiation |
| c0 | p0 | p0 | 99 | Chemotherapy | Radiation |
| c0 | p0 | p0 | 99 | Chemotherapy | Radiation |

| | | | | | |
|---|---|---|---|---|---|
| c0 | p0 | p0 | 99 | Radiation | Chemotherapy |
| c0 | p0 | p0 | 99 | Chemotherapy | Radiation |
| c0 | p0 | p0 | | Chemotherapy | Radiation |
| c0 | pIS | p0 | 0 | Chemotherapy | Chemotherapy |
| c0 | pX | p0 | 99 | Chemotherapy | Radiation |
| c0 | p1 | p0 | 1 | Chemotherapy | Radiation |
| c0 | p0 | p0 | | Chemotherapy crt | Radiation |
| c1A | p0 | p0 | 4A | | |
| c0 | p0 | p0 | 99 | Radiation | Surgery |
| c0 | c0 | p0 | 99 | Surgery | Radiation |
| c0 | c0 | p0 | 99 | Radiation | Surgery |
| c0 | p0 | p0 | 99 | | Surgery |
| c0 | c0 | p0 | 99 | Radiation | Surgery |
| c0 | p0 | p0 | 99 | Radiation | Surgery |
| c0 | p3 | p0 | 2A | | Radiation |
| c0 | p3 | p0 | 2A | | Radiation |
| c0 | p3 | p0 | 2A | | Radiation |
| c0 | p3 | p2A | 3B | Chemotherapy | Radiation |
| c1A | p3 | p1C | 4A | Chemotherapy | Radiation |
| c0 | p4A | p1C | 3B | | Radiation |
| c0 | p3 | p0 | 2A | Chemotherapy | Radiation |
| c0 | p2 | p1 | 3A | | Radiation |
| c0 | p3 | p0 | 2A | | Radiation |
| c0 | p3 | p1 | 3B | Chemotherapy | Radiation |
| c0 | p3 | p0 | 2 | Chemotherapy | Radiation |
| c0 | p2 | p1 | 4 | Chemotherapy | Chemotherapy |

| | | | | | |
|---|---|---|---|---|---|
| c0 | p3 | p0 | 2A | Chemotherapy | Radiation |
| c0 | p3 | p0 | 2A | Chemotherapy | Radiation |
| c1 | p3 | p1 | 4 | Chemotherapy | Radiation |
| c0 | p3 | p1 | 3B | Chemotherapy | Radiation |
| c0 | p3 | p0 | 2A | Chemotherapy | Radiation |
| c0 | p3 | p0 | 2A | Chemotherapy | Radiation |
| c1 | p3 | p2 | 4 | Chemotherapy | Chemotherapy |
| c0 | p3 | p1 | 3B | Chemotherapy | Chemotherapy |
| c0 | p2 | p1 | 3A | Chemotherapy | Radiation |
| c0 | | | 99 | Chemotherapy | Radiation |
| c1 | p3 | p2 | 3B | Chemotherapy | Radiation |
| c0 | p3 | p2 | 3C | Chemotherapy | Radiation |
| c1 | p4 | p1 | 4 | Chemotherapy | Chemotherapy |
| c0 | p3 | p1A | 3B | | Radiation |
| c0 | p3 | p0 | 2A | Chemotherapy | Radiation |
| c0 | p3 | p1 | 3 | Chemotherapy | Radiation |
| c0 | p3 | p0 | 2A | Radiation | Chemotherapy |
| c0 | p3 | p2 | 3C | Chemotherapy | Chemotherapy |
| c0 | p3 | p0 | 2A | Chemotherapy | Radiation |
| c0 | p3 | p1 | 3B | Chemotherapy | Chemotherapy |
| c0 | p3 | p1 | 3 | Chemotherapy | Radiation |
| c0 | p2 | p0 | 1 | Chemotherapy | Radiation |
| c0 | p3 | p0 | 2 | Chemotherapy | Radiation |
| c0 | p3 | p1 | 3B | Radiation | Chemotherapy |
| c0 | p4B | p1 | 3C | Chemotherapy | Chemotherapy |

| | | | | | |
|---|---|---|---|---|---|
| c0 | p3 | p1 | 3B | Chemotherapy | Radiation |
| c0 | p3 | p2 | 3C | | |
| c0 | p3 | p1 | 3B | Radiation | Surgery |
| c0 | p3 | p0 | 2A | Radiation | Surgery |
| c0 | p3 | p0 | 2A | | Radiation |
| c0 | p4B | p1A | 3C | Radiation | Surgery |
| c1A | p1 | p2A | 4A | | |
| c0 | p3 | p0 | 2A | Radiation | Surgery |
| c0 | p3 | p0 | 2A | Radiation | Surgery |
| c0 | p2 | p0 | 1 | Radiation | Surgery |

| Treatment Name III | Treatment Name IV | Treatment Name V |
|---|---|---|
| Surgery | N/a | N/a |
| Surgery | N/a | N/a |
| Surgery | N/a | N/a |
| Surgery | N/a | N/a |
| Surgery | N/a | N/a |
| Surgery | N/a | N/a |
| Surgery | N/a | N/a |
| Chemotherapy | N/a | N/a |
| Surgery | N/a | N/a |
| Surgery | N/a | N/a |
| Surgery | N/a | N/a |
| Surgery | N/a | N/a |
| Surgery | N/a | N/a |
| Surgery | Chemotherapy | N/a |
| Surgery | N/a | N/a |
| Surgery | N/a | N/a |
| Surgery | N/a | N/a |
| Surgery | N/a | N/a |
| Surgery | Chemotherapy | N/a |
| Surgery | Chemotherapy | N/a |
| Surgery | N/a | N/a |
| Surgery | N/a | N/a |
| Surgery | N/a | N/a |

| | | |
|---|---|---|
| Surgery | N/a | N/a |
| Surgery | N/a | N/a |
| Surgery | Chemotherapy | N/a |
| Radiation | Surgery | Chemotherapy |
| Surgery | N/a | N/a |
| Surgery | N/a | N/a |
| Surgery | N/a | N/a |

| | | | |
|---|---|---|---|
| Surgery | N/a | N/a | N/a |
| Surgery | N/a | N/a | N/a |
| Surgery | N/a | N/a | N/a |
| Surgery | N/a | N/a | N/a |
| Surgery | N/a | N/a | N/a |
| Surgery | N/a | N/a | N/a |
| Surgery | N/a | N/a | N/a |
| Surgery | N/a | N/a | N/a |
| Surgery | N/a | N/a | N/a |
| Surgery | N/a | N/a | N/a |
| Surgery | Chemotherapy | N/a | N/a |
| Radiation | Surgery | Chemotherapy | hemotherapy |

| | | | |
|---|---|---|---|
| Surgery | N/a | N/a | N/a |
| Surgery | N/a | N/a | N/a |
| Surgery | N/a | N/a | N/a |
| Surgery | N/a | N/a | N/a |
| Surgery | N/a | N/a | N/a |
| Surgery | N/a | N/a | N/a |
| Radiation | Surgery | Chemotherapy | hemotherapy |
| Radiation | Surgery | Chemotherapy | hemotherapy |
| Surgery | N/a | N/a | N/a |
| Surgery | N/a | N/a | N/a |
| Surgery | N/a | N/a | N/a |
| Surgery | N/a | N/a | N/a |
| Radiation | Surgery | Chemotherapy | N/a |
| Surgery | N/a | N/a | N/a |
| Surgery | N/a | N/a | N/a |
| Surgery | Chemotherapy | N/a | N/a |
| Surgery | N/a | N/a | N/a |
| Radiation | Surgery | Chemotherapy | N/a |
| Surgery | N/a | N/a | N/a |
| Radiation | Surgery | Chemotherapy | N/a |
| Surgery | N/a | N/a | N/a |
| Surgery | N/a | N/a | N/a |
| Surgery | Chemotherapy | Chemotherapy | N/a |
| Surgery | N/a | N/a | N/a |
| Radiation | Surgery | Chemotherapy | N/a |

| | | | |
|---|---|---|---|
| Surgery | Chemotherapy | N/a | N/a |

Surgery

**Supplementary Table 10. Clinical and demographic characteristics of HNSCC cohort (n=90)**

| No Recurrence (coded ID) | Gender | Race | Tested for HPV |
|---|---|---|---|
| 1 | Male | White | Yes |
| 2 | Male | White | Yes |
| 3 | Male | White | No |
| 4 | Female | White | No |
| 5 | Female | White | No |
| 6 | Female | White | No |
| 7 | Male | White | No |
| 8 | Male | White | Yes |
| 9 | Female | White | Yes |
| 10 | Male | White | Yes |
| 11 | Male | White | Yes |
| 12 | Male | White | Yes |
| 13 | Male | White | Yes |
| 14 | Female | White | Yes |
| 15 | Male | White | Yes |
| 16 | Male | White | Yes |
| 17 | Male | Other Asian | Yes |
| 18 | Male | White | Yes |
| 19 | Male | White | No |
| 20 | Male | White | No |
| 21 | Male | White | No |
| 22 | Male | White | Yes |
| 23 | Male | White | No |
| 24 | Female | White | No |
| 25 | Female | White | No |
| 26 | Female | White | No |
| 27 | Female | White | No |
| 28 | Male | White | Yes |
| 29 | Female | White | No |
| 30 | Male | Black | No |

| Recurrence (coded ID) | Gender | Race | Tested for HPV |
|---|---|---|---|
| 1 | Male | White | No |
| 2 | Male | Black | No |
| 3 | Male | White | No |
| 4 | Male | White | Yes |

| | | | |
|---|---|---|---|
| 5 | Female | White | No |
| 6 | Male | White | No |
| 7 | Male | White | Yes |
| 8 | Female | White | No |
| 9 | Male | White | Yes |
| 10 | Male | White | No |
| 11 | Male | White | Yes |
| 12 | Female | White | Yes |
| 13 | Male | White | Yes |
| 14 | Male | White | Yes |
| 15 | Male | White | Yes |
| 16 | Male | White | No |
| 17 | Female | White | No |
| 18 | Male | White | Yes |
| 19 | Male | White | No |
| 20 | Male | White | No |
| 21 | Male | White | No |
| 22 | Male | Black | Yes |
| 23 | Male | White | No |
| 24 | Male | White | No |
| 25 | Male | · Pakistani, NOS (for | No |
| 26 | Female | White | No |
| 27 | Male | White | Yes |
| 28 | Female | White | No |

| **Incomplete Response (coded ID)** | **Gender** | **Race** | **Tested for HPV** |
|---|---|---|---|
| 1 | Male | White | Yes |
| 2 | Male | White | Yes |
| 3 | Male | White | No |
| 4 | Male | White | Yes |
| 5 | Male | White | Yes |
| 6 | Male | White | No |
| 7 | Female | White | No |
| 8 | Male | White | No |
| 9 | Male | White | Yes |
| 10 | Male | White | No |
| 11 | Male | White | Yes |
| 12 | Female | White | No |

| | | | |
|---|---|---|---|
| 13 | Male | White | No |
| 14 | Male | White | No |
| 15 | Male | Black | Yes |
| 16 | Male | White | Yes |
| 17 | Male | White | Yes |
| 18 | Male | White | Yes |
| 19 | Female | White | Yes |
| 20 | Male | White | Yes |
| 21 | Male | White | No |
| 22 | Male | White | Yes |
| 23 | Female | White | Yes |
| 24 | Male | White | Yes |
| 25 | Female | White | Yes |
| 26 | Male | White | Yes |
| 27 | Male | White | No |
| 28 | Male | White | No |
| 29 | Male | White | No |
| 30 | Male | Other | Yes |
| 31 | Male | White | Yes |
| 32 | Male | White | No |

| p16 expression | HPV status/ hrHPV+ | Clinical T Stage | Clinical N Stage | Clinical M Stage |
|---|---|---|---|---|
| Negative | No | c1A | c0 | c0 |
| Positive | No | c3 | c0 | c0 |
| n/a | n/a | c4A | c0 | c0 |
| n/a | n/a | c2 | c0 | c0 |
| n/a | n/a | c3 | c0 | c0 |
| n/a | n/a | c3 | c0 | c0 |
| n/a | n/a | c3 | c2C | c0 |
| Negative | No | c4A | c2A | c0 |
| Positive | Yes | c1 | c1 | c0 |
| Positive | Yes | c4A | c2C | c0 |
| Positive | No | c3 | c2C | c0 |
| Positive | Yes | c1 | c1 | c0 |
| Positive | Yes | c2 | c1 | c0 |
| Negative | No | c4A | c0 | c0 |
| Negative | No | c2 | c1 | c0 |
| Positive | Yes | c2 | c0 | c0 |
| Negative | No | c4A | c0 | c0 |
| Negative | No | c4A | c0 | c0 |
| n/a | n/a | c4A | c2B | c0 |
| n/a | n/a | c4a | c2b | c0 |
| n/a | n/a | c4a | c2b | c0 |
| Positive | Yes | c1 | c2B | c0 |
| n/a | n/a | c4A | c1 | c0 |
| n/a | n/a | c4A | c0 | c0 |
| n/a | n/a | c4A | c0 | c0 |
| n/a | n/a | c2 | c1 | c0 |
| n/a | n/a |  | c0 | c0 |
| Negative | No | c1A | c0 | c0 |
| n/a | n/a | c3 | c0 | c0 |
| n/a | n/a | c2 | c1 | c0 |

| p16 expression | HPV status/ hrHPV+ | Clinical T Stage | Clinical N Stage | Clinical M Stage |
|---|---|---|---|---|
| n/a | n/a | c1A | c0 | c0 |
| n/a | n/a | c1 | c0 | c0 |
| n/a | n/a | c1B | c0 | c0 |
| Positive | Yes | c3 | c2C | c0 |

| | | | | |
|---|---|---|---|---|
| n/a | n/a | c4A | c0 | c0 |
| n/a | n/a | c3 | c0 | c0 |
| Negative | No | c4A | c1 | c0 |
| n/a | n/a | c4A | c0 | c0 |
| Negative | No | c1 | c1 | c0 |
| n/a | n/a | c4A | c0 | c0 |
| Negative | No | c3 | c1 | c0 |
| Negative | No | c2 | c0 | c0 |
| Positive | Yes | c3 | c0 | c0 |
| Negative | No | c2 | c0 | c0 |
| Negative | No | c4B | c2B | c0 |
| n/a | n/a | c2 | c0 | c0 |
| n/a | n/a | | c0 | c0 |
| Negative | No | c2 | c1 | c0 |
| n/a | n/a | c1A | c0 | c0 |
| n/a | n/a | c1 | c0 | c0 |
| n/a | n/a | c1 | c0 | c0 |
| Negative | No | c2 | c0 | c0 |
| n/a | n/a | c1A | c0 | c0 |
| n/a | n/a | c3 | c0 | c0 |
| n/a | n/a | c2 | c0 | c0 |
| n/a | n/a | c4A | c2 | c0 |
| Negative | No | c1 | c0 | c0 |
| n/a | n/a | c3 | c2B | c0 |

| **p16 expression** | **HPV status/ hrHPV+** | **Clinical T Stage** | **Clinical N Stage** | **Clinical M Stage** |
|---|---|---|---|---|
| Positive | Yes | c2 | c0 | c0 |
| Unknown | n/a | 88 | 88 | 88 |
| n/a | n/a | *c1B* | c0 | c0 |
| Negative | No | c3 | c2C | c1 |
| Positive | Yes | c1 | c2B | c0 |
| n/a | n/a | c4A | c2B | c0 |
| n/a | n/a | c1 | c0 | c0 |
| n/a | n/a | c4A | c1 | c0 |
| Negative | No | c2 | c1 | c0 |
| n/a | n/a | c4A | c2B | c0 |
| Negative | No | c2 | c2B | c0 |
| n/a | n/a | c2 | c1 | c0 |

| | | | | |
|---|---|---|---|---|
| n/a | n/a | c1 | c2B | c0 |
| n/a | n/a | c2 | c1 | c0 |
| Negative | No | c3 | c0 | c0 |
| Positive | Yes | c4 | c2 | c0 |
| Negative | No | 88 | 88 | 88 |
| Positive | Yes | c4 | c3 | c0 |
| Negative | No | c2 | c3b | c0 |
| Positive | Yes | c1 | c1 | c0 |
| n/a | n/a | c3 | c1 | c0 |
| Negative | No | c4A | c2C | c0 |
| Positive | Yes | c2 | c2C | c0 |
| Positive | No | c2 | c2B | c0 |
| Negative | No | c4b | c2c | c0 |
| Positive | Yes | c2 | c0 | c0 |
| n/a | n/a | c3 | c2C | c0 |
| n/a | n/a | c3 | c0 | c0 |
| n/a | n/a | c3 | c0 | c0 |
| Negative | No | c3 | c0 | c0 |
| Positive | Yes | c1 | c2B | c0 |
| n/a | n/a | c4A | c0 | c0 |

| Treatment Name I | Treatment Name II |
|---|---|
| Radiation | |
| Radiation | Chemotherapy |
| Chemotherapy | Radiation |
| Chemotherapy | Radiation |
| Chemotherapy | Radiation |
| Chemotherapy | Radiation |
| Chemotherapy | Radiation |
| Chemotherapy | Radiation |
| Radiation | |
| Radiation | Chemotherapy |
| Chemotherapy | |
| Chemotherapy | Radiation |
| Chemotherapy | Radiation |
| Radiation | |
| Surgery | Radiation |
| Surgery | Radiation |
| Radiation | |
| Surgery | Chemotherapy |
| Surgery | Radiation |
| Surgery | Radiation |
| Surgery | Chemotherapy |
| Chemotherapy | Radiation |
| Surgery | Radiation |
| Surgery | Radiation |
| Surgery | Radiation |
| Chemotherapy | Radiation |
| Surgery | Chemotherapy |
| Radiation | |
| Surgery | Chemotherapy |
| Radiation | |

| Treatment Name I | Treatment Name II |
|---|---|
| Radiation | Surgery |
| Radiation | |
| Surgery | |
| Chemotherapy | Radiation |

| | |
|---|---|
| Surgery | Radiation |
| Surgery | |
| Surgery | Radiation |
| Surgery | Radiation |
| Surgery | Radiation |
| Surgery | Radiation |
| Surgery | Radiation |
| Radiation | |
| Radiation | Chemotherapy |
| Surgery | Radiation |
| Chemotherapy | Radiation |
| Surgery | Radiation |
| Surgery | |
| Surgery | Radiation |
| Radiation | Surgery |
| Radiation | Surgery |
| Chemotherapy | Radiation |
| Radiation | |
| Radiation | |
| Radiation | |
| Chemotherapy | Radiation |
| Surgery | Radiation |
| Radiation | |
| Chemotherapy | Radiation |

| Treatment Name I | Treatment Name II |
|---|---|
| Radiation | Chemotherapy |
| Chemotherapy | Radiation |
| Radiation | |
| Immunotherapy | Radiation |
| Surgery | Radiation |
| Surgery | Chemotherapy |
| Surgery | Radiation |
| Surgery | Radiation |
| Surgery | Radiation |
| Surgery | Radiation |
| Surgery | Radiation |
| Surgery | Chemotherapy |

| | |
|---|---|
| Chemotherapy | Radiation |
| Surgery | Radiation |
| Surgery | Chemotherapy |
| Chemotherapy | Radiation |
| Chemotherapy | |
| Chemotherapy | Radiation |
| Chemotherapy | Radiation |
| Immunotherapy | Radiation |
| Chemotherapy | Radiation |
| Chemotherapy | Radiation |
| Chemotherapy | Radiation |
| Surgery | Radiation |
| Chemotherapy | Radiation |
| Surgery | Radiation |
| Chemotherapy | Radiation |
| Surgery | Chemotherapy |
| Radiation | |
| Chemotherapy | Radiation |
| Chemotherapy | |
| Surgery | Radiation |

**Supplementary Table 11. Allele frequency of SNPs in LARC patients (n=30). GnomAD version us**

| Gene | Position | Ref | Alt | AA Change | Total |
|---|---|---|---|---|---|
| **Complete responders** | | | | | |
| *ATM* | rs150757822 | A | C | K1992T | 0.0002163 |
| *BRCA1* | rs1799950 | T | C | Q356R | 0.05506 |
| *BRCA2* | rs28897745 | A | G | D2665G | 0.0004034 |
| *DOT1L* | rs144165419 | G | A | A1053T | 0.001159 |
| *DOT1L* | rs113164803 | C | A | L1379M | 0.01444 |
| *ERCC4* | rs150244523 | T | C | L313P | 0.00003471 |
| *ERCC4* | rs1800124 | A | G | E875G | 0.01531 |
| *FAAP24* | rs36017455 | C | T | S126F | 0.005937 |
| *FAAP24* | rs148106526 | C | T | T212M | 0.002579 |
| *FAN1* | rs771137282 | G | A | V687I | 0.0001221 |
| *FANCM* | rs61746943 | C | T | T1600I | 0.01919 |
| *FANCM* | rs147021911 | C | T | Q1701X | 0.0009548 |
| *KDM3C/JMJD1C* | rs200797706 | A | G | L1458S | 0.00002417 |
| *KDM3C/JMJD1C* | rs200160728 | G | A | P1089S | 0.0005019 |
| *KDM3C/JMJD1C* | rs10761725 | A | T | S464T | 0.7366 |
| *KDM3A* | rs143405154 | G | T | C417F | 0.001672 |
| *KDM3B* | rs200772506 | G | A | A5T | 0.03038 |
| *KDM4A* | rs138721228 | C | T | A493V | 0.0001395 |
| *KDM4C* | rs34369202 | T | C | I603T | 0.001246 |
| *KDM4D* | rs144086807 | C | G | P457A | 0.004423 |
| *KDM5A* | rs781474100 | C | T | R1282H | 0.00003222 |
| *KDM5B* | rs112284833 | C | G | E939Q | 0.02867 |
| *KDM6B* | rs148641957 | G | T | V209L | 0.004077 |
| *KMT2A* | rs9332745 | C | G | A30G | 0.02374 |
| *KMT2A* | rs778675942 | C | T | R3228C | 4.337E-06 |
| *KMT2A* | rs199656572 | C | G | P3531A | 0.000127 |
| *KMT2B* | . | G | A | E117K | NA |
| *KMT2C* | rs142997680 | T | A | T3575S | 0.00005452 |
| *KMT2C* | rs755062207 | C | T | G2986D | 0.00001053 |
| *KMT2D* | rs75937132 | T | C | M3398V | 0.0105 |
| *KMT2E* | rs200289007 | A | T | M316L | 0.0001078 |
| *KMT2E* | rs145923995 | A | G | N1195S | 0.00148 |
| *KMT2E* | rs74959149 | G | A | S1532N | 0.01427 |
| *KMT5C* | rs200852152 | C | A | P328H | 0.0008091 |
| *MBD4* | rs2307293 | C | G | D568H | 0.007184 |
| *MMS19* | rs36023427 | C | T | G1029D | 0.005661 |
| *MMS19* | rs29001280 | G | A | R98W | 0.0005574 |
| *MPG* | rs145824088 | G | A | V179I | 0.000587 |
| *MSH2* | rs63750875 | G | C | A636P | 7.435E-06 |
| *MUS81* | rs34891773 | C | T | R351W | 0.02616 |
| *NEIL3* | rs34007209 | C | T | R38C | 0.0312 |
| *NSD1* | rs768074196 | C | T | R2195C | 8.054E-06 |

| | | | | | |
|---|---|---|---|---|---|
| *NSD1* | rs3733875 | G | T | V345L | 0.1377 |
| *NSD3* | rs185811622 | G | A | S754L | 0.008308 |
| *PALB2* | rs45624036 | C | T | V932M | 0.004741 |
| *PARP2* | rs3093921 | A | G | D235G | 0.01998 |
| *PER1* | rs72845601 | G | A | P37S | 0.01756 |
| *POLI* | rs3218786 | T | C | F532S | 0.02802 |
| *POLQ* | rs3218634 | G | C | L2538V | 0.06677 |
| *POLQ* | rs532411 | G | A | A2304V | 0.067 |
| *PRDM16* | rs201904226 | G | A | G818S | 0.002066 |
| *RAD50* | rs28903086 | G | A | V127I | 0.001356 |
| *RAD51D* | rs28363284 | T | C | E233G | 0.01682 |
| *SETD1A* | . | C | T | R694W | N/A |
| *SETD1A* | rs374966225 | C | T | T1122M | 0.00008439 |
| *SETD1B* | rs58801491 | T | C | L1355P | 0.01211 |
| *SHPRH* | . | C | T | R1479H | N/A |
| *SMYD1* | rs565724752 | G | A | V258I | 0.000153 |
| *TP53BP1* | rs45482998 | A | G | V1031A | 0.0126 |
| *TP53BP1* | rs148009445 | T | C | Q308R | 0.0001184 |
| *XRCC1* | rs2307186 | C | A | R7L | 0.00218 |
| **Poor responders** | | | | | |
| *ASH1L* | rs78904392 | T | C | N1941D | 0.001113 |
| *ATR* | rs77208665 | T | C | K764E | 0.004589 |
| *BRCA2* | rs80358431 | C | G | S476C | 0.00004597 |
| *BRIP1* | rs587781325 | A | G | Y369H | 0.00002974 |
| *CDK7* | rs34584424 | C | T | T285M | 0.02056 |
| *CHAF1A* | rs369272426 | C | T | T904M | 0.0003111 |
| *EME1* | rs148490439 | C | A | P245Q | 0.0002067 |
| *ERCC4* | rs1800067 | G | A | R415Q | 0.06796 |
| *EZH2* | rs2302427 | C | G | D185H | 0.07367 |
| *FANCC* | rs1800361 | G | A | S26F | 0.005791 |
| *FANCI* | rs149223439 | G | C | G53A | 0.0002374 |
| *FANCI* | rs151038616 | T | A | L619Q | 0.00009356 |
| *FANCM* | rs45547534 | A | G | I208M | 0.01418 |
| *HELQ* | rs113520876 | T | A | Y876F | 0.002123 |
| *KDM3C/JMJD1C* | rs34491125 | G | C | D2400E | 0.03031 |
| *KDM3C/JMJD1C* | rs368852030 | T | C | E1577G | 0.0001042 |
| *KDM1B* | rs149734475 | A | C | Q162P | 0.002763 |
| *KDM4C* | rs111451391 | A | C | S534R | 0.002248 |
| *KDM4C* | rs191848178 | G | T | A630S | 0.00002478 |
| *KDM4C* | rs35389625 | A | G | N697S | 0.04486 |
| *KDM4D* | rs372591204 | C | T | R451C | 0.00003346 |
| *KDM5A* | rs200969654 | G | C | S103C | 0.0003761 |
| *KDM6A* | rs138723332 | T | C | C520R | 0.000773 |
| *KDM6A* | rs2230018 | C | A | T778K | 0.1262 |
| *KDM6B* | rs79548905 | G | C | E221D | 0.02436 |
| *KDM6B* | rs749981813 | G | T | R988L | 0.00003139 |
| *KMT2C* | rs138119145 | C | G | D1319H | 0.006602 |

| | | | | | |
|---|---|---|---|---|---|
| *KMT2D* | rs201481646 | C | T | R5266H | 0.0008099 |
| *KMT2D* | rs112921115 | G | A | T1246M | 0.0006531 |
| *KMT2D* | rs201994402 | G | C | R228G | 0.0002146 |
| *KMT5A* | rs376869233 | G | A | A108T | 0.00003531 |
| *LIG1* | rs3730947 | C | T | V349M | 0.002691 |
| *MLH3* | rs138006166 | C | T | A1394T | 0.0003116 |
| *MMS19* | rs29001311 | T | G | Q409P | 0.003389 |
| *MSH6* | rs63749999 | C | T | R1035X | 0.00001055 |
| *MUS81* | rs61754785 | G | A | R432H | 0.01893 |
| *MUTYH* | rs150792276 | G | A | R423C | 0.001309 |
| *MUTYH* | rs200872702 | T | C | I220V | 0.0002007 |
| *POLG* | rs113994096 | G | A | P587L | 0.001985 |
| *POLI* | rs3218784 | A | G | I261M | 0.02309 |
| *POLI* | rs200852409 | C | G | P390R | 0.0001978 |
| *POLM* | rs28382644 | C | G | G220A | 0.008823 |
| *PRDM16* | rs187194973 | G | A | A34T | 0.002882 |
| *PRDM16* | rs554385722 | G | A | A643T | 0.00002298 |
| *PRDM16* | rs149333409 | G | A | V764M | 0.001654 |
| *PRDM16* | rs201814961 | C | T | P889L | 0.0003279 |
| *PRDM2* | rs41269803 | G | A | C334Y | 0.02088 |
| *PRDM2* | rs116238585 | C | G | P949A | 0.0206 |
| *PRDM6* | rs139705595 | A | G | I262V | 0.02423 |
| *PRDM9* | rs202118545 | C | G | T769R | 0.00004862 |
| *PRDM9* | rs77287813 | A | C | N790H | 0.02173 |
| *PRPF19* | rs188856023 | G | T | T91N | 0.0001208 |
| *RAD1* | rs2308957 | C | T | G114D | 0.006455 |
| *RAD51B* | rs34094401 | T | G | L172W | 0.01368 |
| *RAD51B* | rs34594234 | A | G | K243R | 0.01007 |
| *RECQL* | . | G | A | P126L | NA |
| *SETD1B* | rs773794483 | A | C | H8P | 0.004045 |
| *SETD1B* | rs113279920 | C | T | P367L | 0.005827 |
| *SETD1B* | rs144871159 | G | A | A1488T | 0.01712 |
| *SETDB1* | rs150892641 | C | T | A1021V | 0.004248 |
| *SMUG1* | rs3136389 | G | A | R105W | 0.001391 |
| *SMYD2* | rs34259050 | A | G | M384V | 0.01856 |
| *TDG* | rs4135113 | G | A | G199S | 0.03325 |
| *TP53BP1* | rs28903077 | C | T | R1454Q | 0.001453 |
| *UVSSA* | rs116741007 | G | A | G355R | 0.007286 |
| *UVSSA* | rs147609449 | C | T | R445W | 0.001373 |
| *XAB2* | rs61761630 | C | T | E303K | 0.003223 |

ed is gnomAD v4.1.1.

| | | GnomAD frequency | | | |
|---|---|---|---|---|---|
| Admixed American | African/African American | Amish | Ashkenazi Jewish | East Asian | European (Finnish) |
| 0.0004002 | 0.00002685 | 0 | 0.003804 | 0 | 0 |
| 0.03048 | 0.01043 | 0.01645 | 0.07851 | 0.0002005 | 0.07851 |
| 0.001033 | 0.0000533 | 0 | 0 | 0 | 0.00004703 |
| 0.001966 | 0.0002265 | 0 | 0.007028 | 0.0001114 | 0.001334 |
| 0.01074 | 0.002761 | 0.01099 | 0.02785 | 0.00004562 | 0.008271 |
| 0.00001666 | 0 | 0 | 0 | 0 | 0 |
| 0.01088 | 0.003065 | 0.01206 | 0.01716 | 0.00004457 | 0.02079 |
| 0.006664 | 0.00168 | 0 | 0.02276 | 0.00002228 | 0.0002811 |
| 0.002183 | 0.0004263 | 0 | 0.0005066 | 0.00002228 | 0.004107 |
| 0 | 0.00006735 | 0 | 0 | 0 | 0 |
| 0.007322 | 0.003059 | 0.004386 | 0.02431 | 0.00002231 | 0.04287 |
| 0.00006665 | 0.00007993 | 0 | 0 | 0 | 0.009339 |
| 0 | 0 | 0 | 0.0009459 | 0 | 0 |
| 0.00005 | 0.00008009 | 0 | 0 | 0 | 0.00003125 |
| 0.4434 | 0.3778 | 0.8007 | 0.8213 | 0.4271 | 0.7925 |
| 0.0003501 | 0.0003066 | 0 | 0.002027 | 0 | 0.001812 |
| 0.01487 | 0.003992 | 0.04276 | 0.01603 | 0.000204 | 0.05821 |
| 0.00001671 | 0.00008 | 0 | 0 | 0.002741 | 0 |
| 0.0001832 | 0.0002265 | 0 | 0.0001689 | 0 | 0.0000937 |
| 0.0015 | 0.0008396 | 0 | 0.0003716 | 0.00002229 | 0.003624 |
| 0 | 0.00001335 | 0 | 0 | 0.00002227 | 0 |
| 0.01041 | 0.004784 | 0.05154 | 0.01402 | 0.00002228 | 0.07016 |
| 0.00155 | 0.000533 | 0 | 0.001858 | 0.00004457 | 0.0146 |
| 0.007972 | 0.004001 | 0.02212 | 0.00307 | 0 | 0.02795 |
| 0 | 0 | 0 | 0 | 0 | 0 |
| 0.00015 | 0.00001335 | 0 | 0.0051 | 0 | 0 |
| NA | NA | NA | NA | NA | NA |
| 0 | 0.00002669 | 0 | 0 | 0 | 0 |
| 0 | 0 | 0 | 0 | 0 | 0 |
| 0.002835 | 0.00178 | 0 | 0.004979 | 0.0006083 | 0.01598 |
| 0.0006832 | 0.0000533 | 0 | 0 | 0 | 0 |
| 0.0003665 | 0.0002798 | 0 | 0.0007431 | 0 | 0.000203 |
| 0.01133 | 0.002214 | 0.01535 | 0.008545 | 0.02876 | 0.002089 |
| 0.0007048 | 0.0001354 | 0 | 0.0001031 | 0 | 0.00002056 |
| 0.003765 | 0.001213 | 0.0307 | 0.008515 | 0.00004459 | 0.0004883 |
| 0.004019 | 0.001374 | 0.003297 | 0.01396 | 0.00002238 | 0.0002353 |
| 0.001817 | 0.005182 | 0 | 0.0003064 | 0.00002239 | 0.00001574 |
| 0.0005517 | 0.0000934 | 0 | 0.0004758 | 0.00006696 | 0.000048 |
| 0 | 0 | 0 | 0.0002702 | 0 | 0 |
| 0.01376 | 0.01029 | 0.002193 | 0.009356 | 0.00008911 | 0.01241 |
| 0.038 | 0.3207 | 0.03289 | 0.01843 | 0 | 0.01204 |
| 0 | 0.00003996 | 0 | 0 | 0.00002227 | 0 |

| | | | | | |
|---|---|---|---|---|---|
| 0.2081 | 0.07261 | 0.05702 | 0.1134 | 0.5045 | 0.186 |
| 0.002567 | 0.00132 | 0.001096 | 0.01723 | 0 | 0.01416 |
| 0.001817 | 0.001042 | 0 | 0.004532 | 0 | 0.01552 |
| 0.007882 | 0.002759 | 0.004386 | 0.01707 | 0 | 0.06727 |
| 0.006215 | 0.003758 | 0.001096 | 0.002939 | 0.00002228 | 0.003882 |
| 0.01156 | 0.005056 | 0.03728 | 0.04917 | 0.00004466 | 0.03885 |
| 0.04832 | 0.1973 | 0.01754 | 0.07849 | 0.00004458 | 0.05833 |
| 0.04833 | 0.1999 | 0.01754 | 0.07844 | 0.00004456 | 0.05822 |
| 0.0009973 | 0.0002337 | 0 | 0.002952 | 0.00007126 | 0.001059 |
| 0.0019 | 0.0002133 | 0 | 0.009865 | 0.0000223 | 0.00009373 |
| 0.007997 | 0.003123 | 0.01096 | 0.00183 | 0.00002231 | 0.001661 |
| N/A | N/A | N/A | N/A | N/A | N/A |
| 0 | 0 | 0 | 0 | 0.001093 | 0.00001662 |
| 0.01689 | 0.07074 | 0 | 0.06366 | 0.01035 | 0.01489 |
| N/A | N/A | N/A | N/A | N/A | N/A |
| 0.0001166 | 0.00009329 | 0 | 0 | 0 | 0 |
| 0.01066 | 0.002 | 0.008772 | 0.03617 | 0 | 0.0273 |
| 0.0002675 | 0 | 0 | 0 | 0 | 0 |
| 0.002817 | 0.001138 | 0.0008127 | 0.0007014 | 0.00036 | 0.00003383 |
| 0.0006997 | 0.02222 | 0 | 0 | 0 | 0 |
| 0.003293 | 0.0006404 | 0 | 0.006527 | 0 | 0.001984 |
| 0 | 0 | 0 | 0 | 0 | 0 |
| 0 | 0 | 0 | 0 | 0 | 0 |
| 0.01091 | 0.003786 | 0.02544 | 0.02855 | 0.0001137 | 0.01633 |
| 0.0001669 | 0.00005342 | 0 | 0 | 0.00002228 | 0.00003126 |
| 0.0004525 | 0.003553 | 0 | 0 | 0 | 0 |
| 0.05288 | 0.01446 | 0.05592 | 0.1168 | 0.0001782 | 0.03811 |
| 0.04038 | 0.01298 | 0.05263 | 0.09929 | 0.1273 | 0.0956 |
| 0.004665 | 0.00128 | 0 | 0.008781 | 0 | 0.00525 |
| 0 | 0 | 0 | 0.01025 | 0 | 0 |
| 0 | 0.00001334 | 0 | 0.003818 | 0 | 0 |
| 0.00868 | 0.002719 | 0.03194 | 0.002534 | 0.00002229 | 0.005771 |
| 0.002204 | 0.0407 | 0 | 0 | 0 | 0 |
| 0.02043 | 0.006691 | 0.1042 | 0.0856 | 0.0001783 | 0.007125 |
| 0 | 0.00002674 | 0 | 0 | 0 | 0 |
| 0.001433 | 0.0003464 | 0 | 0.00003378 | 0.00002229 | 0.0003279 |
| 0.0545 | 0.001986 | 0 | 0 | 0.0001783 | 0 |
| 0 | 0 | 0 | 0 | 0.0007575 | 0.00001562 |
| 0.02216 | 0.009711 | 0.03838 | 0.02794 | 0.06849 | 0.05791 |
| 0.00006667 | 0.00006672 | 0 | 0 | 0 | 0.00001562 |
| 0.00005001 | 0.0001069 | 0 | 0 | 0 | 0 |
| 0.0001111 | 0.00009023 | 0 | 0.00004639 | 0 | 0.001229 |
| 0.04591 | 0.04176 | 0.03488 | 0.133 | 0.4237 | 0.1604 |
| 0.01541 | 0.005051 | 0.02412 | 0.05435 | 0.00006684 | 0.005295 |
| 0.0004966 | 0.00001363 | 0 | 0 | 0 | 0 |
| 0.003611 | 0.001243 | 0 | 0.001658 | 0.00002237 | 0.006821 |

| | | | | | |
|---|---|---|---|---|---|
| 0.0001054 | 0.0001749 | 0 | 0 | 0 | 0.000016 |
| 0.0002333 | 0.00001334 | 0 | 0.02705 | 0 | 0 |
| 0.00003519 | 0 | 0 | 0.00886 | 0 | 0 |
| 0.00003622 | 0 | 0 | 0.00003439 | 0 | 0 |
| 0.0004664 | 0.0004263 | 0 | 0.009524 | 0.00002228 | 0.01546 |
| 0.00001667 | 0.0000534 | 0 | 0 | 0 | 0.00006247 |
| 0.07453 | 0.001279 | 0 | 0.0002027 | 0.01125 | 0 |
| 0 | 0 | 0 | 0 | 0.00002229 | 0 |
| 0.01128 | 0.003 | 0.1042 | 0.07937 | 0.0001114 | 0.008545 |
| 0.0003499 | 0.0002669 | 0 | 0.00003377 | 0.000401 | 0.0001094 |
| 0.0001167 | 0.00004004 | 0 | 0.00003378 | 0 | 0.00004687 |
| 0.0008995 | 0.0003998 | 0.003289 | 0.001554 | 0 | 0.0005448 |
| 0.01563 | 0.004278 | 0.04276 | 0.04106 | 0.0000224 | 0.02348 |
| 0.00001682 | 0.0001602 | 0 | 0.005707 | 0 | 0 |
| 0.004051 | 0.001532 | 0 | 0.004259 | 0.00004456 | 0.0155 |
| 0.0002166 | 0.0005729 | 0.004386 | 0 | 0.0001337 | 0.002929 |
| 0 | 0.0001225 | 0 | 0 | 0 | 0 |
| 0.0009498 | 0.0003733 | 0 | 0.0002702 | 0.00004461 | 0.0002844 |
| 0.0003001 | 0.00006666 | 0 | 0 | 0.00002229 | 0 |
| 0.006333 | 0.005893 | 0.01535 | 0.02824 | 0.002495 | 0.02455 |
| 0.005484 | 0.002893 | 0.1079 | 0.03003 | 0.01592 | 0.009793 |
| 0.0101 | 0.003731 | 0.007675 | 0.01257 | 0.0001956 | 0.01164 |
| 0.00007622 | 0.0001357 | 0.001887 | 0.0001439 | 0.00004881 | 0.00008777 |
| 0.02474 | 0.01488 | 0.01485 | 0.03916 | 0.00279 | 0.02436 |
| 0 | 0 | 0 | 0.004459 | 0 | 0 |
| 0.005015 | 0.0009861 | 0.001096 | 0.01358 | 0.00002229 | 0.01656 |
| 0.007426 | 0.05433 | 0.004386 | 0.01022 | 0.00004467 | 0.02529 |
| 0.003965 | 0.002175 | 0 | 0.002302 | 0 | 0.008243 |
| NA | NA | NA | NA | NA | NA |
| 0.003167 | 0.009422 | 0.001174 | 0.004969 | 0.0002986 | 0.003564 |
| 0.003313 | 0.0009433 | 0 | 0.001187 | 0.00002445 | 0.003992 |
| 0.01893 | 0.004014 | 0.004484 | 0.008572 | 0 | 0.01977 |
| 0.003433 | 0.03154 | 0 | 0.003547 | 0.004012 | 0 |
| 0.001941 | 0.0002136 | 0 | 0.00003396 | 0 | 0.00001563 |
| 0.01614 | 0.009355 | 0.002193 | 0.06945 | 0.001582 | 0.007122 |
| 0.05989 | 0.1675 | 0.002193 | 0.06206 | 0.165 | 0.01909 |
| 0.0011 | 0.0002799 | 0 | 0.0001689 | 0 | 0.00001562 |
| 0.003466 | 0.001119 | 0 | 0.01365 | 0 | 0.0004546 |
| 0.0002332 | 0.0002665 | 0 | 0.0002702 | 0.00002229 | 0.0002365 |
| 0.001133 | 0.0005995 | 0 | 0.005641 | 0.00002228 | 0.003093 |

| European (Non-Finnish) | Middle Eastern | South Asian | Remaining |
|---|---|---|---|
| 0.0001458 | 0.0008322 | 0.00006625 | 0.0004215 |
| 0.06282 | 0.05856 | 0.01359 | 0.05078 |
| 0.0004797 | 0.000165 | 0 | 0.00024 |
| 0.0009009 | 0.01106 | 0.002075 | 0.001888 |
| 0.01617 | 0.02374 | 0.008549 | 0.01598 |
| 0.00004409 | 0 | 0 | 0.00004802 |
| 0.01751 | 0.01666 | 0.003063 | 0.01489 |
| 0.006437 | 0.01519 | 0.002124 | 0.007748 |
| 0.002746 | 0.003804 | 0.003393 | 0.002384 |
| 0.0001443 | 0.0003318 | 0.00005633 | 0.0002253 |
| 0.02177 | 0.0003674 | 0.001771 | 0.01547 |
| 0.0007442 | 0 | 0 | 0.0008802 |
| 0.000005085 | 0 | 0.00001098 | 0.00006401 |
| 0.0006339 | 0 | 0.00001098 | 0.0008002 |
| 0.7831 | 0.7022 | 0.7281 | 0.71 |
| 0.002044 | 0.0001651 | 0 | 0.001056 |
| 0.03332 | 0.02478 | 0.02086 | 0.02575 |
| 0.0000551 | 0 | 0.0001981 | 0.0001921 |
| 0.001372 | 0.0004949 | 0.003228 | 0.000912 |
| 0.005247 | 0.00165 | 0.003481 | 0.003584 |
| 0.00003898 | 0 | 0.00002196 | 0.00003202 |
| 0.03275 | 0.003629 | 0.002448 | 0.02315 |
| 0.004364 | 0.0009898 | 0.0007905 | 0.003648 |
| 0.02696 | 0.003385 | 0.004986 | 0.0195 |
| 0.000005932 | 0 | 0 | 0 |
| 0.00002034 | 0.0001644 | 0 | 0.0003041 |
| NA | NA | NA | NA |
| 0.00007203 | 0 | 0 | 0.000016 |
| 0.00001441 | 0 | 0 | 0 |
| 0.0124 | 0.001837 | 0.002332 | 0.009483 |
| 0.00005424 | 0.006931 | 0.00001098 | 0.000352 |
| 0.001878 | 0.000165 | 0 | 0.001504 |
| 0.0117 | 0.01402 | 0.0622 | 0.01477 |
| 0.001019 | 0 | 0 | 0.0006465 |
| 0.007988 | 0.00684 | 0.01104 | 0.007812 |
| 0.006198 | 0.01469 | 0.006263 | 0.006191 |
| 0.0002661 | 0.002147 | 0.00004492 | 0.0009475 |
| 0.0007093 | 0.0001803 | 0.0002532 | 0.000402 |
| 0.000001695 | 0 | 0 | 0.00003201 |
| 0.03224 | 0.004454 | 0.001383 | 0.02179 |
| 0.01631 | 0.0515 | 0.008556 | 0.03957 |
| 0.000001695 | 0 | 0.00006587 | 0.000016 |

| | | | |
|---|---|---|---|
| 0.116 | 0.115 | 0.2183 | 0.1477 |
| 0.009427 | 0.00231 | 0.001493 | 0.00744 |
| 0.005081 | 0.0009924 | 0.001044 | 0.003868 |
| 0.02164 | 0.00892 | 0.0008786 | 0.01742 |
| 0.02177 | 0.005114 | 0.006631 | 0.01638 |
| 0.03208 | 0.02938 | 0.005318 | 0.02646 |
| 0.06632 | 0.06426 | 0.01338 | 0.06638 |
| 0.06647 | 0.06467 | 0.01329 | 0.06645 |
| 0.002317 | 0.001878 | 0.002461 | 0.001831 |
| 0.001292 | 0.007763 | 0.0006259 | 0.002112 |
| 0.02112 | 0.007145 | 0.003703 | 0.01522 |
| N/A | N/A | N/A | N/A |
| 0.00005886 | 0 | 0.0001068 | 0.0001159 |
| 0.005268 | 0.04509 | 0.02475 | 0.02061 |
| N/A | N/A | N/A | N/A |
| 0.0001051 | 0.00132 | 0.0008673 | 0.000352 |
| 0.01273 | 0.01601 | 0.008216 | 0.01363 |
| 0.0001339 | 0.000495 | 0.00006598 | 0.000128 |
| 0.000011 | 0 | 0 | 0 |
| 0.00000607 | 0.0006964 | 0.00006295 | 0.001105 |
| 0.005396 | 0.00314 | 0.0008055 | 0.006068 |
| 0.00006276 | 0 | 0 | 0 |
| 0.00003983 | 0 | 0 | 0.00001601 |
| 0.0242 | 0.01186 | 0.004751 | 0.0178 |
| 0.000378 | 0 | 0.0002968 | 0.0001921 |
| 0.000006789 | 0.0009921 | 0.00007713 | 0.000305 |
| 0.07701 | 0.133 | 0.03981 | 0.0669 |
| 0.07502 | 0.09092 | 0.07211 | 0.08081 |
| 0.006517 | 0.02293 | 0.001834 | 0.006079 |
| 0.00003645 | 0 | 0 | 0.0005927 |
| 0.00001695 | 0 | 0 | 0.0002721 |
| 0.01695 | 0.003795 | 0.01072 | 0.01093 |
| 0.00003741 | 0.003153 | 0.0001568 | 0.002883 |
| 0.03006 | 0.06357 | 0.06739 | 0.03371 |
| 0.0001382 | 0 | 0 | 0.00004808 |
| 0.002903 | 0.008746 | 0.007269 | 0.002961 |
| 0.0000322 | 0.001485 | 0 | 0.002464 |
| 0.00000339 | 0 | 0 | 0.000016 |
| 0.04338 | 0.03068 | 0.0934 | 0.04531 |
| 0.00003559 | 0 | 0.00001098 | 0.000016 |
| 0.0004748 | 0 | 0.0001428 | 0.0003687 |
| 0.0009395 | 0.0002368 | 0 | 0.0005926 |
| 0.1184 | 0.09678 | 0.191 | 0.1296 |
| 0.02869 | 0.02177 | 0.005313 | 0.02515 |
| 0.00000868 | 0 | 0 | 0.0001987 |
| 0.007212 | 0.01169 | 0.01041 | 0.00528 |

| | | | |
|---|---|---|---|
| 0.00103 | 0 | 0.00002282 | 0.00102 |
| 0.0001 | 0.0001644 | 0.00003294 | 0.001857 |
| 0.00003417 | 0 | 0 | 0.0006797 |
| 0.00004374 | 0 | 0 | 0.00003252 |
| 0.001972 | 0.001485 | 0.0055 | 0.002784 |
| 0.0004076 | 0 | 0 | 0.000208 |
| 0.00006018 | 0 | 0.001043 | 0.003617 |
| 0.00001357 | 0 | 0 | 0 |
| 0.01965 | 0.01188 | 0.0247 | 0.01813 |
| 0.001667 | 0.0001644 | 0 | 0.001232 |
| 0.0002466 | 0 | 0.00005489 | 0.000224 |
| 0.002444 | 0.00165 | 0.0003623 | 0.001712 |
| 0.0269 | 0.01178 | 0.001869 | 0.02272 |
| 0.00009183 | 0.0003309 | 0 | 0.000434 |
| 0.009969 | 0.009736 | 0.005479 | 0.007073 |
| 0.003521 | 0.000495 | 0.001131 | 0.002209 |
| 0.00001729 | 0 | 0.00005991 | 0.0000332 |
| 0.002016 | 0.0003299 | 0.0009003 | 0.001488 |
| 0.0004145 | 0 | 0.00002197 | 0.0002241 |
| 0.0234 | 0.02903 | 0.01238 | 0.02275 |
| 0.01781 | 0.01898 | 0.08732 | 0.02077 |
| 0.02474 | 0.03057 | 0.06646 | 0.02628 |
| 0.00003624 | 0.0003493 | 0.00004553 | 0.00005039 |
| 0.02283 | 0.03669 | 0.007952 | 0.02446 |
| 0.00003051 | 0 | 0 | 0.0004321 |
| 0.006791 | 0.001815 | 0.001109 | 0.007279 |
| 0.012 | 0.02709 | 0.004967 | 0.01356 |
| 0.01219 | 0.01174 | 0.002971 | 0.008306 |
| NA | NA | NA | NA |
| 0.0038 | 0.009468 | 0.003912 | 0.004974 |
| 0.006877 | 0.0008344 | 0.003652 | 0.005425 |
| 0.01955 | 0.01034 | 0.00487 | 0.01593 |
| 0.0003119 | 0.004639 | 0.03528 | 0.006239 |
| 0.001725 | 0.0008278 | 0.000022 | 0.001091 |
| 0.0193 | 0.07589 | 0.0118 | 0.02257 |
| 0.01825 | 0.07943 | 0.02541 | 0.04375 |
| 0.001824 | 0 | 0.0002086 | 0.001296 |
| 0.00869 | 0.01369 | 0.002712 | 0.007202 |
| 0.001779 | 0 | 0.00004391 | 0.00088 |
| 0.003791 | 0.000495 | 0.0004391 | 0.003296 |

**Supplementary Table 12. Expected genotype frequencies of KDM3C rs10761725 across gnomAD ance**

| Gene | Position | Ref | Alt | AA Change | Total |
|---|---|---|---|---|---|
| *KDM3C/JMJD1C* | rs10761725 | A | T | S464T | 0.7366 |

Expected genotype frequencies were derived from gnomAD allele frequencies assuming Hardy–Weinberg equ

| Formula | Example using Total T allele frequency in KDM3C rs10761725 (q = 0.7366) | Result |
|---|---|---|
| q = T allele frequency | q = 0.7366 | 0.7366 |
| p = 1 − q | p = 1 − 0.7366 | 0.2634 |
| A/A = p² | (0.2634)² | 0.0694 |
| A/T = 2pq | 2 × 0.2634 × 0.7366 | 0.3881 |
| T/T = q² | (0.7366)² | 0.5426 |

| Population | T allele frequency (q) | A allele frequency (p = 1−q) | A/A = p² | A/T = 2pq | T/T = q² |
|---|---|---|---|---|---|
| Total | 0.7366 | 0.2634 | 0.06938 | 0.38804088 | 0.54258 |
| Admixed American | 0.4434 | 0.5566 | 0.309804 | 0.49359288 | 0.196604 |
| African/African | 0.3778 | 0.6222 | 0.387133 | 0.47013432 | 0.142733 |
| Amish | 0.8007 | 0.1993 | 0.03972 | 0.31915902 | 0.64112 |
| Ashkenazi Jewish | 0.8213 | 0.1787 | 0.031934 | 0.29353262 | 0.674534 |
| East Asian | 0.4271 | 0.5729 | 0.328214 | 0.48937118 | 0.182414 |
| European (Finnish) | 0.7925 | 0.2075 | 0.043056 | 0.3288875 | 0.628056 |
| European (Non-Finnish) | 0.7831 | 0.2169 | 0.047046 | 0.33970878 | 0.613246 |
| Middle Eastern | 0.7022 | 0.2978 | 0.088685 | 0.41823032 | 0.493085 |
| South Asian | 0.7281 | 0.2719 | 0.07393 | 0.39594078 | 0.53013 |
| Remaining | 0.71 | 0.29 | 0.0841 | 0.4118 | 0.5041 |

stry groups

| Admixed American | African/African American | Amish | Ashkenazi Jewish | East Asian | European (Finnish) | European (Non-Finnish) | Middle Eastern |
|---|---|---|---|---|---|---|---|
| 0.4434 | 0.3778 | 0.8007 | 0.8213 | 0.4271 | 0.7925 | 0.7831 | 0.7022 |

uilibrium: A/A = $(1-q)^2$, A/T = $2q(1-q)$, and T/T = $q^2$, where q is alternate allele frequency (T) and p represents the

| South Asian | Remaining |
|---|---|
| 0.7281 | 0.71 |

reference allele frequency (A)

**Supplementary Table 13. Somatic mutations in Rectal and HNSC separated by *KDM3C* genotype**

| | |
|---|---|
| **Rectal_APC** | *APC* somatic mutations in rectal seperated by *KDM3C* genotype |
| **Rectal_KRAS** | *KRAS* somatic mutations in rectal seperated by *KDM3C* genotype |
| **Rectal_BRAF** | *BRAF* somatic mutations in rectal seperated by *KDM3C* genotype |
| **Rectal_SMAD4** | *SMAD4* somatic mutations in rectal seperated by *KDM3C* genotype |
| **Rectal_TP53** | *TP53* somatic mutations in rectal seperated by *KDM3C* genotype |
| **HSNC_NOTCH1** | *NOTCH1* somatic mutations in HNSC seperated by *KDM3C* genotype |
| **HNSC_NSD1** | *NSD1* somatic mutations in HNSC seperated by *KDM3C* genotype |
| **HNSC_PIK3CA** | *PIK3CA* somatic mutations in HNSC seperated by *KDM3C* genotype |
| **HNSC_TP53** | *TP53* somatic mutations in HNSC seperated by *KDM3C* genotype |
| **HNSC_CDKN2A** | *CDKN2A* somatic mutations in HNSC seperated by *KDM3C* genotype |
| **HNSC_FAT1** | *FAT1* somatic mutations in HNSC seperated by *KDM3C* genotype |
| **HNSC_KMT2D** | *KMT2D* somatic mutations in HNSC seperated by *KDM3C* genotype |

***APC* somatic mutations in rectal seperated by *KDM3C* genotype**

| Sample ID | Cancer Type | Gene | *KDM3C* genotype | APC protein change |
|---|---|---|---|---|
| 1 | Rectal | *APC* | A/A | R213* |
| 1 | Rectal | *APC* | A/A | S1415Rfs*4 |
| 2 | Rectal | *APC* | A/A | L1382Tfs*4 |
| 2 | Rectal | *APC* | A/A | R564* |
| 3 | Rectal | *APC* | A/A | D1285Afs*5 |
| 4 | Rectal | *APC* | A/A | G1365Vfs*50 |
| 5 | Rectal | *APC* | A/A | R876* |
| 6 | Rectal | *APC* | A/A | I1008Yfs*14 |
| 7 | Rectal | *APC* | A/A | E1397* |
| 7 | Rectal | *APC* | A/A | R1114* |
| 8 | Rectal | *APC* | A/A | D1570Vfs*6 |
| 8 | Rectal | *APC* | A/A | Q757* |
| 9 | Rectal | *APC* | A/A | R213* |
| 9 | Rectal | *APC* | A/A | T1493Rfs*14 |
| 10 | Rectal | *APC* | A/T | W685* |
| 11 | Rectal | *APC* | A/T | Q480Pfs*5 |
| 11 | Rectal | *APC* | A/T | R1331* |
| 12 | Rectal | *APC* | A/T | Q667* |
| 12 | Rectal | *APC* | A/T | S1282* |
| 13 | Rectal | *APC* | A/T | Q1328* |
| 13 | Rectal | *APC* | A/T | S596* |
| 14 | Rectal | *APC* | A/T | Q1367* |
| 15 | Rectal | *APC* | A/T | E582Gfs*20 |
| 16 | Rectal | *APC* | A/T | K1310Dfs*4 |
| 16 | Rectal | *APC* | A/T | Q1260* |
| 17 | Rectal | *APC* | A/T | H1375Lfs*10 |
| 17 | Rectal | *APC* | A/T | R216* |
| 18 | Rectal | *APC* | A/T | P1427Lfs*46 |
| 18 | Rectal | *APC* | A/T | R805* |
| 19 | Rectal | *APC* | A/T | G1288* |
| 19 | Rectal | *APC* | A/T | X582_splice |
| 20 | Rectal | *APC* | A/T | E190* |
| 21 | Rectal | *APC* | A/T | Q1328* |
| 21 | Rectal | *APC* | A/T | S1198* |
| 22 | Rectal | *APC* | A/T | E1379* |
| 22 | Rectal | *APC* | A/T | Q1303* |
| 23 | Rectal | *APC* | A/T | L589* |
| 23 | Rectal | *APC* | A/T | Q1378* |
| 24 | Rectal | *APC* | A/T | E1209* |
| 24 | Rectal | *APC* | A/T | E1397* |
| 25 | Rectal | *APC* | A/T | E1286* |
| 26 | Rectal | *APC* | A/T | R213* |
| 27 | Rectal | *APC* | A/T | S1495Vfs*12 |
| 28 | Rectal | *APC* | A/T | S1327* |
| 29 | Rectal | *APC* | A/T | E1309* |
| 30 | Rectal | *APC* | A/T | R1450* |
| 31 | Rectal | *APC* | A/T | R640Mfs*5 |
| 32 | Rectal | *APC* | A/T | Q1429* |
| 32 | Rectal | *APC* | A/T | R876* |
| 33 | Rectal | *APC* | A/T | R1114* |
| 34 | Rectal | *APC* | A/T | L1488Yfs*25 |
| 34 | Rectal | *APC* | A/T | R856Nfs*6 |

| | | | | |
|---|---|---|---|---|
| 35 | Rectal | *APC* | A/T | K1308* |
| 35 | Rectal | *APC* | A/T | S837* |
| 36 | Rectal | *APC* | A/T | N1548Tfs*17 |
| 36 | Rectal | *APC* | A/T | S583* |
| 37 | Rectal | *APC* | A/T | S1282* |
| 38 | Rectal | *APC* | A/T | G1312* |
| 38 | Rectal | *APC* | A/T | Q1131* |
| 39 | Rectal | *APC* | A/T | R564* |
| 40 | Rectal | *APC* | A/T | Q1367* |
| 41 | Rectal | *APC* | A/T | P1324Sfs*8 |
| 42 | Rectal | *APC* | A/T | Q1338* |
| 43 | Rectal | *APC* | A/T | R1114* |
| 43 | Rectal | *APC* | A/T | R2204* |
| 43 | Rectal | *APC* | A/T | R2237* |
| 43 | Rectal | *APC* | A/T | S1400* |
| 44 | Rectal | *APC* | A/T | Q1378* |
| 45 | Rectal | *APC* | A/T | L1488Yfs*19 |
| 46 | Rectal | *APC* | A/T | S1334F |
| 47 | Rectal | *APC* | A/T | Q1041E |
| 48 | Rectal | *APC* | A/T | S2026Y |
| 48 | Rectal | *APC* | A/T | S294F |
| 49 | Rectal | *APC* | A/T | S948Y |
| 50 | Rectal | *APC* | T/T | E650* |
| 50 | Rectal | *APC* | T/T | P1319Lfs*2 |
| 51 | Rectal | *APC* | T/T | Q542* |
| 51 | Rectal | *APC* | T/T | S1327* |
| 52 | Rectal | *APC* | T/T | R1399Ffs*9 |
| 53 | Rectal | *APC* | T/T | A1446Lfs*27 |
| 53 | Rectal | *APC* | T/T | A426Lfs*28 |
| 54 | Rectal | *APC* | T/T | K1165* |
| 55 | Rectal | *APC* | T/T | T1556Nfs*3 |
| 56 | Rectal | *APC* | T/T | E1295* |
| 57 | Rectal | *APC* | T/T | R283* |
| 58 | Rectal | *APC* | T/T | E1397* |
| 58 | Rectal | *APC* | T/T | N1026Ifs*11 |
| 59 | Rectal | *APC* | T/T | Y935Tfs*20 |
| 60 | Rectal | *APC* | T/T | R1114* |
| 61 | Rectal | *APC* | T/T | S1356* |
| 62 | Rectal | *APC* | T/T | S1042Mfs*6 |
| 62 | Rectal | *APC* | T/T | V1414* |
| 63 | Rectal | *APC* | T/T | Q1294Gfs*6 |
| 63 | Rectal | *APC* | T/T | V830Gfs*12 |
| 64 | Rectal | *APC* | T/T | G1499Pfs*3 |
| 64 | Rectal | *APC* | T/T | R876* |
| 65 | Rectal | *APC* | T/T | Y1376* |
| 66 | Rectal | *APC* | T/T | E1536* |
| 66 | Rectal | *APC* | T/T | R1450* |
| 67 | Rectal | *APC* | T/T | K1085* |
| 67 | Rectal | *APC* | T/T | T1493Rfs*14 |
| 68 | Rectal | *APC* | T/T | L665Ifs*8 |
| 69 | Rectal | *APC* | T/T | P114Ffs*22 |
| 70 | Rectal | *APC* | T/T | E1209* |
| 70 | Rectal | *APC* | T/T | S1411Rfs*4 |
| 71 | Rectal | *APC* | T/T | S1272Ffs*4 |
| 72 | Rectal | *APC* | T/T | E1322* |

| | | | | |
|---|---|---|---|---|
| 73 | Rectal | *APC* | T/T | S837Ifs*7 |
| 74 | Rectal | *APC* | T/T | R856Nfs*6 |
| 74 | Rectal | *APC* | T/T | T1493Rfs*14 |
| 75 | Rectal | *APC* | T/T | R232* |
| 76 | Rectal | *APC* | T/T | Q1378Rfs*37 |
| 77 | Rectal | *APC* | T/T | C1270* |
| 78 | Rectal | *APC* | T/T | E1309* |
| 79 | Rectal | *APC* | T/T | E1322* |
| 79 | Rectal | *APC* | T/T | R302* |
| 80 | Rectal | *APC* | T/T | R1114* |
| 80 | Rectal | *APC* | T/T | R1450* |
| 81 | Rectal | *APC* | T/T | R1450* |
| 82 | Rectal | *APC* | T/T | E428* |
| 82 | Rectal | *APC* | T/T | K1182* |
| 83 | Rectal | *APC* | T/T | S1356* |
| 84 | Rectal | *APC* | T/T | R876* |
| 85 | Rectal | *APC* | T/T | E1353* |
| 86 | Rectal | *APC* | T/T | Q1378* |
| 86 | Rectal | *APC* | T/T | R213* |
| 87 | Rectal | *APC* | T/T | I1311Lfs*10 |
| 87 | Rectal | *APC* | T/T | R876* |
| 88 | Rectal | *APC* | T/T | C1270* |
| 88 | Rectal | *APC* | T/T | R1450* |
| 89 | Rectal | *APC* | T/T | I1417Lfs*2 |
| 89 | Rectal | *APC* | T/T | R876* |
| 90 | Rectal | *APC* | T/T | Q1429* |
| 90 | Rectal | *APC* | T/T | R499* |
| 100 | Rectal | *APC* | T/T | E1353* |
| 100 | Rectal | *APC* | T/T | E1345* |
| 101 | Rectal | *APC* | T/T | R283* |
| 102 | Rectal | *APC* | T/T | R332* |
| 102 | Rectal | *APC* | T/T | T1556Nfs*3 |
| 103 | Rectal | *APC* | T/T | G524Afs*3 |
| 104 | Rectal | *APC* | T/T | Q1429* |
| 105 | Rectal | *APC* | T/T | S1344* |
| 106 | Rectal | *APC* | T/T | R213* |
| 107 | Rectal | *APC* | T/T | Q1338* |
| 108 | Rectal | *APC* | T/T | I1287* |
| 109 | Rectal | *APC* | T/T | E1309* |
| 109 | Rectal | *APC* | T/T | R1114* |
| 110 | Rectal | *APC* | T/T | R216* |
| 111 | Rectal | *APC* | T/T | R230Tfs*20 |
| 112 | Rectal | *APC* | T/T | D849Rfs*2 |
| 113 | Rectal | *APC* | T/T | R876* |
| 113 | Rectal | *APC* | T/T | S1495Vfs*12 |
| 114 | Rectal | *APC* | T/T | A1492Cfs*22 |
| 114 | Rectal | *APC* | T/T | R876* |
| 115 | Rectal | *APC* | T/T | Q1378* |
| 115 | Rectal | *APC* | T/T | R213* |
| 116 | Rectal | *APC* | T/T | E1064* |
| 116 | Rectal | *APC* | T/T | I1580Lfs*69 |
| 117 | Rectal | *APC* | T/T | L1488* |
| 117 | Rectal | *APC* | T/T | R1114* |
| 118 | Rectal | *APC* | T/T | T1556Nfs*3 |
| 119 | Rectal | *APC* | T/T | E1306* |

| | | | | |
|---|---|---|---|---|
| 119 | Rectal | *APC* | T/T | K1165* |
| 120 | Rectal | *APC* | T/T | R876* |
| 120 | Rectal | *APC* | T/T | S1495Vfs*12 |
| 121 | Rectal | *APC* | T/T | R1450* |
| 121 | Rectal | *APC* | T/T | Q1378* |
| 122 | Rectal | *APC* | T/T | S1272* |
| 122 | Rectal | *APC* | T/T | X438_splice |
| 123 | Rectal | *APC* | T/T | Q1367* |
| 124 | Rectal | *APC* | T/T | E1353* |
| 124 | Rectal | *APC* | T/T | R1114* |
| 126 | Rectal | *APC* | T/T | K1165* |
| 126 | Rectal | *APC* | T/T | K1165* |
| 60 | Rectal | *APC* | T/T | N1161K |
| 127 | Rectal | *APC* | T/T | R2431K |
| 128 | Rectal | *APC* | T/T | D774G |
| 128 | Rectal | *APC* | T/T | E136K |
| 128 | Rectal | *APC* | T/T | P2761L |
| 128 | Rectal | *APC* | T/T | S1400L |
| 129 | Rectal | *APC* | T/T | G2709V |
| 130 | Rectal | *APC* | T/T | T518M |
| 131 | Rectal | *APC* | T/T | A3T |
| 131 | Rectal | *APC* | T/T | I718S |
| 131 | Rectal | *APC* | T/T | R332Q |

**Summary Table**

| APC mutation class | A/A | A/T | T/T |
|---|---|---|---|
| Nonsense | 7 | 29 | 49 |
| Frameshift any | 6 | 12 | 30 |
| Splice | 0 | 1 | 1 |
| Missense | 0 | 3 | 6 |

| Mutation type |
|---|
| Nonsense_Mutation |
| Frame_Shift_Del |
| Frame_Shift_Ins |
| Nonsense_Mutation |
| Frame_Shift_Ins |
| Frame_Shift_Del |
| Nonsense_Mutation |
| Frame_Shift_Del |
| Nonsense_Mutation |
| Nonsense_Mutation |
| Frame_Shift_Del |
| Nonsense_Mutation |
| Nonsense_Mutation |
| Frame_Shift_Del |
| Nonsense_Mutation |
| Frame_Shift_Ins |
| Frame_Shift_Ins |
| Nonsense_Mutation |
| Nonsense_Mutation |
| Nonsense_Mutation |
| Nonsense_Mutation |
| Nonsense_Mutation |
| Frame_Shift_Ins |
| Frame_Shift_Del |
| Nonsense_Mutation |
| Frame_Shift_Del |
| Nonsense_Mutation |
| Frame_Shift_Del |
| Nonsense_Mutation |
| Nonsense_Mutation |
| Splice_Site |
| Nonsense_Mutation |
| Nonsense_Mutation |
| Nonsense_Mutation |
| Nonsense_Mutation |
| Nonsense_Mutation |
| Frame_Shift_Del |
| Nonsense_Mutation |
| Nonsense_Mutation |
| Nonsense_Mutation |
| Nonsense_Mutation |
| Nonsense_Mutation |
| Frame_Shift_Del |
| Nonsense_Mutation |
| Nonsense_Mutation |
| Nonsense_Mutation |
| Frame_Shift_Del |
| Nonsense_Mutation |
| Nonsense_Mutation |
| Nonsense_Mutation |
| Frame_Shift_Del |
| Frame_Shift_Ins |

Nonsense_Mutation
Nonsense_Mutation
Frame_Shift_Del
Nonsense_Mutation
Nonsense_Mutation
Nonsense_Mutation
Nonsense_Mutation
Nonsense_Mutation
Nonsense_Mutation
Frame_Shift_Ins
Nonsense_Mutation
Nonsense_Mutation
Nonsense_Mutation
Nonsense_Mutation
Nonsense_Mutation
Nonsense_Mutation
Frame_Shift_Del
Missense_Mutation
Missense_Mutation
Missense_Mutation
Missense_Mutation
Missense_Mutation
Nonsense_Mutation
Frame_Shift_Del
Nonsense_Mutation
Nonsense_Mutation
Frame_Shift_Del
Frame_Shift_Del
Frame_Shift_Del
Nonsense_Mutation
Frame_Shift_Ins
Nonsense_Mutation
Nonsense_Mutation
Frame_Shift_Ins
Frame_Shift_Del
Frame_Shift_Del
Nonsense_Mutation
Nonsense_Mutation
Frame_Shift_Ins
Frame_Shift_Del
Frame_Shift_Del
Frame_Shift_Del
Frame_Shift_Del
Nonsense_Mutation
Nonsense_Mutation
Nonsense_Mutation
Nonsense_Mutation
Nonsense_Mutation
Frame_Shift_Del
Frame_Shift_Del
Frame_Shift_Del
Nonsense_Mutation
Frame_Shift_Del
Frame_Shift_Ins
Nonsense_Mutation

| |
|---|
| Frame_Shift_Ins |
| Frame_Shift_Ins |
| Frame_Shift_Del |
| Nonsense_Mutation |
| Frame_Shift_Del |
| Nonsense_Mutation |
| Nonsense_Mutation |
| Nonsense_Mutation |
| Nonsense_Mutation |
| Nonsense_Mutation |
| Nonsense_Mutation |
| Nonsense_Mutation |
| Nonsense_Mutation |
| Nonsense_Mutation |
| Nonsense_Mutation |
| Nonsense_Mutation |
| Nonsense_Mutation |
| Nonsense_Mutation |
| Nonsense_Mutation |
| Frame_Shift_Del |
| Nonsense_Mutation |
| Nonsense_Mutation |
| Nonsense_Mutation |
| Frame_Shift_Del |
| Nonsense_Mutation |
| Nonsense_Mutation |
| Nonsense_Mutation |
| Nonsense_Mutation |
| Nonsense_Mutation |
| Nonsense_Mutation |
| Nonsense_Mutation |
| Frame_Shift_Ins |
| Frame_Shift_Del |
| Nonsense_Mutation |
| Frame_Shift_Del |
| Nonsense_Mutation |
| Nonsense_Mutation |
| Frame_Shift_Del |
| Nonsense_Mutation |
| Nonsense_Mutation |
| Nonsense_Mutation |
| Frame_Shift_Del |
| Frame_Shift_Ins |
| Nonsense_Mutation |
| Frame_Shift_Del |
| Frame_Shift_Ins |
| Nonsense_Mutation |
| Nonsense_Mutation |
| Nonsense_Mutation |
| Nonsense_Mutation |
| Frame_Shift_Del |
| Nonsense_Mutation |
| Nonsense_Mutation |
| Frame_Shift_Ins |
| Nonsense_Mutation |

| |
|---|
| Nonsense_Mutation |
| Nonsense_Mutation |
| Frame_Shift_Del |
| Nonsense_Mutation |
| Nonsense_Mutation |
| Nonsense_Mutation |
| Splice_Site |
| Nonsense_Mutation |
| Nonsense_Mutation |
| Nonsense_Mutation |
| Nonsense_Mutation |
| Nonsense_Mutation |
| Missense_Mutation |
| Missense_Mutation |
| Missense_Mutation |
| Missense_Mutation |
| Missense_Mutation |
| Missense_Mutation |
| Missense_Mutation |
| Missense_Mutation |
| Missense_Mutation |
| Missense_Mutation |
| Missense_Mutation |

***KRAS* somatic mutations in rectal seperated by *KDM3C* genotype**

| Sample ID | Cancer Type | Gene | *KDM3C* genotype | KRAS protein change |
|---|---|---|---|---|
| 1 | Rectal | *KRAS* | A/A | G12C |
| 2 | Rectal | *KRAS* | A/A | G12A |
| 3 | Rectal | *KRAS* | A/A | G13D |
| 4 | Rectal | *KRAS* | A/A | G12V |
| 5 | Rectal | *KRAS* | A/T | G12C |
| 6 | Rectal | *KRAS* | A/T | G12V |
| 7 | Rectal | *KRAS* | A/T | G12V |
| 8 | Rectal | *KRAS* | A/T | G12D |
| 9 | Rectal | *KRAS* | A/T | Q61E |
| 10 | Rectal | *KRAS* | A/T | Q61H |
| 11 | Rectal | *KRAS* | A/T | G12D |
| 12 | Rectal | *KRAS* | A/T | G13D |
| 13 | Rectal | *KRAS* | A/T | G12S |
| 14 | Rectal | *KRAS* | A/T | G12V |
| 15 | Rectal | *KRAS* | A/T | G12V |
| 16 | Rectal | *KRAS* | A/T | G12D |
| 17 | Rectal | *KRAS* | A/T | G12V |
| 18 | Rectal | *KRAS* | A/T | G12D |
| 19 | Rectal | *KRAS* | A/T | A59T |
| 20 | Rectal | *KRAS* | A/T | G13D |
| 21 | Rectal | *KRAS* | A/T | G12D |
| 22 | Rectal | *KRAS* | A/T | G12A |
| 23 | Rectal | *KRAS* | A/T | G12V |
| 24 | Rectal | *KRAS* | T/T | G12C |
| 25 | Rectal | *KRAS* | T/T | G12C |
| 26 | Rectal | *KRAS* | T/T | G12C |
| 27 | Rectal | *KRAS* | T/T | G12C |
| 28 | Rectal | *KRAS* | T/T | G12C |
| 29 | Rectal | *KRAS* | T/T | G12D |
| 30 | Rectal | *KRAS* | T/T | G13D |
| 31 | Rectal | *KRAS* | T/T | G13D |
| 32 | Rectal | *KRAS* | T/T | G12V |
| 33 | Rectal | *KRAS* | T/T | G13D |
| 34 | Rectal | *KRAS* | T/T | G12V |
| 35 | Rectal | *KRAS* | T/T | G12V |
| 36 | Rectal | *KRAS* | T/T | G12V |
| 37 | Rectal | *KRAS* | T/T | G12D |
| 38 | Rectal | *KRAS* | T/T | G12S |
| 39 | Rectal | *KRAS* | T/T | Q61L |
| 40 | Rectal | *KRAS* | T/T | G12V |
| 41 | Rectal | *KRAS* | T/T | G12D |
| 42 | Rectal | *KRAS* | T/T | G12D |
| 43 | Rectal | *KRAS* | T/T | G12V |
| 44 | Rectal | *KRAS* | T/T | G12S |
| 45 | Rectal | *KRAS* | T/T | G12V |
| 46 | Rectal | *KRAS* | T/T | G12V |
| 47 | Rectal | *KRAS* | T/T | G12D |
| 48 | Rectal | *KRAS* | T/T | G12A |
| 49 | Rectal | *KRAS* | T/T | G12D |
| 50 | Rectal | *KRAS* | T/T | G13D |
| 51 | Rectal | *KRAS* | T/T | G12V |
| 52 | Rectal | *KRAS* | T/T | G13D |

| | | | | |
|---|---|---|---|---|
| 53 | Rectal | *KRAS* | T/T | G12S |
| 54 | Rectal | *KRAS* | T/T | G12S |
| 55 | Rectal | *KRAS* | T/T | K117N |
| 56 | Rectal | *KRAS* | T/T | A146T |
| 57 | Rectal | *KRAS* | T/T | A146T |
| 58 | Rectal | *KRAS* | T/T | A146T |
| 59 | Rectal | *KRAS* | T/T | Q22K |
| 60 | Rectal | *KRAS* | T/T | A11_G12dup |
| 61 | Rectal | *KRAS* | T/T | E98* |

**Summary Table**

| Mutation type | KRAS G12 | A/A | A/T | T/T |
|---|---|---|---|---|
| Missense_Mutation | G12C | 1 | 1 | 5 |
| Missense_Mutation | G12D | 0 | 5 | 6 |
| Missense_Mutation | G12V | 1 | 6 | 9 |
| Missense_Mutation | Other G12 | 1 | 2 | 5 |
| Missense_Mutation | | | | |
| Missense_Mutation | | | | |
| Missense_Mutation | | | | |
| Missense_Mutation | | | | |
| Missense_Mutation | | | | |
| Missense_Mutation | | | | |
| Missense_Mutation | | | | |
| Missense_Mutation | | | | |
| Missense_Mutation | | | | |
| Missense_Mutation | | | | |
| Missense_Mutation | | | | |
| Missense_Mutation | | | | |
| Missense_Mutation | | | | |
| Missense_Mutation | | | | |
| Missense_Mutation | | | | |
| Missense_Mutation | | | | |
| Missense_Mutation | | | | |
| Missense_Mutation | | | | |
| Missense_Mutation | | | | |
| Missense_Mutation | | | | |
| Missense_Mutation | | | | |
| Missense_Mutation | | | | |
| Missense_Mutation | | | | |
| Missense_Mutation | | | | |
| Missense_Mutation | | | | |
| Missense_Mutation | | | | |
| Missense_Mutation | | | | |
| Missense_Mutation | | | | |
| Missense_Mutation | | | | |
| Missense_Mutation | | | | |
| Missense_Mutation | | | | |
| Missense_Mutation | | | | |
| Missense_Mutation | | | | |
| Missense_Mutation | | | | |
| Missense_Mutation | | | | |
| Missense_Mutation | | | | |
| Missense_Mutation | | | | |
| Missense_Mutation | | | | |
| Missense_Mutation | | | | |
| Missense_Mutation | | | | |
| Missense_Mutation | | | | |
| Missense_Mutation | | | | |
| Missense_Mutation | | | | |
| Missense_Mutation | | | | |
| Missense_Mutation | | | | |
| Missense_Mutation | | | | |
| Missense_Mutation | | | | |
| Missense_Mutation | | | | |

| |
|---|
| Missense_Mutation |
| Missense_Mutation |
| Missense_Mutation |
| Missense_Mutation |
| Missense_Mutation |
| Missense_Mutation |
| Missense_Mutation |
| In_Frame_Ins |
| Nonsense_Mutation |

***BRAF* somatic mutations in rectal seperated by *KDM3C* genotype**

| Sample ID | Cancer Type | Gene | *KDM3C* genotype |
|---|---|---|---|
| 1 | Rectal | *BRAF* | A/A |
| 2 | Rectal | *BRAF* | A/T |
| 3 | Rectal | *BRAF* | A/T |
| 4 | Rectal | *BRAF* | T/T |
| 5 | Rectal | *BRAF* | T/T |
| 6 | Rectal | *BRAF* | T/T |

| BRAF protein change | Mutation type |
|---|---|
| V600E | Missense_Mutation |
| F294L | Missense_Mutation |
| R389C | Missense_Mutation |
| H540Q | Missense_Mutation |
| F247L | Missense_Mutation |
| X287_splice | Splice_Site |

**Summary Table**

| BRAF mutations | A/A | A/T |
|---|---|---|
| V600E | 1 | 0 |
| Non-V600 missense | 0 | 2 |
| Splice | 0 | 0 |

| **T/T** |
| --- |
| 0 |
| 2 |
| 1 |

***SMAD4* somatic mutations in rectal seperated by *KDM3C* genotype**

| Sample ID | Cancer Type | Gene | *KDM3C* genotype | SMAD4 protein change |
|---|---|---|---|---|
| 1 | Rectal | *SMAD4* | A/A | R361C |
| 2 | Rectal | *SMAD4* | A/A | D537H |
| 3 | Rectal | *SMAD4* | A/T | R361H |
| 4 | Rectal | *SMAD4* | A/T | R361H |
| 5 | Rectal | *SMAD4* | A/T | D537V |
| 6 | Rectal | *SMAD4* | A/T | F408L |
| 7 | Rectal | *SMAD4* | T/T | G386D |
| 8 | Rectal | *SMAD4* | T/T | D537Y |
| 9 | Rectal | *SMAD4* | T/T | P356L |
| 10 | Rectal | *SMAD4* | T/T | C363del |
| 11 | Rectal | *SMAD4* | T/T | D537E |
| 12 | Rectal | *SMAD4* | T/T | S325Yfs*5 |
| 13 | Rectal | *SMAD4* | T/T | K428M |
| 14 | Rectal | *SMAD4* | T/T | N285Tfs*51 |
| 15 | Rectal | *SMAD4* | T/T | L57V |
| 16 | Rectal | *SMAD4* | T/T | H92Y |
| 17 | Rectal | *SMAD4* | T/T | L47Q |
| 18 | Rectal | *SMAD4* | T/T | L104F |
| 19 | Rectal | *SMAD4* | T/T | L495R |
| 20 | Rectal | *SMAD4* | T/T | R445Q |
| 21 | Rectal | *SMAD4* | T/T | G352_Y353del |

| Mutation type |
| --- |
| Missense_Mutation |
| Missense_Mutation |
| Missense_Mutation |
| Missense_Mutation |
| Missense_Mutation |
| Missense_Mutation |
| Missense_Mutation |
| Missense_Mutation |
| Missense_Mutation |
| In_Frame_Del |
| Missense_Mutation |
| Frame_Shift_Del |
| Missense_Mutation |
| Frame_Shift_Del |
| Missense_Mutation |
| Missense_Mutation |
| Missense_Mutation |
| Missense_Mutation |
| Missense_Mutation |
| Missense_Mutation |
| In_Frame_Del |

**Summary Table**

| SMAD4 mutation class | A/A | A/T | T/T |
| --- | --- | --- | --- |
| Missense | 2 | 4 | 11 |
| Frameshift | 0 | 0 | 2 |
| In-frame deletion | 0 | 0 | 2 |

| SMAD4 hotspot | A/A | A/T | T/T |
| --- | --- | --- | --- |
| R361 | 1 | 2 | 0 |
| D537 | 1 | 1 | 2 |
| C363del | 0 | 0 | 1 |
| G352_Y353del | 0 | 0 | 1 |
| Other SMAD4 mutation | 0 | 1 | 11 |

***TP53* somatic mutations in rectal seperated by *KDM3C* genotype**

| Sample ID | Cancer Type | Gene | *KDM3C* genotype | TP53 protein change |
|---|---|---|---|---|
| 1 | Rectal | *TP53* | A/A | R248Q |
| 2 | Rectal | *TP53* | A/A | R248Q |
| 3 | Rectal | *TP53* | A/A | R175H |
| 4 | Rectal | *TP53* | A/A | R282W |
| 5 | Rectal | *TP53* | A/A | A159V |
| 6 | Rectal | *TP53* | A/A | A138V |
| 7 | Rectal | *TP53* | A/A | H193N |
| 8 | Rectal | *TP53* | A/A | X126_splice |
| 9 | Rectal | *TP53* | A/A | E287* |
| 10 | Rectal | *TP53* | A/T | G245D |
| 11 | Rectal | *TP53* | A/T | R175H |
| 12 | Rectal | *TP53* | A/T | P278A |
| 13 | Rectal | *TP53* | A/T | R273H |
| 14 | Rectal | *TP53* | A/T | R273H |
| 15 | Rectal | *TP53* | A/T | R175H |
| 16 | Rectal | *TP53* | A/T | R175H |
| 17 | Rectal | *TP53* | A/T | R248Q |
| 18 | Rectal | *TP53* | A/T | R282W |
| 19 | Rectal | *TP53* | A/T | C238Y |
| 20 | Rectal | *TP53* | A/T | R273H |
| 21 | Rectal | *TP53* | A/T | R273C |
| 22 | Rectal | *TP53* | A/T | R337S |
| 23 | Rectal | *TP53* | A/T | R273C |
| 24 | Rectal | *TP53* | A/T | R273C |
| 25 | Rectal | *TP53* | A/T | R175H |
| 26 | Rectal | *TP53* | A/T | V272G |
| 27 | Rectal | *TP53* | A/T | P151S |
| 28 | Rectal | *TP53* | A/T | R213* |
| 29 | Rectal | *TP53* | A/T | R213* |
| 30 | Rectal | *TP53* | A/T | I195F |
| 31 | Rectal | *TP53* | A/T | E286G |
| 32 | Rectal | *TP53* | A/T | P278T |
| 33 | Rectal | *TP53* | A/T | X33_splice |
| 34 | Rectal | *TP53* | A/T | S127P |
| 35 | Rectal | *TP53* | A/T | E258G |
| 36 | Rectal | *TP53* | A/T | X33_splice |
| 37 | Rectal | *TP53* | A/T | K319Afs*19 |
| 38 | Rectal | *TP53* | A/T | R306* |
| 39 | Rectal | *TP53* | A/T | E180* |
| 40 | Rectal | *TP53* | A/T | F109Lfs*36 |
| 41 | Rectal | *TP53* | A/T | Q136* |
| 42 | Rectal | *TP53* | A/T | W53Mfs*4 |
| 43 | Rectal | *TP53* | A/T | T211Ffs*4 |
| 44 | Rectal | *TP53* | T/T | R248Q |
| 45 | Rectal | *TP53* | T/T | E286K |
| 46 | Rectal | *TP53* | T/T | R175H |
| 47 | Rectal | *TP53* | T/T | C275Y |
| 48 | Rectal | *TP53* | T/T | R248Q |
| 49 | Rectal | *TP53* | T/T | R175H |
| 50 | Rectal | *TP53* | T/T | R248W |
| 51 | Rectal | *TP53* | T/T | L194R |
| 52 | Rectal | *TP53* | T/T | R282W |

| | | | | |
|---|---|---|---|---|
| 53 | Rectal | *TP53* | T/T | P250L |
| 54 | Rectal | *TP53* | T/T | R273C |
| 55 | Rectal | *TP53* | T/T | R248W |
| 56 | Rectal | *TP53* | T/T | R273H |
| 57 | Rectal | *TP53* | T/T | R282W |
| 58 | Rectal | *TP53* | T/T | G245S |
| 59 | Rectal | *TP53* | T/T | R175H |
| 60 | Rectal | *TP53* | T/T | R282W |
| 61 | Rectal | *TP53* | T/T | R282W |
| 62 | Rectal | *TP53* | T/T | R282W |
| 63 | Rectal | *TP53* | T/T | R282W |
| 64 | Rectal | *TP53* | T/T | R248W |
| 65 | Rectal | *TP53* | T/T | R248Q |
| 66 | Rectal | *TP53* | T/T | G245S |
| 67 | Rectal | *TP53* | T/T | C275Y |
| 68 | Rectal | *TP53* | T/T | R175H |
| 69 | Rectal | *TP53* | T/T | R248W |
| 70 | Rectal | *TP53* | T/T | P151H |
| 71 | Rectal | *TP53* | T/T | G266V |
| 72 | Rectal | *TP53* | T/T | A138V |
| 73 | Rectal | *TP53* | T/T | R213* |
| 74 | Rectal | *TP53* | T/T | R213Q |
| 75 | Rectal | *TP53* | T/T | I195T |
| 76 | Rectal | *TP53* | T/T | V274L |
| 77 | Rectal | *TP53* | T/T | I195T |
| 78 | Rectal | *TP53* | T/T | I232N |
| 79 | Rectal | *TP53* | T/T | Y205D |
| 80 | Rectal | *TP53* | T/T | X331_splice |
| 81 | Rectal | *TP53* | T/T | G262V |
| 82 | Rectal | *TP53* | T/T | Y205S |
| 83 | Rectal | *TP53* | T/T | S215I |
| 84 | Rectal | *TP53* | T/T | X187_splice |
| 85 | Rectal | *TP53* | T/T | X126_splice |
| 86 | Rectal | *TP53* | T/T | P219Afs*3 |
| 87 | Rectal | *TP53* | T/T | S94Cfs*30 |
| 88 | Rectal | *TP53* | T/T | R342* |
| 89 | Rectal | *TP53* | T/T | R335Qfs*2 |
| 90 | Rectal | *TP53* | T/T | V122Dfs*26 |
| 91 | Rectal | *TP53* | T/T | Q331* |
| 92 | Rectal | *TP53* | T/T | L35Pfs*10 |
| 93 | Rectal | *TP53* | T/T | E285* |
| 94 | Rectal | *TP53* | T/T | Y234Pfs*7 |
| 95 | Rectal | *TP53* | T/T | R196* |
| 96 | Rectal | *TP53* | T/T | G108_H115del |
| 97 | Rectal | *TP53* | T/T | R306* |
| 98 | Rectal | *TP53* | T/T | W146* |
| 99 | Rectal | *TP53* | T/T | R196* |
| 100 | Rectal | *TP53* | T/T | E171* |
| 101 | Rectal | *TP53* | T/T | C124Ffs*47 |
| 102 | Rectal | *TP53* | T/T | E51* |

| Mutation type |
| --- |
| Missense_Mutation |
| Missense_Mutation |
| Missense_Mutation |
| Missense_Mutation |
| Missense_Mutation |
| Missense_Mutation |
| Missense_Mutation |
| Splice_Site |
| Nonsense_Mutation |
| Missense_Mutation |
| Missense_Mutation |
| Missense_Mutation |
| Missense_Mutation |
| Missense_Mutation |
| Missense_Mutation |
| Missense_Mutation |
| Missense_Mutation |
| Missense_Mutation |
| Missense_Mutation |
| Missense_Mutation |
| Missense_Mutation |
| Missense_Mutation |
| Missense_Mutation |
| Missense_Mutation |
| Missense_Mutation |
| Missense_Mutation |
| Missense_Mutation |
| Nonsense_Mutation |
| Nonsense_Mutation |
| Missense_Mutation |
| Missense_Mutation |
| Missense_Mutation |
| Splice_Site |
| Missense_Mutation |
| Missense_Mutation |
| Splice_Site |
| Frame_Shift_Ins |
| Nonsense_Mutation |
| Nonsense_Mutation |
| Frame_Shift_Del |
| Nonsense_Mutation |
| Frame_Shift_Ins |
| Frame_Shift_Del |
| Missense_Mutation |
| Missense_Mutation |
| Missense_Mutation |
| Missense_Mutation |
| Missense_Mutation |
| Missense_Mutation |
| Missense_Mutation |
| Missense_Mutation |
| Missense_Mutation |

**Summary Table**

| **TP53 mutations** | **A/A** | **A/T** | **T/T** |
| --- | --- | --- | --- |
| Missense | 7 | 24 | 38 |
| Nonsense | 1 | 5 | 10 |
| Splice | 1 | 2 | 3 |
| Frameshift any | 0 | 4 | 7 |
| In-frame deletion | 0 | 0 | 1 |

| **TP53 mutation subtype** | **A/A** | **A/T** | **T/T** |
| --- | --- | --- | --- |
| R175H | 1 | 4 | 4 |
| R248Q | 2 | 1 | 3 |
| R248W | 0 | 0 | 4 |
| R273H | 0 | 3 | 1 |
| R273C | 0 | 3 | 1 |
| R282W | 1 | 1 | 6 |
| G245S | 0 | 0 | 2 |
| Y220C | 0 | 0 | 0 |
| Other TP53 mutation | 5 | 22 | 38 |

| |
|---|
| Missense_Mutation |
| Missense_Mutation |
| Missense_Mutation |
| Missense_Mutation |
| Missense_Mutation |
| Missense_Mutation |
| Missense_Mutation |
| Missense_Mutation |
| Missense_Mutation |
| Missense_Mutation |
| Missense_Mutation |
| Missense_Mutation |
| Missense_Mutation |
| Missense_Mutation |
| Missense_Mutation |
| Missense_Mutation |
| Missense_Mutation |
| Missense_Mutation |
| Missense_Mutation |
| Missense_Mutation |
| Nonsense_Mutation |
| Missense_Mutation |
| Missense_Mutation |
| Missense_Mutation |
| Missense_Mutation |
| Missense_Mutation |
| Missense_Mutation |
| Splice_Site |
| Missense_Mutation |
| Missense_Mutation |
| Missense_Mutation |
| Splice_Site |
| Splice_Site |
| Frame_Shift_Ins |
| Frame_Shift_Ins |
| Nonsense_Mutation |
| Frame_Shift_Ins |
| Frame_Shift_Del |
| Nonsense_Mutation |
| Frame_Shift_Ins |
| Nonsense_Mutation |
| Frame_Shift_Ins |
| Nonsense_Mutation |
| In_Frame_Del |
| Nonsense_Mutation |
| Nonsense_Mutation |
| Nonsense_Mutation |
| Nonsense_Mutation |
| Frame_Shift_Ins |
| Nonsense_Mutation |

***NOTCH1* somatic mutations in HNSC seperated by *KDM3C* genotype**

| Sample ID | Cancer Type | Gene | *KDM3C* genotype | NOTCH1 protein change |
|---|---|---|---|---|
| 1 | HNSC | *NOTCH1* | A/A | C739* |
| 2 | HNSC | *NOTCH1* | A/A | G1119* |
| 3 | HNSC | *NOTCH1* | A/A | E1450* |
| 4 | HNSC | *NOTCH1* | A/A | X2061_splice |
| 5 | HNSC | *NOTCH1* | A/A | S647Pfs*123 |
| 6 | HNSC | *NOTCH1* | A/A | A2023T |
| 7 | HNSC | *NOTCH1* | A/A | C1085W |
| 8 | HNSC | *NOTCH1* | A/A | P2064L |
| 9 | HNSC | *NOTCH1* | A/A | G1765D |
| 10 | HNSC | *NOTCH1* | A/T | E455K |
| 11 | HNSC | *NOTCH1* | A/T | C440F |
| 12 | HNSC | *NOTCH1* | A/T | A465T |
| 13 | HNSC | *NOTCH1* | A/T | C381Vfs*250 |
| 14 | HNSC | *NOTCH1* | A/T | X2060_splice |
| 15 | HNSC | *NOTCH1* | A/T | X1215_splice |
| 16 | HNSC | *NOTCH1* | A/T | Q1978Pfs*17 |
| 17 | HNSC | *NOTCH1* | A/T | Q1845* |
| 18 | HNSC | *NOTCH1* | A/T | Q1080* |
| 19 | HNSC | *NOTCH1* | A/T | Q1974* |
| 20 | HNSC | *NOTCH1* | A/T | G635Qfs*138 |
| 21 | HNSC | *NOTCH1* | A/T | G684Afs*88 |
| 22 | HNSC | *NOTCH1* | A/T | E124Gfs*19 |
| 23 | HNSC | *NOTCH1* | A/T | G1710Afs*88 |
| 24 | HNSC | *NOTCH1* | A/T | E294* |
| 25 | HNSC | *NOTCH1* | A/T | Q1864* |
| 26 | HNSC | *NOTCH1* | A/T | L2047F |
| 27 | HNSC | *NOTCH1* | A/T | C387Y |
| 28 | HNSC | *NOTCH1* | A/T | P67T |
| 29 | HNSC | *NOTCH1* | A/T | L97V |
| 30 | HNSC | *NOTCH1* | A/T | C954Y |
| 31 | HNSC | *NOTCH1* | A/T | G484V |
| 32 | HNSC | *NOTCH1* | A/T | C478Y |
| 33 | HNSC | *NOTCH1* | A/T | D1520H |
| 34 | HNSC | *NOTCH1* | A/T | N454T |
| 35 | HNSC | *NOTCH1* | A/T | C429Y |
| 36 | HNSC | *NOTCH1* | A/T | E2071K |
| 37 | HNSC | *NOTCH1* | A/T | V324D |
| 38 | HNSC | *NOTCH1* | A/T | D352V |
| 39 | HNSC | *NOTCH1* | A/T | G1401E |
| 40 | HNSC | *NOTCH1* | A/T | E424K |
| 41 | HNSC | *NOTCH1* | A/T | G1894S |
| 42 | HNSC | *NOTCH1* | A/T | L830Q |
| 43 | HNSC | *NOTCH1* | T/T | E455K |
| 44 | HNSC | *NOTCH1* | T/T | E455K |
| 45 | HNSC | *NOTCH1* | T/T | F357del |
| 46 | HNSC | *NOTCH1* | T/T | C440R |
| 47 | HNSC | *NOTCH1* | T/T | A465T |
| 48 | HNSC | *NOTCH1* | T/T | Q513Hfs*116 |
| 49 | HNSC | *NOTCH1* | T/T | C138* |
| 50 | HNSC | *NOTCH1* | T/T | D1942Rfs*51 |
| 51 | HNSC | *NOTCH1* | T/T | E383* |
| 52 | HNSC | *NOTCH1* | T/T | E948* |

| | | | | |
|---|---|---|---|---|
| 53 | HNSC | *NOTCH1* | T/T | L418Rfs*214 |
| 54 | HNSC | *NOTCH1* | T/T | K542Sfs*89 |
| 55 | HNSC | *NOTCH1* | T/T | E1148Sfs*31 |
| 56 | HNSC | *NOTCH1* | T/T | C1063* |
| 57 | HNSC | *NOTCH1* | T/T | Q803* |
| 58 | HNSC | *NOTCH1* | T/T | C1225* |
| 59 | HNSC | *NOTCH1* | T/T | W1089* |
| 60 | HNSC | *NOTCH1* | T/T | C1311Afs*134 |
| 61 | HNSC | *NOTCH1* | T/T | L1531Cfs*49 |
| 62 | HNSC | *NOTCH1* | T/T | D1124Afs*15 |
| 63 | HNSC | *NOTCH1* | T/T | Y550* |
| 64 | HNSC | *NOTCH1* | T/T | Y662* |
| 65 | HNSC | *NOTCH1* | T/T | C1169Afs*10 |
| 66 | HNSC | *NOTCH1* | T/T | D1267Gfs*24 |
| 67 | HNSC | *NOTCH1* | T/T | C461Y |
| 68 | HNSC | *NOTCH1* | T/T | C456Y |
| 69 | HNSC | *NOTCH1* | T/T | L468R |
| 70 | HNSC | *NOTCH1* | T/T | G481C |
| 71 | HNSC | *NOTCH1* | T/T | D142H |
| 72 | HNSC | *NOTCH1* | T/T | C1085Y |
| 73 | HNSC | *NOTCH1* | T/T | C478S |
| 74 | HNSC | *NOTCH1* | T/T | P422S |
| 75 | HNSC | *NOTCH1* | T/T | E1305K |
| 76 | HNSC | *NOTCH1* | T/T | V1222M |
| 77 | HNSC | *NOTCH1* | T/T | D338N |
| 78 | HNSC | *NOTCH1* | T/T | G434V |
| 79 | HNSC | *NOTCH1* | T/T | E1404K |
| 80 | HNSC | *NOTCH1* | T/T | N1875K |
| 81 | HNSC | *NOTCH1* | T/T | C893S |
| 82 | HNSC | *NOTCH1* | T/T | R879W |
| 83 | HNSC | *NOTCH1* | T/T | S385F |
| 84 | HNSC | *NOTCH1* | T/T | K1317N |
| 85 | HNSC | *NOTCH1* | T/T | T1996M |
| 86 | HNSC | *NOTCH1* | T/T | C980G |
| 87 | HNSC | *NOTCH1* | T/T | C933F |
| 88 | HNSC | *NOTCH1* | T/T | D680N |
| 89 | HNSC | *NOTCH1* | T/T | P262A |
| 90 | HNSC | *NOTCH1* | T/T | S608C |
| 91 | HNSC | *NOTCH1* | T/T | W327C |
| 92 | HNSC | *NOTCH1* | T/T | R353C |
| 93 | HNSC | *NOTCH1* | T/T | P422T |

**Summary Table**

| NOTCH1 mutations | A/A | A/T | T/T |
|---|---|---|---|
| Missense | 4 | 20 | 31 |
| Nonsense | 3 | 5 | 9 |
| Frameshift any | 1 | 6 | 11 |
| Splice | 1 | 2 | 0 |
| In-frame deletion | 0 | 0 | 1 |

| Mutation type |
|---|
| Nonsense_Mutation |
| Nonsense_Mutation |
| Nonsense_Mutation |
| Splice_Site |
| Frame_Shift_Del |
| Missense_Mutation |
| Missense_Mutation |
| Missense_Mutation |
| Missense_Mutation |
| Missense_Mutation |
| Missense_Mutation |
| Missense_Mutation |
| Frame_Shift_Del |
| Splice_Site |
| Splice_Site |
| Frame_Shift_Ins |
| Nonsense_Mutation |
| Nonsense_Mutation |
| Nonsense_Mutation |
| Frame_Shift_Ins |
| Frame_Shift_Del |
| Frame_Shift_Ins |
| Frame_Shift_Del |
| Nonsense_Mutation |
| Nonsense_Mutation |
| Missense_Mutation |
| Missense_Mutation |
| Missense_Mutation |
| Missense_Mutation |
| Missense_Mutation |
| Missense_Mutation |
| Missense_Mutation |
| Missense_Mutation |
| Missense_Mutation |
| Missense_Mutation |
| Missense_Mutation |
| Missense_Mutation |
| Missense_Mutation |
| Missense_Mutation |
| Missense_Mutation |
| Missense_Mutation |
| Missense_Mutation |
| Missense_Mutation |
| Missense_Mutation |
| In_Frame_Del |
| Missense_Mutation |
| Missense_Mutation |
| Frame_Shift_Del |
| Nonsense_Mutation |
| Frame_Shift_Del |
| Nonsense_Mutation |
| Nonsense_Mutation |

| |
|---|
| Frame_Shift_Ins |
| Frame_Shift_Del |
| Frame_Shift_Del |
| Nonsense_Mutation |
| Nonsense_Mutation |
| Nonsense_Mutation |
| Nonsense_Mutation |
| Frame_Shift_Del |
| Frame_Shift_Del |
| Frame_Shift_Del |
| Nonsense_Mutation |
| Nonsense_Mutation |
| Frame_Shift_Del |
| Frame_Shift_Ins |
| Missense_Mutation |
| Missense_Mutation |
| Missense_Mutation |
| Missense_Mutation |
| Missense_Mutation |
| Missense_Mutation |
| Missense_Mutation |
| Missense_Mutation |
| Missense_Mutation |
| Missense_Mutation |
| Missense_Mutation |
| Missense_Mutation |
| Missense_Mutation |
| Missense_Mutation |
| Missense_Mutation |
| Missense_Mutation |
| Missense_Mutation |
| Missense_Mutation |
| Missense_Mutation |
| Missense_Mutation |
| Missense_Mutation |
| Missense_Mutation |
| Missense_Mutation |
| Missense_Mutation |
| Missense_Mutation |
| Missense_Mutation |
| Missense_Mutation |

***NSD1* somatic mutations in HNSC seperated by *KDM3C* genotype**

| Sample ID | Cancer Type | Gene | *KDM3C* genotype | NSD1 protein change |
|---|---|---|---|---|
| 1 | HNSC | *NSD1* | A/A | G1928* |
| 2 | HNSC | *NSD1* | A/A | G959* |
| 3 | HNSC | *NSD1* | A/A | Q679* |
| 4 | HNSC | *NSD1* | A/A | G2666Efs*95 |
| 5 | HNSC | *NSD1* | A/A | R788* |
| 6 | HNSC | *NSD1* | A/A | L2054R |
| 7 | HNSC | *NSD1* | A/A | G1678W |
| 8 | HNSC | *NSD1* | A/A | S1528Y |
| 9 | HNSC | *NSD1* | A/T | C1710S |
| 10 | HNSC | *NSD1* | A/T | E990* |
| 11 | HNSC | *NSD1* | A/T | F1292Ifs*4 |
| 12 | HNSC | *NSD1* | A/T | G1095Kfs*2 |
| 13 | HNSC | *NSD1* | A/T | W1160* |
| 14 | HNSC | *NSD1* | A/T | R1948Pfs*16 |
| 15 | HNSC | *NSD1* | A/T | R1700* |
| 16 | HNSC | *NSD1* | A/T | R1984* |
| 17 | HNSC | *NSD1* | A/T | C792* |
| 18 | HNSC | *NSD1* | A/T | K601* |
| 19 | HNSC | *NSD1* | A/T | R1320* |
| 20 | HNSC | *NSD1* | A/T | A1837Sfs*10 |
| 21 | HNSC | *NSD1* | A/T | T922Rfs*6 |
| 22 | HNSC | *NSD1* | A/T | P1665L |
| 23 | HNSC | *NSD1* | A/T | C1897F |
| 24 | HNSC | *NSD1* | A/T | R1948L |
| 25 | HNSC | *NSD1* | A/T | R1948S |
| 26 | HNSC | *NSD1* | A/T | Y1997C |
| 27 | HNSC | *NSD1* | A/T | E2467Q |
| 28 | HNSC | *NSD1* | A/T | S96C |
| 29 | HNSC | *NSD1* | A/T | C1619S |
| 30 | HNSC | *NSD1* | A/T | V366L |
| 31 | HNSC | *NSD1* | A/T | P1665del |
| 32 | HNSC | *NSD1* | A/T | R2005Q |
| 33 | HNSC | *NSD1* | A/T | R1634Q |
| 34 | HNSC | *NSD1* | A/T | G566E |
| 35 | HNSC | *NSD1* | T/T | R1200Lfs*5 |
| 36 | HNSC | *NSD1* | T/T | L590Ifs*5 |
| 37 | HNSC | *NSD1* | T/T | K1433* |
| 38 | HNSC | *NSD1* | T/T | S744* |
| 39 | HNSC | *NSD1* | T/T | E1391* |
| 40 | HNSC | *NSD1* | T/T | W2032* |
| 41 | HNSC | *NSD1* | T/T | X2155_splice |
| 42 | HNSC | *NSD1* | T/T | E1501* |
| 43 | HNSC | *NSD1* | T/T | E1979* |
| 44 | HNSC | *NSD1* | T/T | M1531Nfs*4 |
| 45 | HNSC | *NSD1* | T/T | M1525Cfs*49 |
| 46 | HNSC | *NSD1* | T/T | Q1989* |
| 47 | HNSC | *NSD1* | T/T | R1473* |
| 48 | HNSC | *NSD1* | T/T | E1534* |
| 49 | HNSC | *NSD1* | T/T | S1359* |
| 50 | HNSC | *NSD1* | T/T | S1086Rfs*6 |
| 51 | HNSC | *NSD1* | T/T | D984* |
| 52 | HNSC | *NSD1* | T/T | E1758Rfs*5 |

| | | | | |
|---|---|---|---|---|
| 53 | HNSC | *NSD1* | T/T | P1942Sfs*4 |
| 54 | HNSC | *NSD1* | T/T | T2029Hfs*6 |
| 55 | HNSC | *NSD1* | T/T | K754Nfs*14 |
| 56 | HNSC | *NSD1* | T/T | G1095Afs*3 |
| 57 | HNSC | *NSD1* | T/T | G1953Vfs*16 |
| 58 | HNSC | *NSD1* | T/T | E1516* |
| 59 | HNSC | *NSD1* | T/T | I1873Kfs*18 |
| 60 | HNSC | *NSD1* | T/T | E1853* |
| 61 | HNSC | *NSD1* | T/T | E1970A |
| 62 | HNSC | *NSD1* | T/T | W1769C |
| 63 | HNSC | *NSD1* | T/T | D1489N |
| 64 | HNSC | *NSD1* | T/T | T723S |
| 65 | HNSC | *NSD1* | T/T | A2009T |
| 66 | HNSC | *NSD1* | T/T | D1992H |
| 67 | HNSC | *NSD1* | T/T | D2002H |
| 68 | HNSC | *NSD1* | T/T | E1520K |
| 69 | HNSC | *NSD1* | T/T | H1616Y |

**Summary Table**

| NSD1 mutations | A/A | A/T | T/T |
|---|---|---|---|
| Missense | 3 | 11 | 7 |
| Nonsense | 4 | 7 | 11 |
| Frameshift any | 1 | 5 | 13 |
| Splice | 0 | 0 | 1 |
| In-frame deletion | 0 | 1 | 0 |

| Mutation type |
|---|
| Nonsense_Mutation |
| Nonsense_Mutation |
| Nonsense_Mutation |
| Frame_Shift_Del |
| Nonsense_Mutation |
| Missense_Mutation |
| Missense_Mutation |
| Missense_Mutation |
| Missense_Mutation |
| Nonsense_Mutation |
| Frame_Shift_Ins |
| Frame_Shift_Del |
| Nonsense_Mutation |
| Frame_Shift_Ins |
| Nonsense_Mutation |
| Nonsense_Mutation |
| Nonsense_Mutation |
| Nonsense_Mutation |
| Nonsense_Mutation |
| Frame_Shift_Ins |
| Frame_Shift_Del |
| Missense_Mutation |
| Missense_Mutation |
| Missense_Mutation |
| Missense_Mutation |
| Missense_Mutation |
| Missense_Mutation |
| Missense_Mutation |
| Missense_Mutation |
| Missense_Mutation |
| In_Frame_Del |
| Missense_Mutation |
| Missense_Mutation |
| Missense_Mutation |
| Frame_Shift_Del |
| Frame_Shift_Del |
| Nonsense_Mutation |
| Nonsense_Mutation |
| Nonsense_Mutation |
| Nonsense_Mutation |
| Splice_Site |
| Nonsense_Mutation |
| Nonsense_Mutation |
| Frame_Shift_Ins |
| Frame_Shift_Del |
| Nonsense_Mutation |
| Nonsense_Mutation |
| Nonsense_Mutation |
| Nonsense_Mutation |
| Frame_Shift_Del |
| Frame_Shift_Ins |
| Frame_Shift_Del |

Frame_Shift_Ins
Frame_Shift_Del
Frame_Shift_Del
Frame_Shift_Del
Frame_Shift_Del
Nonsense_Mutation
Frame_Shift_Del
Nonsense_Mutation
Missense_Mutation
Missense_Mutation
Missense_Mutation
Missense_Mutation
Missense_Mutation
Missense_Mutation
Missense_Mutation
Missense_Mutation
Missense_Mutation

***PIK3CA* somatic mutations in HNSC seperated by *KDM3C* genotype**

| Sample ID | Cancer Type | Gene | *KDM3C* genotype | PIK3CA protein change |
|---|---|---|---|---|
| 1 | HNSC | *PIK3CA* | A/A | E545K |
| 2 | HNSC | *PIK3CA* | A/A | R88Q |
| 3 | HNSC | *PIK3CA* | A/A | E542K |
| 4 | HNSC | *PIK3CA* | A/A | E542K |
| 5 | HNSC | *PIK3CA* | A/A | M1043V |
| 6 | HNSC | *PIK3CA* | A/A | E542K |
| 7 | HNSC | *PIK3CA* | A/A | G1007R |
| 8 | HNSC | *PIK3CA* | A/T | E545K |
| 9 | HNSC | *PIK3CA* | A/T | E545K |
| 10 | HNSC | *PIK3CA* | A/T | E545K |
| 11 | HNSC | *PIK3CA* | A/T | E545K |
| 12 | HNSC | *PIK3CA* | A/T | E545K |
| 13 | HNSC | *PIK3CA* | A/T | E545K |
| 14 | HNSC | *PIK3CA* | A/T | E545K |
| 15 | HNSC | *PIK3CA* | A/T | E545K |
| 16 | HNSC | *PIK3CA* | A/T | E545K |
| 17 | HNSC | *PIK3CA* | A/T | E542K |
| 18 | HNSC | *PIK3CA* | A/T | C420R |
| 19 | HNSC | *PIK3CA* | A/T | E542K |
| 20 | HNSC | *PIK3CA* | A/T | E542K |
| 21 | HNSC | *PIK3CA* | A/T | M1043V |
| 22 | HNSC | *PIK3CA* | A/T | E542K |
| 23 | HNSC | *PIK3CA* | A/T | E542K |
| 24 | HNSC | *PIK3CA* | A/T | E542K |
| 25 | HNSC | *PIK3CA* | A/T | K111N |
| 26 | HNSC | *PIK3CA* | A/T | E542K |
| 27 | HNSC | *PIK3CA* | A/T | E542K |
| 28 | HNSC | *PIK3CA* | A/T | H1047R |
| 29 | HNSC | *PIK3CA* | A/T | H1047R |
| 30 | HNSC | *PIK3CA* | A/T | E542K |
| 31 | HNSC | *PIK3CA* | A/T | E542K |
| 32 | HNSC | *PIK3CA* | A/T | H1047R |
| 33 | HNSC | *PIK3CA* | A/T | H1047R |
| 34 | HNSC | *PIK3CA* | A/T | E542K |
| 35 | HNSC | *PIK3CA* | A/T | E542K |
| 36 | HNSC | *PIK3CA* | A/T | Q75E |
| 37 | HNSC | *PIK3CA* | A/T | E110del |
| 38 | HNSC | *PIK3CA* | A/T | N1068Kfs*5 |
| 39 | HNSC | *PIK3CA* | A/T | E81K |
| 40 | HNSC | *PIK3CA* | A/T | G363A |
| 41 | HNSC | *PIK3CA* | A/T | R975S |
| 42 | HNSC | *PIK3CA* | A/T | E726K |
| 43 | HNSC | *PIK3CA* | A/T | E365V |
| 44 | HNSC | *PIK3CA* | A/T | E418K |
| 45 | HNSC | *PIK3CA* | A/T | R335G |
| 46 | HNSC | *PIK3CA* | A/T | M1040I |
| 47 | HNSC | *PIK3CA* | T/T | E545K |
| 48 | HNSC | *PIK3CA* | T/T | E545K |
| 49 | HNSC | *PIK3CA* | T/T | Q546R |
| 50 | HNSC | *PIK3CA* | T/T | E545K |
| 51 | HNSC | *PIK3CA* | T/T | E545K |

| | | | | |
|---|---|---|---|---|
| 52 | HNSC | *PIK3CA* | T/T | E545K |
| 53 | HNSC | *PIK3CA* | T/T | E545K |
| 54 | HNSC | *PIK3CA* | T/T | E545K |
| 55 | HNSC | *PIK3CA* | T/T | E545K |
| 56 | HNSC | *PIK3CA* | T/T | E545K |
| 57 | HNSC | *PIK3CA* | T/T | E545K |
| 58 | HNSC | *PIK3CA* | T/T | Q546R |
| 59 | HNSC | *PIK3CA* | T/T | E545K |
| 60 | HNSC | *PIK3CA* | T/T | E545K |
| 61 | HNSC | *PIK3CA* | T/T | E545K |
| 62 | HNSC | *PIK3CA* | T/T | R88Q |
| 63 | HNSC | *PIK3CA* | T/T | E545K |
| 64 | HNSC | *PIK3CA* | T/T | H1047L |
| 65 | HNSC | *PIK3CA* | T/T | H1047R |
| 66 | HNSC | *PIK3CA* | T/T | E542K |
| 67 | HNSC | *PIK3CA* | T/T | H1047R |
| 68 | HNSC | *PIK3CA* | T/T | H1047R |
| 69 | HNSC | *PIK3CA* | T/T | H1047R |
| 70 | HNSC | *PIK3CA* | T/T | H1047R |
| 71 | HNSC | *PIK3CA* | T/T | H1047R |
| 72 | HNSC | *PIK3CA* | T/T | E453K |
| 73 | HNSC | *PIK3CA* | T/T | C604R |
| 74 | HNSC | *PIK3CA* | T/T | E542K |
| 75 | HNSC | *PIK3CA* | T/T | H1047Q |
| 76 | HNSC | *PIK3CA* | T/T | N345K |
| 77 | HNSC | *PIK3CA* | T/T | H1047R |
| 78 | HNSC | *PIK3CA* | T/T | H1047R |
| 79 | HNSC | *PIK3CA* | T/T | E542K |
| 80 | HNSC | *PIK3CA* | T/T | E542K |
| 81 | HNSC | *PIK3CA* | T/T | V344G |
| 82 | HNSC | *PIK3CA* | T/T | E970K |
| 83 | HNSC | *PIK3CA* | T/T | C901F |
| 84 | HNSC | *PIK3CA* | T/T | G451R |
| 85 | HNSC | *PIK3CA* | T/T | C971R |
| 86 | HNSC | *PIK3CA* | T/T | E726K |
| 87 | HNSC | *PIK3CA* | T/T | R519G |
| 88 | HNSC | *PIK3CA* | T/T | V71I |
| 89 | HNSC | *PIK3CA* | T/T | C905S |

**Summary Table**

| Mutation type | PIK3CA mutations | A/A | A/T | T/T |
|---|---|---|---|---|
| Missense_Mutation | E542K | 3 | 12 | 4 |
| Missense_Mutation | E545K | 1 | 9 | 14 |
| Missense_Mutation | Q546R | 0 | 0 | 2 |
| Missense_Mutation | H1047R | 0 | 4 | 8 |
| Missense_Mutation | H1047L | 0 | 0 | 1 |
| Missense_Mutation | H1047Q | 0 | 0 | 1 |
| Missense_Mutation | | | | |
| Missense_Mutation | | | | |
| Missense_Mutation | | | | |
| Missense_Mutation | | | | |
| Missense_Mutation | | | | |
| Missense_Mutation | | | | |
| Missense_Mutation | | | | |
| Missense_Mutation | | | | |
| Missense_Mutation | | | | |
| Missense_Mutation | | | | |
| Missense_Mutation | | | | |
| Missense_Mutation | | | | |
| Missense_Mutation | | | | |
| Missense_Mutation | | | | |
| Missense_Mutation | | | | |
| Missense_Mutation | | | | |
| Missense_Mutation | | | | |
| Missense_Mutation | | | | |
| Missense_Mutation | | | | |
| Missense_Mutation | | | | |
| Missense_Mutation | | | | |
| Missense_Mutation | | | | |
| Missense_Mutation | | | | |
| Missense_Mutation | | | | |
| Missense_Mutation | | | | |
| Missense_Mutation | | | | |
| Missense_Mutation | | | | |
| Missense_Mutation | | | | |
| Missense_Mutation | | | | |
| Missense_Mutation | | | | |
| In_Frame_Del | | | | |
| Frame_Shift_Ins | | | | |
| Missense_Mutation | | | | |
| Missense_Mutation | | | | |
| Missense_Mutation | | | | |
| Missense_Mutation | | | | |
| Missense_Mutation | | | | |
| Missense_Mutation | | | | |
| Missense_Mutation | | | | |
| Missense_Mutation | | | | |
| Missense_Mutation | | | | |
| Missense_Mutation | | | | |
| Missense_Mutation | | | | |
| Missense_Mutation | | | | |
| Missense_Mutation | | | | |

Missense_Mutation
Missense_Mutation
Missense_Mutation
Missense_Mutation
Missense_Mutation
Missense_Mutation
Missense_Mutation
Missense_Mutation
Missense_Mutation
Missense_Mutation
Missense_Mutation
Missense_Mutation
Missense_Mutation
Missense_Mutation
Missense_Mutation
Missense_Mutation
Missense_Mutation
Missense_Mutation
Missense_Mutation
Missense_Mutation
Missense_Mutation
Missense_Mutation
Missense_Mutation
Missense_Mutation
Missense_Mutation
Missense_Mutation
Missense_Mutation
Missense_Mutation
Missense_Mutation
Missense_Mutation
Missense_Mutation
Missense_Mutation
Missense_Mutation
Missense_Mutation
Missense_Mutation
Missense_Mutation
Missense_Mutation
Missense_Mutation

***TP53* somatic mutations in HNSC seperated by *KDM3C* genotype**

| Sample ID | Cancer Type | Gene | *KDM3C* genotype | TP53 protein change |
|---|---|---|---|---|
| 1 | HNSC | *TP53* | A/A | Y220C |
| 2 | HNSC | *TP53* | A/A | Y220C |
| 3 | HNSC | *TP53* | A/A | R175H |
| 4 | HNSC | *TP53* | A/A | R273H |
| 5 | HNSC | *TP53* | A/A | C176Y |
| 6 | HNSC | *TP53* | A/A | G245D |
| 7 | HNSC | *TP53* | A/A | H179Y |
| 8 | HNSC | *TP53* | A/A | H193L |
| 9 | HNSC | *TP53* | A/A | R282W |
| 10 | HNSC | *TP53* | A/A | R273H |
| 11 | HNSC | *TP53* | A/A | G245S |
| 12 | HNSC | *TP53* | A/A | Y163C |
| 13 | HNSC | *TP53* | A/A | R248W |
| 14 | HNSC | *TP53* | A/A | R175H |
| 15 | HNSC | *TP53* | A/A | R248Q |
| 16 | HNSC | *TP53* | A/A | R248W |
| 17 | HNSC | *TP53* | A/A | N239D |
| 18 | HNSC | *TP53* | A/A | Y126S |
| 19 | HNSC | *TP53* | A/A | R158H |
| 20 | HNSC | *TP53* | A/A | V157F |
| 21 | HNSC | *TP53* | A/A | R213* |
| 22 | HNSC | *TP53* | A/A | I195F |
| 23 | HNSC | *TP53* | A/A | Y205C |
| 24 | HNSC | *TP53* | A/A | Y126C |
| 25 | HNSC | *TP53* | A/A | X32_splice |
| 26 | HNSC | *TP53* | A/A | R202_L206del |
| 27 | HNSC | *TP53* | A/A | R196P |
| 28 | HNSC | *TP53* | A/A | L137Q |
| 29 | HNSC | *TP53* | A/A | D48Gfs*4 |
| 30 | HNSC | *TP53* | A/A | E298* |
| 31 | HNSC | *TP53* | A/A | Q331* |
| 32 | HNSC | *TP53* | A/A | E224D |
| 33 | HNSC | *TP53* | A/A | T256Hfs*8 |
| 34 | HNSC | *TP53* | A/A | S166* |
| 35 | HNSC | *TP53* | A/A | R342Efs*3 |
| 36 | HNSC | *TP53* | A/A | R306* |
| 37 | HNSC | *TP53* | A/A | G266* |
| 38 | HNSC | *TP53* | A/A | R196* |
| 39 | HNSC | *TP53* | A/A | I195Lfs*53 |
| 40 | HNSC | *TP53* | A/A | Q52* |
| 41 | HNSC | *TP53* | A/A | E294Sfs*51 |
| 42 | HNSC | *TP53* | A/A | Q144* |
| 43 | HNSC | *TP53* | A/T | Y220C |
| 44 | HNSC | *TP53* | A/T | L194P |
| 45 | HNSC | *TP53* | A/T | H193L |
| 46 | HNSC | *TP53* | A/T | V272M |
| 47 | HNSC | *TP53* | A/T | R248W |
| 48 | HNSC | *TP53* | A/T | M237I |
| 49 | HNSC | *TP53* | A/T | R337C |
| 50 | HNSC | *TP53* | A/T | V173M |
| 51 | HNSC | *TP53* | A/T | R248W |
| 52 | HNSC | *TP53* | A/T | G245S |

| | | | | |
|---|---|---|---|---|
| 53 | HNSC | *TP53* | A/T | R175H |
| 54 | HNSC | *TP53* | A/T | D281Y |
| 55 | HNSC | *TP53* | A/T | R248Q |
| 56 | HNSC | *TP53* | A/T | H179R |
| 57 | HNSC | *TP53* | A/T | R280S |
| 58 | HNSC | *TP53* | A/T | R273C |
| 59 | HNSC | *TP53* | A/T | R273H |
| 60 | HNSC | *TP53* | A/T | P278S |
| 61 | HNSC | *TP53* | A/T | R282W |
| 62 | HNSC | *TP53* | A/T | C242S |
| 63 | HNSC | *TP53* | A/T | R248W |
| 64 | HNSC | *TP53* | A/T | C275F |
| 65 | HNSC | *TP53* | A/T | H193R |
| 66 | HNSC | *TP53* | A/T | R273H |
| 67 | HNSC | *TP53* | A/T | C242F |
| 68 | HNSC | *TP53* | A/T | R249S |
| 69 | HNSC | *TP53* | A/T | P278S |
| 70 | HNSC | *TP53* | A/T | R273H |
| 71 | HNSC | *TP53* | A/T | C176F |
| 72 | HNSC | *TP53* | A/T | R175H |
| 73 | HNSC | *TP53* | A/T | C275Y |
| 74 | HNSC | *TP53* | A/T | R248Q |
| 75 | HNSC | *TP53* | A/T | R337C |
| 76 | HNSC | *TP53* | A/T | H179R |
| 77 | HNSC | *TP53* | A/T | Y236C |
| 78 | HNSC | *TP53* | A/T | R273C |
| 79 | HNSC | *TP53* | A/T | R249S |
| 80 | HNSC | *TP53* | A/T | R248Q |
| 81 | HNSC | *TP53* | A/T | R175H |
| 82 | HNSC | *TP53* | A/T | R282W |
| 83 | HNSC | *TP53* | A/T | R273C |
| 84 | HNSC | *TP53* | A/T | A159V |
| 85 | HNSC | *TP53* | A/T | S127Y |
| 86 | HNSC | *TP53* | A/T | G266E |
| 87 | HNSC | *TP53* | A/T | G266E |
| 88 | HNSC | *TP53* | A/T | P151H |
| 89 | HNSC | *TP53* | A/T | V157F |
| 90 | HNSC | *TP53* | A/T | R110L |
| 91 | HNSC | *TP53* | A/T | E180K |
| 92 | HNSC | *TP53* | A/T | R283P |
| 93 | HNSC | *TP53* | A/T | R283H |
| 94 | HNSC | *TP53* | A/T | R283P |
| 95 | HNSC | *TP53* | A/T | R213L |
| 96 | HNSC | *TP53* | A/T | R213* |
| 97 | HNSC | *TP53* | A/T | R213* |
| 98 | HNSC | *TP53* | A/T | F134V |
| 99 | HNSC | *TP53* | A/T | I195T |
| 100 | HNSC | *TP53* | A/T | H179N |
| 101 | HNSC | *TP53* | A/T | Q136E |
| 102 | HNSC | *TP53* | A/T | M237V |
| 103 | HNSC | *TP53* | A/T | M133K |
| 104 | HNSC | *TP53* | A/T | C141F |
| 105 | HNSC | *TP53* | A/T | M246I |
| 106 | HNSC | *TP53* | A/T | C277G |
| 107 | HNSC | *TP53* | A/T | Y236D |

| | | | | |
|---|---|---|---|---|
| 108 | HNSC | *TP53* | A/T | E271K |
| 109 | HNSC | *TP53* | A/T | H193Y |
| 110 | HNSC | *TP53* | A/T | I251N |
| 111 | HNSC | *TP53* | A/T | E271K |
| 112 | HNSC | *TP53* | A/T | P177_C182del |
| 113 | HNSC | *TP53* | A/T | Q331= |
| 114 | HNSC | *TP53* | A/T | T125= |
| 115 | HNSC | *TP53* | A/T | V274_C275dup |
| 116 | HNSC | *TP53* | A/T | X32_splice |
| 117 | HNSC | *TP53* | A/T | X331_splice |
| 118 | HNSC | *TP53* | A/T | P151T |
| 119 | HNSC | *TP53* | A/T | X307_splice |
| 120 | HNSC | *TP53* | A/T | X331_splice |
| 121 | HNSC | *TP53* | A/T | A159P |
| 122 | HNSC | *TP53* | A/T | X187_splice |
| 123 | HNSC | *TP53* | A/T | R110C |
| 124 | HNSC | *TP53* | A/T | V173_R175del |
| 125 | HNSC | *TP53* | A/T | X125_splice |
| 126 | HNSC | *TP53* | A/T | V274dup |
| 127 | HNSC | *TP53* | A/T | X33_splice |
| 128 | HNSC | *TP53* | A/T | T125= |
| 129 | HNSC | *TP53* | A/T | X187_splice |
| 130 | HNSC | *TP53* | A/T | X224_splice |
| 131 | HNSC | *TP53* | A/T | X261_splice |
| 132 | HNSC | *TP53* | A/T | T125= |
| 133 | HNSC | *TP53* | A/T | F341L |
| 134 | HNSC | *TP53* | A/T | L201* |
| 135 | HNSC | *TP53* | A/T | Q167* |
| 136 | HNSC | *TP53* | A/T | P219Lfs*2 |
| 137 | HNSC | *TP53* | A/T | Q144* |
| 138 | HNSC | *TP53* | A/T | G199Efs*48 |
| 139 | HNSC | *TP53* | A/T | T329Hfs*8 |
| 140 | HNSC | *TP53* | A/T | D184Afs*62 |
| 141 | HNSC | *TP53* | A/T | Y103Lfs*46 |
| 142 | HNSC | *TP53* | A/T | R209Kfs*6 |
| 143 | HNSC | *TP53* | A/T | R283Afs*62 |
| 144 | HNSC | *TP53* | A/T | E298* |
| 145 | HNSC | *TP53* | A/T | E298* |
| 146 | HNSC | *TP53* | A/T | I195Sfs*52 |
| 147 | HNSC | *TP53* | A/T | E294* |
| 148 | HNSC | *TP53* | A/T | L383Cfs*38 |
| 149 | HNSC | *TP53* | A/T | L32Cfs*12 |
| 150 | HNSC | *TP53* | A/T | C229Yfs*10 |
| 151 | HNSC | *TP53* | A/T | P80Sfs*69 |
| 152 | HNSC | *TP53* | A/T | G154Afs*16 |
| 153 | HNSC | *TP53* | A/T | S37Pfs*7 |
| 154 | HNSC | *TP53* | A/T | S106R |
| 155 | HNSC | *TP53* | A/T | D41Mfs*3 |
| 156 | HNSC | *TP53* | A/T | S90Vfs*55 |
| 157 | HNSC | *TP53* | A/T | E56* |
| 158 | HNSC | *TP53* | A/T | Q331* |
| 159 | HNSC | *TP53* | A/T | G302Efs*3 |
| 160 | HNSC | *TP53* | A/T | V203Gfs*44 |
| 161 | HNSC | *TP53* | A/T | R110Wfs*12 |
| 162 | HNSC | *TP53* | A/T | R306* |

| | | | | |
|---|---|---|---|---|
| 163 | HNSC | *TP53* | A/T | Y103* |
| 164 | HNSC | *TP53* | A/T | Q104* |
| 165 | HNSC | *TP53* | A/T | R306* |
| 166 | HNSC | *TP53* | A/T | T118Qfs*5 |
| 167 | HNSC | *TP53* | A/T | H178Pfs*3 |
| 168 | HNSC | *TP53* | A/T | R342* |
| 169 | HNSC | *TP53* | A/T | W146Vfs*3 |
| 170 | HNSC | *TP53* | A/T | H178Tfs*69 |
| 171 | HNSC | *TP53* | A/T | V197Ifs*42 |
| 172 | HNSC | *TP53* | A/T | S215Kfs*7 |
| 173 | HNSC | *TP53* | A/T | L348S |
| 174 | HNSC | *TP53* | A/T | S260Pfs*85 |
| 175 | HNSC | *TP53* | A/T | R306* |
| 176 | HNSC | *TP53* | A/T | Q192* |
| 177 | HNSC | *TP53* | A/T | C238Lfs*9 |
| 178 | HNSC | *TP53* | A/T | H178Pfs*70 |
| 179 | HNSC | *TP53* | A/T | E204* |
| 180 | HNSC | *TP53* | A/T | E294* |
| 181 | HNSC | *TP53* | A/T | A84Pfs*39 |
| 182 | HNSC | *TP53* | A/T | D281Efs*26 |
| 183 | HNSC | *TP53* | A/T | G245C |
| 184 | HNSC | *TP53* | A/T | V218G |
| 185 | HNSC | *TP53* | A/T | T125M |
| 186 | HNSC | *TP53* | A/T | D259N |
| 187 | HNSC | *TP53* | T/T | Y220C |
| 188 | HNSC | *TP53* | T/T | Y220C |
| 189 | HNSC | *TP53* | T/T | Y220C |
| 190 | HNSC | *TP53* | T/T | G245S |
| 191 | HNSC | *TP53* | T/T | Y236C |
| 192 | HNSC | *TP53* | T/T | R175H |
| 193 | HNSC | *TP53* | T/T | R282W |
| 194 | HNSC | *TP53* | T/T | C238F |
| 195 | HNSC | *TP53* | T/T | C242Y |
| 196 | HNSC | *TP53* | T/T | R273C |
| 197 | HNSC | *TP53* | T/T | V173M |
| 198 | HNSC | *TP53* | T/T | G244V |
| 199 | HNSC | *TP53* | T/T | C176Y |
| 200 | HNSC | *TP53* | T/T | C242F |
| 201 | HNSC | *TP53* | T/T | C275F |
| 202 | HNSC | *TP53* | T/T | H179R |
| 203 | HNSC | *TP53* | T/T | R273H |
| 204 | HNSC | *TP53* | T/T | H179Y |
| 205 | HNSC | *TP53* | T/T | R273H |
| 206 | HNSC | *TP53* | T/T | R273C |
| 207 | HNSC | *TP53* | T/T | R175H |
| 208 | HNSC | *TP53* | T/T | G245V |
| 209 | HNSC | *TP53* | T/T | P278S |
| 210 | HNSC | *TP53* | T/T | G245S |
| 211 | HNSC | *TP53* | T/T | R248W |
| 212 | HNSC | *TP53* | T/T | G245D |
| 213 | HNSC | *TP53* | T/T | C238F |
| 214 | HNSC | *TP53* | T/T | R273C |
| 215 | HNSC | *TP53* | T/T | R282W |
| 216 | HNSC | *TP53* | T/T | R249S |
| 217 | HNSC | *TP53* | T/T | R280T |

| | | | | |
|---|---|---|---|---|
| 218 | HNSC | *TP53* | T/T | C176S |
| 219 | HNSC | *TP53* | T/T | G245V |
| 220 | HNSC | *TP53* | T/T | H179R |
| 221 | HNSC | *TP53* | T/T | R248Q |
| 222 | HNSC | *TP53* | T/T | R175H |
| 223 | HNSC | *TP53* | T/T | R282W |
| 224 | HNSC | *TP53* | T/T | R248W |
| 225 | HNSC | *TP53* | T/T | R337L |
| 226 | HNSC | *TP53* | T/T | Y236C |
| 227 | HNSC | *TP53* | T/T | R248Q |
| 228 | HNSC | *TP53* | T/T | V173L |
| 229 | HNSC | *TP53* | T/T | H193L |
| 230 | HNSC | *TP53* | T/T | Y163C |
| 231 | HNSC | *TP53* | T/T | E285K |
| 232 | HNSC | *TP53* | T/T | R248W |
| 233 | HNSC | *TP53* | T/T | Y234C |
| 234 | HNSC | *TP53* | T/T | V173M |
| 235 | HNSC | *TP53* | T/T | R282W |
| 236 | HNSC | *TP53* | T/T | K132N |
| 237 | HNSC | *TP53* | T/T | R175H |
| 238 | HNSC | *TP53* | T/T | E285K |
| 239 | HNSC | *TP53* | T/T | P278S |
| 240 | HNSC | *TP53* | T/T | Y236C |
| 241 | HNSC | *TP53* | T/T | R280K |
| 242 | HNSC | *TP53* | T/T | R175H |
| 243 | HNSC | *TP53* | T/T | R248Q |
| 244 | HNSC | *TP53* | T/T | R280S |
| 245 | HNSC | *TP53* | T/T | H193L |
| 246 | HNSC | *TP53* | T/T | R337C |
| 247 | HNSC | *TP53* | T/T | Y234C |
| 248 | HNSC | *TP53* | T/T | R273H |
| 249 | HNSC | *TP53* | T/T | V173G |
| 250 | HNSC | *TP53* | T/T | Y163H |
| 251 | HNSC | *TP53* | T/T | Y163C |
| 252 | HNSC | *TP53* | T/T | L194R |
| 253 | HNSC | *TP53* | T/T | H179R |
| 254 | HNSC | *TP53* | T/T | R337C |
| 255 | HNSC | *TP53* | T/T | G245S |
| 256 | HNSC | *TP53* | T/T | E286V |
| 257 | HNSC | *TP53* | T/T | N239D |
| 258 | HNSC | *TP53* | T/T | Y163C |
| 259 | HNSC | *TP53* | T/T | H179Y |
| 260 | HNSC | *TP53* | T/T | G245S |
| 261 | HNSC | *TP53* | T/T | R248Q |
| 262 | HNSC | *TP53* | T/T | S241C |
| 263 | HNSC | *TP53* | T/T | R175H |
| 264 | HNSC | *TP53* | T/T | R282W |
| 265 | HNSC | *TP53* | T/T | R248Q |
| 266 | HNSC | *TP53* | T/T | G245S |
| 267 | HNSC | *TP53* | T/T | G245S |
| 268 | HNSC | *TP53* | T/T | R248Q |
| 269 | HNSC | *TP53* | T/T | H193L |
| 270 | HNSC | *TP53* | T/T | R282W |
| 271 | HNSC | *TP53* | T/T | V272M |
| 272 | HNSC | *TP53* | T/T | R175H |

| | | | | |
|---|---|---|---|---|
| 273 | HNSC | *TP53* | T/T | V157F |
| 274 | HNSC | *TP53* | T/T | A161T |
| 275 | HNSC | *TP53* | T/T | V157F |
| 276 | HNSC | *TP53* | T/T | G266R |
| 277 | HNSC | *TP53* | T/T | P151S |
| 278 | HNSC | *TP53* | T/T | V216M |
| 279 | HNSC | *TP53* | T/T | R267P |
| 280 | HNSC | *TP53* | T/T | R110L |
| 281 | HNSC | *TP53* | T/T | G266E |
| 282 | HNSC | *TP53* | T/T | P151S |
| 283 | HNSC | *TP53* | T/T | R158L |
| 284 | HNSC | *TP53* | T/T | R158L |
| 285 | HNSC | *TP53* | T/T | V157F |
| 286 | HNSC | *TP53* | T/T | R283P |
| 287 | HNSC | *TP53* | T/T | R156P |
| 288 | HNSC | *TP53* | T/T | R213* |
| 289 | HNSC | *TP53* | T/T | R213* |
| 290 | HNSC | *TP53* | T/T | R213* |
| 291 | HNSC | *TP53* | T/T | R213* |
| 292 | HNSC | *TP53* | T/T | R213L |
| 293 | HNSC | *TP53* | T/T | Q136P |
| 294 | HNSC | *TP53* | T/T | R280G |
| 295 | HNSC | *TP53* | T/T | H193P |
| 296 | HNSC | *TP53* | T/T | R249M |
| 297 | HNSC | *TP53* | T/T | R280G |
| 298 | HNSC | *TP53* | T/T | H179L |
| 299 | HNSC | *TP53* | T/T | C238S |
| 300 | HNSC | *TP53* | T/T | E271V |
| 301 | HNSC | *TP53* | T/T | M246I |
| 302 | HNSC | *TP53* | T/T | C141Y |
| 303 | HNSC | *TP53* | T/T | H193P |
| 304 | HNSC | *TP53* | T/T | E258D |
| 305 | HNSC | *TP53* | T/T | X225_splice |
| 306 | HNSC | *TP53* | T/T | V143M |
| 307 | HNSC | *TP53* | T/T | T125= |
| 308 | HNSC | *TP53* | T/T | G105V |
| 309 | HNSC | *TP53* | T/T | X307_splice |
| 310 | HNSC | *TP53* | T/T | X125_splice |
| 311 | HNSC | *TP53* | T/T | H168L |
| 312 | HNSC | *TP53* | T/T | X33_splice |
| 313 | HNSC | *TP53* | T/T | X126_splice |
| 314 | HNSC | *TP53* | T/T | X332_splice |
| 315 | HNSC | *TP53* | T/T | X261_splice |
| 316 | HNSC | *TP53* | T/T | L265R |
| 317 | HNSC | *TP53* | T/T | X225_splice |
| 318 | HNSC | *TP53* | T/T | G105V |
| 319 | HNSC | *TP53* | T/T | X32_splice |
| 320 | HNSC | *TP53* | T/T | X307_splice |
| 321 | HNSC | *TP53* | T/T | E224= |
| 322 | HNSC | *TP53* | T/T | X307_splice |
| 323 | HNSC | *TP53* | T/T | V274_G279del |
| 324 | HNSC | *TP53* | T/T | P177_C182del |
| 325 | HNSC | *TP53* | T/T | F270C |
| 326 | HNSC | *TP53* | T/T | V143M |
| 327 | HNSC | *TP53* | T/T | T155_A161del |

| | | | | |
|---|---|---|---|---|
| 328 | HNSC | *TP53* | T/T | E258A |
| 329 | HNSC | *TP53* | T/T | G105C |
| 330 | HNSC | *TP53* | T/T | S127F |
| 331 | HNSC | *TP53* | T/T | T125= |
| 332 | HNSC | *TP53* | T/T | X224_splice |
| 333 | HNSC | *TP53* | T/T | X224_splice |
| 334 | HNSC | *TP53* | T/T | G262V |
| 335 | HNSC | *TP53* | T/T | X187_splice |
| 336 | HNSC | *TP53* | T/T | X125_splice |
| 337 | HNSC | *TP53* | T/T | X224_splice |
| 338 | HNSC | *TP53* | T/T | X187_splice |
| 339 | HNSC | *TP53* | T/T | X225_splice |
| 340 | HNSC | *TP53* | T/T | A276D |
| 341 | HNSC | *TP53* | T/T | Q331H |
| 342 | HNSC | *TP53* | T/T | Q331H |
| 343 | HNSC | *TP53* | T/T | R196P |
| 344 | HNSC | *TP53* | T/T | E343* |
| 345 | HNSC | *TP53* | T/T | V73Rfs*76 |
| 346 | HNSC | *TP53* | T/T | E224* |
| 347 | HNSC | *TP53* | T/T | X367_splice |
| 348 | HNSC | *TP53* | T/T | R196* |
| 349 | HNSC | *TP53* | T/T | P58Qfs*65 |
| 350 | HNSC | *TP53* | T/T | E171* |
| 351 | HNSC | *TP53* | T/T | Q136* |
| 352 | HNSC | *TP53* | T/T | E294* |
| 353 | HNSC | *TP53* | T/T | L201* |
| 354 | HNSC | *TP53* | T/T | G154Afs*16 |
| 355 | HNSC | *TP53* | T/T | Y205* |
| 356 | HNSC | *TP53* | T/T | A79Rfs*40 |
| 357 | HNSC | *TP53* | T/T | R65Qfs*84 |
| 358 | HNSC | *TP53* | T/T | K139Cfs*6 |
| 359 | HNSC | *TP53* | T/T | E298* |
| 360 | HNSC | *TP53* | T/T | L330Ffs*15 |
| 361 | HNSC | *TP53* | T/T | Q136* |
| 362 | HNSC | *TP53* | T/T | R306* |
| 363 | HNSC | *TP53* | T/T | G266* |
| 364 | HNSC | *TP53* | T/T | H297Pfs*48 |
| 365 | HNSC | *TP53* | T/T | V157Pfs*23 |
| 366 | HNSC | *TP53* | T/T | K101* |
| 367 | HNSC | *TP53* | T/T | R196* |
| 368 | HNSC | *TP53* | T/T | R196* |
| 369 | HNSC | *TP53* | T/T | P85Lfs*38 |
| 370 | HNSC | *TP53* | T/T | T211Ffs*4 |
| 371 | HNSC | *TP53* | T/T | W91* |
| 372 | HNSC | *TP53* | T/T | Q192* |
| 373 | HNSC | *TP53* | T/T | L206Wfs*41 |
| 374 | HNSC | *TP53* | T/T | T211I |
| 375 | HNSC | *TP53* | T/T | S99Rfs*23 |
| 376 | HNSC | *TP53* | T/T | P27Lfs*17 |
| 377 | HNSC | *TP53* | T/T | E171* |
| 378 | HNSC | *TP53* | T/T | P151Rfs*27 |
| 379 | HNSC | *TP53* | T/T | Q38* |
| 380 | HNSC | *TP53* | T/T | R196* |
| 381 | HNSC | *TP53* | T/T | E339* |
| 382 | HNSC | *TP53* | T/T | S240Gfs*20 |

| | | | | |
|---|---|---|---|---|
| 383 | HNSC | *TP53* | T/T | C135Afs*35 |
| 384 | HNSC | *TP53* | T/T | X25_splice |
| 385 | HNSC | *TP53* | T/T | E298* |
| 386 | HNSC | *TP53* | T/T | E298* |
| 387 | HNSC | *TP53* | T/T | R156Hfs*26 |
| 388 | HNSC | *TP53* | T/T | L137Pfs*32 |
| 389 | HNSC | *TP53* | T/T | E286* |
| 390 | HNSC | *TP53* | T/T | P191Qfs*51 |
| 391 | HNSC | *TP53* | T/T | P128Lfs*42 |
| 392 | HNSC | *TP53* | T/T | E68* |
| 393 | HNSC | *TP53* | T/T | P223Afs*3 |
| 394 | HNSC | *TP53* | T/T | K320Nfs*19 |
| 395 | HNSC | *TP53* | T/T | E336* |
| 396 | HNSC | *TP53* | T/T | P58Qfs*65 |
| 397 | HNSC | *TP53* | T/T | T329Hfs*8 |
| 398 | HNSC | *TP53* | T/T | S90Pfs*34 |
| 399 | HNSC | *TP53* | T/T | T256I |
| 400 | HNSC | *TP53* | T/T | E56* |
| 401 | HNSC | *TP53* | T/T | E221* |
| 402 | HNSC | *TP53* | T/T | Y103* |
| 403 | HNSC | *TP53* | T/T | R196* |
| 404 | HNSC | *TP53* | T/T | X367_splice |
| 405 | HNSC | *TP53* | T/T | R196* |
| 406 | HNSC | *TP53* | T/T | H214Qfs*33 |
| 407 | HNSC | *TP53* | T/T | N200Ifs*47 |
| 408 | HNSC | *TP53* | T/T | P250Hfs*13 |
| 409 | HNSC | *TP53* | T/T | Y163* |
| 410 | HNSC | *TP53* | T/T | R306* |
| 411 | HNSC | *TP53* | T/T | Y205* |
| 412 | HNSC | *TP53* | T/T | E285* |
| 413 | HNSC | *TP53* | T/T | Q192* |
| 414 | HNSC | *TP53* | T/T | E68* |
| 415 | HNSC | *TP53* | T/T | G199* |
| 416 | HNSC | *TP53* | T/T | V73Rfs*76 |
| 417 | HNSC | *TP53* | T/T | S106R |
| 418 | HNSC | *TP53* | T/T | Q136H |
| 419 | HNSC | *TP53* | T/T | V218G |
| 420 | HNSC | *TP53* | T/T | N268_R273del |

| Mutation type |
|---|
| Missense_Mutation |
| Missense_Mutation |
| Missense_Mutation |
| Missense_Mutation |
| Missense_Mutation |
| Missense_Mutation |
| Missense_Mutation |
| Missense_Mutation |
| Missense_Mutation |
| Missense_Mutation |
| Missense_Mutation |
| Missense_Mutation |
| Missense_Mutation |
| Missense_Mutation |
| Missense_Mutation |
| Missense_Mutation |
| Missense_Mutation |
| Missense_Mutation |
| Missense_Mutation |
| Missense_Mutation |
| Nonsense_Mutation |
| Missense_Mutation |
| Missense_Mutation |
| Missense_Mutation |
| Splice_Site |
| In_Frame_Del |
| Missense_Mutation |
| Missense_Mutation |
| Frame_Shift_Ins |
| Nonsense_Mutation |
| Nonsense_Mutation |
| Missense_Mutation |
| Frame_Shift_Ins |
| Nonsense_Mutation |
| Frame_Shift_Del |
| Nonsense_Mutation |
| Nonsense_Mutation |
| Nonsense_Mutation |
| Frame_Shift_Ins |
| Nonsense_Mutation |
| Frame_Shift_Del |
| Nonsense_Mutation |
| Missense_Mutation |
| Missense_Mutation |
| Missense_Mutation |
| Missense_Mutation |
| Missense_Mutation |
| Missense_Mutation |
| Missense_Mutation |
| Missense_Mutation |
| Missense_Mutation |
| Missense_Mutation |

**Summary Table**

| **TP53 mutations** | **A/A** | **A/T** | **T/T** |
|---|---|---|---|
| Missense | 26 | 77 | 134 |
| Nonsense | 9 | 19 | 41 |
| Frameshift any | 5 | 30 | 31 |
| Splice/splice-region | 1 | 14 | 24 |
| In-frame indel/deletion | 1 | 4 | 4 |

| **TP53 hotspot mutations** | **A/A** | **A/T** | **T/T** |
|---|---|---|---|
| Y220C | 2 | 1 | 3 |
| R175H | 2 | 3 | 7 |
| R248Q | 1 | 3 | 6 |
| R248W | 2 | 3 | 3 |
| R273H | 2 | 3 | 3 |
| R273C | 0 | 3 | 3 |
| R282W | 1 | 2 | 6 |
| G245S | 1 | 1 | 6 |
| G245D | 1 | 0 | 1 |
| G245V | 0 | 0 | 2 |

Missense_Mutation
Missense_Mutation
Missense_Mutation
Missense_Mutation
Missense_Mutation
Missense_Mutation
Missense_Mutation
Missense_Mutation
Missense_Mutation
Missense_Mutation
Missense_Mutation
Missense_Mutation
Missense_Mutation
Missense_Mutation
Missense_Mutation
Missense_Mutation
Missense_Mutation
Missense_Mutation
Missense_Mutation
Missense_Mutation
Missense_Mutation
Missense_Mutation
Missense_Mutation
Missense_Mutation
Missense_Mutation
Missense_Mutation
Missense_Mutation
Missense_Mutation
Missense_Mutation
Missense_Mutation
Missense_Mutation
Missense_Mutation
Missense_Mutation
Missense_Mutation
Missense_Mutation
Missense_Mutation
Missense_Mutation
Missense_Mutation
Missense_Mutation
Missense_Mutation
Missense_Mutation
Missense_Mutation
Missense_Mutation
Nonsense_Mutation
Nonsense_Mutation
Missense_Mutation
Missense_Mutation
Missense_Mutation
Missense_Mutation
Missense_Mutation
Missense_Mutation
Missense_Mutation
Missense_Mutation
Missense_Mutation
Missense_Mutation

| |
|---|
| Missense_Mutation |
| Missense_Mutation |
| Missense_Mutation |
| Missense_Mutation |
| In_Frame_Del |
| Splice_Region |
| Splice_Region |
| In_Frame_Ins |
| Splice_Site |
| Splice_Site |
| Missense_Mutation |
| Splice_Site |
| Splice_Site |
| Missense_Mutation |
| Splice_Site |
| Missense_Mutation |
| In_Frame_Del |
| Splice_Site |
| In_Frame_Ins |
| Splice_Site |
| Splice_Region |
| Splice_Site |
| Splice_Site |
| Splice_Site |
| Splice_Region |
| Missense_Mutation |
| Nonsense_Mutation |
| Nonsense_Mutation |
| Frame_Shift_Del |
| Nonsense_Mutation |
| Frame_Shift_Del |
| Frame_Shift_Ins |
| Frame_Shift_Del |
| Frame_Shift_Ins |
| Frame_Shift_Del |
| Frame_Shift_Del |
| Nonsense_Mutation |
| Nonsense_Mutation |
| Frame_Shift_Del |
| Nonsense_Mutation |
| Frame_Shift_Del |
| Frame_Shift_Del |
| Frame_Shift_Del |
| Frame_Shift_Ins |
| Frame_Shift_Del |
| Frame_Shift_Del |
| Missense_Mutation |
| Frame_Shift_Del |
| Frame_Shift_Del |
| Nonsense_Mutation |
| Nonsense_Mutation |
| Frame_Shift_Del |
| Frame_Shift_Del |
| Frame_Shift_Del |
| Nonsense_Mutation |

| |
|---|
| Nonsense_Mutation |
| Nonsense_Mutation |
| Nonsense_Mutation |
| Frame_Shift_Del |
| Frame_Shift_Ins |
| Nonsense_Mutation |
| Frame_Shift_Ins |
| Frame_Shift_Del |
| Frame_Shift_Del |
| Frame_Shift_Ins |
| Missense_Mutation |
| Frame_Shift_Del |
| Nonsense_Mutation |
| Nonsense_Mutation |
| Frame_Shift_Del |
| Frame_Shift_Ins |
| Nonsense_Mutation |
| Nonsense_Mutation |
| Frame_Shift_Del |
| Frame_Shift_Ins |
| Missense_Mutation |
| Missense_Mutation |
| Missense_Mutation |
| Missense_Mutation |
| Missense_Mutation |
| Missense_Mutation |
| Missense_Mutation |
| Missense_Mutation |
| Missense_Mutation |
| Missense_Mutation |
| Missense_Mutation |
| Missense_Mutation |
| Missense_Mutation |
| Missense_Mutation |
| Missense_Mutation |
| Missense_Mutation |
| Missense_Mutation |
| Missense_Mutation |
| Missense_Mutation |
| Missense_Mutation |
| Missense_Mutation |
| Missense_Mutation |
| Missense_Mutation |
| Missense_Mutation |
| Missense_Mutation |
| Missense_Mutation |
| Missense_Mutation |
| Missense_Mutation |
| Missense_Mutation |
| Missense_Mutation |
| Missense_Mutation |
| Missense_Mutation |
| Missense_Mutation |
| Missense_Mutation |
| Missense_Mutation |

Missense_Mutation
Missense_Mutation
Missense_Mutation
Missense_Mutation
Missense_Mutation
Missense_Mutation
Missense_Mutation
Missense_Mutation
Missense_Mutation
Missense_Mutation
Missense_Mutation
Missense_Mutation
Missense_Mutation
Missense_Mutation
Missense_Mutation
Missense_Mutation
Missense_Mutation
Missense_Mutation
Missense_Mutation
Missense_Mutation
Missense_Mutation
Missense_Mutation
Missense_Mutation
Missense_Mutation
Missense_Mutation
Missense_Mutation
Missense_Mutation
Missense_Mutation
Missense_Mutation
Missense_Mutation
Missense_Mutation
Missense_Mutation
Missense_Mutation
Missense_Mutation
Missense_Mutation
Missense_Mutation
Missense_Mutation
Missense_Mutation
Missense_Mutation
Missense_Mutation
Missense_Mutation
Missense_Mutation
Missense_Mutation
Missense_Mutation
Missense_Mutation
Missense_Mutation
Missense_Mutation
Missense_Mutation
Missense_Mutation
Missense_Mutation
Missense_Mutation
Missense_Mutation
Missense_Mutation
Missense_Mutation
Missense_Mutation

| |
|---|
| Missense_Mutation |
| Missense_Mutation |
| Missense_Mutation |
| Missense_Mutation |
| Missense_Mutation |
| Missense_Mutation |
| Missense_Mutation |
| Missense_Mutation |
| Missense_Mutation |
| Missense_Mutation |
| Missense_Mutation |
| Missense_Mutation |
| Missense_Mutation |
| Missense_Mutation |
| Missense_Mutation |
| Nonsense_Mutation |
| Nonsense_Mutation |
| Nonsense_Mutation |
| Nonsense_Mutation |
| Missense_Mutation |
| Missense_Mutation |
| Missense_Mutation |
| Missense_Mutation |
| Missense_Mutation |
| Missense_Mutation |
| Missense_Mutation |
| Missense_Mutation |
| Missense_Mutation |
| Missense_Mutation |
| Missense_Mutation |
| Missense_Mutation |
| Missense_Mutation |
| Splice_Site |
| Missense_Mutation |
| Splice_Region |
| Missense_Mutation |
| Splice_Site |
| Splice_Site |
| Missense_Mutation |
| Splice_Site |
| Splice_Site |
| Splice_Site |
| Splice_Site |
| Missense_Mutation |
| Splice_Site |
| Missense_Mutation |
| Splice_Site |
| Splice_Site |
| Splice_Region |
| Splice_Site |
| In_Frame_Del |
| In_Frame_Del |
| Missense_Mutation |
| Missense_Mutation |
| In_Frame_Del |

| |
|---|
| Missense_Mutation |
| Missense_Mutation |
| Missense_Mutation |
| Splice_Region |
| Splice_Site |
| Splice_Site |
| Missense_Mutation |
| Splice_Site |
| Splice_Site |
| Splice_Site |
| Splice_Site |
| Splice_Site |
| Missense_Mutation |
| Missense_Mutation |
| Missense_Mutation |
| Missense_Mutation |
| Nonsense_Mutation |
| Frame_Shift_Ins |
| Frame_Shift_Ins |
| Splice_Site |
| Nonsense_Mutation |
| Frame_Shift_Del |
| Nonsense_Mutation |
| Nonsense_Mutation |
| Nonsense_Mutation |
| Nonsense_Mutation |
| Frame_Shift_Del |
| Nonsense_Mutation |
| Frame_Shift_Del |
| Frame_Shift_Ins |
| Frame_Shift_Del |
| Nonsense_Mutation |
| Frame_Shift_Del |
| Nonsense_Mutation |
| Nonsense_Mutation |
| Nonsense_Mutation |
| Frame_Shift_Del |
| Frame_Shift_Del |
| Nonsense_Mutation |
| Nonsense_Mutation |
| Nonsense_Mutation |
| Frame_Shift_Del |
| Frame_Shift_Del |
| Nonsense_Mutation |
| Nonsense_Mutation |
| Frame_Shift_Del |
| Missense_Mutation |
| Frame_Shift_Del |
| Frame_Shift_Del |
| Nonsense_Mutation |
| Frame_Shift_Del |
| Nonsense_Mutation |
| Nonsense_Mutation |
| Nonsense_Mutation |
| Frame_Shift_Del |

| |
|---|
| Frame_Shift_Del |
| Splice_Site |
| Nonsense_Mutation |
| Nonsense_Mutation |
| Frame_Shift_Ins |
| Frame_Shift_Del |
| Nonsense_Mutation |
| Frame_Shift_Del |
| Frame_Shift_Del |
| Nonsense_Mutation |
| Frame_Shift_Ins |
| Frame_Shift_Del |
| Nonsense_Mutation |
| Frame_Shift_Del |
| Frame_Shift_Ins |
| Frame_Shift_Ins |
| Missense_Mutation |
| Nonsense_Mutation |
| Nonsense_Mutation |
| Nonsense_Mutation |
| Nonsense_Mutation |
| Splice_Site |
| Nonsense_Mutation |
| Frame_Shift_Del |
| Frame_Shift_Del |
| Frame_Shift_Del |
| Nonsense_Mutation |
| Nonsense_Mutation |
| Nonsense_Mutation |
| Nonsense_Mutation |
| Nonsense_Mutation |
| Nonsense_Mutation |
| Nonsense_Mutation |
| Frame_Shift_Ins |
| Missense_Mutation |
| Missense_Mutation |
| Missense_Mutation |
| In_Frame_Del |

***CDKN2A* somatic mutations in HNSC seperated by *KDM3C* genotype**

| Sample ID | Cancer Type | Gene | *KDM3C* genotype | CDKN2A protein change |
|---|---|---|---|---|
| 1 | HNSC | *CDKN2A* | A/A | Y129* |
| 2 | HNSC | *CDKN2A* | A/A | X153_splice |
| 3 | HNSC | *CDKN2A* | A/A | R58* |
| 4 | HNSC | *CDKN2A* | A/A | P114L |
| 5 | HNSC | *CDKN2A* | A/A | E88* |
| 6 | HNSC | *CDKN2A* | A/A | E88* |
| 7 | HNSC | *CDKN2A* | A/A | R80* |
| 8 | HNSC | *CDKN2A* | A/T | W110* |
| 9 | HNSC | *CDKN2A* | A/T | W110* |
| 10 | HNSC | *CDKN2A* | A/T | W110* |
| 11 | HNSC | *CDKN2A* | A/T | E69* |
| 12 | HNSC | *CDKN2A* | A/T | H83R |
| 13 | HNSC | *CDKN2A* | A/T | R80* |
| 14 | HNSC | *CDKN2A* | A/T | E88* |
| 15 | HNSC | *CDKN2A* | A/T | R99Gfs*47 |
| 16 | HNSC | *CDKN2A* | A/T | Y44* |
| 17 | HNSC | *CDKN2A* | A/T | R58* |
| 18 | HNSC | *CDKN2A* | A/T | R58* |
| 19 | HNSC | *CDKN2A* | A/T | E88* |
| 20 | HNSC | *CDKN2A* | A/T | R80* |
| 21 | HNSC | *CDKN2A* | A/T | R58* |
| 22 | HNSC | *CDKN2A* | A/T | X153_splice |
| 23 | HNSC | *CDKN2A* | A/T | R80* |
| 24 | HNSC | *CDKN2A* | A/T | R80* |
| 25 | HNSC | *CDKN2A* | A/T | A36Gfs*8 |
| 26 | HNSC | *CDKN2A* | A/T | D156Ifs*37 |
| 27 | HNSC | *CDKN2A* | A/T | R58* |
| 28 | HNSC | *CDKN2A* | A/T | R80* |
| 29 | HNSC | *CDKN2A* | A/T | G35W |
| 30 | HNSC | *CDKN2A* | A/T | R80* |
| 31 | HNSC | *CDKN2A* | A/T | X153_splice |
| 32 | HNSC | *CDKN2A* | A/T | X51_splice |
| 33 | HNSC | *CDKN2A* | A/T | R80* |
| 34 | HNSC | *CDKN2A* | A/T | W15Pfs*22 |
| 35 | HNSC | *CDKN2A* | A/T | T77Ifs*69 |
| 36 | HNSC | *CDKN2A* | A/T | R29_A34del |
| 37 | HNSC | *CDKN2A* | A/T | Y44Lfs*76 |
| 38 | HNSC | *CDKN2A* | A/T | X153_splice |
| 39 | HNSC | *CDKN2A* | A/T | A68P |
| 40 | HNSC | *CDKN2A* | A/T | E10* |
| 41 | HNSC | *CDKN2A* | A/T | X153_splice |
| 42 | HNSC | *CDKN2A* | A/T | L117Rfs*29 |
| 43 | HNSC | *CDKN2A* | A/T | Y44C |
| 44 | HNSC | *CDKN2A* | A/T | F90L |
| 45 | HNSC | *CDKN2A* | T/T | R87P |
| 46 | HNSC | *CDKN2A* | T/T | D108Y |
| 47 | HNSC | *CDKN2A* | T/T | W110* |
| 48 | HNSC | *CDKN2A* | T/T | W110* |
| 49 | HNSC | *CDKN2A* | T/T | W110* |
| 50 | HNSC | *CDKN2A* | T/T | P48L |
| 51 | HNSC | *CDKN2A* | T/T | W110* |
| 52 | HNSC | *CDKN2A* | T/T | W110* |

| | | | | |
|---|---|---|---|---|
| 53 | HNSC | *CDKN2A* | T/T | W110* |
| 54 | HNSC | *CDKN2A* | T/T | D84G |
| 55 | HNSC | *CDKN2A* | T/T | D108Y |
| 56 | HNSC | *CDKN2A* | T/T | D74N |
| 57 | HNSC | *CDKN2A* | T/T | E69* |
| 58 | HNSC | *CDKN2A* | T/T | D156Ifs*37 |
| 59 | HNSC | *CDKN2A* | T/T | X153_splice |
| 60 | HNSC | *CDKN2A* | T/T | R80* |
| 61 | HNSC | *CDKN2A* | T/T | Q50R |
| 62 | HNSC | *CDKN2A* | T/T | R58* |
| 63 | HNSC | *CDKN2A* | T/T | R58* |
| 64 | HNSC | *CDKN2A* | T/T | X153_splice |
| 65 | HNSC | *CDKN2A* | T/T | E120* |
| 66 | HNSC | *CDKN2A* | T/T | R80* |
| 67 | HNSC | *CDKN2A* | T/T | G45Vfs*8 |
| 68 | HNSC | *CDKN2A* | T/T | R58* |
| 69 | HNSC | *CDKN2A* | T/T | R80* |
| 70 | HNSC | *CDKN2A* | T/T | D74V |
| 71 | HNSC | *CDKN2A* | T/T | X51_splice |
| 72 | HNSC | *CDKN2A* | T/T | R80* |
| 73 | HNSC | *CDKN2A* | T/T | A60Rfs*86 |
| 74 | HNSC | *CDKN2A* | T/T | R80* |
| 75 | HNSC | *CDKN2A* | T/T | N42H |
| 76 | HNSC | *CDKN2A* | T/T | A102Rfs*14 |
| 77 | HNSC | *CDKN2A* | T/T | R80* |
| 78 | HNSC | *CDKN2A* | T/T | D108V |
| 79 | HNSC | *CDKN2A* | T/T | X153_splice |
| 80 | HNSC | *CDKN2A* | T/T | R80* |
| 81 | HNSC | *CDKN2A* | T/T | R80* |
| 82 | HNSC | *CDKN2A* | T/T | R58* |
| 83 | HNSC | *CDKN2A* | T/T | A147Pfs*46 |
| 84 | HNSC | *CDKN2A* | T/T | R80* |
| 85 | HNSC | *CDKN2A* | T/T | A76Rfs*44 |
| 86 | HNSC | *CDKN2A* | T/T | R58* |
| 87 | HNSC | *CDKN2A* | T/T | R22Gfs*4 |
| 88 | HNSC | *CDKN2A* | T/T | R80* |
| 89 | HNSC | *CDKN2A* | T/T | L78Hfs*41 |
| 90 | HNSC | *CDKN2A* | T/T | R58* |
| 91 | HNSC | *CDKN2A* | T/T | R112H |
| 92 | HNSC | *CDKN2A* | T/T | E88Rfs*58 |
| 93 | HNSC | *CDKN2A* | T/T | R80* |
| 94 | HNSC | *CDKN2A* | T/T | R80* |
| 95 | HNSC | *CDKN2A* | T/T | E120* |
| 96 | HNSC | *CDKN2A* | T/T | V51Sfs*2 |
| 97 | HNSC | *CDKN2A* | T/T | M53Ifs*2 |
| 98 | HNSC | *CDKN2A* | T/T | R80* |
| 99 | HNSC | *CDKN2A* | T/T | R58* |
| 100 | HNSC | *CDKN2A* | T/T | R80* |
| 101 | HNSC | *CDKN2A* | T/T | A68Rfs*78 |
| 102 | HNSC | *CDKN2A* | T/T | R58* |
| 103 | HNSC | *CDKN2A* | T/T | R80* |
| 104 | HNSC | *CDKN2A* | T/T | R80* |
| 105 | HNSC | *CDKN2A* | T/T | T18_A21del |
| 106 | HNSC | *CDKN2A* | T/T | A100_L104del |
| 107 | HNSC | *CDKN2A* | T/T | M54del |

| | | | | |
|---|---|---|---|---|
| 108 | HNSC | *CDKN2A* | T/T | P135S |

| Mutation type |
|---|
| Nonsense_Mutation |
| Splice_Site |
| Nonsense_Mutation |
| Missense_Mutation |
| Nonsense_Mutation |
| Nonsense_Mutation |
| Nonsense_Mutation |
| Nonsense_Mutation |
| Nonsense_Mutation |
| Nonsense_Mutation |
| Nonsense_Mutation |
| Missense_Mutation |
| Nonsense_Mutation |
| Nonsense_Mutation |
| Frame_Shift_Del |
| Nonsense_Mutation |
| Nonsense_Mutation |
| Nonsense_Mutation |
| Nonsense_Mutation |
| Nonsense_Mutation |
| Nonsense_Mutation |
| Splice_Site |
| Nonsense_Mutation |
| Nonsense_Mutation |
| Frame_Shift_Ins |
| Frame_Shift_Del |
| Nonsense_Mutation |
| Nonsense_Mutation |
| Missense_Mutation |
| Nonsense_Mutation |
| Splice_Site |
| Splice_Site |
| Nonsense_Mutation |
| Frame_Shift_Del |
| Frame_Shift_Del |
| In_Frame_Del |
| Frame_Shift_Ins |
| Splice_Site |
| Missense_Mutation |
| Nonsense_Mutation |
| Splice_Site |
| Frame_Shift_Del |
| Missense_Mutation |
| Missense_Mutation |
| Missense_Mutation |
| Missense_Mutation |
| Nonsense_Mutation |
| Nonsense_Mutation |
| Nonsense_Mutation |
| Missense_Mutation |
| Nonsense_Mutation |
| Nonsense_Mutation |

**Summary Table**

| CDKN2A mutations | A/A | A/T | T/T |
|---|---|---|---|
| Missense | 1 | 5 | 12 |
| Nonsense | 5 | 19 | 33 |
| Frameshift any | 0 | 7 | 12 |
| Splice | 1 | 5 | 4 |
| In-frame deletion | 0 | 1 | 3 |

| CDKN2A recurrent mutations | A/A | A/T | T/T |
|---|---|---|---|
| R80* | 1 | 7 | 16 |
| R58* | 1 | 4 | 8 |
| W110* | 0 | 3 | 6 |
| E88* / E88fs | 2 | 2 | 1 |
| X153_splice | 1 | 4 | 3 |
| E120* | 0 | 0 | 2 |
| D108Y/V | 0 | 0 | 3 |

Nonsense_Mutation
Missense_Mutation
Missense_Mutation
Missense_Mutation
Nonsense_Mutation
Frame_Shift_Del
Splice_Site
Nonsense_Mutation
Missense_Mutation
Nonsense_Mutation
Nonsense_Mutation
Splice_Site
Nonsense_Mutation
Nonsense_Mutation
Frame_Shift_Del
Nonsense_Mutation
Nonsense_Mutation
Missense_Mutation
Splice_Site
Nonsense_Mutation
Frame_Shift_Del
Nonsense_Mutation
Missense_Mutation
Frame_Shift_Del
Nonsense_Mutation
Missense_Mutation
Splice_Site
Nonsense_Mutation
Nonsense_Mutation
Nonsense_Mutation
Frame_Shift_Del
Nonsense_Mutation
Frame_Shift_Ins
Nonsense_Mutation
Frame_Shift_Del
Nonsense_Mutation
Frame_Shift_Del
Nonsense_Mutation
Missense_Mutation
Frame_Shift_Del
Nonsense_Mutation
Nonsense_Mutation
Nonsense_Mutation
Frame_Shift_Del
Frame_Shift_Ins
Nonsense_Mutation
Nonsense_Mutation
Nonsense_Mutation
Frame_Shift_Del
Nonsense_Mutation
Nonsense_Mutation
Nonsense_Mutation
In_Frame_Del
In_Frame_Del
In_Frame_Del

Missense_Mutation

***FAT1* somatic mutations in HNSC seperated by *KDM3C* genotype**

| Sample ID | Cancer Type | Gene | *KDM3C* genotype | FAT1 protein change |
|---|---|---|---|---|
| 1 | HNSC | *FAT1* | A/A | R1795* |
| 2 | HNSC | *FAT1* | A/A | S2838* |
| 3 | HNSC | *FAT1* | A/A | R1262* |
| 4 | HNSC | *FAT1* | A/A | V3229Cfs*3 |
| 5 | HNSC | *FAT1* | A/A | E1602* |
| 6 | HNSC | *FAT1* | A/A | Q2321* |
| 7 | HNSC | *FAT1* | A/A | Q725* |
| 8 | HNSC | *FAT1* | A/A | Q2830* |
| 9 | HNSC | *FAT1* | A/A | G2758Wfs*32 |
| 10 | HNSC | *FAT1* | A/A | R3087Pfs*2 |
| 11 | HNSC | *FAT1* | A/T | R937* |
| 12 | HNSC | *FAT1* | A/T | P4007Ifs*45 |
| 13 | HNSC | *FAT1* | A/T | S2928Efs*2 |
| 14 | HNSC | *FAT1* | A/T | N2075Mfs*32 |
| 15 | HNSC | *FAT1* | A/T | E160* |
| 16 | HNSC | *FAT1* | A/T | X3285_splice |
| 17 | HNSC | *FAT1* | A/T | F2333Sfs*24 |
| 18 | HNSC | *FAT1* | A/T | R3872* |
| 19 | HNSC | *FAT1* | A/T | Q1114* |
| 20 | HNSC | *FAT1* | A/T | L777Ffs*5 |
| 21 | HNSC | *FAT1* | A/T | Q1527* |
| 22 | HNSC | *FAT1* | A/T | G3960* |
| 23 | HNSC | *FAT1* | A/T | E4274* |
| 24 | HNSC | *FAT1* | A/T | X3077_splice |
| 25 | HNSC | *FAT1* | A/T | E3024* |
| 26 | HNSC | *FAT1* | A/T | Y3610* |
| 27 | HNSC | *FAT1* | A/T | P2007Vfs*12 |
| 28 | HNSC | *FAT1* | A/T | Q2775* |
| 29 | HNSC | *FAT1* | A/T | Y3559* |
| 30 | HNSC | *FAT1* | A/T | R191* |
| 31 | HNSC | *FAT1* | A/T | R628* |
| 32 | HNSC | *FAT1* | A/T | G2299* |
| 33 | HNSC | *FAT1* | A/T | Y2529* |
| 34 | HNSC | *FAT1* | A/T | R97Ifs*9 |
| 35 | HNSC | *FAT1* | A/T | S1669* |
| 36 | HNSC | *FAT1* | A/T | Q1494* |
| 37 | HNSC | *FAT1* | A/T | S1200* |
| 38 | HNSC | *FAT1* | A/T | S3404* |
| 39 | HNSC | *FAT1* | A/T | E1430* |
| 40 | HNSC | *FAT1* | A/T | V1689Gfs*14 |
| 41 | HNSC | *FAT1* | A/T | T3332Nfs*15 |
| 42 | HNSC | *FAT1* | A/T | T1364Yfs*8 |
| 43 | HNSC | *FAT1* | A/T | E2929* |
| 44 | HNSC | *FAT1* | A/T | E3558* |
| 45 | HNSC | *FAT1* | A/T | E1839* |
| 46 | HNSC | *FAT1* | A/T | R548Tfs*16 |
| 47 | HNSC | *FAT1* | A/T | Q1694* |
| 48 | HNSC | *FAT1* | A/T | R3400* |
| 49 | HNSC | *FAT1* | A/T | R937* |
| 50 | HNSC | *FAT1* | A/T | F2283Cfs*11 |
| 51 | HNSC | *FAT1* | A/T | S3353F |

| | | | | |
|---|---|---|---|---|
| 52 | HNSC | *FAT1* | A/T | P2180L |
| 53 | HNSC | *FAT1* | A/T | V2204G |
| 54 | HNSC | *FAT1* | A/T | F2186V |
| 55 | HNSC | *FAT1* | A/T | P2594del |
| 56 | HNSC | *FAT1* | A/T | L2994R |
| 57 | HNSC | *FAT1* | A/T | P147L |
| 58 | HNSC | *FAT1* | A/T | G3155R |
| 59 | HNSC | *FAT1* | A/T | G2626D |
| 60 | HNSC | *FAT1* | A/T | H2435Y |
| 61 | HNSC | *FAT1* | A/T | I2422K |
| 62 | HNSC | *FAT1* | A/T | I3042M |
| 63 | HNSC | *FAT1* | A/T | I661R |
| 64 | HNSC | *FAT1* | T/T | K1445Qfs*7 |
| 65 | HNSC | *FAT1* | T/T | K751Sfs*14 |
| 66 | HNSC | *FAT1* | T/T | E1344* |
| 67 | HNSC | *FAT1* | T/T | T1555Rfs*30 |
| 68 | HNSC | *FAT1* | T/T | K1225Qfs*13 |
| 69 | HNSC | *FAT1* | T/T | Y468Sfs*4 |
| 70 | HNSC | *FAT1* | T/T | I1343Yfs*2 |
| 71 | HNSC | *FAT1* | T/T | F3230Sfs*39 |
| 72 | HNSC | *FAT1* | T/T | N104Ifs*9 |
| 73 | HNSC | *FAT1* | T/T | R132* |
| 74 | HNSC | *FAT1* | T/T | S3373* |
| 75 | HNSC | *FAT1* | T/T | S3763Nfs*7 |
| 76 | HNSC | *FAT1* | T/T | T944Nfs*12 |
| 77 | HNSC | *FAT1* | T/T | Q1694* |
| 78 | HNSC | *FAT1* | T/T | R3400* |
| 79 | HNSC | *FAT1* | T/T | V623Ifs*52 |
| 80 | HNSC | *FAT1* | T/T | Q600* |
| 81 | HNSC | *FAT1* | T/T | S2838* |
| 82 | HNSC | *FAT1* | T/T | S3268* |
| 83 | HNSC | *FAT1* | T/T | N1330Kfs*38 |
| 84 | HNSC | *FAT1* | T/T | G3740* |
| 85 | HNSC | *FAT1* | T/T | D866Tfs*8 |
| 86 | HNSC | *FAT1* | T/T | X3784_splice |
| 87 | HNSC | *FAT1* | T/T | Q4023* |
| 88 | HNSC | *FAT1* | T/T | E1994* |
| 89 | HNSC | *FAT1* | T/T | R1070* |
| 90 | HNSC | *FAT1* | T/T | Q3524* |
| 91 | HNSC | *FAT1* | T/T | S3560* |
| 92 | HNSC | *FAT1* | T/T | E4290* |
| 93 | HNSC | *FAT1* | T/T | S625* |
| 94 | HNSC | *FAT1* | T/T | S1336* |
| 95 | HNSC | *FAT1* | T/T | Q1477* |
| 96 | HNSC | *FAT1* | T/T | S2826Kfs*2 |
| 97 | HNSC | *FAT1* | T/T | Q1494Afs*4 |
| 98 | HNSC | *FAT1* | T/T | R885* |
| 99 | HNSC | *FAT1* | T/T | S377Kfs*2 |
| 100 | HNSC | *FAT1* | T/T | E685* |
| 101 | HNSC | *FAT1* | T/T | S3373* |
| 102 | HNSC | *FAT1* | T/T | Q958* |
| 103 | HNSC | *FAT1* | T/T | R1627* |
| 104 | HNSC | *FAT1* | T/T | T3083Pfs*33 |
| 105 | HNSC | *FAT1* | T/T | R885* |
| 106 | HNSC | *FAT1* | T/T | S3373* |

| | | | | |
|---|---|---|---|---|
| 107 | HNSC | *FAT1* | T/T | S3004* |
| 108 | HNSC | *FAT1* | T/T | E38* |
| 109 | HNSC | *FAT1* | T/T | Q3902* |
| 110 | HNSC | *FAT1* | T/T | R1627* |
| 111 | HNSC | *FAT1* | T/T | L1883Ffs*3 |
| 112 | HNSC | *FAT1* | T/T | A274Cfs*7 |
| 113 | HNSC | *FAT1* | T/T | E3391* |
| 114 | HNSC | *FAT1* | T/T | S1200* |
| 115 | HNSC | *FAT1* | T/T | F2438Lfs*18 |
| 116 | HNSC | *FAT1* | T/T | Y2395Mfs*15 |
| 117 | HNSC | *FAT1* | T/T | X3881_splice |
| 118 | HNSC | *FAT1* | T/T | Q1848Hfs*11 |
| 119 | HNSC | *FAT1* | T/T | E28* |
| 120 | HNSC | *FAT1* | T/T | S2826Kfs*2 |
| 121 | HNSC | *FAT1* | T/T | A173P |
| 122 | HNSC | *FAT1* | T/T | T3031I |
| 123 | HNSC | *FAT1* | T/T | P925R |
| 124 | HNSC | *FAT1* | T/T | S291C |
| 125 | HNSC | *FAT1* | T/T | M530L |
| 126 | HNSC | *FAT1* | T/T | Y3876C |
| 127 | HNSC | *FAT1* | T/T | E4283K |
| 128 | HNSC | *FAT1* | T/T | C4136W |
| 129 | HNSC | *FAT1* | T/T | D3399Y |
| 130 | HNSC | *FAT1* | T/T | P1454R |
| 131 | HNSC | *FAT1* | T/T | T590S |
| 132 | HNSC | *FAT1* | T/T | D175H |
| 133 | HNSC | *FAT1* | T/T | F4371L |
| 134 | HNSC | *FAT1* | T/T | S4367C |
| 135 | HNSC | *FAT1* | T/T | Y2427C |
| 136 | HNSC | *FAT1* | T/T | S520N |
| 137 | HNSC | *FAT1* | T/T | D3693G |
| 138 | HNSC | *FAT1* | T/T | R3858_S3863del |

| Mutation type |
|---|
| Nonsense_Mutation |
| Nonsense_Mutation |
| Nonsense_Mutation |
| Frame_Shift_Ins |
| Nonsense_Mutation |
| Nonsense_Mutation |
| Nonsense_Mutation |
| Nonsense_Mutation |
| Frame_Shift_Del |
| Frame_Shift_Ins |
| Nonsense_Mutation |
| Frame_Shift_Ins |
| Frame_Shift_Ins |
| Frame_Shift_Del |
| Nonsense_Mutation |
| Splice_Site |
| Frame_Shift_Del |
| Nonsense_Mutation |
| Nonsense_Mutation |
| Frame_Shift_Ins |
| Nonsense_Mutation |
| Nonsense_Mutation |
| Nonsense_Mutation |
| Splice_Site |
| Nonsense_Mutation |
| Nonsense_Mutation |
| Frame_Shift_Del |
| Nonsense_Mutation |
| Nonsense_Mutation |
| Nonsense_Mutation |
| Nonsense_Mutation |
| Nonsense_Mutation |
| Nonsense_Mutation |
| Frame_Shift_Del |
| Nonsense_Mutation |
| Nonsense_Mutation |
| Nonsense_Mutation |
| Nonsense_Mutation |
| Nonsense_Mutation |
| Frame_Shift_Ins |
| Frame_Shift_Ins |
| Frame_Shift_Ins |
| Nonsense_Mutation |
| Nonsense_Mutation |
| Nonsense_Mutation |
| Frame_Shift_Ins |
| Nonsense_Mutation |
| Nonsense_Mutation |
| Nonsense_Mutation |
| Frame_Shift_Del |
| Missense_Mutation |

**Summary Table**

| FAT1 mutations | A/A | A/T | T/T |
|---|---|---|---|
| Missense | 0 | 12 | 17 |
| Nonsense | 7 | 26 | 31 |
| Frameshift any | 3 | 12 | 24 |
| Splice | 0 | 2 | 2 |
| In-frame deletion | 0 | 1 | 1 |

| FAT1 recurrent mutations | A/A | A/T | T/T |
|---|---|---|---|
| R937* | 0 | 2 | 0 |
| S3373* | 0 | 0 | 3 |
| R885* | 0 | 0 | 2 |
| R1627* | 0 | 0 | 2 |
| S2826Kfs*2 | 0 | 0 | 2 |

| |
|---|
| Missense_Mutation |
| Missense_Mutation |
| Missense_Mutation |
| In_Frame_Del |
| Missense_Mutation |
| Missense_Mutation |
| Missense_Mutation |
| Missense_Mutation |
| Missense_Mutation |
| Missense_Mutation |
| Missense_Mutation |
| Missense_Mutation |
| Frame_Shift_Ins |
| Frame_Shift_Del |
| Frame_Shift_Ins |
| Frame_Shift_Del |
| Frame_Shift_Ins |
| Frame_Shift_Del |
| Frame_Shift_Ins |
| Frame_Shift_Del |
| Frame_Shift_Del |
| Nonsense_Mutation |
| Nonsense_Mutation |
| Frame_Shift_Ins |
| Frame_Shift_Ins |
| Nonsense_Mutation |
| Nonsense_Mutation |
| Frame_Shift_Del |
| Nonsense_Mutation |
| Nonsense_Mutation |
| Nonsense_Mutation |
| Frame_Shift_Del |
| Nonsense_Mutation |
| Frame_Shift_Del |
| Splice_Site |
| Nonsense_Mutation |
| Nonsense_Mutation |
| Nonsense_Mutation |
| Nonsense_Mutation |
| Nonsense_Mutation |
| Nonsense_Mutation |
| Nonsense_Mutation |
| Nonsense_Mutation |
| Nonsense_Mutation |
| Frame_Shift_Ins |
| Frame_Shift_Ins |
| Nonsense_Mutation |
| Frame_Shift_Ins |
| Nonsense_Mutation |
| Nonsense_Mutation |
| Nonsense_Mutation |
| Nonsense_Mutation |
| Frame_Shift_Del |
| Nonsense_Mutation |
| Nonsense_Mutation |

| |
|---|
| Nonsense_Mutation |
| Nonsense_Mutation |
| Nonsense_Mutation |
| Nonsense_Mutation |
| Frame_Shift_Ins |
| Frame_Shift_Ins |
| Nonsense_Mutation |
| Nonsense_Mutation |
| Frame_Shift_Del |
| Frame_Shift_Del |
| Splice_Site |
| Frame_Shift_Del |
| Nonsense_Mutation |
| Frame_Shift_Ins |
| Missense_Mutation |
| Missense_Mutation |
| Missense_Mutation |
| Missense_Mutation |
| Missense_Mutation |
| Missense_Mutation |
| Missense_Mutation |
| Missense_Mutation |
| Missense_Mutation |
| Missense_Mutation |
| Missense_Mutation |
| Missense_Mutation |
| Missense_Mutation |
| Missense_Mutation |
| Missense_Mutation |
| Missense_Mutation |
| Missense_Mutation |
| In_Frame_Del |

***KMT2D* somatic mutations in HNSC seperated by *KDM3C* genotype**

| Sample ID | Cancer Type | Gene | *KDM3C* genotype | KMT2D protein change |
|---|---|---|---|---|
| 1 | HNSC | *KMT2D* | A/A | E1490* |
| 2 | HNSC | *KMT2D* | A/A | R5027Tfs*12 |
| 3 | HNSC | *KMT2D* | A/A | L761Hfs*169 |
| 4 | HNSC | *KMT2D* | A/A | Q4687* |
| 5 | HNSC | *KMT2D* | A/A | Q1361* |
| 6 | HNSC | *KMT2D* | A/A | G5310R |
| 7 | HNSC | *KMT2D* | A/A | E5234K |
| 8 | HNSC | *KMT2D* | A/A | G5041R |
| 9 | HNSC | *KMT2D* | A/A | P1669S |
| 10 | HNSC | *KMT2D* | A/T | R5432Q |
| 11 | HNSC | *KMT2D* | A/T | C780* |
| 12 | HNSC | *KMT2D* | A/T | P648Tfs*2 |
| 13 | HNSC | *KMT2D* | A/T | X3480_splice |
| 14 | HNSC | *KMT2D* | A/T | E1902* |
| 15 | HNSC | *KMT2D* | A/T | Q2416* |
| 16 | HNSC | *KMT2D* | A/T | K1466Qfs*25 |
| 17 | HNSC | *KMT2D* | A/T | A3292Gfs*7 |
| 18 | HNSC | *KMT2D* | A/T | X3452_splice |
| 19 | HNSC | *KMT2D* | A/T | L1032Pfs*33 |
| 20 | HNSC | *KMT2D* | A/T | K4653* |
| 21 | HNSC | *KMT2D* | A/T | T3053Yfs*2 |
| 22 | HNSC | *KMT2D* | A/T | R1252* |
| 23 | HNSC | *KMT2D* | A/T | W2006* |
| 24 | HNSC | *KMT2D* | A/T | X1581_splice |
| 25 | HNSC | *KMT2D* | A/T | R1252* |
| 26 | HNSC | *KMT2D* | A/T | E614* |
| 27 | HNSC | *KMT2D* | A/T | Q2540* |
| 28 | HNSC | *KMT2D* | A/T | C5062Rfs*10 |
| 29 | HNSC | *KMT2D* | A/T | X1956_splice |
| 30 | HNSC | *KMT2D* | A/T | X1695_splice |
| 31 | HNSC | *KMT2D* | A/T | Q3744* |
| 32 | HNSC | *KMT2D* | A/T | S4789Cfs*27 |
| 33 | HNSC | *KMT2D* | A/T | P4766L |
| 34 | HNSC | *KMT2D* | A/T | S920C |
| 35 | HNSC | *KMT2D* | A/T | R2774W |
| 36 | HNSC | *KMT2D* | A/T | R3536H |
| 37 | HNSC | *KMT2D* | T/T | S2788= |
| 38 | HNSC | *KMT2D* | T/T | S654Pfs*276 |
| 39 | HNSC | *KMT2D* | T/T | R2410* |
| 40 | HNSC | *KMT2D* | T/T | Q2337* |
| 41 | HNSC | *KMT2D* | T/T | Q2380* |
| 42 | HNSC | *KMT2D* | T/T | X2037_splice |
| 43 | HNSC | *KMT2D* | T/T | E1158* |
| 44 | HNSC | *KMT2D* | T/T | Q1035Pfs*33 |
| 45 | HNSC | *KMT2D* | T/T | G1916Vfs*129 |
| 46 | HNSC | *KMT2D* | T/T | A1109Gfs*2 |
| 47 | HNSC | *KMT2D* | T/T | E1649* |
| 48 | HNSC | *KMT2D* | T/T | E308* |
| 49 | HNSC | *KMT2D* | T/T | V4472Afs*14 |
| 50 | HNSC | *KMT2D* | T/T | X59_splice |
| 51 | HNSC | *KMT2D* | T/T | Q2380* |

| | | | | |
|---|---|---|---|---|
| 52 | HNSC | *KMT2D* | T/T | Q56* |
| 53 | HNSC | *KMT2D* | T/T | S1632* |
| 54 | HNSC | *KMT2D* | T/T | K287Vfs*18 |
| 55 | HNSC | *KMT2D* | T/T | P4684Lfs*113 |
| 56 | HNSC | *KMT2D* | T/T | R5501* |
| 57 | HNSC | *KMT2D* | T/T | Q2000* |
| 58 | HNSC | *KMT2D* | T/T | R2282Gfs*4 |
| 59 | HNSC | *KMT2D* | T/T | V1244Gfs*6 |
| 60 | HNSC | *KMT2D* | T/T | F2739Lfs*18 |
| 61 | HNSC | *KMT2D* | T/T | V3089Wfs*30 |
| 62 | HNSC | *KMT2D* | T/T | E2723* |
| 63 | HNSC | *KMT2D* | T/T | L238Cfs*23 |
| 64 | HNSC | *KMT2D* | T/T | E649Pfs*276 |
| 65 | HNSC | *KMT2D* | T/T | L1461Tfs*30 |
| 66 | HNSC | *KMT2D* | T/T | R4484* |
| 67 | HNSC | *KMT2D* | T/T | R1615* |
| 68 | HNSC | *KMT2D* | T/T | P2218Lfs*46 |
| 69 | HNSC | *KMT2D* | T/T | S1133* |
| 70 | HNSC | *KMT2D* | T/T | H5114Y |
| 71 | HNSC | *KMT2D* | T/T | S2483C |
| 72 | HNSC | *KMT2D* | T/T | E1391K |
| 73 | HNSC | *KMT2D* | T/T | G4356E |
| 74 | HNSC | *KMT2D* | T/T | C5062F |
| 75 | HNSC | *KMT2D* | T/T | E2259D |
| 76 | HNSC | *KMT2D* | T/T | R5214H |
| 77 | HNSC | *KMT2D* | T/T | P3494L |
| 78 | HNSC | *KMT2D* | T/T | P1931T |
| 79 | HNSC | *KMT2D* | T/T | L213P |
| 80 | HNSC | *KMT2D* | T/T | A1716S |
| 81 | HNSC | *KMT2D* | T/T | A262T |
| 82 | HNSC | *KMT2D* | T/T | G5189R |
| 83 | HNSC | *KMT2D* | T/T | R371G |
| 84 | HNSC | *KMT2D* | T/T | R5159W |
| 85 | HNSC | *KMT2D* | T/T | D5518G |
| 86 | HNSC | *KMT2D* | T/T | G2899S |
| 87 | HNSC | *KMT2D* | T/T | Q3745del |
| 88 | HNSC | *KMT2D* | T/T | S714L |

| Mutation type |
|---|
| Nonsense_Mutation |
| Frame_Shift_Ins |
| Frame_Shift_Del |
| Nonsense_Mutation |
| Nonsense_Mutation |
| Missense_Mutation |
| Missense_Mutation |
| Missense_Mutation |
| Missense_Mutation |
| Missense_Mutation |
| Nonsense_Mutation |
| Frame_Shift_Ins |
| Splice_Site |
| Nonsense_Mutation |
| Nonsense_Mutation |
| Frame_Shift_Ins |
| Frame_Shift_Del |
| Splice_Site |
| Frame_Shift_Del |
| Nonsense_Mutation |
| Frame_Shift_Ins |
| Nonsense_Mutation |
| Nonsense_Mutation |
| Splice_Site |
| Nonsense_Mutation |
| Nonsense_Mutation |
| Nonsense_Mutation |
| Frame_Shift_Del |
| Splice_Site |
| Splice_Site |
| Nonsense_Mutation |
| Frame_Shift_Del |
| Missense_Mutation |
| Missense_Mutation |
| Missense_Mutation |
| Missense_Mutation |
| Splice_Region |
| Frame_Shift_Del |
| Nonsense_Mutation |
| Nonsense_Mutation |
| Nonsense_Mutation |
| Splice_Site |
| Nonsense_Mutation |
| Frame_Shift_Ins |
| Frame_Shift_Del |
| Frame_Shift_Del |
| Nonsense_Mutation |
| Nonsense_Mutation |
| Frame_Shift_Del |
| Splice_Site |
| Nonsense_Mutation |

**Summary Table**

| KMT2D mutations | A/A | A/T | T/T |
|---|---|---|---|
| Missense | 4 | 5 | 18 |
| Nonsense | 3 | 10 | 15 |
| Frameshift any | 2 | 7 | 15 |
| Splice/splice-region | 0 | 5 | 3 |
| In-frame deletion | 0 | 0 | 1 |

Nonsense_Mutation
Nonsense_Mutation
Frame_Shift_Ins
Frame_Shift_Del
Nonsense_Mutation
Nonsense_Mutation
Frame_Shift_Del
Frame_Shift_Ins
Frame_Shift_Del
Frame_Shift_Del
Nonsense_Mutation
Frame_Shift_Del
Frame_Shift_Del
Frame_Shift_Ins
Nonsense_Mutation
Nonsense_Mutation
Frame_Shift_Del
Nonsense_Mutation
Missense_Mutation
Missense_Mutation
Missense_Mutation
Missense_Mutation
Missense_Mutation
Missense_Mutation
Missense_Mutation
Missense_Mutation
Missense_Mutation
Missense_Mutation
Missense_Mutation
Missense_Mutation
Missense_Mutation
Missense_Mutation
Missense_Mutation
Missense_Mutation
Missense_Mutation
In_Frame_Del
Missense_Mutation

**Supplementary Table 14. Phosphorylation sites detected in MDC1 in CAL-27 cells bearing KMD3C WT-S464**

| | KDM3C A/A (no IR) | KDM3C A/A (post IR) | KDM3C T/T (no IR) | KDM3C T/T (post IR) |
|---|---|---|---|---|
| **T17** | 1 | 0 | 0 | 0 |
| **S38** | 0 | 0 | 0 | 1 |
| **S70** | 1 | 0 | 0 | 0 |
| **S72** | 1 | 0 | 0 | 0 |
| **S94** | 0 | 1 | 0 | 1 |
| **Y130** | 1 | 0 | 0 | 0 |
| **S136** | 1 | 1 | 0 | 1 |
| **S141** | 0 | 0 | 1 | 1 |
| **T150** | 0 | 1 | 0 | 0 |
| **S168** | 1 | 0 | 0 | 0 |
| **S176** | 1 | 0 | 0 | 0 |
| **S196** | 1 | 0 | 0 | 0 |
| **T230** | 1 | 0 | 0 | 0 |
| **S234** | 0 | 1 | 0 | 0 |
| **S235** | 0 | 1 | 0 | 0 |
| **T242** | 1 | 1 | 1 | 0 |
| **T248** | 0 | 1 | 0 | 0 |
| **T254** | 1 | 1 | 1 | 0 |
| **S329** | 1 | 0 | 0 | 0 |
| **T331** | 1 | 0 | 0 | 0 |
| **T358** | 1 | 1 | 1 | 1 |
| **S372 (SQ)** | 1 | 1 | 0 | 0 |
| **S376** | 1 | 0 | 1 | 0 |
| **S394 (SQ)** | 1 | 0 | 0 | 0 |
| **S397 (SQ)** | 1 | 0 | 0 | 0 |
| **S411** | 0 | 0 | 1 | 0 |
| **T415** | 1 | 0 | 1 | 0 |
| **S422 (SQ)** | 1 | 0 | 0 | 0 |
| **S422** | 0 | 0 | 1 | 0 |
| **S433** | 0 | 0 | 1 | 0 |
| **T447** | 1 | 0 | 0 | 0 |
| **T449 (TQ)** | 0 | 0 | 1 | 0 |
| **T475** | 1 | 0 | 0 | 0 |
| **S485** | 0 | 1 | 1 | 0 |
| **S513 (SQ)** | 0 | 1 | 0 | 0 |
| **T548** | 1 | 0 | 0 | 0 |
| **T585** | 1 | 1 | 0 | 0 |
| **T588** | 0 | 1 | 1 | 0 |
| **S590** | 0 | 0 | 1 | 0 |
| **S598 (SQ)** | 1 | 0 | 0 | 0 |
| **S641** | 0 | 0 | 1 | 0 |
| **T646** | 0 | 0 | 1 | 0 |
| **T652** | 0 | 1 | 1 | 0 |
| **T654** | 0 | 1 | 1 | 0 |
| **T670** | 1 | 1 | 1 | 0 |
| **T681** | 0 | 1 | 0 | 0 |
| **S713** | 0 | 0 | 1 | 0 |

**Key:**
1 denotes that phosph
0 denotes that phosph

| | | | | |
|---|---|---|---|---|
| **T725** | 1 | 0 | 0 | 0 |
| **T739** | 0 | 0 | 1 | 0 |
| **T740** | 0 | 0 | 1 | 0 |
| **S780** | 1 | 0 | 1 | 0 |
| **S793** | 0 | 0 | 1 | 0 |
| **T809** | 0 | 0 | 1 | 0 |
| **T820** | 1 | 0 | 1 | 0 |
| **T844** | 0 | 0 | 1 | 0 |
| **T847** | 1 | 0 | 0 | 0 |
| **T853** | 1 | 0 | 0 | 0 |
| **S889** | 1 | 0 | 0 | 1 |
| **S896** | 1 | 0 | 0 | 0 |
| **T896** | 0 | 1 | 0 | 0 |
| **S912** | 0 | 1 | 0 | 1 |
| **T946 (TQ)** | 1 | 1 | 1 | 0 |
| **S1018** | 1 | 0 | 1 | 1 |
| **S1027** | 1 | 0 | 0 | 0 |
| **T1081** | 0 | 1 | 0 | 0 |
| **S1086 (SQ)** | 0 | 1 | 0 | 0 |
| **T1138** | 0 | 0 | 1 | 0 |
| **T1142** | 0 | 1 | 0 | 0 |
| **S1143 (SQ)** | 0 | 1 | 0 | 0 |
| **S1153** | 0 | 1 | 0 | 0 |
| **T1157** | 0 | 1 | 0 | 1 |
| **S1179** | 1 | 0 | 0 | 0 |
| **T1183** | 0 | 1 | 0 | 0 |
| **T1187** | 0 | 0 | 1 | 0 |
| **S1192** | 0 | 1 | 0 | 0 |
| **S1194** | 0 | 1 | 0 | 0 |
| **S1195** | 1 | 1 | 0 | 0 |
| **S1197** | 0 | 0 | 1 | 0 |
| **T1205** | 0 | 1 | 0 | 0 |
| **S1212** | 0 | 1 | 0 | 0 |
| **S1235** | 1 | 1 | 0 | 0 |
| **S1239** | 1 | 0 | 0 | 0 |
| **S1253** | 0 | 0 | 1 | 0 |
| **T1269** | 1 | 0 | 0 | 0 |
| **S1276** | 0 | 1 | 0 | 0 |
| **S1277** | 0 | 1 | 0 | 0 |
| **T1280** | 1 | 1 | 0 | 0 |
| **T1287** | 1 | 0 | 0 | 0 |
| **S1287** | 0 | 0 | 1 | 0 |
| **S1294** | 0 | 1 | 0 | 0 |
| **T1302** | 1 | 0 | 0 | 0 |
| **T1309** | 0 | 1 | 0 | 0 |
| **T1310** | 1 | 0 | 0 | 0 |
| **S1314** | 1 | 0 | 0 | 0 |
| **S1318** | 0 | 0 | 1 | 0 |
| **T1328** | 1 | 0 | 0 | 1 |
| **S1337** | 0 | 1 | 0 | 0 |

| | | | | |
|---|---|---|---|---|
| **S1365** | 1 | 0 | 0 | 0 |
| **T1366** | 0 | 1 | 0 | 0 |
| **T1384** | 1 | 1 | 0 | 0 |
| **T1388** | 0 | 1 | 0 | 0 |
| **T1403** | 1 | 0 | 1 | 0 |
| **T1444** | 0 | 0 | 1 | 0 |
| **T1451** | 0 | 1 | 0 | 0 |
| **T1466** | 0 | 1 | 0 | 0 |
| **T1470** | 0 | 1 | 0 | 0 |
| **T1474** | 0 | 1 | 0 | 0 |
| **T1485** | 1 | 0 | 0 | 0 |
| **T1567** | 0 | 0 | 1 | 0 |
| **S1581** | 0 | 1 | 1 | 1 |
| **T1581** | 0 | 0 | 1 | 0 |
| **S1583** | 0 | 1 | 0 | 0 |
| **T1589** | 1 | 0 | 1 | 1 |
| **T1608** | 1 | 1 | 0 | 0 |
| **T1630** | 0 | 0 | 1 | 0 |
| **S1645** | 0 | 1 | 1 | 0 |
| **S1646** | 0 | 0 | 1 | 0 |
| **S1681** | 1 | 1 | 1 | 0 |
| **T1683** | 0 | 0 | 1 | 0 |
| **S1686** | 0 | 0 | 1 | 1 |
| **S1687** | 0 | 0 | 1 | 0 |
| **T1696** | 0 | 1 | 0 | 0 |
| **S1766** | 1 | 0 | 1 | 0 |
| **S1768** | 1 | 0 | 0 | 0 |
| **S1775** | 1 | 1 | 1 | 0 |
| **T1781** | 1 | 0 | 0 | 0 |
| **S1786 (SQ)** | 1 | 0 | 1 | 0 |
| **S1797** | 0 | 0 | 1 | 0 |
| **T1800** | 0 | 1 | 0 | 0 |
| **S1808 (SQ)** | 0 | 1 | 0 | 0 |
| **S1814** | 0 | 1 | 0 | 0 |
| **S1820** | 0 | 1 | 0 | 0 |
| **S1833 (SQ)** | 1 | 1 | 0 | 0 |
| **T1836** | 1 | 0 | 0 | 0 |
| **S1890** | 1 | 0 | 0 | 0 |
| **T1898** | 0 | 1 | 0 | 0 |
| **S1915** | 0 | 1 | 0 | 0 |
| **S1919** | 0 | 0 | 0 | 1 |
| **S1924** | 0 | 1 | 0 | 0 |
| **T1928** | 1 | 1 | 1 | 0 |
| **T1934** | 1 | 1 | 0 | 0 |
| **S1949** | 1 | 0 | 1 | 0 |
| **S1956** | 1 | 1 | 1 | 0 |
| **T1971** | 0 | 1 | 0 | 0 |
| **S1982** | 1 | 0 | 0 | 1 |
| **S1988** | 1 | 0 | 0 | 0 |
| **Y1999** | 0 | 1 | 0 | 0 |

| | | | | |
|---|---|---|---|---|
| **T2004** | 1 | 1 | 0 | 0 |
| **S2028** | 1 | 1 | 0 | 0 |
| **S2032** | 0 | 1 | 0 | 0 |
| **S2082** | 1 | 1 | 1 | 0 |
| **S2087** | 1 | 0 | 0 | 0 |

**[illegible] or SNP-S464T at baseline and 1h post 10Gy IR**

**Supplementary Table 15. Protein-pair models with ipTM ≥ 0.6 and ipSAE ≥0.4 out of 50 models**

**Domain names for KDM3C and its partner proteins, listed in columns B and D, respectively, are based on t**

| Domain Pair model name | KDM3C Domain Assignment | KDM3C domain residue range |
|---|---|---|
| KDM3C-E5_RNF8-E1 | SH3 | 6-180 |
| KDM3C-E4_RNF8-E1 | Double-stranded beta-helix | 2156-2495 |
| KDM3C-nd2_MDC1-E3 | Inter-domain | 1926-2155 |
| KDM3C-nd1_RNF8-E2 | Inter-domain | 256-1695 |
| KDM3C-nd1_RNF8-E1 | Inter-domain | 256-1695 |
| KDM3C-E3_MDC1-E1 | SH3 | 181-255 |
| KDM3C-E2_MDC1-nd2 | LIM domain-like | 1776-1875 |
| KDM3C-nd1_MDC1-E1 | Inter-domain | 256-1695 |
| KDM3C-nd2_RNF8-E1 | Inter-domain | 1926-2155 |
| KDM3C-E3_MDC1-E2 | SH3 | 181-255 |
| KDM3C-nd2_MDC1-E1 | Inter-domain | 1926-2155 |
| KDM3C-nd1_MDC1-E2 | Inter-domain | 256-1695 |
| KDM3C-E2_RNF8-E3 | LIM domain-like | 1776-1875 |
| KDM3C-E4_MDC1-E2 | Double-stranded beta-helix | 2156-2495 |

:he ECOD database (Reference 63). The inter-domain region of KDM3C refers to its intrinsically disordered r

| Partner Protein Domain Assignment | Partner Protein domain residue range | No. of Models (ipTM ≥ 0.6) out of 50 |
|---|---|---|
| RNF8_SMAD/FHA domain | 16-135 | 41 |
| RNF8_SMAD/FHA domain | 16-135 | 37 |
| MDC1_SMAD/FHA domain | 31-135 | 29 |
| RNF8_CT398 helical hairpin | 236-330 | 23 |
| RNF8_SMAD/FHA domain | 16-135 | 15 |
| MDC1_BRCT domain | 1891-1990 | 14 |
| MDC1_Inter-domain | 136-1925 | 13 |
| MDC1_BRCT domain | 1891-1990 | 11 |
| RNF8_SMAD/FHA domain | 16-135 | 8 |
| MDC1_BRCT domain | 1991-2085 | 9 |
| MDC1_BRCT domain | 1891-1990 | 8 |
| MDC1_BRCT domain | 1991-2085 | 8 |
| RNF8_RING/U-box-like | 386-480 | 7 |
| MDC1_BRCT domain | 991-2085 | 7 |

region.

| No of model ipsae ≥ 0.4 out of 50 |
|---|
| 41 |
| 34 |
| 17 |
| 0 |
| 0 |
| 8 |
| 0 |
| 0 |
| 1 |
| 6 |
| 7 |
| 0 |
| 2 |
| 0 |

**Supplementary Table 16. NetPhos prediction results**

| Sequence | # x | Context (p.S464) | Score | Kinase | Answer | Note |
|---|---|---|---|---|---|---|
| # Sequence | 5S | MIIHSSEQS | 0.461 | GSK3 | . | NetPhos s[illegible] |
| # Sequence | 5S | MIIHSSEQS | 0.461 | CDC2 | . | Score abo[illegible] |
| # Sequence | 5S | MIIHSSEQS | 0.446 | PKC | . | |
| # Sequence | 5S | MIIHSSEQS | 0.423 | CaM-II | . | |
| # Sequence | 5S | MIIHSSEQS | 0.423 | CKII | . | |
| # Sequence | 5S | MIIHSSEQS | 0.406 | unsp | . | |
| # Sequence | 5S | MIIHSSEQS | 0.405 | DNAPK | . | |
| # Sequence | 5S | MIIHSSEQS | 0.378 | CKI | . | |
| # Sequence | 5S | MIIHSSEQS | 0.344 | p38MAPK | . | |
| # Sequence | 5S | MIIHSSEQS | 0.3 | ATM | . | |
| # Sequence | 5S | MIIHSSEQS | 0.266 | PKG | . | |
| # Sequence | 5S | MIIHSSEQS | 0.259 | RSK | . | |
| # Sequence | 5S | MIIHSSEQS | 0.25 | PKA | . | |
| # Sequence | 5S | MIIHSSEQS | 0.146 | cdk5 | . | |
| # Sequence | 5S | MIIHSSEQS | 0.081 | PKB | . | |

| Sequence | # x | Context (p.T464) | Score | Kinase | Answer |
|---|---|---|---|---|---|
| # Sequence | 5T | MIIHTSEQS | 0.56 | PKC | Yes |
| # Sequence | 5T | MIIHSSEQS | 0.453 | GSK3 | . |
| # Sequence | 5T | MIIHSSEQS | 0.429 | cdc2 | . |
| # Sequence | 5T | MIIHSSEQS | 0.416 | CaM-II | . |
| # Sequence | 5T | MIIHSSEQS | 0.412 | CKII | . |
| # Sequence | 5T | MIIHSSEQS | 0.37 | CKI | . |
| # Sequence | 5T | MIIHSSEQS | 0.364 | DNAPK | . |
| # Sequence | 5T | MIIHSSEQS | 0.36 | p38MAPK | . |
| # Sequence | 5T | MIIHSSEQS | 0.273 | PKG | . |
| # Sequence | 5T | MIIHSSEQS | 0.253 | ATM | . |
| # Sequence | 5T | MIIHSSEQS | 0.203 | RSK | . |
| # Sequence | 5T | MIIHSSEQS | 0.146 | cdk5 | . |
| # Sequence | 5T | MIIHSSEQS | 0.13 | PKA | . |
| # Sequence | 5T | MIIHSSEQS | 0.08 | PKB | . |
| # Sequence | 5T | MIIHSSEQS | 0.05 | unsp | . |

ve the standard 0.5 threshold indicates a strong positive prediction for potential phosphorylation.

**Supplementary Table 17. Analysis of baseline differentially expressed genes across KDM3C SNP-S464[illegible]**

| | |
|---|---|
| **Downregulated genes** | Baseline downregulated genes in KDM3C SNP-S464T vs WT[illegible] |
| **Upregulated genes** | Baseline upregulated genes in KDM3C SNP-S464T vs WT-S[illegible] |
| **Analysis** | Overlap of baseline differentially expressed genes across KD[illegible] |

**T vs. WT-S464 isogenic models**

T-S464 isogenic models

464 isogenic models

M3C SNP-S464T vs. WT-S464 isogenic models

**Baseline downregulated genes in KDM3C SNP-S464T vs WT-S464 isogenic models**

| RCM-1 | CAL-27 | SCC-9 |
|---|---|---|
| *ABCC2* | *AARSD1* | *ABCA1* |
| *AC005822.1* | *ABI3BP* | *AC009220.3* |
| *AC012065.5* | *AC003681.1* | *AC078899.1* |
| *AC012501.2* | *AC004801.2* | *AC132938.2* |
| *AC078883.1* | *AC008267.3* | *ACACB* |
| *AC091804.1* | *AC008982.2* | *ACTG1P15* |
| *AC098934.1* | *AC009090.6* | *ADAMTS2* |
| *AC098934.2* | *AC009533.1* | *ADAMTSL4* |
| *AC109322.1* | *AC010326.3* | *ADGRF1* |
| *AC126696.2* | *AC010761.1* | *ADGRL1* |
| *ACAP1* | *AC011510.1* | *ADGRL3* |
| *ACBD7* | *AC012615.6* | *AGPAT4* |
| *ACE2* | *AC027644.1* | *AIM2* |
| *ACKR3* | *AC066613.1* | *AK8* |
| *ACOT13* | *AC087385.1* | *AKR1C3* |
| *ACP5* | *AC090425.2* | *AKR1C7P* |
| *ADAMTS17* | *AC093495.1* | *AL158071.4* |
| *ADAP2* | *AC132872.4* | *AL662797.1* |
| *ADAT3* | *AC135279.2* | *ALDH1A2* |
| *ADGRF1* | *AC243654.2* | *ALOX15* |
| *AGT* | *ACAA2* | *ALPG* |
| *AL031320.2* | *ADAMTS1* | *ALPPL2* |
| *AL136295.5* | *ADAMTS12* | *AMOT* |
| *AL162151.2* | *ADAMTS16* | *ANK1* |
| *AL512380.2* | *ADAMTS6* | *AP000679.1* |
| *ALAS2* | *ADAMTS7* | *AP001267.3* |
| *ANGPTL4* | *ADRA1B* | *APLN* |
| *ANPEP* | *ADSL* | *APOL1* |
| *AP000879.1* | *AEN* | *APOL3* |
| *APCDD1* | *AGAP5* | *ARHGEF37* |
| *APLN* | *AKAP12* | *ARHGEF6* |
| *APOBEC3C* | *AL023807.1* | *ARL6IP4* |
| *APOBEC3F* | *AL049840.1* | *BARX1* |
| *APOBEC3G* | *AL135903.1* | *BCAM* |
| *APOBEC3H* | *AL161452.1* | *BCL11B* |
| *ARHGAP44* | *AL355802.2* | *BCL2A1* |
| *ARSE* | *AL391058.1* | *BDKRB2* |
| *ASB4* | *AL512791.2* | *BOLA2B* |
| *ATP6V0A4* | *AL590762.1* | *C3* |
| *AVPR2* | *AL731571.1* | *C7* |
| *BAMBI* | *ALOXE3* | *CACNA2D3* |
| *BCL2* | *AMH* | *CACNA2D4* |
| *BMP2* | *ANAPC15* | *CACNG7* |
| *BMPR1AP1* | *ANGPTL4* | *CALCRL* |
| *BORCS8* | *ANKRD1* | *CALML5* |
| *BSN* | *ANKRD10* | *CARD14* |
| *CA9* | *ANP32E* | *CASP1* |
| *CACNA1H* | *ANTXR2* | *CBFA2T3* |
| *CAMK1D* | *AP000347.1* | *CBLC* |
| *CAMK2N2* | *AP001107.6* | *CCDC8* |
| *CARD11* | *AP003352.1* | *CCDC80* |

*CBX2*
*CCL5*
*CCR1*
*CCR3*
*CD24*
*CD74*
*CDH11*
*CDH16*
*CDH26*
*CDH3*
*CDK5R1*
*CDYL2*
*CEL*
*CELF2*
*CENPS*
*CHRNA3*
*CIDEC*
*CIITA*
*CKB*
*CKMT2*
*CLDN2*
*CMPK2*
*CNTN1*
*COCH*
*COL5A3*
*CPE*
*CPLX2*
*CRIP2*
*CTAG2*
*CTNNA2*
*CX3CL1*
*CXCL5*
*CXCR3*
*CYP26B1*
*CYP2C18*
*CYP2W1*
*DAAM2*
*DAPK1*
*DIAPH3-AS1*
*DISP3*
*DLL1*
*DLX3*
*DPYSL3*
*DQX1*
*DSC3*
*DSG3*
*DTX1*
*EDN2*
*EGLN3*
*EMILIN2*
*ENC1*
*EPHB1*
*ETV2*
*EVA1A*
*FBXO17*
*AQP11*
*AR*
*ARHGAP44*
*ARL9*
*ASB16*
*ASF1B*
*ASPM*
*ATAD3B*
*ATF3*
*AURKA*
*AURKB*
*AXL*
*BIRC5*
*BLM*
*BMP2*
*BMP4*
*BMP5*
*BMS1P4*
*BORA*
*BRCA1*
*BRIP1*
*BUB1*
*BUB1B*
*CAV1*
*CAVIN4*
*CCDC39*
*CCDC80*
*CCL2*
*CCL20*
*CCL5*
*CCNB1*
*CCNB2*
*CCND1*
*CCND2*
*CCP110*
*CD3EAP*
*CD70*
*CDC20*
*CDC25C*
*CDC42*
*CDC45*
*CDC6*
*CDCA5*
*CDCA8*
*CDK1*
*CDK2*
*CDK5RAP3*
*CDT1*
*CENPA*
*CENPE*
*CENPF*
*CENPI*
*CENPK*
*CENPL*
*CENPM*
*CD14*
*CD24*
*CD70*
*CDH16*
*CDH18*
*CDH3*
*CDK18*
*CDKN1C*
*CEACAM1*
*CEBPA*
*CERS4*
*CFD*
*CGN*
*CKB*
*CLCA2*
*CLDN4*
*CLDN7*
*CLSTN2*
*CMKLR1*
*COL1A2*
*COX6B2*
*COX7B2*
*CREB3L1*
*CRIP2*
*CSF1R*
*CXADR*
*CXCL10*
*CXXC5*
*CYP1A1*
*CYP1B1*
*CYP24A1*
*CYP2B6*
*DAPK1*
*DBP*
*DCLK1*
*DHRS3*
*DLK2*
*DOCK8*
*DPYSL3*
*DRAM1*
*DSC1*
*DSCAM*
*DSG3*
*EDAR*
*EFNA3*
*EGFL7*
*ELANE*
*ELF3*
*ELOVL3*
*EPHA1*
*EPHB6*
*EPPK1*
*ESPN*
*FAM86B1*
*FBXL16*

| | | |
|---|---|---|
| *FBXO24* | *CENPN* | *FGD3* |
| *FCGRT* | *CENPO* | *FGF7P6* |
| *FGFBP3* | *CENPU* | *FGFBP1* |
| *FHL1* | *CENPW* | *FGFR3* |
| *FLRT3* | *CEP152* | *FKBP1B* |
| *FOLR1* | *CEP192* | *FLRT2* |
| *FOXL2* | *CEP70* | *FLRT3* |
| *FRMPD3* | *CEP72* | *FOXA1* |
| *FUT7* | *CGB8* | *FP565260.3* |
| *FYB1* | *CHAF1A* | *FRK* |
| *FZD8* | *CHAF1B* | *FST* |
| *G0S2* | *CHEK1* | *FYB1* |
| *GAS7* | *CHEK2* | *GAL* |
| *GDF9* | *CKAP5* | *GALC* |
| *GGT5* | *COL12A1* | *GBP6* |
| *GGT6* | *COL18A1* | *GCNT3* |
| *GJA1* | *COL1A1* | *GDAP1* |
| *GJB5* | *COL1A2* | *GFI1* |
| *GPC4* | *COL4A1* | *GGT4P* |
| *GTF2IP7* | *COL4A2* | *GGT6* |
| *GUCA2A* | *COL4A6* | *GJA1* |
| *GZMB* | *COL5A1* | *GJA5* |
| *HAAO* | *COL6A1* | *GMPR* |
| *HACD1* | *COL9A2* | *GNAO1* |
| *HES2* | *CSF2* | *GRAMD4* |
| *HEY1* | *CSPG4* | *GRHL1* |
| *HEY2* | *CTBP1-AS* | *GRHL3* |
| *HIST1H2AC* | *CTHRC1* | *GRIN3B* |
| *HIST1H2AG* | *CXCL1* | *GTF2IP14* |
| *HIST1H2BC* | *CXCL10* | *GUCY1A1* |
| *HIST1H2BD* | *CXCL11* | *HAP1* |
| *HIST1H2BG* | *CXCL2* | *HCAR2* |
| *HIST1H2BK* | *CXCL3* | *HCAR3* |
| *HIST1H3A* | *CXCL5* | *HES2* |
| *HIST1H3E* | *CXCL8* | *HES7* |
| *HIST1H3H* | *CXCL9* | *HIST1H3H* |
| *HIST1H4H* | *CYP27B1* | *HIST2H4B* |
| *HIST1H4I* | *DAG1* | *HLA-F* |
| *HLA-DPA1* | *DAPK1* | *HMGN5* |
| *HLA-DPB1* | *DCTN1* | *HOXA5* |
| *HLA-DRA* | *DDX12P* | *HRASLS2* |
| *HLA-DRB1* | *DDX47* | *HRK* |
| *HMGCS2* | *DIO2* | *HSH2D* |
| *HOXD9* | *DKK1* | *HYAL4* |
| *HPGD* | *DLG1* | *ICOSLG* |
| *HTRA1* | *DLGAP5* | *ID1* |
| *HYAL1* | *DMC1* | *ID2* |
| *IFITM1* | *DNA2* | *ID4* |
| *IGFBP2* | *DSCC1* | *IDO1* |
| *IGFBP7* | *EBI3* | *IFI27* |
| *IL10* | *ECT2* | *IFITM1* |
| *IL27RA* | *EDN1* | *IFITM3* |
| *IRX2* | *EFEMP2* | *IGFBP6* |
| *ITGA9* | *EGFR-AS1* | *IGHV3-43* |
| *ITPKA* | *EGLN2* | *IL1RN* |

*ITPR2*
*KBTBD11*
*KCNIP3*
*KIT*
*KLHDC8A*
*KLHL29*
*KLK7*
*KRT16*
*KRT17*
*LAP3P2*
*LAPTM5*
*LIMS2*
*LINC00957*
*LINC02001*
*LRP4*
*LYPD3*
*MAFB*
*MAGEA2B*
*MAGEA3*
*MAP1LC3A*
*MAT1A*
*MATN2*
*METTL7A*
*MICB*
*MME*
*MMP9*
*MPP1*
*MRPL53*
*MSN*
*MUC1*
*MUC16*
*MYCN*
*MYO3B*
*NANOGP1*
*NDP*
*NDRG1*
*NEDD9*
*NFE2*
*NGFR*
*NNMT*
*NOD2*
*NQO2*
*NRXN3*
*NUPR1*
*OXCT1*
*P2RX5-TAX1BP3*
*PAGE1*
*PCDH19*
*PCDH7*
*PCDHB2*
*PDE2A*
*PDE8B*
*PDK3*
*PECAM1*
*PEX11G*
*EIF3CL*
*EIF4A1*
*EME1*
*EML1*
*ENG*
*ERCC2*
*ESCO2*
*ESPL1*
*ESR2*
*ETV4*
*ETV5*
*EVI2B*
*EYA1*
*F3*
*FAM83D*
*FANCD2*
*FAS*
*FBLN1*
*FBN2*
*FBXO5*
*FERMT3*
*FGF1*
*FGF5*
*FGFBP1*
*FGFR3*
*FHL1*
*FN1*
*FOSL1*
*FOXC2*
*FRMD6*
*FUT4*
*GAL*
*GAS6-AS1*
*GBX2*
*GEN1*
*GINS1*
*GINS2*
*GINS4*
*GNG13*
*GOLGA2P7*
*GOLGA8B*
*GPAT3*
*GPD1*
*GPER1*
*GPI*
*GPR3*
*GPR75*
*GRB14*
*GRHL3*
*GRIN3B*
*GRPR*
*H2AFX*
*HASPIN*
*HAUS1*
*HAUS4*
*IL20RA*
*IL9RP3*
*IQSEC3*
*IRF6*
*ITGB7*
*IVL*
*JAG2*
*JUP*
*KCNH5*
*KIF26A*
*KLHDC7B*
*KLHL29*
*KLK7*
*KPNA7*
*KREMEN2*
*KRT1*
*KRT14*
*KRT16*
*KRT17*
*KRT19*
*KRT5*
*L1CAM*
*LAMA4*
*LDHAP7*
*LGALS7*
*LMX1B*
*LOC107984053*
*LOXL4*
*LPAR5*
*LPCAT4*
*LRG1*
*LTB4R*
*LTBP1*
*LTF*
*MACC1*
*MACROD2*
*MAFA*
*MAG*
*MAGEA3*
*MAPK4*
*MARCO*
*MATN2*
*MB*
*MCC*
*ME3*
*MEF2C*
*MFAP5*
*MGMT*
*MGST1*
*MMP28*
*MMP7*
*MMP9*
*MPP7*
*MPZL2*
*MSX1*

*PGAM4*
*PLAC8*
*PLAGL1*
*PLPP3*
*PPBP*
*PPP2R2C*
*PRDM12*
*PRF1*
*PROC*
*PRODH*
*PRSS2*
*PSD*
*PTAFR*
*PTGER4*
*PXDN*
*PYCARD*
*PYCARD-AS1*
*RAB3A*
*RASD1*
*RASD2*
*RGS9*
*RNASE1*
*RNASE6*
*RNF144A*
*ROBO4*
*RPL13AP7*
*RRBP1*
*S100A9*
*SCARA3*
*SCN5A*
*SELENOP*
*SFMBT2*
*SFRP4*
*SHC3*
*SIRPA*
*SLC2A3*
*SLC37A2*
*SLC3A1*
*SLC6A4*
*SMG1P5*
*SMPD3*
*SNORC*
*SOAT1*
*SOCS1*
*SOHLH2*
*SOX18*
*SPHK1*
*SPON2*
*SSTR5*
*STMN3*
*SULF2*
*SYNE4*
*TAL1*
*TBL1X*
*TESK2*
*HAUS7*
*HAUS8*
*HELLS*
*HHEX*
*HIST1H2BL*
*HJURP*
*HMCN1*
*HMGA2*
*HMGB1*
*HMGB2*
*HMGB3*
*HSP90AA1*
*HSP90AA2P*
*HSP90AB3P*
*HSPA1A*
*HSPA2*
*HSPD1P11*
*HSPG2*
*ICAM1*
*IL11*
*IL1A*
*IL1B*
*IL1R1*
*IL1R2*
*IL1RL1*
*IL27RA*
*IL36G*
*IL36RN*
*IL6*
*IL7*
*IL7R*
*IL9R*
*INCENP*
*ITGAV*
*ITGB1BP2*
*KIAA1324*
*KIF14*
*KIF18A*
*KIF18B*
*KIF20B*
*KIF22*
*KIF23*
*KIF2C*
*KIF4A*
*KIFC1*
*KITLG*
*KLHL31*
*KNL1*
*KNSTRN*
*KNTC1*
*LAMA3*
*LAMA4*
*LAMB1*
*LAMB2*
*LIG1*
*MUC1*
*MX1*
*MYB*
*MYH14*
*MYLIP*
*MYO16*
*MYO3B*
*NAP1L2*
*NAPSA*
*NECTIN1*
*NECTIN4*
*NEFL*
*NFASC*
*NFE2*
*NKX2-1*
*NLGN4X*
*NLGN4Y*
*NMU*
*NOTCH3*
*NOXA1*
*NPNT*
*NRARP*
*NTN1*
*OAS1*
*OAS2*
*OCLN*
*OLR1*
*OVOL1*
*PADI2*
*PADI3*
*PARD6A*
*PAX9*
*PCDHB16*
*PCDHB2*
*PCDHGA1*
*PCDHGA10*
*PCSK9*
*PGAM1P8*
*PGLYRP4*
*PKIB*
*PKP1*
*PLA2G2F*
*PLCB1*
*PLCG2*
*POU3F1*
*PPL*
*PPM1N*
*PRORSD1P*
*PTGIS*
*PTN*
*PTP4A3*
*PTPRG*
*PTPRZ1*
*PXN-AS1*
*PYGL*

*TGFB2*
*TIAM1*
*TINAG*
*TLE1*
*TLE2*
*TMEM117*
*TNC*
*TNFSF9*
*TNMD*
*TPTEP1*
*TRIB2*
*TRPV2*
*TSPEAR*
*U2AF1*
*UGT1A6*
*ULK2*
*USP18*
*VASH2*
*VGF*
*VRTN*
*WIPF3*
*WNT7A*
*WNT9A*
*XPNPEP2*
*Z92544.2*
*ZBTB46*
*ZNF304*
*ZNF85*
*ZNF93*
*ZPLD1*
*LILRB3*
*LIME1*
*LIPG*
*LOC102724159*
*LOX*
*LOXL2*
*LRRC4*
*LSAMP*
*LUM*
*LY6G5B*
*MAD2L1*
*MAMLD1*
*MAP1B*
*MARCH4*
*MASTL*
*MATN3*
*MATR3*
*MCM2*
*MCM5*
*MCM6*
*MEIOC*
*MELK*
*MFNG*
*MGLL*
*MICAL2*
*MIR17HG*
*MIR25*
*MIS18BP1*
*MKI67*
*MMP1*
*MMP10*
*MMP13*
*MMP2*
*MMP23B*
*MMP7*
*MND1*
*MPP3*
*MSH2*
*MSH5*
*MTBP*
*MTND2P28*
*MTND4P12*
*MUSK*
*MYBL1*
*MYPN*
*NASP*
*NAV3*
*NCAPD2*
*NCAPD3*
*NCAPG*
*NCAPG2*
*NCAPH*
*NCBP2-AS1*
*NDC80*
*NEK2*
*RAB11FIP4*
*RAET1E*
*RAET1G*
*RARG*
*RASSF10*
*RASSF5*
*RBM8B*
*RBP4*
*RHOBTB1*
*RHOV*
*RNASE1*
*RNF122*
*RPL23AP77*
*RPL39P5*
*RPS20P33*
*RPS6KA5*
*RSAD2*
*RTN4RL1*
*S100A7*
*S100A8*
*S100P*
*S1PR3*
*SCAMP5*
*SCARB1*
*SCD*
*SCEL*
*SEMA3E*
*SEMA3F*
*SEMA5A*
*SEMA6A*
*SERPINB13*
*SERPINB2*
*SERPING1*
*SH2B2*
*SH3BGRL*
*SLC25A23*
*SLC2A5*
*SLC6A2*
*SLC7A7*
*SLC7A8*
*SLPI*
*SMAD6*
*SNN*
*SOCS2*
*SOHLH2*
*SOX18*
*SPEF1*
*SPON2*
*SREBF1*
*ST14*
*STEAP4*
*STK32A*
*SUSD2*
*TACSTD2*
*TBX6*

*NEURL1*
*NFATC2*
*NGF*
*NGFR*
*NLRP10*
*NLRP7*
*NOG*
*NOP2*
*NPNT*
*NRG1*
*NT5E*
*NTRK2*
*NUF2*
*NUP37*
*NUSAP1*
*ODC1*
*ODF2*
*OIP5*
*OLFML2A*
*OXTR*
*P4HA3*
*PAM16*
*PARP1*
*PCNA*
*PDE10A*
*PDSS1*
*PFKFB4*
*PHKG1*
*PIF1*
*PIK3R2*
*PILRB*
*PKM*
*PKMYT1*
*PLA2G12AP1*
*PLCB4*
*PLK1*
*PLK4*
*PML*
*POLA1*
*POLA2*
*POLD1*
*POLE*
*POLE2*
*POLR3G*
*PPIEL*
*PRC1*
*PRIM1*
*PRIM2*
*PRKCA*
*PRSS41*
*PSMD12P*
*PSRC1*
*PTGS2*
*PTMA*
*PTN*
*TCAP*
*TFDP3*
*TGM1*
*THEMIS2*
*TLR1*
*TLR6*
*TMEM178A*
*TMPRSS2*
*TMSB4Y*
*TNFRSF18*
*TNFSF10*
*TNFSF9*
*TPD52L1*
*TREM2*
*TRIM22*
*TSPY1*
*TSPY3*
*TUSC1*
*U2AF1*
*UBA7*
*UCN*
*UNC5B*
*UQCRHL*
*USP6*
*VAV3*
*VGLL1*
*VTCN1*
*WISP1*
*WISP3*
*WNT10A*
*WNT10B*
*WNT11*
*WNT4*
*ZC3H11A*
*ZMYND15*
*ZNF219*
*ZNF385A*
*ZNF750*

*PTTG1*
*PWP2*
*PYCARD*
*PYGO1*
*RAB3B*
*RACGAP1*
*RAD54B*
*RAD54L*
*RBBP7*
*RBM14-RBM4*
*RBM24*
*RCC1*
*RECQL*
*RECQL4*
*RFC2*
*RFC3*
*RFC5*
*RPA3*
*RPL10P6*
*RPL14P1*
*RPL23AP64*
*RPL36A*
*RPS10*
*RPS15AP38*
*RPS24P13*
*RPS29P16*
*RPS6P8*
*RPSAP52*
*RTEL1*
*RYR3*
*S1PR1*
*SBK3*
*SBSPON*
*SCG2*
*SEMA3D*
*SEMA7A*
*SERPINB2*
*SERPINE1*
*SERPINF1*
*SETSIP*
*SF3A3P1*
*SKA1*
*SKA2*
*SKA3*
*SLC1A2*
*SLC25A5*
*SLIT2*
*SMARCE1*
*SMC1A*
*SMC1B*
*SMC2*
*SMC4*
*SMG1P2*
*SMG1P6*
*SMG1P7*

*SMOC2*
*SOCS2*
*SPAG5*
*SPARC*
*SPC25*
*SPDL1*
*SPRY4*
*SRPX*
*SRPX2*
*STAMBPL1*
*SUGT1P4-STRA6LP*
*TACC3*
*TBX3*
*TEX15*
*TGFB3*
*TGFBR2*
*THBS1*
*THBS4*
*TMC8*
*TNF*
*TNFAIP3*
*TNFRSF11A*
*TNFRSF11B*
*TNFRSF19*
*TNFRSF21*
*TNFSF10*
*TOP2A*
*TPX2*
*TRIP13*
*TTK*
*TUBA1A*
*TUBB*
*TUBB2B*
*TUBB4B*
*UBAP1L*
*UBB*
*UBXN8*
*VWA1*
*WNK4*
*WNT5A*
*WWOX*
*XRCC2*
*ZEB2*
*ZNF239*
*ZNF365*
*ZW10*
*ZWINT*

**Baseline upregulated genes in KDM3C SNP-S464T vs WT-S464 isogenic models**

| RCM-1 | CAL-27 | SCC-9 |
|---|---|---|
| *AAK1* | *ABCA1* | *A1BG* |
| *ABCB1* | *ABCA13* | *AC015813.5* |
| *AC012073.1* | *ABCG1* | *AC022532.1* |
| *AC025198.1* | *ABI3BP* | *AC068946.1* |
| *AC068491.4* | *ABTB1* | *AC084262.2* |
| *AC080023.2* | *AC007114.1* | *AC084337.1* |
| *AC097512.1* | *AC007688.1* | *AC093690.1* |
| *AC103810.1* | *AC012158.1* | *AC105245.1* |
| *AC132812.1* | *AC022075.1* | *AC106865.1* |
| *AC133644.3* | *AC025580.1* | *AC116913.1* |
| *ACAD11* | *AC026333.3* | *AC135983.2* |
| *ACVR1C* | *AC078883.1* | *AC239859.6* |
| *ADAMTS10* | *AC079250.1* | *AC245595.1* |
| *ADAMTS14* | *ACKR3* | *ACADL* |
| *ADAMTS9* | *ACP5* | *ACHE* |
| *ADAMTSL2* | *ACPP* | *ADAM12* |
| *ADPRH* | *ACSM3* | *ADAMTS12* |
| *ADRA2A* | *ADA* | *ADAMTS3* |
| *ADRB2* | *ADAMTS14* | *ADAMTS5* |
| *ADSSL1* | *ADAMTSL4* | *ADAMTS7* |
| *AGAP5* | *ADGRV1* | *ADAMTSL1* |
| *AGBL2* | *AGO4* | *ADARB2* |
| *AHRR* | *AGR2* | *ADGRL3* |
| *AKNA* | *AGT* | *ADM2* |
| *AL022324.2* | *AGXT* | *AGAP5* |
| *AL031281.2* | *AK7* | *AKAP12* |
| *AL080250.1* | *AKAP6* | *AKAP5* |
| *AL137782.1* | *AKAP7* | *AL355472.3* |
| *AL390719.1* | *AKR1C1* | *AL663070.1* |
| *AL391832.2* | *AKR1C2* | *ALPK2* |
| *ALOX5* | *AL080250.1* | *ANGPTL4* |
| *ANKRD1* | *AL096870.2* | *ANK2* |
| *AOC3* | *AL133517.1* | *ANKRD1* |
| *AP001992.1* | *AL391832.2* | *ANKRD53* |
| *AP4S1* | *ALAS2* | *AP000866.6* |
| *APH1B* | *ALDOC* | *AP1S2* |
| *APOBEC3B* | *ALOX12* | *ARFGEF3* |
| *APOD* | *ALOX15* | *ARHGAP22* |
| *APOE* | *ALOX5AP* | *ATF3* |
| *ARC* | *ANG* | *ATP2A3* |
| *ARHGAP24* | *ANGPT1* | *ATP6V1G2* |
| *ARHGAP27P1-BPTFP1-KPN* [illegible] | *ANK3* | *BCKDHA* |
| *ARHGAP4* | *APH1B* | *BCL2* |
| *ARHGAP8* | *APOBEC3A* | *BEX4* |
| *ARHGEF10* | *APOL1* | *BLK* |
| *ARL11* | *AREG* | *BMS1P2* |
| *ASB14* | *ARHGEF10L* | *BMS1P7* |
| *ASB2* | *ARHGEF37* | *BRSK1* |
| *ASB9* | *ARMCX1* | *C15orf48* |
| *ASNS* | *ARSD* | *CAMK1* |

*ATP6*
*ATP8*
*ATP8A1*
*BCAM*
*BEX3*
*BHLHA15*
*BHLHE41*
*BICC1*
*BLNK*
*BRD7P5*
*BRSK1*
*C3*
*C5AR2*
*CA2*
*CAPN8*
*CAVIN2*
*CCDC14*
*CCDC187*
*CCDC88B*
*CCL26*
*CCND2*
*CD8B*
*CDHR1*
*CDK5RAP3*
*CENPJ*
*CEP126*
*CEP295NL*
*CFAP44*
*CFH*
*CFL1P5*
*CHN1*
*CHRM3*
*CNGA1*
*COL1A1*
*COX1*
*COX2*
*COX3*
*CREB3L1*
*CSF1*
*CXCL8*
*CYP24A1*
*CYP2D6*
*CYTB*
*DAB2*
*DACH1*
*DAPK2*
*DCLK1*
*DDO*
*DDX11L2*
*DHPS*
*DHRS9*
*DNAH2*
*DOCK11*
*DRD4*
*ASB2*
*ASS1*
*ATOH8*
*ATP2A3*
*ATP6V1B1*
*BAIAP3*
*BATF2*
*BBC3*
*BBS12*
*BCL11A*
*BCL2*
*BCL2L1*
*BCL2L14*
*BCL6*
*BDNF-AS*
*BGN*
*BHLHE40*
*BHLHE41*
*BLNK*
*BMF*
*BMP6*
*BMPR1B*
*BMT2*
*BTC*
*BTG2*
*BTN3A3*
*C15orf48*
*C17orf113*
*C1R*
*C1RL*
*C1S*
*C3*
*C4A*
*C4B*
*C5*
*C7orf31*
*CA9*
*CABYR*
*CALCOCO1*
*CALML3*
*CAMK1D*
*CARD16*
*CASP5*
*CCL2*
*CCL20*
*CCL22*
*CCL5*
*CCNG2*
*CD14*
*CD34*
*CD69*
*CD70*
*CDH23*
*CDKN1A*
*CAPN5*
*CARD11*
*CARD18*
*CD34*
*CD74*
*CDH11*
*CDH2*
*CDHR5*
*CDYL2*
*CELF4*
*CLDN18*
*CLGN*
*CMTM8*
*CNTFR*
*COL10A1*
*COL13A1*
*COL4A4*
*COL5A2*
*COL8A1*
*CPEB4*
*CREB5*
*CRYAB*
*CSF2*
*CSF3*
*CXCL2*
*CXCL3*
*CXCL8*
*CYGB*
*DACT3*
*DCHS1*
*DCLK2*
*DISP1*
*DMTN*
*DNAH7*
*DOCK10*
*DOK7*
*DUSP5*
*EIF3C*
*ELMOD1*
*ENHO*
*EPHA6*
*EREG*
*ESM1*
*ETV1*
*ETV5*
*EVI2B*
*EYA4*
*FAT4*
*FBLN7*
*FBN1*
*FBXO39*
*FGF1*
*FGF22*
*FLI1*

| | | |
|---|---|---|
| *DUOX2* | *CDKN2B* | *FOXC2* |
| *DYNC1I1* | *CEACAM1* | *GAB2* |
| *EFEMP2* | *CEBPB* | *GDNF* |
| *EFNA5* | *CEBPD* | *GDPD5* |
| *EGFL8* | *CEL* | *GK* |
| *EIF4A1* | *CERCAM* | *GNG2* |
| *EIF4E3* | *CERS3* | *GPAT3* |
| *ENG* | *CFB* | *GPR173* |
| *ENPEP* | *CFH* | *GPR3* |
| *EPHB6* | *CFI* | *GSTM3* |
| *EYA4* | *CHD4* | *HAAO* |
| *FAR2P1* | *CHD5* | *HAS2* |
| *FARP1* | *CIB2* | *HBEGF* |
| *FBXL2* | *CKB* | *HHIP* |
| *FERMT2* | *CLDN1* | *HIST1H1C* |
| *FFAR4* | *CLDN16* | *HIST1H4H* |
| *FGD3* | *CLDN7* | *HLA-DOB* |
| *FGF13* | *CLEC7A* | *HOXC6* |
| *FGR* | *CMPK2* | *IFNWP19* |
| *FLRT1* | *CMTM8* | *IGFBP1* |
| *FOXA1* | *COL21A1* | *IL23A* |
| *FOXJ1* | *COL6A2* | *IL24* |
| *FOXP1* | *COL8A2* | *IL6* |
| *FOXP2* | *COL9A2* | *IMPG2* |
| *FRAS1* | *CP* | *INHA* |
| *GABBR1* | *CPE* | *INHBE* |
| *GALC* | *CPHL1P* | *INO80D* |
| *GAS6-AS1* | *CPSF1P1* | *IRAK3* |
| *GATA2* | *CR1L* | *ITGB3* |
| *GBP2* | *CREBRF* | *JCAD* |
| *GNAI1* | *CSF1* | *KBTBD8* |
| *GOLGA8A* | *CSF1R* | *KCNH1* |
| *GOLGA8B* | *CSF3* | *KIRREL3* |
| *GPC2* | *CTF1* | *KIT* |
| *GPR89B* | *CTSB* | *KLF2* |
| *GPX3* | *CTSF* | *KLHL35* |
| *GPX8* | *CTSO* | *LCP1* |
| *GSTM2* | *CX3CL1* | *LEF1* |
| *GVINP1* | *CXCL1* | *LIF* |
| *H2AFY2* | *CXCL14* | *LINC00327* |
| *HAUS7* | *CXCL17* | *LPXN* |
| *HDAC10* | *CXCL3* | *LRRN1* |
| *HDAC9* | *CXCL8* | *LZTS1* |
| *HECW2* | *CYP24A1* | *MAGEA2* |
| *HES7* | *DAPK3* | *MAGEC2* |
| *HFM1* | *DBH* | *MAP1B* |
| *HIPK2* | *DBP* | *MAP2K6* |
| *HIST1H4L* | *DDIT4* | *MCAM* |
| *HLA-H* | *DDX60* | *MCOLN2* |
| *HOXC13* | *DEC1* | *MEX3B* |
| *HSF4* | *DEFB1* | *MIR221* |
| *HSPD1P11* | *DEPP1* | *MOCS1* |
| *HYLS1* | *DEPTOR* | *MPP1* |
| *IDUA* | *DHRS3* | *MPP2* |

IL11RA
IL12A
IL1RL2
IQGAP2
ITGA7
IZUMO1
JCAD
JMJD7-PLA2G4B
KATNAL2
KLF12
KLHL31
KLHL5
KLK11
KLK13
KLK5
L3MBTL3
LCN2
LIX1L
LOXL4
LTB4R2
LTBP3
LY6G5B
MAGI2
MAMSTR
MAPK15
MAST4
MATR3
MDFIC
MEIS2
MEX3A
MIR17HG
MIR196A1
MIR200A
MIR25
MIR31HG
MITF
MLPH
MMP7
MRPL38
MSH5
MSS51
MT-ATP6
MT-ATP8
MT-CO1
MT-CO2
MT-CO3
MT-CYB
MT-ND1
MT-ND2
MT-ND3
MT-ND4
MT-ND4L
MT-ND5
MT-ND6
DHRS9
DHX58
DLK2
DLL3
DNAH3
DQX1
DUOX2
DUSP1
DUSP5
DUSP5P1
DUSP9
EBI3
ECM2
ECT2L
EDARADD
EDN2
EGLN3
EGR2
EGR3
EGR4
EIF4E3
ELMO1
ELOVL3
EPHA3
EPHB6
ESRRG
FAM131B
FBLN1
FBLN5
FBLN7
FCGRT
FGD2
FGF2
FGFR3
FKBP10
FOS
FOXI1
FOXO1
FOXO4
FRK
FUZ
FYB1
FYN
GAA
GAMT
GBP1P1
GBP2
GBP4
GBP5
GCNT3
GDF15
GFI1
GGT6
GLUL
MPP4
MRAP2
MSRA
MYEF2
MYOM1
NDNF
NECAB2
NEO1
NEURL1B
NGF
NID1
NPIPB4
NR2F1
NR4A1
NRG2
NRP1
NTM
PAG1
PAK3
PALM2
PAPSS2
PAQR5
PARM1
PCDH10
PCDHGA7
PCOLCE2
PDE1C
PDE2A
PDGFRA
PGF
PLA2G4D
PLCB4
PNPLA7
POSTN
POU5F1
PPP2R2C
PREX1
PRKAA2
PRKAR2B
PRKCA
PRKD1
PROX2
PTGER1
PTPRO
PTX3
PYGO1
QRFPR
RAB36
RAB40A
RBFOX3
RBMS3
RERG
RGS4
RNF182

*MTATP6P1*
*MTCO1P12*
*MTND2P28*
*MUC2*
*MUC4*
*MYEF2*
*MYLK2*
*NAIP*
*NAV2*
*ND1*
*ND2*
*ND3*
*ND4*
*ND5*
*ND6*
*NFATC4*
*NFKBIZ*
*NLGN3*
*NOL12*
*NPFFR1*
*NPHP3*
*NPIPB5*
*NRP2*
*NTN4*
*NTNG2*
*NYNRIN*
*OBSCN*
*OGDHL*
*OGT*
*P2RY1*
*PABPC1L*
*PALM2*
*PAX5*
*PDE4D*
*PDGFRB*
*PDK4*
*PDZRN3*
*PEAR1*
*PELI2*
*PID1*
*PILRB*
*PLA1A*
*PLA2G1B*
*PLA2G4A*
*PLA2G7*
*PLCE1*
*PLCL1*
*PLCL2*
*PLSCR4*
*PLXNA2*
*PLXNB3*
*PLXNC1*
*PNPLA7*
*POLI*
*GNMT*
*GPLD1*
*GPNMB*
*GPR20*
*GPSM3*
*GPX7*
*GRM5*
*GSN*
*H1F0*
*HAPLN3*
*HBP1*
*HCAR2*
*HCAR3*
*HEG1*
*HERC2P3*
*HIC1*
*HIST1H1C*
*HIST1H2AC*
*HIST1H2BD*
*HIST1H3E*
*HIST2H2BE*
*HIST2H2BF*
*HIST3H2BB*
*HLA-DPA1*
*HLA-DRB5*
*HLA-F*
*HLA-J*
*HMGA2*
*HMGCL*
*HMOX1*
*HOMEZ*
*HRASLS2*
*HSH2D*
*HTRA1*
*ICAM1*
*ICOSLG*
*IDO1*
*IFI27*
*IFI27L2*
*IFI44L*
*IFI6*
*IFIT1*
*IFIT2*
*IFIT3*
*IFITM1*
*IFNE*
*IFT140*
*IL11*
*IL13RA2*
*IL17C*
*IL17D*
*IL17RE*
*IL1RL1*
*IL20RA*
*RPL12P14*
*RPL23AP81*
*RPL24P2*
*RPL29P11*
*RPL37P6*
*RPLP0P2*
*RPSAP52*
*SACS*
*SARM1*
*SBSPON*
*SCG2*
*SCG5*
*SDCBP2*
*SEC31B*
*SEMA7A*
*SHANK2*
*SHISA2*
*SLAMF7*
*SLC7A11*
*SLC7A2*
*SLC8A1*
*SLIT3*
*SMG1P4*
*SMG1P6*
*SMOC1*
*SPATA18*
*SPIN2B*
*SPNS2*
*SPRED3*
*SPRY4*
*SPTB*
*SRGN*
*SRPK3*
*SRPX*
*STC2*
*SULT1A1*
*SUSD5*
*SVEP1*
*SYCP2*
*SYNE1*
*SYNE3*
*SYT11*
*TACR1*
*TENM1*
*TENM4*
*TEX15*
*TFAP2E*
*TLN2*
*TLX2*
*TNFRSF10D*
*TNFRSF11B*
*TNFRSF19*
*TNIP3*
*TREM1*

*POLR2J3*
*PPP1R9A*
*PRDM5*
*PREX1*
*PRKACB*
*PRKCG*
*PRKG1*
*PRSS1*
*PTGER1*
*PTGES*
*PTPN13*
*PTPN7*
*PTPRO*
*PYGM*
*RAB37*
*RASGEF1A*
*RASGRP3*
*RBP4*
*RIPK3*
*ROBO1*
*RPL10P13*
*RPL10P19*
*RPL32P3*
*RPL35P5*
*RPL36A*
*RPS20P33*
*RPS6KA6*
*RTN4RL2*
*RUNDC3A*
*SALL4*
*SARDH*
*SATB1*
*SBK3*
*SCART1*
*SCEL*
*SCG5*
*SEC31B*
*SEMA3A*
*SEMA3D*
*SEMA3E*
*SEMA3F*
*SEMA5A*
*SERPINA5*
*SHANK1*
*SIX4*
*SLC16A5*
*SLC6A7*
*SLC8A1*
*SLCO1B3*
*SMG1P4*
*SNCG*
*SOBP*
*SOX2*
*SPEG*
*IL20RB*
*IL34*
*INHBB*
*INPP5J*
*INSIG1*
*INSR*
*IPCEF1*
*IPO4*
*IRAK3*
*ISG15*
*ISG20*
*JUNB*
*KAZALD1*
*KBTBD3*
*KCNJ5*
*KCNN3*
*KIF13B*
*KIF5C*
*KLHDC7B*
*KLHL24*
*KLRC2*
*KRT17*
*KRT19*
*LAMB3*
*LCN2*
*LGALS3*
*LGALS9*
*LHX2*
*LINC00957*
*LMO2*
*LNX1*
*LOX*
*LOXL4*
*LRG1*
*LTC4S*
*LTF*
*LY6D*
*MACROD2*
*MAFB*
*MAP2K6*
*MAP3K8*
*MARCKS*
*MATN1*
*MB*
*MBL1P*
*MCF2L*
*MEOX1*
*METTL7A*
*MME*
*MMP1*
*MMP10*
*MMP11*
*MMP12*
*MMP13*
*TRO*
*TRPM2*
*TTC23L*
*TUBB2B*
*VCAN*
*WHAMMP2*
*WNT16*
*ZEB1*
*ZNF704*
*ZNF90*
*ZSWIM5*

*SPICE1*
*SPTBN5*
*SRPX*
*STRA6*
*STX1B*
*SYCP2*
*SYNPO2L*
*SYT8*
*SYTL2*
*SYTL5*
*TBX3*
*TDRD6*
*TEX22*
*THBS1*
*TIAF1*
*TLR5*
*TNFAIP2*
*TOMM5*
*TRAF1*
*TRIM31*
*TRIM67*
*TRPS1*
*TSPAN4*
*TTC28*
*TTLL10*
*TTLL3*
*TTLL7*
*UBAP1L*
*VNN1*
*WDPCP*
*WEE2-AS1*
*WNT8B*
*XDH*
*XIAPP3*
*ZBED6*
*ZC3H11A*
*ZNF675*
*MMP17*
*MMP28*
*MMP7*
*MR1*
*MTHFR*
*MUC1*
*MUC20*
*MUC4*
*MVP*
*MX1*
*MX2*
*MXD4*
*MYLIP*
*NCF1*
*NCOA7*
*NDRG1*
*NDRG2*
*NECTIN1*
*NECTIN4*
*NEDD9*
*NEURL3*
*NFAM1*
*NFASC*
*NFATC4*
*NFE2*
*NGFR*
*NKD1*
*NKX2-8*
*NLRP6*
*NMNAT3*
*NNMT*
*NOTCH3*
*NOXA1*
*NPNT*
*NR2E1*
*NR4A1*
*NR4A3*
*NRARP*
*NRROS*
*NTF4*
*NTNG2*
*NTRK2*
*NUPR1*
*OAS1*
*OAS2*
*OASL*
*OLFM4*
*ORAI3*
*P2RX7*
*PADI2*
*PADI3*
*PAOX*
*PBX1*
*PBXIP1*

*PCAT7*
*PCDHB13*
*PCDHGA2*
*PCDHGB1*
*PCDHGB2*
*PCMTD1*
*PCSK9*
*PDCD4*
*PDGFA*
*PELI2*
*PEX11G*
*PGLYRP3*
*PGLYRP4*
*PHF21A*
*PI3*
*PIGR*
*PIK3AP1*
*PIK3C2G*
*PIK3IP1*
*PIP*
*PKN1*
*PLA2G4C*
*PLA2R1*
*PLAT*
*PLD1*
*PLPP3*
*PLSCR4*
*PLXND1*
*PNRC1*
*PODNL1*
*POLR3GL*
*PPARGC1A*
*PPP1R12B*
*PRKAA2*
*PRKAR2B*
*PRXL2A*
*PSMB8-AS1*
*PTGES*
*PTGS2*
*PTP4A3*
*PTPRZ1*
*PYCARD*
*QPCT*
*RAB19*
*RAB26*
*RAB3D*
*RAB7B*
*RARB*
*RARRES2*
*RASD1*
*RASSF6*
*RCOR2*
*RET*
*RGCC*

*RGS12*
*RGS9*
*RHOV*
*RND1*
*RNF128*
*RNF144B*
*RNF152*
*RPS10P7*
*RPS23P9*
*RPS26P47*
*RSAD2*
*RYR1*
*S100A14*
*S100A7*
*S100A8*
*S100A9*
*S100P*
*SAA1*
*SASH1*
*SATB1*
*SCAMP5*
*SCD*
*SCGB3A1*
*SCO2*
*SDR16C5*
*SEMA3E*
*SEMA3F*
*SEMA4A*
*SEMA4B*
*SERPINA1*
*SERPINA5*
*SERPINB13*
*SERPINB3*
*SERPINB4*
*SERPINF2*
*SH2D3C*
*SH3PXD2B*
*SHH*
*SIDT1*
*SIPA1*
*SKOR1*
*SLC26A1*
*SLC27A1*
*SLC2A3*
*SLC52A1*
*SLPI*
*SMARCA1*
*SMOC2*
*SNCAIP*
*SOCS3*
*SOD2*
*SOX18*
*SPATA18*
*SPDEF*

*SREBF1*
*SSBP2*
*SSC5D*
*ST6GAL1*
*STC1*
*SUSD4*
*SYNPO*
*TCIM*
*TET1*
*TFPI*
*TIMP3*
*TJP3*
*TLL2*
*TLR5*
*TLR6*
*TM7SF2*
*TMEM135*
*TMEM173*
*TMEM176B*
*TMEM8B*
*TMPRSS2*
*TNFRSF10B*
*TNFSF10*
*TNFSF15*
*TNFSF4*
*TNIP3*
*TP53AIP1*
*TP53INP1*
*TP53INP2*
*TP73*
*TREM1*
*TRIM22*
*TRIM29*
*TRIM31*
*TRIM69*
*TRPM8*
*TRPV4*
*TSC22D3*
*TSLP*
*TSTD1*
*TTC12*
*TTLL1*
*TYMP*
*U2AF1*
*UBASH3A*
*UFSP1*
*UNC5B*
*VAV3*
*VEGFA*
*VNN3*
*VSIR*
*VTCN1*
*WISP2*
*WNT4*

*WNT5A*
*XAF1*
*YPEL5*
*ZC3H6*
*ZFP36*
*ZNF385A*
*ZNF385C*
*ZNF750*
*ZP3*

**Overlap of baseline differentially expressed genes across KDM3C SNP-S464T vs. WT-S464 isogenic m**

| | RCM-1 (total genes) | CAL-27 (total genes) | SCC-9 (total genes) |
|---|---|---|---|
| **downregulated genes** | 301 | 484 | 364 |
| **upregulated genes** | 357 | 547 | 277 |

| | number of genes only in RCM-1 | number of genes only in CAL-27 | number of genes only in SCC-9 |
|---|---|---|---|
| **downregulated genes** | 257 | 457 | 314 |
| **upregulated genes** | 308 | 495 | 241 |

| | number of genes (RCM1 + CAL27) | number of genes (RCM1 + SCC9) | number of genes (CAL27 + SCC9) |
|---|---|---|---|
| **downregulated genes** | 10 | 33 | 16 |
| **upregulated genes** | 31 | 17 | 18 |

odels

**Supplementary Table 18. Bulk RNA-sequencing results**

RCM-1 WT vs SNP enriched genes

RCM-1 enrichedgenes 24h post IR

Cal27 WT vs SNP enriched genes

Cal27 enrichedgenes 24h post IR

SCC-9 WT vs SNP enriched genes

SCC-9 enrichedgenes 24h post IR

Analysis of enriched genes from the comparison between RCM-1 cells bearing KDM3C SNP-S464T vers

Analysis of enriched genes from the comparison between RCM-1 cells bearing KDM3C SNP-S464T vers

Analysis of enriched genes from the comparison between CAL-27 cells bearing *KDM3C* SNP-S464T ve

Analysis of enriched genes from the comparison between CAL-27 cells bearing KDM3C SNP-S464T ver

Analysis of enriched genes from the comparison between SCC-9 cells bearing KDM3C SNP-S464T vers

Analysis of enriched genes from the comparison between SCC-9 cells bearing KDM3C SNP-S464T vers sus  KDM3C WT-S464 allele at baseline

sus KDM3C WT-S464 allele 24h after 2Gy IR

rsus *KDM3C* WT-S464 allele at baseline

rsus KDM3C WT-S464 24hrs after 2Gy IR

sus KDM3C WT-S464 allele at baseline

sus KDM3C WT-S464 allele 24h after 2Gy IR

**Analysis of enriched genes from the comparison between RC**

**Downregulated Genes**

| **Term** | **Description** |
|---|---|
| R-HSA-5334118 | DNA methylation |
| R-HSA-3214815 | HDACs deacetylate histones |
| GO:0009986 | cell surface |
| GO:0000786 | nucleosome |
| GO:0006954 | inflammatory response |
| GO:0044815 | DNA packaging complex |
| R-HSA-2559582 | Senescence-Associated Secretory Phenotype (SASP) |
| hsa04060 | cytokine-cytokine receptor interaction |
| GO:0004896 | cytokine receptor activity |
| GO:0098742 | cell-cell adhesion via plasma-membrane adhesion molecules |
| GO:0032993 | protein-DNA complex |
| GO:0005125 | cytokine activity |

**Upregulated Genes**

| **Term** | **Description** |
|---|---|
| GO:0043062 | extracellular structure organization |
| hsa00190 | oxidative phosphorylation |
| GO:0043269 | regulation of ion transport |
| GO:0006820 | anion transport |
| GO:0099537 | trans-synaptic signaling |
| GO:0072593 | reactive oxygen species metabolic process |
| GO:0005746 | mitochondrial respiratory chain |
| GO:0008236 | serine-type peptidase activity |
| R-HSA-909733 | Interferon alpha/beta signaling |
| R-HSA-1474244 | Extracellular matrix organization |
| GO:0005125 | cytokine activity |
| GO:0001077 | transcriptional activator activity, RNA polymerase II proximal promoter sequence-specific DNA binding |
| GO:0050840 | extracellular matrix binding |

**:M-1 cells bearing KDM3C SNP-S464T versus KDM3C WT-S464 allele at baseline**

| Genes | P value | FDR |
|---|---|---|
| *HIST1H3H* | 3.87E-06 | 0.000424632 |
| *TBL1X, HIST1H3H* | 5.45E-06 | 0.000433978 |
| *ABCC2, ANPEP, BMP2, CD24, CDH11, CDH3, CXCR3, GPC4, HLA-DPA1, HLA-DRA, LRP4, MICB, MSN, NOD2, SULF2, WNT7A* | 1.30E-06 | 0.000497946 |
| *HIST1H3H* | 2.22E-05 | 0.002025249 |
| *AGT, BMP2, CAMK1D, CCL5, CXCR3, CYP26B1, GJA1, KRT16, NOD2, PPBP, SPHK1* | 1.54E-05 | 0.003235416 |
| *HIST1H3H* | 5.03E-05 | 0.003673513 |
| *HIST1H3H* | 0.00012952 | 0.003705128 |
| *BMP2, CCL5, CXCR3, IL27RA, PPBP* | 8.90E-05 | 0.008126457 |
| *CXCR3, IL27RA* | 0.00026222 | 0.009737987 |
| *BMP2,CDH11, CDH16, CDH3, DSC3, DSG3, FLRT3, PCDH7, PCDHB2* | 6.5229E-05 | 0.014950796 |
| *HIST1H3H* | 0.0004243 | 0.017586223 |
| *BMP2, CCL5, PPBP, WNT7A* | 0.00040088 | 0.037638058 |

| Genes | P value | FDR |
|---|---|---|
| *ENG, LOXL4, KLK5, APOE, PRSS1, CREB3L1, ITGA7, ADAMTS14, PLA2G7, COL1A1* | 6.01E-07 | 0.00067432 |
| *MTND2P28, MT-ND2, MT-ND1, MT-ND4L, MT-ND4, MT-CYB, MT-ATP6, MTATP6P1, MT-ND3, MT-CO1, MTCO1P12, MT-CO3* | 2.89E-05 | 0.002636443 |
| *ADRA2A, ADRB2, APOE, ARC, ATP8A1, CA2, DRD4, GPR89B, PDGFRB, PLA2G1B, SLC8A1, SNCG* | 6.78E-06 | 0.004229839 |
| *PLA2G1B, APOE, STRA6, ATP8A1, SLC6A7, CA2, DRD4, SLC16A5, PLA2G4A, SLCO1B3, GPR89B* | 2.00E-05 | 0.006405144 |
| *SYTL5, SYTL2, APOE, ADRB2, CA2, PPP1R9A, NFATC4, DRD4, SYT8, SNCG, PLCL1, ARC, CHRM3* | 5.13E-05 | 0.011605265 |
| *GPX3, MT-ND2, DUOX2, PDK4, PDGFRB* | 7.30E-05 | 0.013111569 |
| *MT-ND2, MT-ND1, MT-ND4L, MT-ND4, MT-CYB, MT-ND3, MT-CO1, MT-CO3* | 0.00023868 | 0.018664877 |
| *KLK5, PRSS1, C3, KLK11, KLK13* | 0.00028364 | 0.030609969 |
| *GBP2* | 0.00012758 | 0.034787821 |
| *LOXL4, PRSS1, LTBP3, ITGA7, ADAMTS14, CAPN8, NTN4, COL1A1* | 0.00014428 | 0.034787821 |
| *CXCL8, CCL26, CSF1* | 0.00066958 | 0.036330215 |
| *GATA2, FOXJ1, CREB3L1, FOXA1, SIX4, MITF, AKNA, HOXC13, MEIS2* | 0.00084489 | 0.036330215 |
| *BCAM, NTN4* | 0.00143512 | 0.048294865 |

**negative Log10**

| |
|---|
| 3.371987113 |
| 3.362532348 |
| 3.30281773 |
| 2.693521538 |
| 2.490069932 |
| 2.43491846 |
| 2.431196741 |
| 2.090098774 |
| 2.011530832 |
| 1.825335699 |
| 1.754827417 |
| 1.424372797 |

**negative Log10**

| |
|---|
| 3.171133959 |
| 2.578981644 |
| 2.373676163 |
| 2.193471102 |
| 1.935344938 |
| 1.882345335 |
| 1.728974864 |
| 1.51413711 |
| 1.458572779 |
| 1.458572779 |
| 1.439732032 |
| 1.439732032 |
| 1.316099044 |

**Analysis of enriched genes from the comparison between RC**

**Downregulated Genes**

| Term | Description |
|---|---|
| R-HSA-5334118 | DNA methylation |
| R-HSA-3214815 | HDACs deacetylate histones |
| GO:0009986 | cell surface |
| GO:0000786 | nucleosome |
| GO:0044815 | DNA packaging complex |
| GO:0005216 | ion channel activity |
| R-HSA-2559582 | Senescence-Associated Secretory Phenotype (SASP) |
| R-HSA-73864 | RNA Polymerase I Transcription |
| GO:0008083 | growth factor activity |
| GO:0005509 | calcium ion binding |
| R-HSA-380108 | Chemokine receptors bind chemokines |
| GO:0005262 | calcium channel activity |
| hsa00190 | oxidative phosphorylation |
| GO:0098742 | cell-cell adhesion via plasma-membrane adhesion molecules |
| GO:0000788 | nuclear nucleosome |
| GO:0032993 | protein-DNA complex |
| GO:0005746 | mitochondrial respiratory chain |

**Upregulated Genes**

| Term | Description |
|---|---|
| GO:0043062 | extracellular structure organization |
| GO:0045814 | negative regulation of gene expression, epigenetic |
| GO:0045596 | negative regulation of cell differentiation |
| GO:0006342 | chromatin silencing |
| GO:0006959 | humoral immune response |
| GO:0032993 | protein-DNA complex |
| GO:0008083 | growth factor activity |
| GO:0005125 | cytokine activity |

**:M-1 cells bearing KDM3C SNP-S464T versus KDM3C WT-S464 allele 24h after 2Gy IR**

| Genes | P value | FDR |
|---|---|---|
| *HIST1H3E* | 5.49E-07 | 0.000140314 |
| *HIST1H3E* | 5.96E-07 | 0.000140314 |
| *CXCR3, HLA-DRA, HLA-DPA1, ACE2, CDH3, CD24, SULF2, CDH11, LRP4, CYP2W1, ABCC2, MUC17, ANPEP, VAMP1, NOD2, MSN, ITGAE, BTNL8, CD36* | 9.65E-07 | 0.000412891 |
| *HIST1H3E* | 1.95E-06 | 0.000418254 |
| *HIST1H3E* | 5.97E-06 | 0.000601056 |
| *SHROOM2, ITPR2, CACNA1C, KCNMA1, HCN3, PDE2A, CACNA1D* | 1.27E-05 | 0.002201425 |
| *HIST1H3E* | 9.78E-05 | 0.002597568 |
| *HIST1H3E* | 0.0001352 | 0.00307918 |
| *VGF, CLEC11A, PPBP, AGT* | 7.72E-05 | 0.00890919 |
| *DSG3, NELL2, PCDH7, DSC3, CDH16, PCDHB2, CDH11, SULF2, CDH3, ITPR2, ANXA13, LRP4, DLL1, EGFLAM, VSNL1, PADI1, C2CD4A, TCHH* | 0.00017535 | 0.017335086 |
| *CXCR3, PPBP* | 0.00186542 | 0.025823239 |
| *ITPR2, CACNA1C, PDE2A, CACNA1D* | 0.00075269 | 0.027413631 |
| *NDUFB1* | 0.00014396 | 0.030215614 |
| *DSG3, PCDH7, DSC3, CDH16, PCDHB2, CDH11, CDH3, TENM1, HIST1H3E* | 2.06E-05 | 0.030563642 |
| *HIST1H3E* | 0.00073356 | 0.034884944 |
| *HIST1H3E* | 0.00084663 | 0.035055948 |
| *NDUFB1* | 0.00034993 | 0.035885239 |

| Genes | P value | FDR |
|---|---|---|
| *ADAMTS9, BCAN, ENG, KLK5, LOXL4, PLA2G7, PRKACB, PRSS1, THSD4* | 2.91E-05 | 4.82E-03 |
| *HIST1H2AH* | 4.52E-05 | 0.006327478 |
| *FGF13, FOXA1, FOXJ1, ZNF675, GATA2, SEMA5A, SIX4, SOX2, INPP5J, EFNA5, ARHGAP4* | 9.91E-05 | 0.011614249 |
| *HIST1H2AH* | 5.61E-06 | 0.001158052 |
| *PLA2G1B, LCN2, FOXJ1, KLK5, CFH, C3* | 0.00022068 | 0.019934408 |
| *HIST1H2AH* | 1.02E-10 | 1.19E-08 |
| *FGF13, INHBE* | 0.00019227 | 0.011939876 |
| *INHBE* | 0.00075616 | 0.026393573 |

| negative Log10 |
|---|
| 3.85289934 |
| 3.85289934 |
| 3.384164233 |
| 3.378559586 |
| 3.221084845 |
| 2.657296204 |
| 2.585433029 |
| 2.511564938 |
| 2.050161794 |
| 1.761074009 |
| 1.587989279 |
| 1.562033432 |
| 1.519768575 |
| 1.514794897 |
| 1.45736197 |
| 1.455238282 |
| 1.445084155 |

| negative Log10 |
|---|
| 2.317297645 |
| 2.198769346 |
| 1.935008869 |
| 2.936271896 |
| 1.700396664 |
| 7.92546706 |
| 1.923000199 |
| 1.578501808 |

**Analysis of enriched genes from the comparison betweer**

**Downregulated Genes**

| **Term** | **Description** |
| --- | --- |
| GO:0051383 | kinetochore organization |
| GO:0022616 | DNA strand elongation |
| GO:0050000 | chromosome localization |
| GO:0007059 | chromosome segregation |
| GO:0071103 | DNA conformation change |
| GO:0042769 | DNA damage response, detection of DNA damage |
| GO:0034502 | protein localization to chromosome |
| GO:0006301 | postreplication repair |
| GO:0051653 | spindle localization |
| GO:0044839 | cell cycle G2/M phase transition |

| | |
|---|---|
| GO:0000075 | cell cycle checkpoint |
| GO:0006298 | mismatch repair |
| GO:0006260 | DNA replication |
| GO:0006333 | chromatin assembly or disassembly |
| GO:0006338 | chromatin remodeling |
| GO:0097191 | extrinsic apoptotic signaling pathway |
| GO:0051302 | regulation of cell division |
| GO:0006302 | double-strand break repair |
| GO:0006289 | nucleotide-excision repair |
| GO:0044843 | cell cycle G1/S phase transition |
| GO:0006310 | DNA recombination |

| | |
|---|---|
| GO:0043062 | extracellular structure organization |
| GO:0001525 | angiogenesis |
| GO:0042770 | signal transduction in response to DNA damage |
| GO:0006959 | humoral immune response |
| GO:0032635 | interleukin-6 production |
| GO:0070371 | ERK1 and ERK2 cascade |
| GO:0070555 | response to interleukin-1 |

**Upregulated Genes**

| **Term** | **Description** |
|---|---|
| GO:0032633 | interleukin-4 production |
| GO:0010463 | mesenchymal cell proliferation |
| GO:0032602 | chemokine production |
| GO:0097191 | extrinsic apoptotic signaling pathway |
| GO:0043954 | cellular component maintenance |
| GO:0097696 | STAT cascade |
| GO:0006333 | chromatin assembly or disassembly |
| GO:0050866 | negative regulation of cell activation |
| GO:0006959 | humoral immune response |
| GO:0071103 | DNA conformation change |
| GO:2001233 | regulation of apoptotic signaling pathway |

| | |
|---|---|
| GO:0051302 | regulation of cell division |
| GO:2000147 | positive regulation of cell motility |
| GO:0043062 | extracellular structure organization |
| GO:0043491 | protein kinase B signaling |
| GO:0070371 | ERK1 and ERK2 cascade |
| GO:0050673 | epithelial cell proliferation |
| GO:0001819 | positive regulation of cytokine production |
| GO:0001525 | angiogenesis |
| GO:0050727 | regulation of inflammatory response |
| GO:0002697 | regulation of immune effector process |
| GO:0002250 | adaptive immune response |
| GO:0002521 | leukocyte differentiation |

**n CAL-27 cells bearing KDM3C SNP-S464T versus KDM3C WT-S464 allele at baseline**

**Genes**

*CDT1, CENPA, CENPE, CENPF, CENPK, CENPN, CENPW, NDC80, NUF2, SMC2, SMC4*

*DNA2, GINS1, GINS2, GINS4, LIG1, PCNA, POLA1, POLE, RFC3*

*CCNB1, CDCA5, CDCA8, CDT1,CENPE, CENPF, DLGAP5, FAM83D, KIF14, KIF18A, KIF22, KIF2C, KIFC1, NDC80, NUF2, PSRC1, SPAG5, SPDL1, ZW10*

*ANAPC15, AURKB, BIRC5, BLM, BRCA1, BRIP1, BUB1, BUB1B, CCNB1, CDC20, CDC42, CDC6, CDCA5, CDCA8, CDT1, CENPE, CENPF, CENPN, CENPW, DLGAP5, DMC1, DSCC1, ECT2, EME1, ERCC2, ESCO2, ESPL1, FAM83D, FANCD2, FBXO5, GEN1, HASPIN, HJURP, INCENP, KIF14, KIF18A, KIF18B, KIF22, KIF23, KIF2C, KIF4A, KIFC1, KNL1, KNSTRN, MAD2L1, MEIOC, MKI67, NCAPD2, NCAPD3, NCAPG, NCAPH, NDC80, NEK2, NUF2, NUP37, NUSAP1, OIP5, PLK1, PRC1, PSRC1, PTTG1, RACGAP1, RCC1, SKA1, SKA2, SKA3, SLC25A5, SMC1A, SMC2, SMC4, SPAG5, SPC25, SPDL1, TACC3, TOP2A, TRIP13, TTK, ZW10, ZWINT*

*RAD54B, RAD54L, RBBP7, RECQL, RECQL4, RTEL1, SMC2, SMC4, TOP2A, HJURP, HMGB1, HMGB2, HMGB3, KNL1, MCM2, MCM6, MIS18BP1, NASP, NCAPD2, NCAPD3, NCAPG, NCAPG2, NCAPH, NUSAP1, OIP5, PIF1, ASF1B, BLM, BRIP1, CCNB1, CDC45,CDCA5, CDK1, CENPA, CENPI, CENPK, CENPL, CENPM, CENPN, CENPO, CENPU, CENPW, CHAF1A, CHAF1B, DNA2, ERCC2, GINS1, GINS2, GINS4, GPER1, H2AFX, HELLS, HHEX, HIST1H2BL*

*PARP1, PCNA, POLD1, RFC2, RFC3, RFC5, RPA3, UBB*

*AURKB, BUB1B, CDCA5, CDK1, CDT1, CENPA, ESCO2, HASPIN, KNL1, MSH2, MTBP, NDC80, PLK1, SPDL1, TTK, ZW10*
*BRCA1, MSH2, PCNA, POLD1, POLE2, RFC2, RFC3, RFC5, RPA3, UBB*

*ASPM, CENPA, DCTN1, ESPL1, NDC80, NUSAP1, SPDL1, ZW10*
*AURKA, AURKB, BLM, BORA, BRCA1, CCNB1, CCNB2, CCP110, CDC25C, CDC6, CDK1, CDK2, CENPF, CEP152, CEP192, CEP70, CEP72, CHEK1, CHEK2, CKAP5, DCTN1, FBXO5, HAUS1, HAUS4, HAUS8, HSP90AA1, HSPA2, KIF14, MASTL, MELK, NEK2, ODF2, PKMYT1, PLK1, PLK4, RECQL4, TPX2, TUBA1A, TUBB, TUBB4B*

*ANAPC15, AURKA, AURKB, BLM, BRCA1, BRIP1, BUB1, BUB1B, CCNB1, CDC20, CDC25C, CDC45, CDC6, CDK1, CDK2, CDT1, CENPF, CHEK1, CHEK2, DLG1, DNA2, EME1, GEN1, H2AFX, KNTC1, MAD2L1, MSH2, NDC80, PCNA, PLK1, PML, SPDL1, TOP2A, TRIP13, TTK, ZW10, ZWINT*

*HMGB1, LIG1, MSH2, PCNA, RPA3*

*BLM, BRCA1, BRIP1, CDC42, CDC45, CDC6, CDK1, CDK2, CDT1, CHAF1A, CHAF1B, CHEK1, CHEK2, DNA2, DSCC1, EME1, ESCO2, FBXO5, GEN1, GINS1, GINS2, GINS4, LIG1, MCM2, MCM5, MCM6, NASP, PCNA, PIF1, POLA1, POLA2, POLD1, POLE, POLE2, PRIM1, PRIM2, RBBP7, RECQL4, RFC2, RFC3, RFC5, RPA3, RTEL1, SMC1A*

*ASF1B, CENPA, CENPI, CENPK, CENPL, CENPM, CENPN, CENPO, CENPU, CENPW, CHAF1A, CHAF1B, H2AFX, HELLS, HIST1H2BL, HJURP, HMGB1, HMGB2, KNL1, MCM2, MIS18BP1, NASP, OIP5, RBBP7*

*ANP32E, CENPA, CENPI, CENPK, CENPL, CENPM, CENPN, CENPO, CENPU, CENPW, CHEK1, HELLS, HJURP, HMGB1, HMGB2, HMGB3, KNL1, MIS18BP1, NASP, OIP5, PTMA, RBBP7*

*AR, ATF3, BMP4, BMP5, BRCA1, CAV1, CD70, CSF2, DAPK1, EYA1, FAS, FGFR3, GPER1, HMGB2, HSPA1A, ICAM1, IL1A, IL1B, IL7, ITGAV, KITLG, PML, PYCARD, SCG2, SERPINE1, SRPX, THBS1, TMC8, TNF, TNFAIP3, TNFSF10, WWOX*

*ASPM, AURKA, AURKB, BLM, CCP110, CDC42, CDC6, ECT2, FGF1, IL1A, IL1B, INCENP, KIF14, KIF18B, KIF20B, KIF23, PLK1, PRC1, PTN, RACGAP1, TGFB3, THBS4*

*BLM, BRCA1, BRIP1, CDC45, CDCA5, CHEK1, CHEK2, DMC1, DNA2, EME1, ESCO2, EYA1, GEN1, GINS2, GINS4, H2AFX, MEIOC, MSH2, PARP1, PML, POLA1, RAD54B, RAD54L, RECQL, RECQL4, RPA3, RTEL1, TRIP13, XRCC2*

*BRIP1, ERCC2, HMGB1, LIG1, PARP1, PCNA, POLA1, POLD1, POLE, RFC2, RFC3, RFC5, RPA3, UBB*

*ADAMTS1, AURKA, CCL2, CCNB1, CDC25C, CDC45, CDC6, CDK1, CDK2, CDT1, CHEK2, FAM83D, FBXO5, KIF14, MCM2, MCM5, MCM6, MTBP, NASP, PCNA, PML, POLA1, POLA2, POLE, POLE2, PRIM1, PRIM2, RCC1, RPA3*

*BLM, BRCA1, BRIP1, CDC45, CHEK1, DMC1, EME1, ERCC2, GEN1, GINS2, GINS4, H2AFX, HMGB1, HMGB2, HMGB3, IL27RA, IL7R, LIG1, MND1, MSH2, PARP1, PIF1, RAD54B, RAD54L, RECQL, RECQL4, RPA3, RTEL1, TOP2A, TRIP13, XRCC2*

*ABI3BP, CCDC80, COL12A1, COL18A1, COL1A1, COL1A2, COL4A1, COL4A2, COL4A6, COL5A1, COL6A1, COL9A2, DAG1, EFEMP2, ENG, ERCC2, FBLN1, FBN2, FN1, HSPG2, ICAM1, ITGAV, LAMA3, LAMA4, LAMB1, LAMB2, LOX, LOXL2, LUM, MATN3, MMP1, MMP10, MMP13, MMP2, MMP7, NPNT, OLFML2A, SERPINE1, SMOC2, SPARC, THBS1, TNF, TNFRSF11B, VWA1*

*ANGPTL4, BMP4, BRCA1, CAV1, CCL2, CDC42, COL18A1, COL4A1, COL4A2, CSPG4, CXCL10, CXCL8, DAG1, EDN1, ENG, F3, FGF1, FGFBP1, FN1, HHEX, HMGB1, HSPG2, IL1A, IL1B, IL6, ITGAV, LOXL2, MMP2, NGFR, PKM, PML, PRKCA, PTGS2, PTN, SCG2, SERPINE1, SERPINF1, SLIT2, SMOC2, SPARC, SRPX2, TGFBR2, THBS1, THBS4, TNFAIP3, WNT5A*

*AURKA, BRCA1, CCNB1, CDC25C, CDK1, CDK2, CHEK1, CHEK2, GEN1, PCNA, PLK1, PML*

*CCL2, CXCL1, CXCL10, CXCL11, CXCL2, CXCL3, CXCL5, CXCL8, CXCL9, EBI3, GPI, IL36RN, IL6, IL7, MMP7, S100A9, TNF, TNFRSF21*

*HMGB1, IL1B, IL36G, IL36RN, IL6, PYCARD, TNF, TNFAIP3, WNT5A*

*ATF3, BMP2, BMP4, CCL2, CCL20, CCL5, DLG1, FAM83D, FBLN1, FGFR3, FN1, GPER1, HMGB1, ICAM1, IL1B, ITGAV, NPNT, NTRK2, PRKCA, PYCARD, TNF, TNFRSF11A*

*ADAMTS12, ADAMTS7, CCL2, CCL20, CCL5, CXCL8, EDN1, ICAM1, IL1A, IL1B, IL1R1, IL1R2, IL6, NLRP7, PRKCA, PYCARD, TNFRSF11A, UBB*

**Genes**

*CEBPB, IL20RB, ZP3*

*PDGFA, SHH, VEGFA*

*C5, CSF1R, HMOX1, IL1RL1, TSLP*

*AGT, BCL2, BCL2L1, BMPR1B, DAPK3, FYN, HMOX1, LGALS3, RET, TIMP3, TNFRSF10B, UNC5B*

*CSF1R, FYN, INSR*

*AGT, CSF1R, CTF1, FYN, IFNE, RET, TSLP, VEGFA*

*H1F0, HIST1H1C, HIST1H2BD, HIST2H2BE, HIST2H2BF, HIST3H2BB, HMGA2, IPO4*

*CEBPB, HMOX1, IL13RA2, IL20RB, LGALS3, PDGFA, PKN1,SHH*

*BCL2, C5, DEFB1, HIST1H2BD, HIST2H2BE, IFNE, PI3, S100A7, SLPI, ZP3*

*CHD4, CHD5, H1F0, HIST1H1C, HIST1H2BD, HIST2H2BE, HIST2H2BF, HIST3H2BB, HMGA2, IPO4*

*AGT, BCL2, BCL2L1, BMPR1B, DAPK3, FYN, HMOX1, IL20RA, INHBB, LGALS3, RET, TIMP3, TNFRSF10B, UNC5B*

*BCL2L1, FGF2, PDGFA, SHH, VEGFA*

*AGT, BCL2, CSF1R, DAPK3, DEFB1, FGF2, HMOX1, INSR, LGALS3, PDGFA, RET, S100A14, S100A7, VEGFA, ZP3*
*ADAMTS14, AGT, BGN, COL6A2, COL8A2, ECM2, FGF2, HTRA1, LAMB3, MATN1, MMP11, PDGFA*

*FGF2, FYN, GDF15, INSR, PDGFA, RET*

*AGT, CSF1R, INSR, PDGFA, RET, S100A7, TIMP3*

*AREG, BMP6, CEBPB, COL8A2, FGF2, HMOX1, HTRA1, SHH, VEGFA*

*AGT, C5, CEBPB, CSF1R, HMOX1, IL17D, IL1RL1, IL20RB, TSLP, ZP3*

*AGT, C5, FGF2, HMGA2, HMOX1, PDGFA, S100A7, SHH, UNC5B, VEGFA*

*AGT, C5, CEBPB, IL17D, IL1RL1, IL20RB, TSLP, ZP3*

*C5, HMGA2, HMOX1, IL13RA2, IL20RB, LGALS3, PKN1, ZP3*

*C5, FYN, IFNE, IL13RA2, IL1RL1, IL20RB, PKN1,ZP3*
*BCL2, CEBPB, CSF1R, IFNE, IL11, IL34, LGALS3, SHH, VEGFA*

| P value | FDR | negative Log10 |
|---|---|---|
| 2.29E-13 | 6.49E-12 | 11.18800297 |
| 1.54E-09 | 2.98E-08 | 7.526133644 |
| 5.55E-16 | 2.25E-14 | 13.64841626 |
| <2.2e-16 | 2.20E-16 | 15.65757732 |
| <2.2e-16 | 2.20E-16 | 15.65757732 |
| 2.3527E-06 | 0.000025639 | 4.591098918 |
| 3.65E-11 | 7.96E-10 | 9.099163322 |
| 1.98E-07 | 2.8974E-06 | 5.537991544 |
| 4.2507E-06 | 0.000044062 | 4.355935794 |
| <2.2e-16 | 2.20E-16 | 15.65757732 |

| | | |
|---|---|---|
| <2.2e-16 | 2.20E-16 | 15.65757732 |
| 0.00055069 | 0.0031627 | 2.499942001 |
| <2.2e-16 | 2.20E-16 | 15.65757732 |
| 1.23E-13 | 4.01E-12 | 11.39639017 |
| 4.08E-12 | 1.02E-10 | 9.991740585 |
| <2.2e-16 | 2.20E-16 | 15.65757732 |
| 1.18E-11 | 2.79E-10 | 9.555143613 |
| 1.89E-14 | 6.98E-13 | 12.15644956 |
| 2.18E-07 | 3.0872E-06 | 5.510435234 |
| 4.92E-13 | 1.35E-11 | 10.87011684 |
| 1.85E-13 | 5.62E-12 | 11.25065024 |

| | | |
|---|---|---|
| <2.2e-16 | 2.20E-16 | 15.65757732 |
| 2.22E-16 | 9.44E-15 | 14.02517065 |
| 0.000048589 | 0.00038599 | 3.413423947 |
| 0.00001254 | 0.00011398 | 3.943171347 |
| 0.0030856 | 0.014652 | 1.83410309 |
| 6.6304E-06 | 0.000067094 | 4.173316316 |
| 1.03E-08 | 1.74E-07 | 6.758528232 |
| **P value** | **FDR** | **negative Log10** |
| 0.0012856 | 0.018521 | 1.732335568 |
| 0.0028989 | 0.034199 | 1.465986593 |
| 0.00014409 | 0.0037115 | 2.430450535 |
| 1.40E-08 | 2.9809E-06 | 5.525652593 |
| 0.0051205 | 0.048865 | 1.311002097 |
| 4.4404E-06 | 0.0002696 | 3.569280112 |
| 6.2531E-06 | 0.00031266 | 3.504927676 |
| 0.000017915 | 0.00076139 | 3.118392831 |
| 3.0686E-06 | 0.00021736 | 3.662820375 |
| 3.4253E-06 | 0.00022396 | 3.649829541 |
| 1.35E-07 | 0.000016333 | 4.786934038 |

| | | |
|---|---|---|
| 0.0033239 | 0.037175 | 1.429749023 |
| 4.58E-07 | 0.000043279 | 4.363722783 |
| 8.2902E-06 | 0.00037088 | 3.430766586 |
| 0.0035876 | 0.039603 | 1.402271914 |
| 0.0010912 | 0.017177 | 1.765052684 |
| 0.00058721 | 0.01062 | 1.973875483 |
| 0.00031501 | 0.0065308 | 2.185033616 |
| 0.00025584 | 0.005576 | 2.253677235 |
| 0.002078 | 0.027387 | 1.562455538 |
| 0.002898 | 0.034199 | 1.465986593 |
| 0.0029448 | 0.034199 | 1.465986593 |
| 0.0042196 | 0.043594 | 1.36057328 |

**Analysis of enriched genes from the comparison between CAL-27**

**Downregulated Genes**

| Term | Description |
|---|---|
| hsa00190 | oxidative phosphorylation |
| GO:0098800 | inner mitochondrial membrane protein complex |
| GO:0098803 | respiratory chain complex |
| GO:0005746 | mitochondrial respiratory chain |
| GO:0070469 | respiratory chain |
| GO:0044455 | mitochondrial membrane part |
| GO:0006119 | oxidative phosphorylation |
| GO:0008137 | NADH dehydrogenase (ubiquinone) activity |
| GO:0050136 | NADH dehydrogenase (quinone) activity |
| GO:0003954 | NADH dehydrogenase activity |
| GO:0045271 | respiratory chain complex I |
| GO:0030964 | NADH dehydrogenase complex |
| GO:0005747 | mitochondrial respiratory chain complex I |
| hsa05208 | Chemical carcinogenesis - reactive oxygen species |
| GO:0098798 | mitochondrial protein complex |
| GO:0042775 | mitochondrial ATP synthesis coupled electron transport |
| GO:0042773 | ATP synthesis coupled electron transport |
| GO:0016655 | oxidoreductase activity |
| GO:0022904 | respiratory electron transport chain |

**Upregulated Genes**

| Term | Description |
|---|---|
| GO:0006959 | humoral immune response |
| GO:0002253 | activation of immune response |
| GO:0045088 | regulation of innate immune response |

| | |
|---|---|
| GO:0006954 | inflammatory response |
| GO:0034341 | response to interferon-gamma |
| GO:0045089 | positive regulation of innate immune response |
| GO:0071346 | cellular response to interferon-gamma |
| GO:0002764 | immune response-regulating signaling pathway |
| GO:0060326 | cell chemotaxis |
| GO:0002526 | acute inflammatory response |
| GO:0071621 | granulocyte chemotaxis |
| GO:0002757 | immune response-activating signal transduction |
| GO:0002697 | regulation of immune effector process |
| GO:0009986 | cell surface |
| GO:0030593 | neutrophil chemotaxis |
| GO:0050663 | cytokine secretion |
| GO:0002224 | toll-like receptor signaling pathway |
| GO:0030449 | regulation of complement activation |
| GO:2000257 | regulation of protein activation cascade |
| GO:0001819 | positive regulation of cytokine production |
| GO:0002758 | innate immune response-activating signal transduction |
| GO:0050727 | regulation of inflammatory response |
| GO:0045785 | positive regulation of cell adhesion |
| GO:0048247 | lymphocyte chemotaxis |
| GO:0006956 | complement activation |
| R-HSA-977606 | regulation of complement cascade |
| GO:0002218 | activation of innate immune response |
| GO:0002920 | regulation of humoral immune response |
| GO:0004866 | endopeptidase inhibitor activity |
| R-HSA-166658 | complement cascade |
| GO:0002449 | lymphocyte mediated immunity |
| GO:0001906 | cell killing |
| GO:0050707 | regulation of cytokine secretion |
| GO:0031341 | regulation of cell killing |
| GO:0050701 | interleukin-1 secretion |

| | |
|---|---|
| GO:0052548 | regulation of endopeptidase activity |
| GO:0071357 | cellular response to type I interferon |
| GO:0004252 | serine-type endopeptidase activity |
| GO:0005539 | glycosaminoglycan binding |
| GO:0004867 | serine-type endopeptidase inhibitor activity |
| GO:0061134 | peptidase regulator activity |
| GO:0022407 | regulation of cell-cell adhesion |
| GO:0001525 | angiogenesis |
| GO:0014066 | regulation of phosphatidylinositol 3-kinase signaling |
| GO:0008329 | signaling pattern recognition receptor activity |
| GO:0034142 | toll-like receptor 4 signaling pathway |
| R-HSA-877300 | Interferon gamma signaling |

**cells bearing KDM3C SNP-S464T versus KDM3C WT-S464 24hrs after 2Gy IR**

| Genes | P value | FDR |
|---|---|---|
| *MT-ATP6, MTATP6P1, MT-ATP8, MT-CO1, MT-CYB, MT-ND1, MT-ND2, MTND2P28, MT-ND4, MT-ND4L, MT-ND5* | 3.82E-07 | 0.001149353 |
| *MT-ATP6, MT-ATP8, MT-CO1, MT-CYB, MT-ND1, MT-ND2, MT-ND4, MT-ND4L, MT-ND5* | 5.03E-06 | 0.001323871 |
| *MT-CO1, MT-CYB, MT-ND1, MT-ND2, MT-ND4, MT-ND4L, MT-ND5* | 1.27E-05 | 0.001323871 |
| *MT-CO1, MT-CYB, MT-ND1, MT-ND2, MT-ND4, MT-ND4L, MT-ND5* | 2.48E-05 | 0.001754997 |
| *MT-CO1, MT-CYB, MT-ND1, MT-ND2, MT-ND4, MT-ND4L, MT-ND5* | 6.11E-06 | 0.001754997 |
| *MT-ATP6, MT-ATP8, MT-CO1, MT-CYB, MT-ND1, MT-ND2, MT-ND4, MT-ND4L, MT-ND5* | 1.87E-05 | 0.003488865 |
| *MT-ATP6, MT-ATP8, MT-CO1, MT-CYB, MT-ND1, MT-ND2, MT-ND4, MT-ND4L, MT-ND5* | 1.90E-06 | 0.004542701 |
| *MT-ND1, MT-ND2, MT-ND4, MT-ND4L, MT-ND5* | 1.04E-05 | 0.005480448 |
| *MT-ND1, MT-ND2, MT-ND4, MT-ND4L, MT-ND5* | 1.04E-05 | 0.005480448 |
| *MT-ND1, MT-ND2, MT-ND4, MT-ND4L, MT-ND5* | 1.40E-05 | 0.005480448 |
| *MT-ND1, MT-ND2, MT-ND4, MT-ND4L, MT-ND5* | 0.000250633 | 0.005576693 |
| *MT-ND1, MT-ND2, MT-ND4, MT-ND4L, MT-ND5* | 0.000250633 | 0.005576693 |
| *MT-ND1, MT-ND2, MT-ND4, MT-ND4L, MT-ND5* | 0.000250633 | 0.005576693 |
| *MT-ATP6, MTATP6P1, MT-ATP8, MT-CO1, MT-CYB, MT-ND1, MT-ND2, MTND2P28, MT-ND4, MT-ND4L, MT-ND5* | 3.72E-06 | 0.005906754 |
| *MT-ATP6, MT-ATP8, MT-CO1, MT-CYB, MT-ND1, MT-ND2, MT-ND4, MT-ND4L, MT-ND5* | 0.00018209 | 0.006293761 |
| *MT-CO1, MT-CYB, MT-ND1, MT-ND2, MT-ND4, MT-ND4L, MT-ND5* | 3.66E-06 | 0.010772022 |
| *MT-CO1, MT-CYB, MT-ND1, MT-ND2, MT-ND4, MT-ND4L, MT-ND5* | 4.04E-06 | 0.010772022 |
| *MT-ND1, MT-ND2, MT-ND4, MT-ND4L, MT-ND5* | 5.10E-05 | 0.011424149 |
| *MT-CO1, MT-CYB, MT-ND1, MT-ND2, MT-ND4, MT-ND4L, MT-ND5* | 1.82E-05 | 0.024640177 |

| Genes | P value | FDR |
|---|---|---|
| *BLNK, C1R, CFB, CFH, LTF, PGLYRP3, PGLYRP4, S100A7, S100A8, S100A9, SLPI* | 8.45E-09 | 0.00000519 |
| *C1R, CD14, CEACAM1, CFB, CFH, GFI1, LTF, MUC4, PGLYRP3, PGLYRP4, PIK3AP1, RSAD2, S100A8, S100A9, TLR5, TNIP3, VTCN1* | 1.97E-07 | 0.00000519 |
| *CD14, CEACAM1, GBP5, GFI1, LTF, MUC4, PGLYRP3, PGLYRP4, PIK3AP1, RASGRP1, RSAD2, S100A8, S100A9, SERPINB4, TLR5, TNIP3, WNT5A* | 2.36E-06 | 0.000576 |

| | | |
|---|---|---|
| *BLNK, C1R, CCL22, CD14, CFB, CFH, CX3CL1, GBP5, IDO1, NGFR, NUPR1, PIK3AP1, RASGRP1, S100A8, S100A9, SAA2, TLR5, TNIP3, WNT5A* | 4.14E-11 | 0.00078 |
| *CCL22, CX3CL1, GBP2, GBP5, IFITM1, OASL, WNT5A* | 9.11E-07 | 0.000842 |
| *CD14, GBP5, GFI1, LTF, MUC4, PGLYRP3, PGLYRP4, PIK3AP1, RASGRP1, RSAD2, S100A8, S100A9, TLR5, TNIP3, WNT5A* | 2.13E-06 | 0.000886 |
| *CCL22, CX3CL1, GBP2, GBP5, OASL, WNT5A* | 2.73E-06 | 0.00116 |
| *CD14, CEACAM1, GFI1, LTF, MUC4, PGLYRP3, PGLYRP4, PIGR, PIK3AP1, RSAD2, S100A8, S100A9, TLR5, TNIP3, VTCN1* | 1.46E-05 | 0.00129 |
| *CCL22, CX3CL1, EGR3, PADI2, S100A7, S100A8, S100A9, SAA2, WNT5A* | 5.05E-09 | 0.00152 |
| *C1R, CFB, CFH, NUPR1, S100A8, SAA2* | 5.11E-07 | 0.00182 |
| *CCL22, CX3CL1, S100A7, S100A9* | 4.68E-10 | 0.00204 |
| *CD14, CEACAM1, GFI1, LTF, MUC4, PGLYRP3, PGLYRP4, PIK3AP1, RSAD2, S100A8, S100A9, TLR5, TNIP3, VTCN1* | 6.09E-05 | 0.00224 |
| *ANGPT1, C1R, CEACAM1, CFB, CFH, PGLYRP3, RASGRP1, RSAD2, SERPINB4, WNT5A* | 7.83E-05 | 0.00244 |
| *ABCA1, ABCG1, ADGRV1, CD14, CD34, CEACAM1,* | 0.000113924 | 0.00265 |
| *CCL22, CX3CL1, S100A8, S100A9* | 5.01E-08 | 0.00279 |
| *ABCA1, ANGPT1, CARD17, CD14, CD34, GBP5, RASGRP1, TLR5, VTCN1, WNT5A* | 0.000137151 | 0.00341 |
| *CD14, GFI1, LTF, PIK3AP1, RSAD2, S100A8, S100A9, TLR5, TNIP3* | 1.58E-05 | 0.00364 |
| *C1R, CFB, CFH* | 5.79E-05 | 0.00408 |
| *C1R, CFB, CFH* | 5.79E-05 | 0.00408 |
| *CD14, CD34, CX3CL1, GBP5, IDO1, LUM, RASGRP1, RSAD2, TLR5, VTCN1, WNT5A* | 0.000162124 | 0.00474 |
| *CD14, GFI1, LTF, MUC4, PGLYRP3, PGLYRP4, PIK3AP1, RSAD2, S100A8, S100A9, TLR5, TNIP3* | 2.40E-05 | 0.00491 |
| *C1R, CFB, CFH, CX3CL1, GBP5, IDO1, PIK3AP1, S100A8, S100A9, WNT5A* | 1.93E-06 | 0.0057 |
| *AGR2, ANGPT1, CX3CL1, OLFM4, RASGRP1, VTCN1, WNT4, WNT5A* | 0.000343426 | 0.00622 |
| *CCL22, CX3CL1, PADI2, S100A7, WNT5A* | 0.000334639 | 0.00627 |
| *C1R, CFB, CFH* | 3.65E-05 | 0.00654 |
| *C1R, CFB, CFH* | 3.86E-05 | 0.0072 |
| *CD14, GFI1, LTF, MUC4, PGLYRP3, PGLYRP4, PIK3AP1, RSAD2, S100A8, S100A9, TLR5, TNIP3* | 3.02E-05 | 0.00749 |
| *C1R, CFB, CFH* | 7.01E-05 | 0.00969 |
| *CARD17, LTF, SERPINA5, SERPINB3, SERPINB4, SLPI, WFDC2* | 1.80E-05 | 0.0102 |
| *C1R, CFB, CFH* | 0.000137691 | 0.013 |
| *C1R, CEACAM1, GCNT3, RASGRP1, RSAD2, SERPINB4* | 0.000463487 | 0.013 |
| *CEACAM1, LTF, PGLYRP3, PGLYRP4, RASGRP1, SERPINB4* | 0.001132894 | 0.014 |
| *ANGPT1, CARD17, CD14, CD34, RASGRP1, TLR5, VTCN1, WNT5A* | 0.001224469 | 0.0143 |
| *CEACAM1, PGLYRP3, PGLYRP4, RASGRP1, SERPINB4* | 0.001410279 | 0.0149 |
| *ABCA1, CARD17, GBP5, WNT5A* | 0.000914756 | 0.0185 |

| | | |
|---|---|---|
| *CARD17, CTGF, LTF, S100A8, S100A9, SERPINA5, SERPINB3, SERPINB4, SLPI, TNFSF10, WFDC2* | 1.61E-05 | 0.0191 |
| *GBP2, IFITM1, OASL, RSAD2, WNT5A* | 1.79E-10 | 0.0197 |
| *C1R, CFB, LTF, MMP7, TMPRSS3, TMPRSS4* | 7.74E-10 | 0.0208 |
| *CFH, CTGF, EVA1C, LTF, MMP7, PGLYRP3, PGLYRP4, SERPINA5* | 4.38E-07 | 0.0208 |
| *SERPINA5, SERPINB3, SERPINB4, SLPI, WFDC2* | 8.12E-05 | 0.0222 |
| *CARD17, LTF, SERPINA5, SERPINB3, SERPINB4, SLPI, WFDC2* | 0.000121277 | 0.0225 |
| *CEACAM1, CX3CL1, IDO1, RASGRP1, VTCN1, WNT4, WNT5A* | 0.002355728 | 0.0229 |
| *ANGPT1, CD34, CEACAM1, CTGF, CX3CL1, EGR3, FOXO4, LOXL4, LRG1, NGFR, S100A7, WNT5A* | 0.001801723 | 0.0257 |
| *ANGPT1, CEACAM1, CSF3, PIK3AP1, PIK3IP1, RASGRP1* | 0.002648058 | 0.0296 |
| *CD14, PGLYRP3, PGLYRP4* | 0.000812717 | 0.0315 |
| *CD14, LTF, PIK3AP1, TNIP3* | 0.0024198 | 0.0381 |
| *GBP2, GBP5, OASL* | 0.00109692 | 0.0427 |

**negative Log10**

2.939546566

2.878154331

2.878154331

2.755723622

2.755723622

2.457315835

2.342685847

2.261183939

2.261183939
2.261183939
2.253623263
2.253623263
2.253623263
2.228651116

2.201089753

1.967702768

1.967702768

1.942176141
1.608356177

**negative Log10**

5.284832642

5.284832642

3.239577517

3.107905397

3.074687909

3.052566278

2.935542011
2.88941029

2.818156412

2.739928612
2.690369833
2.649751982

2.612610174

2.576754126
2.554395797
2.467245621

2.438898616

2.389339837
2.389339837
2.324221658

2.308918508

2.244125144

2.206209615

2.202732459
2.184422252
2.142667504
2.125518182

2.013676223
1.991399828

1.886056648
1.886056648

1.853871964

1.844663963

1.826813732

1.732828272

1.718966633

1.705533774
1.681936665
1.681936665

1.653647026
1.647817482

1.640164518

1.590066877

1.528708289

1.501689446
1.419075024
1.369572125

**Analysis of enriched genes from the comparison betwe**

**Downregulated Genes**

| Term | Description |
|---|---|
| GO:0031012 | extracellular matrix |
| GO:0004888 | transmembrane signaling receptor activity |
| GO:0043062 | extracellular structure organization |
| GO:0071774 | response to fibroblast growth factor |
| R-HSA-1474244 | Extracellular matrix organization |
| R-HSA-388396 | GPCR downstream signalling |

**Upregulated Genes**

| Term | Description |
|---|---|
| GO:0048018 | receptor ligand activity |
| GO:0008083 | growth factor activity |
| GO:0060231 | mesenchymal to epithelial transition |
| GO:0097191 | extrinsic apoptotic signaling pathway |
| GO:0018212 | peptidyl-tyrosine modification |
| hsa04060 | Cytokine-cytokine receptor interaction |

**en SCC-9 cells bearing KDM3C SNP-S464T versus KDM3C WT-S464 allele at baseline**

| Genes | P value | FDR |
|---|---|---|
| *ADAMTS2, COL1A2, FLRT2, FLRT3, LAMA4, LTBP1, MMP9, PTN, PTPRZ1, WISP1* | 3.01E-09 | 1.01E-06 |
| *ABCA1, ADGRL3, LTB4R, CMKLR1, FGFR3, LTBP1, PTPRZ1* | 3.34E-06 | 0.002017 |
| *ABCA1, ADAMTS2, COL1A2, CREB3L1, FLRT2, LAMA4, MMP9* | 1.53E-07 | 0.00013037 |
| *CREB3L1, FGFR3, FLRT2, FLRT3* | 0.000017512 | 0.0074426 |
| *ADAMTS2, COL1A2, LAMA4, LTBP1, MMP9* | 2.82E-05 | 0.009617802 |
| *LTB4R, PLCB1* | 1.54E-03 | 0.039482645 |

| Genes | P value | FDR |
|---|---|---|
| *CSF2, CXCL8, EREG, GDNF, LIF, NGF, TNFRSF11B* | 5.36E-08 | 2.36E-05 |
| *CSF2, EREG, GDNF, LIF, NGF* | 2.80E-07 | 4.05E-05 |
| *GDNF, LIF* | 0.000048297 | 0.013684 |
| *CSF2, GDNF, NGF* | 1.71E-04 | 2.92E-02 |
| *CD74, CSF2, EREG, LIF* | 0.0000302 | 0.013684 |
| *CSF2, CXCL8, LIF, NGF* | 3.71E-06 | 0.000641773 |

| negative Log10 |
|---|
| 5.995678626 |
| 2.695294102 |
| 3.884822334 |
| 2.128275321 |
| 2.016924168 |
| 1.403593761 |

| negative Log10 |
|---|
| 4.627087997 |
| 4.392544977 |
| 1.863786934 |
| 1.535331645 |
| 1.863786934 |
| 3.192618558 |

**Analysis of enriched genes from the comparison between SCC-9 c**

**Downregulated Genes**

| **Term** | **Description** |
|---|---|
| GO:0009913 | epidermal cell differentiation |
| GO:0005882 | intermediate filament |
| GO:0045111 | intermediate filament cytoskeleton |
| GO:0031012 | extracellular matrix |
| GO:0016327 | apicolateral plasma membrane |
| GO:0043296 | apical junction complex |
| GO:0005911 | cell-cell junction |
| GO:0070160 | occluding junction |
| GO:0005578 | proteinaceous extracellular matrix |
| GO:0005200 | structural constituent of cytoskeleton |
| GO:0008236 | serine-type peptidase activity |
| GO:0004252 | serine-type endopeptidase activity |
| hsa04915 | estrogen signaling pathway |

**Upregulated Genes**

| **Term** | **Description** |
|---|---|
| GO:0010469 | regulation of signaling receptor activity |
| GO:0031012 | extracellular matrix |
| GO:0008083 | growth factor activity |
| GO:0005125 | cytokine activity |

**ells bearing KDM3C SNP-S464T versus KDM3C WT-S464 allele 24h after 2Gy IR**

| **Genes** | **P value** | **FDR** |
|---|---|---|
| *AKR1C3, AQP3, CDH3, DSG3, IVL, JAG2, JUP, KLK14, KLK8, KRT13, KRT14, KRT15, KRT16, KRT17, KRT19, KRT20, KRT5, KRT6C, PPL, SCEL, SERPINB13, TGM1* | 4.45E-13 | 5.18E-09 |
| *GJA1, JUP, KRT13, KRT14, KRT15, KRT16, KRT17, KRT19, KRT20, KRT5, KRT6C, NEFL* | 2.37E-09 | 2.46E-06 |
| *GJA1, JUP, KRT13, KRT14, KRT15, KRT16, KRT17, KRT19, KRT20, KRT5, KRT6C, NEFL, S100A8* | 1.20E-07 | 3.09E-05 |
| *COL1A2, EGFL7, FGFBP1, L1CAM, LAMA4, MFAP5, MMP7, MMP9, PTN, VWF* | 1.07E-06 | 7.40E-04 |
| *CLDN4, CXADR, JUP, KRT19, OCLN* | 4.23E-07 | 1.55E-03 |
| *AMOT, CLDN4, CLDN9, CXADR, JUP, MPP7, OCLN, POF1B* | 0.00013879 | 1.27E-02 |
| *AMOT, AQP3, CDH3, CLDN4, CLDN9, CXADR, DSG3, GJA1, JUP, MPP7, OCLN, POF1B, PPL* | 3.40E-05 | 1.07E-02 |
| *AMOT, CLDN4, CLDN9, CXADR, MPP7, OCLN, POF1B* | 0.000619651 | 1.88E-02 |
| *COL1A2, LAMA4, MFAP5, MMP7, MMP9, PTN, VWF* | 8.33E-05 | 1.40E-02 |
| *KRT14, KRT15, KRT16, KRT17, KRT19, KRT20, KRT5, NEFL, PPL* | 0.000457335 | 1.45E-02 |
| *KLK14, KLK8, MMP7, MMP9* | 0.000457335 | 2.53E-02 |
| *KLK14, KLK8, MMP7, MMP9* | 7.67E-05 | 1.96E-02 |
| *KRT13, KRT14, KRT15, KRT16, KRT17, KRT19, KRT20, MMP9* | 0.000376153 | 1.53E-02 |

| **Genes** | **P value** | **FDR** |
|---|---|---|
| *CSF2, CXCL3, EREG, FBN1, GDF15, GDNF, IL24, IL6, NRP1, TNFRSF11B* | 7.06E-08 | 0.01141531 |
| *ADAMTS3, ANGPTL4, CDH2, FBN1, SLIT3, TNFRSF11B, VCAN* | 4.67E-06 | 0.00364029 |
| *CSF2, EREG, GDF15, GDNF, IL6* | 5.69E-06 | 0.013300052 |
| *CSF2, CXCL3, GDF15, IL24, IL6, TNFRSF11B* | 0.001914219 | 0.041453054 |

**negative Log10**

8.285974468

5.608639027

4.510710887

3.130817912

2.809753188

1.895501358

1.972067651

1.726878809

1.855293945

1.837833416

1.596980389

1.707871219

1.816065257

**negative Log10**

1.942512277

2.438864056

1.876146649

1.382443472

**Supplementary Table 19. Primer sequence used for SNP genotyping**

| Target gene | Forward (5'-3') | Reverse (5'-3') |
|---|---|---|
| *KDM3C* (S464T) | ACATTTTGGTGTGGGTGGTC | GGCAGGAGAAGAGACCCTAAA |

**Supplementary Table 20. Primer sequences for target genes used in the stu**

| **Target gene** | **Forward (5'-3')** |
|---|---|
| human *36B4* | CAGATTGGCTACCCAACTGTT |
| human *IFNB* | ATGACCAACAAGTGTCTCCTCC |
| human *CXCL10* | GTGGCATTCAAGGAGTACCTC |

**udy**

| Reverse (5'-3') |
| --- |
| GGAAGGTGTAATCCGTCTCCAC |
| GGAATCCAAGCAAGTTGTAGCTC |
| TGATGGCCTTCGATTCTGGATT |
